# Prevalence of comorbidities and their impact on survival among older adults with the five most common cancers in Taiwan: A population study

**DOI:** 10.1101/2022.10.29.22281698

**Authors:** Li-Hsin Chien, Tzu-Jui Tseng, Tzu-Yu Chen, Chung-Hsing Chen, Chia-Yu Chen, Hsin-Fang Jiang, Fang-Yu Tsai, Hsiu-Ying Ku, Shih Sheng Jiang, Chao A. Hsiung, Tsang-Wu Liu, I-Shou Chang

## Abstract

Because of the cancer incidence increase and population aging in Taiwan, we aimed to assess the cancer prevalence, to summarize the comorbidities of older patients with the five most common cancers (i.e., breast, colorectal, liver, lung, and oral), and to develop a Taiwan comorbidity index (TCI) for studying their actual prognosis. The linkage of the Taiwan Cancer Registry, Cause of Death Database, and National Health Insurance Research Database was used. We followed the standard statistical learning steps to obtain a survival model with good discriminatory accuracy in predicting death due to noncancer causes, from which we obtained the TCI and defined comorbidity levels. We reported the actual prognosis by age, stage, and comorbidity level. In Taiwan, cancer prevalence nearly doubled in 2004–2014, and comorbidities were common among older patients. Stage was the major predictor of patients’ actual prognoses. For localized and regional breast, colorectal, and oral cancers, comorbidities correlated with noncancer-related deaths. Compared with the US, the chances of dying from comorbidities in Taiwan were lower and the chances of dying from cancer were higher for breast, colorectal, and male lung cancers. These actual prognoses could help clinicians and patients in treatment decision-making and help policymakers in resource planning.

## Introduction

Cancer is an important disease, and comorbidities are common among older adults with cancer in developed countries. Comorbidities can affect treatment decisions and outcomes. Clinical management and treatment decision-making must be improved for older adults with cancer because of their comorbidities. One critical strategy is to develop standardized comorbidity measurements to assess the impact of specific combinations of comorbidities on older adults with cancer [1-4].

Important advances in this direction include determining the 5-year chances of dying from cancer and from noncancer by age, stage, and comorbidity levels for older patients with breast, prostate, colorectal, or lung cancers in the US [5-7]. These measures of patients’ actual prognoses provide important information for clinicians and patients to determine their treatment options, for policymakers to allocate healthcare resources, and for researchers to design trials for cancer treatments in older adults with cancer and pre-existing comorbidities [8-13].

The comorbidity level in these studies was determined jointly by clinical judgment and the National Cancer Institute combined comorbidity index (NCICI), which reflects the hazard ratio associated with the time from cancer diagnosis to noncancer-related death [14-16].

Because industrialization in Taiwan started in the 1960s, the cancer incidence is increasing, and the population is aging. The number of Taiwanese residents aged ≥65 years increased from 2.5% in 1955 to 13.9% in 2017 [17-19]. These observations, together with an improved net survival for patients with cancer in Taiwan [19], suggest that cancer survivors are likely to become more prevalent, and studies of comorbidities among patients with cancer may be timely for better clinical management and surveillance [20-22].

This study aimed to report the prevalence of cancer survivors in Taiwan and their comorbidities before cancer diagnosis, to develop a Taiwan comorbidity index (TCI) for older patients with the five most common cancers (i.e., breast, colorectal, liver, lung, and oral) and to use it to study their actual prognoses by age, stage, and comorbidity levels. Because the prevalence of comorbidities and their effects on cancer patients in Taiwan may be different from those in developed countries, we followed the NCICI to develop the TCI by slightly modifying the coding of a few comorbidities.

## Methods

### Overview

This study was based on the linkage of the Taiwan Cancer Registry (TCR), National Health Insurance Research Database (NHIRD), and Cause of Death Database (TCOD); they were used in our earlier studies [23-25]. Taking advantage of these large datasets, we followed the standard three steps in statistical learning (i.e., model training, model selection, and model assessment) to obtain a survival model with good discriminatory accuracy in predicting death due to noncancer causes; see, for example, Chapter 7 of Hastie, Tibshirani, and Friedman [26]. We systematically fitted several Cox’s regression models using different training sets and selected among them models having a large area under the receiver operating characteristic curve (AUC) using validation sets; we reported the performances using test sets. This AUC, a predictive accuracy measure, was the time-dependent extension of the analysis by Heagerty and Zheng [27].

Because the main effects of certain comorbidities may have hazard ratios <1 in fitting a Cox regression model with several comorbidities, which happens when correlation exists among them, and the index derived from such models are less intuitive, we deleted the comorbid conditions having negative coefficients and refitted the Cox models. It was among these more intuitive ones that we performed model selection using validation sets[26]. Having chosen the model, we reported its performance using the test set. In addition, we carefully determined the time periods for assessing comorbidities before developing the TCI [28, 29]. Using the TCI and clinical judgements, we considered three comorbidity levels. The actual prognoses of cancer patients were studies by age, cancer stage, and comorbidity level.

### Study population

We examined patients’ first invasive primary cancer reported in the TCR during 2004–2014. We obtained patients’ vital statuses from the TCOD for 2004–2016 and their comorbidity information from the NHIRD from 2000 to the time of cancer diagnosis.

The TCR collects information on patients of primary cancers at all hospitals in Taiwan having 50 or more beds. The quality of the TCR is improving and was reviewed before [17, 30]. The TCR included 1,934,198 records for 1979–2014, with one record for each primary cancer. After basic data checks and cleaning using birth date and sex, 1,852,694 cancer cases involving 1,699,907 patients were included.

Taiwan’s National Health Insurance (NHI) program (implemented in 1995 by the NHI Administration) provides compulsory universal health insurance and covers all health care services for more than 99% of Taiwan’s population. The NHIRD is built on data from this program, and we used the 2000–2015 data in this study. Data in the NHIRD have proven to be valuable resources for health science research [31].

TCOD has included the cause of death for individuals in Taiwan since 1971 and used the national identification card number (NICN) since 1985 [32]. The original TCOD contained 4,191,373 individual records for 1985–2016. Eliminating inconsistencies in the NICN, sex, birth date, death date, and cause of death yielded 4,054,632 unique death records for this period.

We studied patients diagnosed with primary breast, colorectal, liver, lung, and oral cancers between 2004 and 2014. Table S1 presents the ICD9 codes for these cancers. From 2004, 27 hospitals, and from 2007, all participating hospitals were required to use the TCR long-form to collect information on patients with these cancers, including the stage. Table S2 reports the numbers of patients with these five cancers in the TCR.

### Noncancer death

This study used the Surveillance, Epidemiology, and End Results Program (SEER) cause-specific death classification algorithm to define noncancer-related death [33]. This algorithm has been essential for cancer survivorship studies [5-7, 16]. Our earlier publication used this algorithm and showed that cause-specific survival and relative survival for common cancers in Taiwan are comparable, thus suggesting the validity of this SEER algorithm in Taiwan [25].

### Comorbidity definition

The comorbidities considered were mainly adopted from those in NCICI except including hypertension without and with complications and modifying mild liver diseases by incorporating viral hepatitis B and C to reflect their high prevalence in Taiwan and their roles in cancer development. We considered 18 comorbidities in this study; they are shown in Table 1, and their ICD 9 codes are shown in Table S3. They included 16 of the 19 comorbidities defining the Charlson Comorbidity Index but excluded solid cancer, leukemia, and lymphoma because of our study focus [34].

**Table 1.**
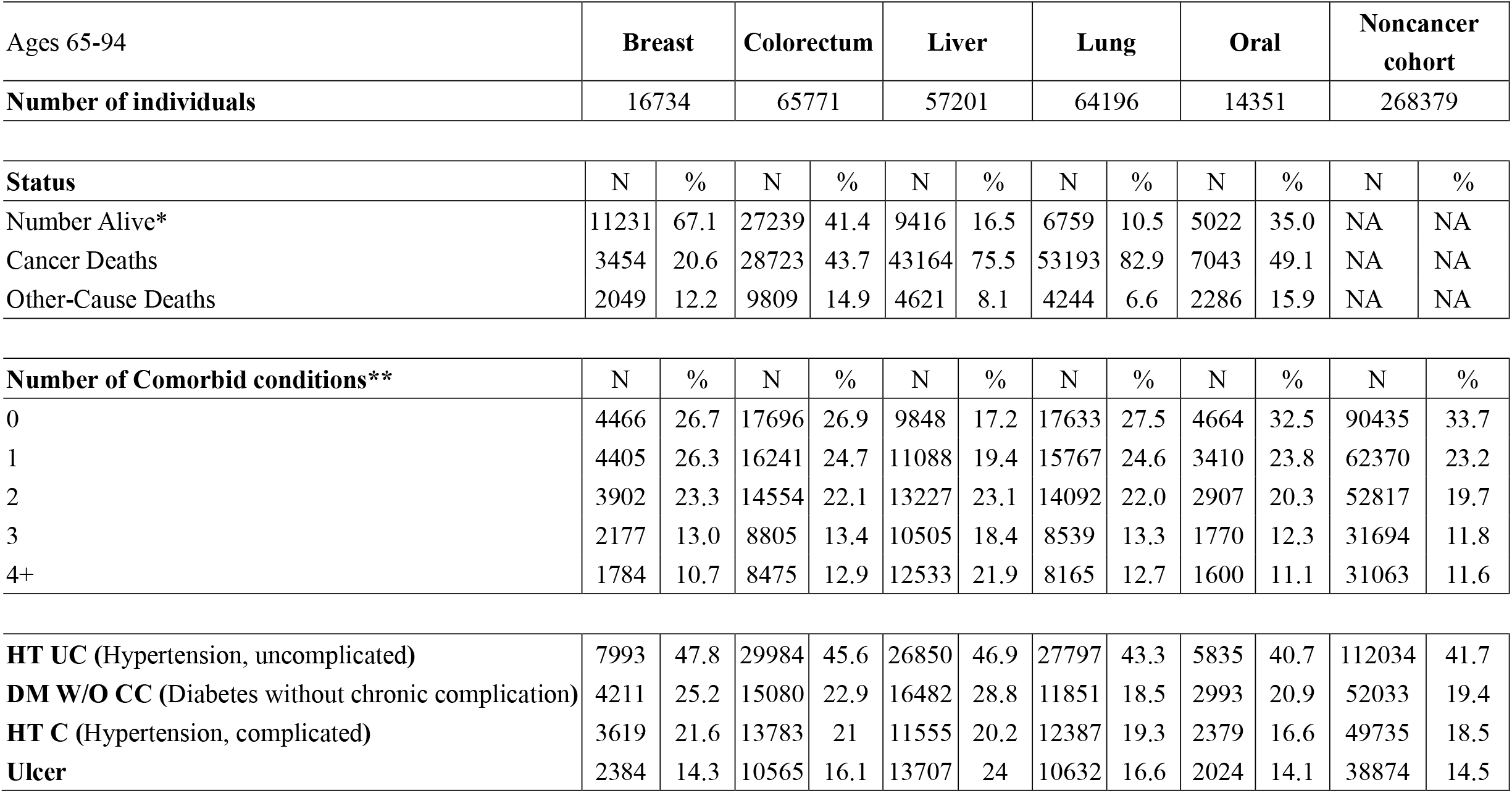
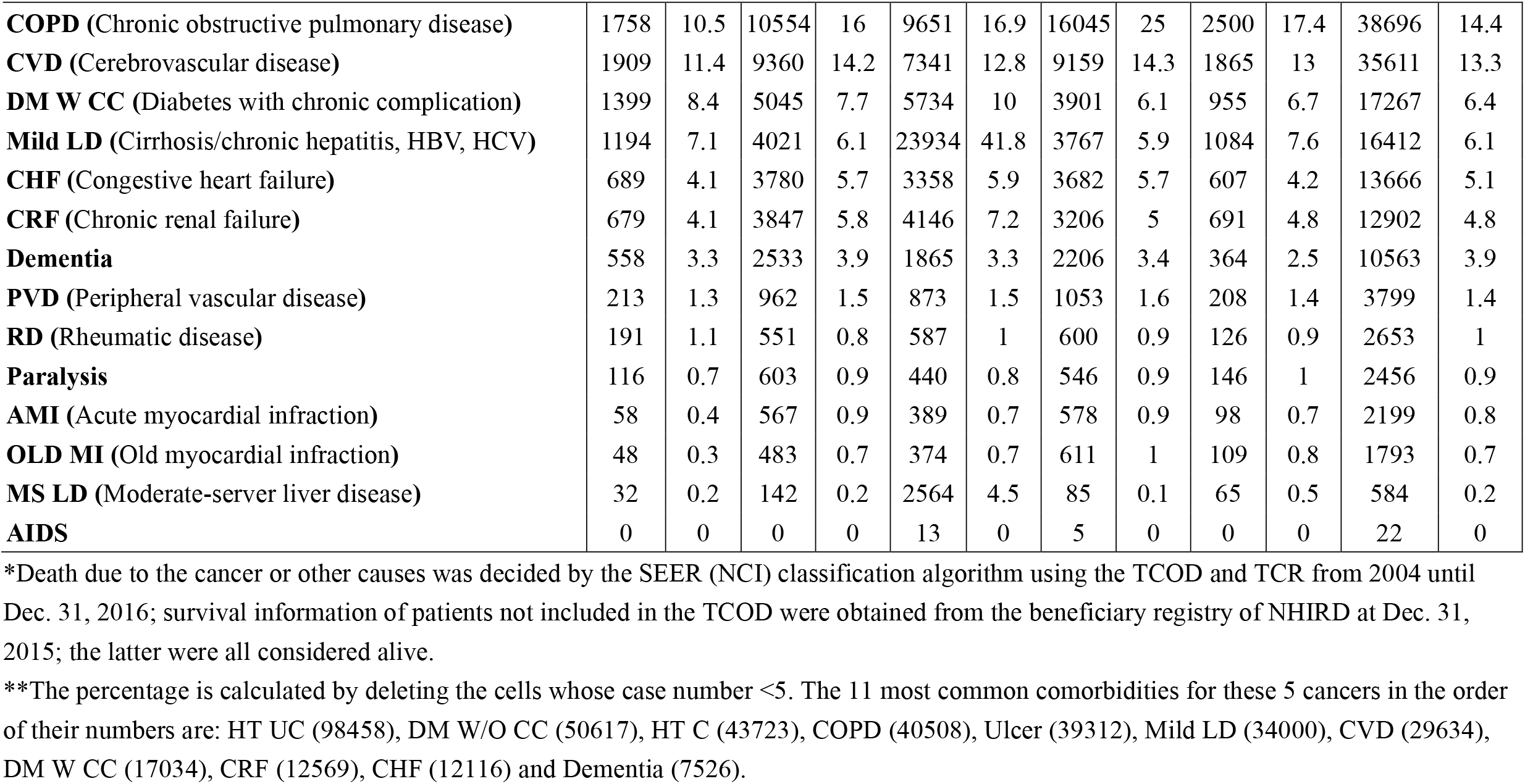
The prevalence of selected comorbidities for the 5 most common cancers in Taiwan diagnosed in 2004-2014 and for individuals without cancer sampled in 2004-2014, ages 65-94.

| Ages 65-94 | **Breast** |  | **Colorectum** |  | **Liver** |  | **Lung** |  | **Oral** |  | **Noncancer cohort** |
| --- | --- | --- | --- | --- | --- | --- | --- | --- | --- | --- | --- |
| **Number of individuals** | 16734 |  | 65771 |  | 57201 |  | 64196 |  | 14351 |  | 268379 |

| **Status** | N | % | N | % | N | % | N | % | N | % | N | % |
| --- | --- | --- | --- | --- | --- | --- | --- | --- | --- | --- | --- | --- |
| Number Alive* | 11231 | 67.1 | 27239 | 41.4 | 9416 | 16.5 | 6759 | 10.5 | 5022 | 35.0 | NA | NA |
| Cancer Deaths | 3454 | 20.6 | 28723 | 43.7 | 43164 | 75.5 | 53193 | 82.9 | 7043 | 49.1 | NA | NA |
| Other-Cause Deaths | 2049 | 12.2 | 9809 | 14.9 | 4621 | 8.1 | 4244 | 6.6 | 2286 | 15.9 | NA | NA |

| **Number of Comorbid conditions\*\*** | N | % | N | % | N | % | N | % | N | % | N | % |
| --- | --- | --- | --- | --- | --- | --- | --- | --- | --- | --- | --- | --- |
| 0 | 4466 | 26.7 | 17696 | 26.9 | 9848 | 17.2 | 17633 | 27.5 | 4664 | 32.5 | 90435 | 33.7 |
| 1 | 4405 | 26.3 | 16241 | 24.7 | 11088 | 19.4 | 15767 | 24.6 | 3410 | 23.8 | 62370 | 23.2 |
| 2 | 3902 | 23.3 | 14554 | 22.1 | 13227 | 23.1 | 14092 | 22.0 | 2907 | 20.3 | 52817 | 19.7 |
| 3 | 2177 | 13.0 | 8805 | 13.4 | 10505 | 18.4 | 8539 | 13.3 | 1770 | 12.3 | 31694 | 11.8 |
| 4+ | 1784 | 10.7 | 8475 | 12.9 | 12533 | 21.9 | 8165 | 12.7 | 1600 | 11.1 | 31063 | 11.6 |
| **HT UC (Hypertension, uncomplicated)** | 7993 | 47.8 | 29984 | 45.6 | 26850 | 46.9 | 27797 | 43.3 | 5835 | 40.7 | 112034 | 41.7 |
| **DM W/O CC (Diabetes without chronic complication)** | 4211 | 25.2 | 15080 | 22.9 | 16482 | 28.8 | 11851 | 18.5 | 2993 | 20.9 | 52033 | 19.4 |
| **HT C (Hypertension, complicated)** | 3619 | 21.6 | 13783 | 21 | 11555 | 20.2 | 12387 | 19.3 | 2379 | 16.6 | 49735 | 18.5 |
| **Ulcer** | 2384 | 14.3 | 10565 | 16.1 | 13707 | 24 | 10632 | 16.6 | 2024 | 14.1 | 38874 | 14.5 |
| **COPD** (Chronic obstructive pulmonary disease) | 1758 | 10.5 | 10554 | 16 | 9651 | 16.9 | 16045 | 25 | 2500 | 17.4 | 38696 | 14.4 |
| **CVD** (Cerebrovascular disease) | 1909 | 11.4 | 9360 | 14.2 | 7341 | 12.8 | 9159 | 14.3 | 1865 | 13 | 35611 | 13.3 |
| **DM W CC** (Diabetes with chronic complication) | 1399 | 8.4 | 5045 | 7.7 | 5734 | 10 | 3901 | 6.1 | 955 | 6.7 | 17267 | 6.4 |
| **Mild LD** (Cirrhosis/chronic hepatitis, HBV, HCV) | 1194 | 7.1 | 4021 | 6.1 | 23934 | 41.8 | 3767 | 5.9 | 1084 | 7.6 | 16412 | 6.1 |
| **CHF** (Congestive heart failure) | 689 | 4.1 | 3780 | 5.7 | 3358 | 5.9 | 3682 | 5.7 | 607 | 4.2 | 13666 | 5.1 |
| **CRF** (Chronic renal failure) | 679 | 4.1 | 3847 | 5.8 | 4146 | 7.2 | 3206 | 5 | 691 | 4.8 | 12902 | 4.8 |
| **Dementia** | 558 | 3.3 | 2533 | 3.9 | 1865 | 3.3 | 2206 | 3.4 | 364 | 2.5 | 10563 | 3.9 |
| **PVD** (Peripheral vascular disease) | 213 | 1.3 | 962 | 1.5 | 873 | 1.5 | 1053 | 1.6 | 208 | 1.4 | 3799 | 1.4 |
| **RD** (Rheumatic disease) | 191 | 1.1 | 551 | 0.8 | 587 | 1 | 600 | 0.9 | 126 | 0.9 | 2653 | 1 |
| **Paralysis** | 116 | 0.7 | 603 | 0.9 | 440 | 0.8 | 546 | 0.9 | 146 | 1 | 2456 | 0.9 |
| **AMI** (Acute myocardial infraction) | 58 | 0.4 | 567 | 0.9 | 389 | 0.7 | 578 | 0.9 | 98 | 0.7 | 2199 | 0.8 |
| **OLD MI** (Old myocardial infraction) | 48 | 0.3 | 483 | 0.7 | 374 | 0.7 | 611 | 1 | 109 | 0.8 | 1793 | 0.7 |
| **MS LD** (Moderate-severe liver disease) | 32 | 0.2 | 142 | 0.2 | 2564 | 4.5 | 85 | 0.1 | 65 | 0.5 | 584 | 0.2 |
| **AIDS** | 0 | 0 | 0 | 0 | 13 | 0 | 5 | 0 | 0 | 0 | 22 | 0 |

\*Death due to the cancer or other causes was decided by the SEER (NCI) classification algorithm using the TCOD and TCR from 2004 until Dec. 31, 2016; survival information of patients not included in the TCOD were obtained from the beneficiary registry of NHIRD at Dec. 31, 2015; the latter were all considered alive.
\*\*The percentage is calculated by deleting the cells whose case number <5. The 11 most common comorbidities for these 5 cancers in the order of their numbers are: HT UC (98458), DM W/O CC (50617), HT C (43723), COPD (40508), Ulcer (39312), Mild LD (34000), CVD (29634), DM W CC (17034), CRF (12569), CHF (12116) and Dementia (7526).

### Intervals defining comorbidity

Because comorbidity assessment depends on the time-interval before cancer diagnosis, we followed Maringe et al.’s study to determine the interval for comorbidity assessment [29]. We present in Table S4 the hazard ratios from fitting Cox regression models, including only single comorbidity as the covariate of interest and using time from cancer diagnosis to noncancer-related death as the outcome, based on all patients with colorectal cancer in the TCR from 2006–2014. Tables S4-1 and 4-2 regard patients aged 15— 64 and 65—94, respectively. We considered three comorbidity assessment time-intervals, which were 30 months, 54 months, and 78 months, before the cancer diagnosis and explored the sex-specific comorbidity effect. In fact, we excluded comorbidities that appeared only in the six months immediately before the cancer diagnosis to reduce the comorbidities caused by the cancers. A patient was said to have a specific comorbidity if their inpatient files contained a diagnosis of this comorbidity within the earlier 24, 48, or 72 months or if their outpatient files contained two diagnoses of this comorbidity in these periods with a gap >1 month. The supplementary materials and Table S4 give more details in this regard.

For the age group 65—94, Table S4-2 shows that for the vast majority of the comorbidities, the differences in hazard ratios among these three assessment periods were small; thus, we decided to consider a 30-month period for the assessment to include more patients in the study.

### Taiwan comorbidity index

We acquired data from the NHIRD and TCOD for each patient with the five studied cancers in the TCR during 2004–2014, aged between 15 and 94 years. This dataset was called “Five-Cancer”. We report in Tables S5-1, for those aged 15–64, and S5-2, for those aged 65–94, the numbers and percentages of these patients who had any of the 18 comorbidities and who were alive at the end of 2016. Five-Cancer was randomly divided into three disjoint parts: one half was the training set, one quarter was the validation set, and the remaining quarter was the test set. Five-Cancer had a total of 501,572 patients, shown in Table S2.

Using the Five-Cancer training set, we fitted Cox’s regression models with time from diagnosis to noncancer-related death as the outcome. Censoring events included cancer-associated deaths or loss to follow-up as per the linkage of the TCR, TCOD, and NHIRD. Table S6-1 presents the estimated coefficients of the Cox model, including all 18 comorbidities and the interactions of any two of the 11 most common comorbidities (“Main18&11”) as covariates. It shows that three of the estimated main effects of the comorbidities were negative. We deleted the comorbidities with negative coefficients altogether and refitted the model until all the main effects were positive; whenever a comorbidity was deleted, interaction terms involving it were also deleted. The resulting model was termed “Main18&11.ND”, for whom Table S6-2 presents the hazard ratios and coefficients.

A patient’s TCI in this study was defined to be the sum of the coefficients in the Cox model Main18&11.ND corresponding to the patient’s comorbid conditions and interaction terms. We chose this for its excellent performance and simplicity.

In fact, we systematically considered 24 subsets of Five-Cancer defined by age, sex, and cancer site and divided each of them randomly into a training set, a validation set, and a test set. These divisions were compatible among these 24 subsets in the sense that if one subset was included in another subset, the training set, validation set, and test set of the former were included in the counterparts of the latter. We fitted several Cox’s regression models to each of the 24 training sets and computed the time-dependent AUC at 1 year, 2 years, and 5 years from diagnosis in each validation set. Table S7 presents the 5-year AUCs evaluated in each of the sex- and site-specific validation set of cancer patients aged 65—94. According to Tables S7-1–S7-9, the more intuitive Cox model Main18&11.ND trained by Five-Cancer performed very well generally across all these validation sets. The supplementary materials detail the construction of the training sets, validation sets, and test sets and the Cox’s models considered and assessed.

### Noncancer cohort comorbidity

We constructed a cohort representing the 2004–2014 Taiwan population without cancer diagnoses using the TCR, TCOD, NHIRD, and Monthly Bulletin of Interior Statistics. Details are given in the supplementary materials.

### Statistical analysis

The fitting of Cox’s models in the training sets was carried out using the R-package ‘survival’. The time-dependent AUCs for the Cox models were obtained using the R-package ‘risksetROC’. The actual prognoses were computed using the R-package ‘cmprsk’.

All methods were performed in accordance with the relevant guidelines and regulations of Scientific Reports. This study used only datasets for which all the personal information had been deidentified by the Health and Welfare Data Science Center, Ministry of Health and Welfare of Taiwan. There was no patient contact for the study; therefore, there was no patient consent process. The Institutional Review Board of National Health Research Institutes, Taiwan approved this study (EC1030707-E) and waived the need for informed consent for this study as part of the study approval. Indeed, all the analyses were conducted in a secured area administered by the Health and Welfare Data Science Center, Ministry of Health and Welfare of Taiwan (https://dep.mohw.gov.tw/DOS/sp-GS-113.html? Query and https://dep.mohw.gov.tw/DOS/sp-GS-113.html?Query). Only summary tables could be brought out after verification by the officials.

## Results

### A rapid increase in the number of cancer survivors

Figure 1 shows that the total number of cancer survivors increased drastically, from 314,107 in 2004 to 610,712 in 2014, and the number of long-term survivors who survived >15 years increased from 29,953 in 2004 to 115,021 in 2014, a 4-fold increase, which was much faster than that in the US [35]. Figure S1 presents the corresponding numbers for each of the five most common cancers, indicating breast cancer in women had more long-term survivors than the other four cancers. Thus, cancer survivorship warrants immediate attention in Taiwan.

**Figure 1.**
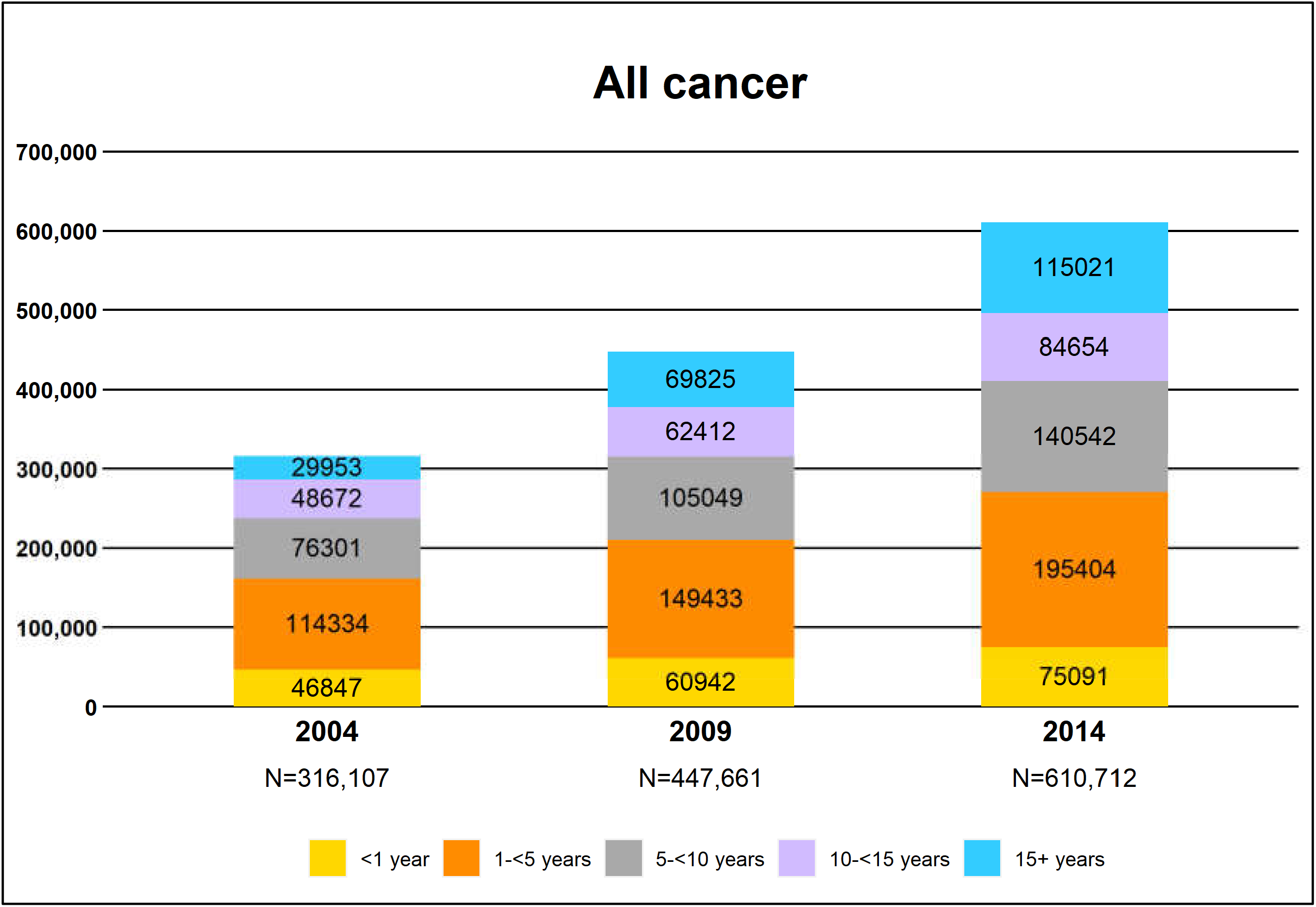
Prevalence of cancer survivors by calendar year and number of years from diagnosis: All cancer

### Prevalence of comorbidities

Table 1 reports the prevalence of the 18 comorbidities in the cancer patient cohorts and the noncancer cohort aged 65–94 years; Tables S4-1 and S4-2 show that a much higher comorbidity prevalence existed among those aged 65–94 than among those aged 15–64 years. For example, 73.3% of the breast cancer patients aged 65–94 had at least one comorbidity, while 26.5% of those aged 15-64 had at least one comorbidity. Among the old patients in Taiwan, hypertension, diabetes, ulcer disease, COPD, and CVD were the most common comorbidities, with a prevalence higher than 10%; liver disease, CHF, and CRF were the next most common comorbidities, with a prevalence 5–10%. There were differences in comorbidity prevalence compared to those in the US and UK [5, 16, 36]. For example, CHF and PVD had higher ranks in the US and UK, and liver disease and ulcer disease had higher ranks in Taiwan. In Taiwan, the noncancer and oral cancer cohorts had the fewest comorbidities; patients with liver cancers had the most comorbidities, and 25%–28% of patients with breast, colorectal and lung cancer had no comorbidities. In the US, breast cancer had a similar comorbidity prevalence to the noncancer cohort, and lung cancer had a much higher comorbidity prevalence [5]. However, COPD was most prevalent in patients with lung cancer in both the US and Taiwan.

### TCI and comorbidity levels

We used the more intuitive Cox’s model Main18&11.ND to define the TCI; Table 2 (Table S6-2) reports the weights in the Cox’s model defining the TCI. Based on the TCI and clinical judgment, we followed Cho and colleagues and Edwards and colleagues to consider three comorbidity levels [5, 7]. Patients with none of the 18 comorbidities were coded as 0. Patients were considered to have a severe comorbidity and coded as 2 if their TCIs were >0.66 or they had severe illnesses, such as COPD, liver dysfunction, chronic renal failure, dementia, or congestive heart failure, which frequently lead to organ failure or systemic dysfunction and usually require adjusting the cancer treatment. Among patients with exactly one comorbidity in the US, they were the patients with NCI index weights >0.66 [5]; this statement also held true in this study, except for those with COPD only. Patients coded as neither 0 nor 2 were coded as 1 and said to have a low/moderate comorbidity. Note that the cutoff of 0.66 was coincidentally the same as that of Edwards et al. [5].

**Table 2.**
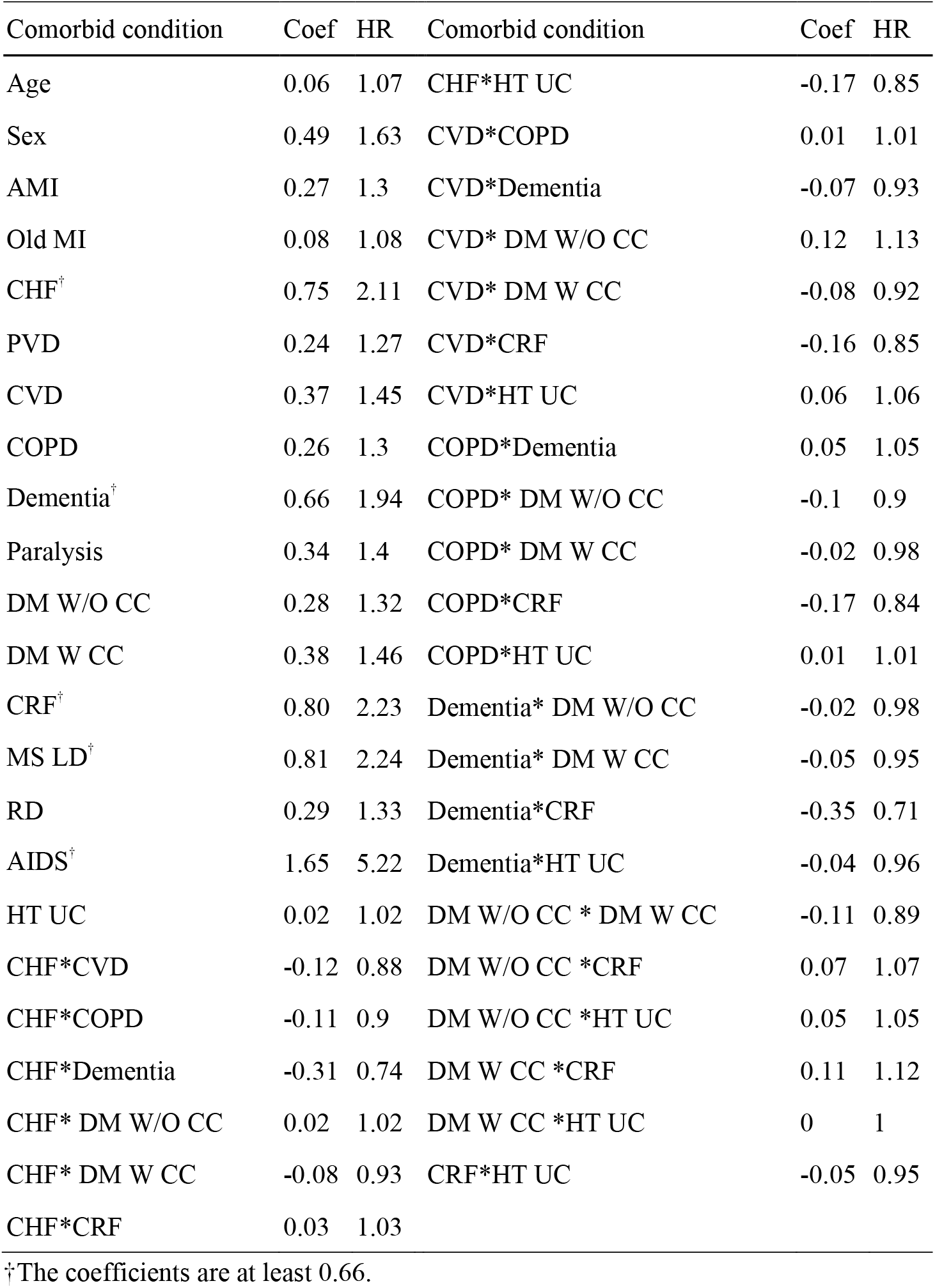
Hazard ratios and the coefficients from Negative Deleted Model Main18 & 11.ND based on the training set of Five-Cancer.

Table 3 reports the numbers and percentages of patients by stage, age, and comorbidity levels for each cancer. Here, the cancer stage follows the SEER summary stage described in Table S8, which converts the stage at diagnosis from TNM to the SEER summary stage. Tables S9–S11 provide additional information about Table 3. Table 3 shows that comorbidity prevalence increased with age; breast cancer, colorectal cancer, and liver cancer had more patients diagnosed with early stages, oral cancer had more in regional stage, and lung cancer had majority in late stage.

**Table 3.**
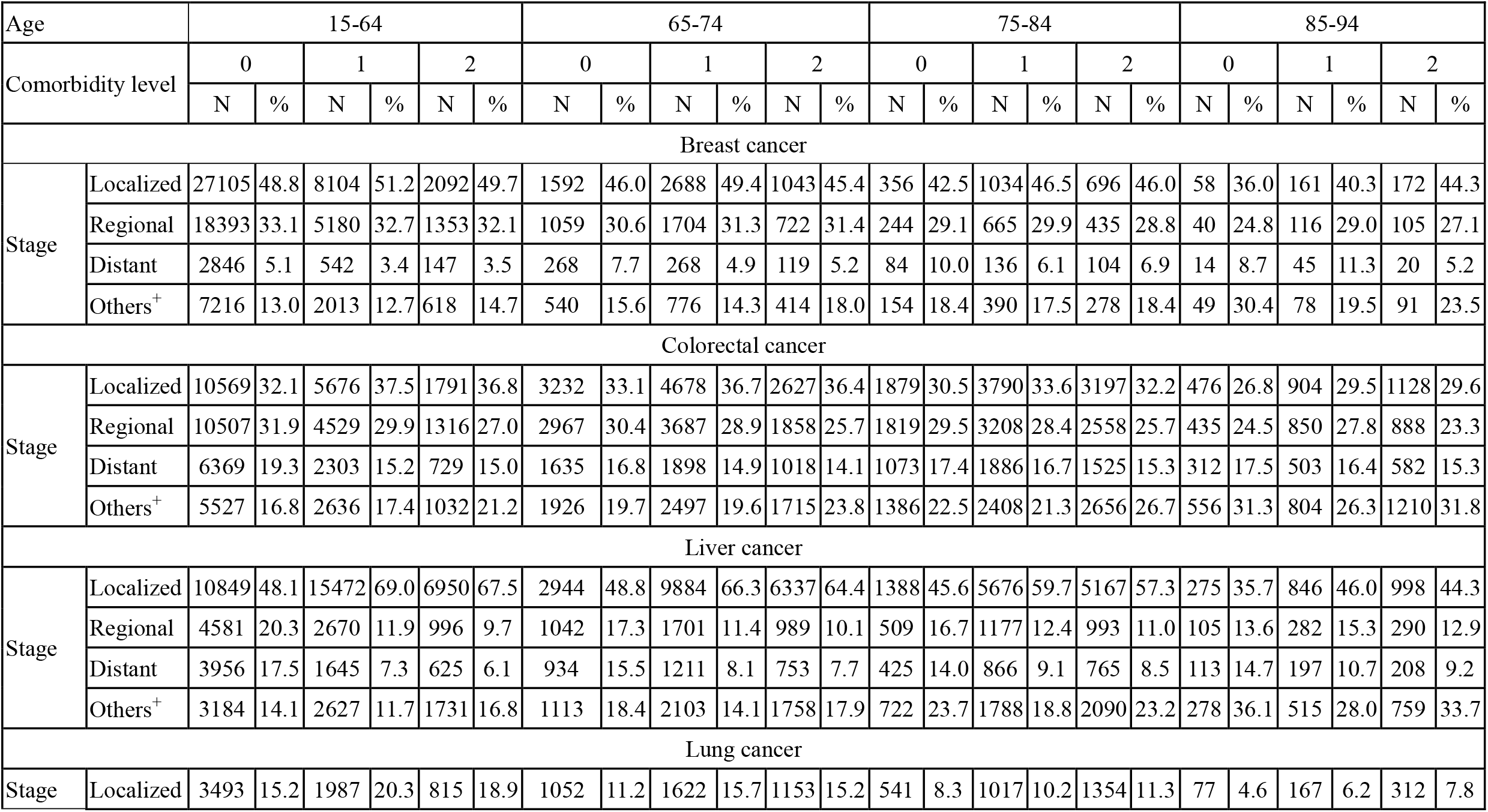
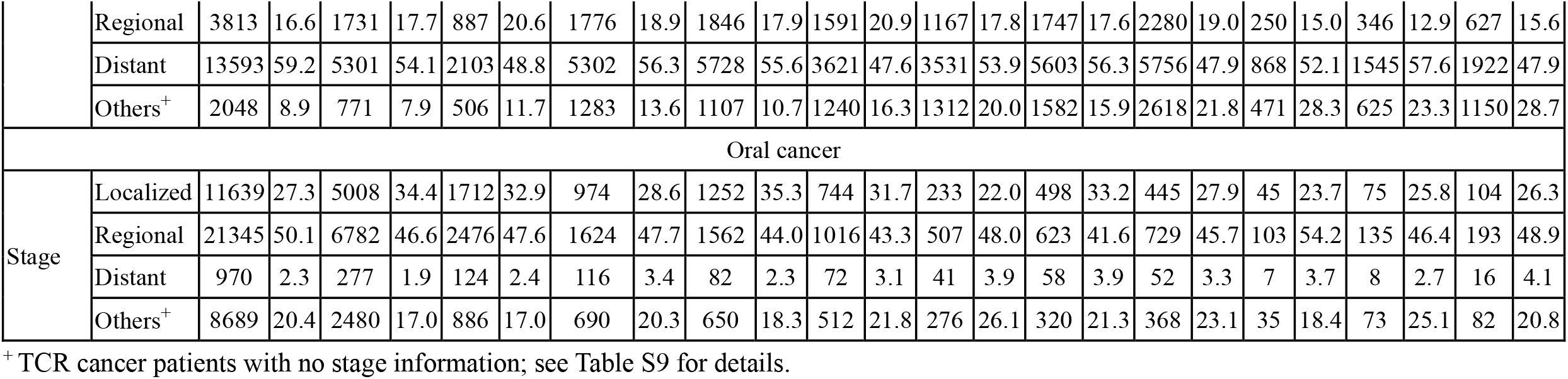
Number and percentage of cancer patients by age, stage, and comorbidity level.

| Age |  | 15-64 |  |  |  |  |  | 65-74 |  |  |  |  |  | 75-84 |  |  |  |  |  | 85-94 |  |  |  |  |  |
| --- | --- | --- | --- | --- | --- | --- | --- | --- | --- | --- | --- | --- | --- | --- | --- | --- | --- | --- | --- | --- | --- | --- | --- | --- | --- |
| Comorbidity level |  | 0 |  | 1 |  | 2 |  | 0 |  | 1 |  | 2 |  | 0 |  | 1 |  | 2 |  | 0 |  | 1 |  | 2 |  |
|  |  | N | % | N | % | N | % | N | % | N | % | N | % | N | % | N | % | N | % | N | % | N | % | N | % |
| Breast cancer |  |  |  |  |  |  |  |  |  |  |  |  |  |  |  |  |  |  |  |  |  |  |  |  |  |
| Stage | Localized | 27105 | 48.8 | 8104 | 51.2 | 2092 | 49.7 | 1592 | 46.0 | 2688 | 49.4 | 1043 | 45.4 | 356 | 42.5 | 1034 | 46.5 | 696 | 46.0 | 58 | 36.0 | 161 | 40.3 | 172 | 44.3 |
|  | Regional | 18393 | 33.1 | 5180 | 32.7 | 1353 | 32.1 | 1059 | 30.6 | 1704 | 31.3 | 722 | 31.4 | 244 | 29.1 | 665 | 29.9 | 435 | 28.8 | 40 | 24.8 | 116 | 29.0 | 105 | 27.1 |
|  | Distant | 2846 | 5.1 | 542 | 3.4 | 147 | 3.5 | 268 | 7.7 | 268 | 4.9 | 119 | 5.2 | 84 | 10.0 | 136 | 6.1 | 104 | 6.9 | 14 | 8.7 | 45 | 11.3 | 20 | 5.2 |
|  | Others<sup>+</sup> | 7216 | 13.0 | 2013 | 12.7 | 618 | 14.7 | 540 | 15.6 | 776 | 14.3 | 414 | 18.0 | 154 | 18.4 | 390 | 17.5 | 278 | 18.4 | 49 | 30.4 | 78 | 19.5 | 91 | 23.5 |
| Colorectal cancer |  |  |  |  |  |  |  |  |  |  |  |  |  |  |  |  |  |  |  |  |  |  |  |  |  |
| Stage | Localized | 10569 | 32.1 | 5676 | 37.5 | 1791 | 36.8 | 3232 | 33.1 | 4678 | 36.7 | 2627 | 36.4 | 1879 | 30.5 | 3790 | 33.6 | 3197 | 32.2 | 476 | 26.8 | 904 | 29.5 | 1128 | 29.6 |
|  | Regional | 10507 | 31.9 | 4529 | 29.9 | 1316 | 27.0 | 2967 | 30.4 | 3687 | 28.9 | 1858 | 25.7 | 1819 | 29.5 | 3208 | 28.4 | 2558 | 25.7 | 435 | 24.5 | 850 | 27.8 | 888 | 23.3 |
|  | Distant | 6369 | 19.3 | 2303 | 15.2 | 729 | 15.0 | 1635 | 16.8 | 1898 | 14.9 | 1018 | 14.1 | 1073 | 17.4 | 1886 | 16.7 | 1525 | 15.3 | 312 | 17.5 | 503 | 16.4 | 582 | 15.3 |
|  | Others<sup>+</sup> | 5527 | 16.8 | 2636 | 17.4 | 1032 | 21.2 | 1926 | 19.7 | 2497 | 19.6 | 1715 | 23.8 | 1386 | 22.5 | 2408 | 21.3 | 2656 | 26.7 | 556 | 31.3 | 804 | 26.3 | 1210 | 31.8 |
| Liver cancer |  |  |  |  |  |  |  |  |  |  |  |  |  |  |  |  |  |  |  |  |  |  |  |  |  |
| Stage | Localized | 10849 | 48.1 | 15472 | 69.0 | 6950 | 67.5 | 2944 | 48.8 | 9884 | 66.3 | 6337 | 64.4 | 1388 | 45.6 | 5676 | 59.7 | 5167 | 57.3 | 275 | 35.7 | 846 | 46.0 | 998 | 44.3 |
|  | Regional | 4581 | 20.3 | 2670 | 11.9 | 996 | 9.7 | 1042 | 17.3 | 1701 | 11.4 | 989 | 10.1 | 509 | 16.7 | 1177 | 12.4 | 993 | 11.0 | 105 | 13.6 | 282 | 15.3 | 290 | 12.9 |
|  | Distant | 3956 | 17.5 | 1645 | 7.3 | 625 | 6.1 | 934 | 15.5 | 1211 | 8.1 | 753 | 7.7 | 425 | 14.0 | 866 | 9.1 | 765 | 8.5 | 113 | 14.7 | 197 | 10.7 | 208 | 9.2 |
|  | Others<sup>+</sup> | 3184 | 14.1 | 2627 | 11.7 | 1731 | 16.8 | 1113 | 18.4 | 2103 | 14.1 | 1758 | 17.9 | 722 | 23.7 | 1788 | 18.8 | 2090 | 23.2 | 278 | 36.1 | 515 | 28.0 | 759 | 33.7 |
| Lung cancer |  |  |  |  |  |  |  |  |  |  |  |  |  |  |  |  |  |  |  |  |  |  |  |  |  |
| Stage | Localized | 3493 | 15.2 | 1987 | 20.3 | 815 | 18.9 | 1052 | 11.2 | 1622 | 15.7 | 1153 | 15.2 | 541 | 8.3 | 1017 | 10.2 | 1354 | 11.3 | 77 | 4.6 | 167 | 6.2 | 312 | 7.8 |
|  | Regional | 3813 | 16.6 | 1731 | 17.7 | 887 | 20.6 | 1776 | 18.9 | 1846 | 17.9 | 1591 | 20.9 | 1167 | 17.8 | 1747 | 17.6 | 2280 | 19.0 | 250 | 15.0 | 346 | 12.9 | 627 | 15.6 |
|  | Distant | 13593 | 59.2 | 5301 | 54.1 | 2103 | 48.8 | 5302 | 56.3 | 5728 | 55.6 | 3621 | 47.6 | 3531 | 53.9 | 5603 | 56.3 | 5756 | 47.9 | 868 | 52.1 | 1545 | 57.6 | 1922 | 47.9 |
|  | Others<sup>+</sup> | 2048 | 8.9 | 771 | 7.9 | 506 | 11.7 | 1283 | 13.6 | 1107 | 10.7 | 1240 | 16.3 | 1312 | 20.0 | 1582 | 15.9 | 2618 | 21.8 | 471 | 28.3 | 625 | 23.3 | 1150 | 28.7 |
| Oral cancer |  |  |  |  |  |  |  |  |  |  |  |  |  |  |  |  |  |  |  |  |  |  |  |  |  |
| Stage | Localized | 11639 | 27.3 | 5008 | 34.4 | 1712 | 32.9 | 974 | 28.6 | 1252 | 35.3 | 744 | 31.7 | 233 | 22.0 | 498 | 33.2 | 445 | 27.9 | 45 | 23.7 | 75 | 25.8 | 104 | 26.3 |
|  | Regional | 21345 | 50.1 | 6782 | 46.6 | 2476 | 47.6 | 1624 | 47.7 | 1562 | 44.0 | 1016 | 43.3 | 507 | 48.0 | 623 | 41.6 | 729 | 45.7 | 103 | 54.2 | 135 | 46.4 | 193 | 48.9 |
|  | Distant | 970 | 2.3 | 277 | 1.9 | 124 | 2.4 | 116 | 3.4 | 82 | 2.3 | 72 | 3.1 | 41 | 3.9 | 58 | 3.9 | 52 | 3.3 | 7 | 3.7 | 8 | 2.7 | 16 | 4.1 |
|  | Others<sup>+</sup> | 8689 | 20.4 | 2480 | 17.0 | 886 | 17.0 | 690 | 20.3 | 650 | 18.3 | 512 | 21.8 | 276 | 26.1 | 320 | 21.3 | 368 | 23.1 | 35 | 18.4 | 73 | 25.1 | 82 | 20.8 |

<sup>+</sup> TCR cancer patients with no stage information; see Table S9 for details.

### Survival measures considering competing risks of death by comorbidity level

Figure 2 presents the 5-year probabilities of dying from cancer, dying from competing causes, and survival stratified by sex, stage, age, and comorbidity level for the five cancers. Table S12 reports their actual values and the corresponding 1-year and 2-year probabilities; Figure S2 presents the corresponding figures. Strata with fewer than 100/50 patients are marked with */+.

**Figure 2.**
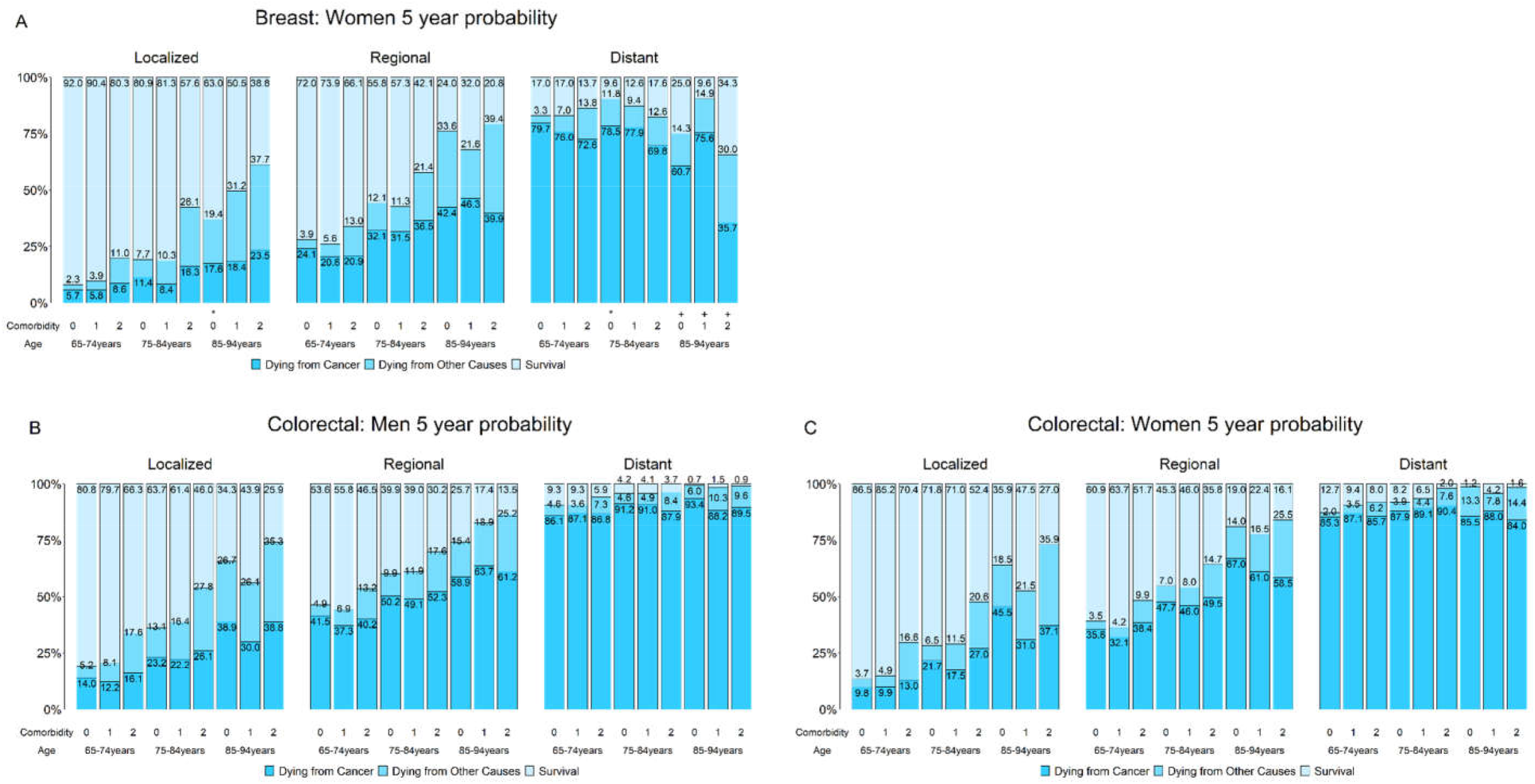
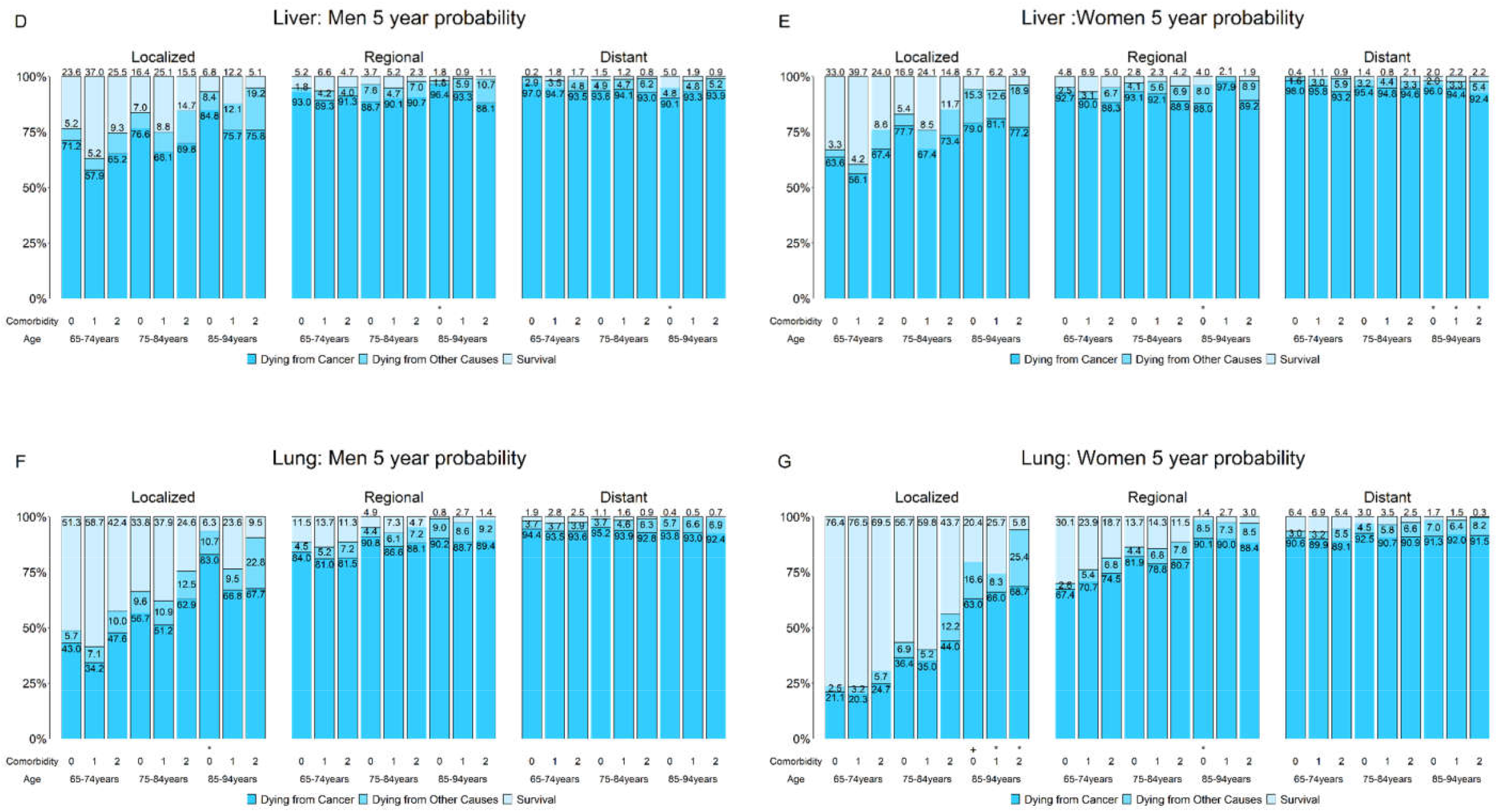
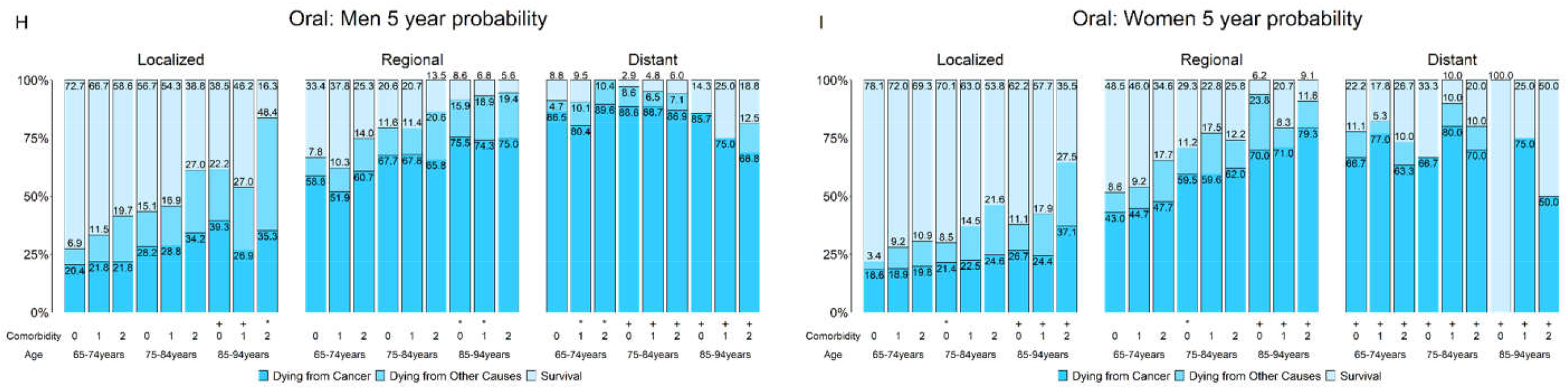
Five-year probabilities of dying from cancer, dying from other causes, and survival stratified by stage, age, comorbidity level, and sex for breast, colorectal, liver, lung, and oral cancer; strata having patients less than 100/50 are marked */+.

Among patients with localized and regional stage cancers, those with older age or severe comorbidity had lower survival rates, mainly due to increased deaths from competing causes. For patients with distant-stage cancers, age and comorbidities had a reduced effect, and the chances of dying from cancer were high. Although comorbidities affected both cancer-related and noncancer-related deaths, the effect was larger for noncancer-related deaths. Stage had a much larger effect on survival than age or comorbidity. Thus, the impact of age, comorbidity, and stage on the actual prognosis was generally similar to those reported in the US [6].

Despite these similarities, there were considerable differences between Taiwan and the US. In Taiwan, patients with local or regional breast, colorectal, and lung cancers had lower chances of dying from competing causes and higher chances of dying from the cancer, except for women with lung cancer.

Figure 2 show that patients with liver and lung cancer had the highest probabilities of cancer-related death, and their comorbidities had smaller influences on death. Figure 2 also show similar patterns for oral cancers when there were enough patients in the strata.

### Lung cancer subtypes

Because of the sample size, we considered patients aged 30–94 years with distant stages in the lung cancer subtype studies, although TCI was not evaluated for young patients. Figure 3-1 shows that the overall survival for lung adenocarcinoma (ADC) was better than that for squamous cell carcinoma (SCC) of the lung and that for small cell lung cancer (SCLC); the difference was most obvious for one-year overall survival. Figure 3-2 shows that for lung ADC, the overall survival was better in 2011–2014 than in 2004–2010, and the difference was also most obvious for one-year overall survival. Figure 3-3 shows that the one-year overall survival was best for women with lung ADC, next for men with lung ADC, and worst for men with lung SCC. Supplementary Figures S3–S4 include other prognoses.

**Figure 3-1.**
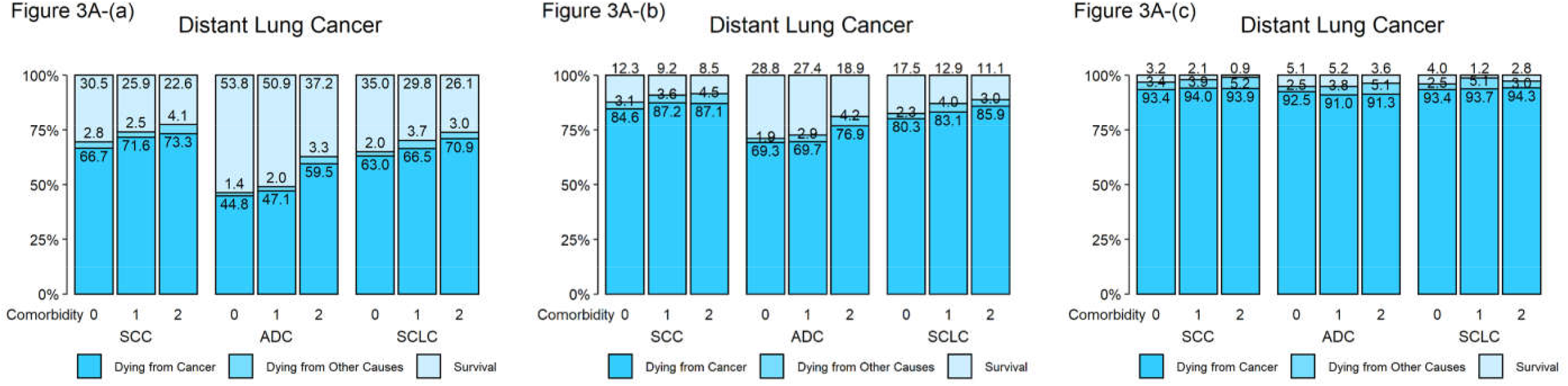
(a) One-, (b) two-, (c) five-year probabilities of dying from cancer, dying from other causes, and survival stratified by comorbidity level and subtype for distant lung cancer patients ages 30—94.

**Figure 3-2.**
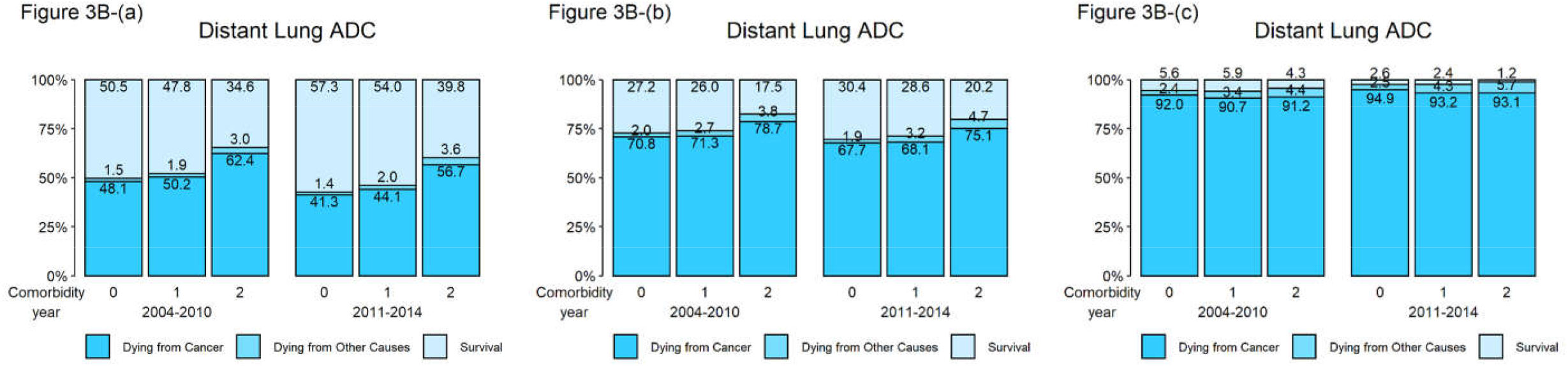
(a) One-, (b) two-, (c) five-year probabilities of dying from cancer, dying from other causes, and survival stratified by comorbidity level and year of diagnosis for distant lung ADC patients ages 30—94.

**Figure 3-3.**
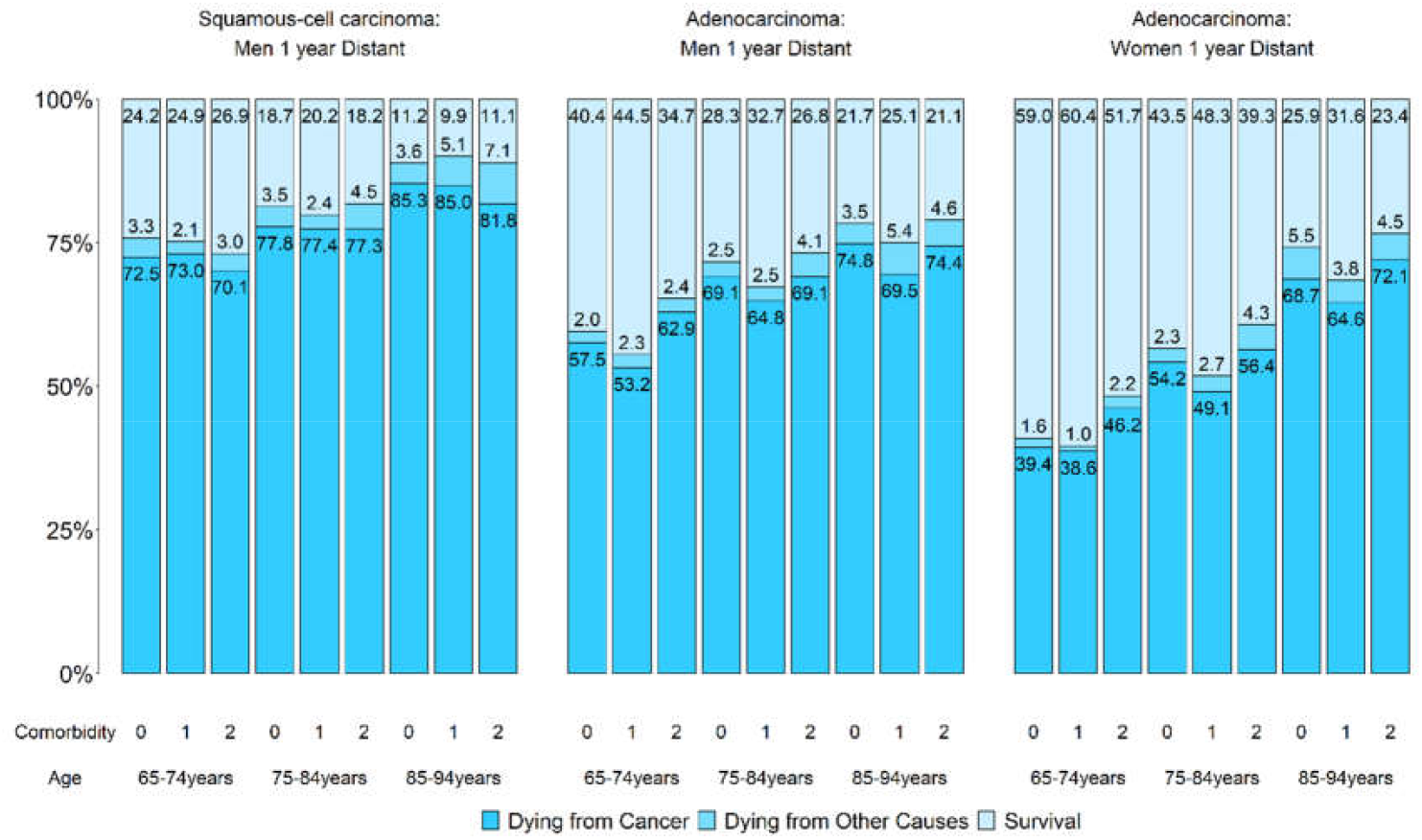
One-year probabilities of dying from cancer, dying from other causes, and survival stratified by comorbidity level, age and certain sex-subtypes for distant lung cancer.

Tables S13–S14 present the corresponding point estimates, confidence intervals, and other related statistics.

All of these findings are consistent with the 2011 Taiwan NHI Program policy that reimburses patients with late-stage lung ADC who have *EGFR* mutations for tyrosine kinase inhibitors (TKIs); *EGFR* mutations are common among never-smoking female lung ADC patients in Taiwan [37].

The above observations from the 1-year probabilities became less prominent for the 2-year probabilities and nearly vanished for the 5-year probabilities (Figures S2–S3). This may reflect the palliative nature of the TKIs.

### Model performances

Table S7 reports the AUCs regarding the 5-year survival of noncancer deaths for indices based on different models and training sets. It is interesting to see from Tables S7-1—S7-9 that the AUCs did not change much by deleting the comorbidities with negative coefficients but did decrease clearly by backward stepwise variable selection, where one of the coefficients was still negative; see Table S6-3. They also varied little with training sets. Table S7-10 reports the 5-year AUCs using the test sets of those aged 65–94: 0.73 (breast), 0.71 (male colorectal), 0.75 (female colorectal), 0.68 (male liver), 0.69 (female liver), 0.64 (male lung), 0.72 (female lung), 0.65 (male oral), and 0.71 (female oral). These AUCs for breast, colorectal and lung cancer are 2% to 9% higher than those in the US [16]. The observation that male cancer patients had smaller AUCs might be caused by more deaths due to lung cancer.

## Discussion

This study reports that in Taiwan, the number of cancer survivors increased rapidly, comorbidities among older patients with cancer were common, and the comorbidity profile among Taiwanese older patients differed from those in the US and UK. Using the three comorbidity levels defined by the TCI and clinical judgment, we reported the actual prognoses of these patients. This study could help clinicians and patients in risk communication, policymakers in resource allocation, and researchers in designing health studies.

Here are some remarks on the methodology about the TCI. Although we modified the condition of mild liver disease by adding viral hepatitis B and C and included hypertension to reflect the high prevalence of these diseases in Taiwan, all the remaining comorbidities were adopted from the CCI and NCICI. Although these comorbidities are well-established, we found some of them had negative coefficients for their main effects in the resulting Cox’s models, suggesting the existence of correlation among them. To make the model more intuitive, we considered the procedure to eliminate the comorbidities with negative coefficients. Tables S7-1—S7-9 show that this procedure help make the model more intuitive without sacrificing performance. These tables also suggest that stepwise variable selection may suffer severe disadvantages. All the above are in line with those discussed in Steyerberg [38] and Harrel [39]. Because we followed standard model development, selection, and assessment procedures strictly, the AUCs reported in Table S7-10 was based on the test sets, and the test sets were held back until the final assessment, the performance of the TCI is likely reliable. Finally, we note that in the model selection step, we chose Main18&11, instead of Main11, because the former performed better in 7 of the 9 cancers, although only slightly.

To better estimate the actual prognoses, one might need to consider other comorbid conditions, treatments, and other risk factors [10, 40-42]. It is also important to see if socioeconomic status affects the actual prognosis of cancer patients, given the comorbidity level [36].

Compared with the SEER studies, the chances of dying from competing causes are lower and those of dying from cancer are higher in Taiwan for local and regional breast, colorectal and male lung cancers. This seems to be in line with the results based on net survival. Indeed, a comparison between the 5-year cancer cause-specific survival in Taiwan during 2000—2010, based on Table 3 in Chien and colleagues [25], and that in the US SEER study during 1992— 2004, based on Table 3 in Howlader and colleagues [33], suggests that cancer survival of the breast and the colon and rectum in Taiwan seemed to be poorer than those in the US. Comparing Table 2 with Stedman et al. [16] suggests that COPD and chronic renal failure (CRF) exhibited the largest difference in hazard ratios. While the large hazard ratio for CRF might reflect the serious renal disease problem in Taiwan [43], further studies are needed to understand the low hazard ratios for COPD in Taiwan. Because tobacco smoking is an important risk factor for both lung cancer and COPD and a large proportion of lung cancer patients are never-smokers in Taiwan [37, 44], it might be worthwhile to study the prognosis of lung cancer by smoking status.

A recent study suggested that targeted therapies may have contributed to the reduced mortality from nonsmall-cell lung cancer in the US population [45, 46]. Our results on the actual prognoses for patients with distant-stage disease provide additional population-level support for the positive effects of recent advances in lung cancer treatment on patient outcomes, reflecting the 2011 reimbursement policy of the Taiwan NHI program.

Figure 2 indicate that for localized liver cancer, 5-year overall survival rates were better for those at comorbidity level 1 than for those without comorbidities. This might be related to the 2003 NHI policy that reimburses antiviral medications [47] and suggests a future study that considers the actual prognoses of patients with liver cancer separately for those with and without hepatitis viral infections.

## Conclusions

The rapid increase in long-term cancer survivors and the widespread comorbidities among older cancer patients in Taiwan demand attention on their actual prognoses. In addition to provide information for patients and clinicians regarding treatment decisions and for policymakers regarding resource allocation, this study suggested important future research topics.

## Data Availability

All the datasets used in this study were provided by and all the analyses were carried out in one of the secure labs of the Health and Welfare Data Science Center, Ministry of Health and Welfare, Taiwan. All the data are de-identified. For information on how to submit an application for gaining access to these datasets, please follow the instructions at https://www.apre.mohw.gov.tw/ If some one wants to request the data from this study, please contact the corresponding author Dr. I-Shou Chang for more detailed information.

## References

[1] Boyd CM, Darer J, Boult C, Fried LP, Boult L, Wu AW. Clinical practice guidelines and quality of care for older patients with multiple comorbid diseases: implications for pay for performance. JAMA. 2005;294:716–24.

[2] Sogaard M, Thomsen RW, Bossen KS, Sorensen HT, Norgaard M. The impact of comorbidity on cancer survival: a review. Clinical epidemiology. 2013;5:3–29.

[3] Sarfati D, Koczwara B, Jackson C. The impact of comorbidity on cancer and its treatment. CA: a cancer journal for clinicians. 2016;66:337–50.

[4] Williams GR, Mackenzie A, Magnuson A, Olin R, Chapman A, Mohile S, et al. Comorbidity in older adults with cancer. J Geriatr Oncol. 2016;7:249–57.

[5] Edwards BK, Noone AM, Mariotto AB, Simard EP, Boscoe FP, Henley SJ, et al. Annual Report to the Nation on the status of cancer, 1975-2010, featuring prevalence of comorbidity and impact on survival among persons with lung, colorectal, breast, or prostate cancer. Cancer. 2014;120:1290–314.

[6] Howlader N, Mariotto AB, Woloshin S, Schwartz LM. Providing clinicians and patients with actual prognosis: cancer in the context of competing causes of death. Journal of the National Cancer Institute Monographs. 2014;2014:255–64.

[7] Cho H, Mariotto AB, Mann BS, Klabunde CN, Feuer EJ. Assessing non-cancer-related health status of US cancer patients: other-cause survival and comorbidity prevalence. American journal of epidemiology. 2013;178:339–49.

[8] Levit LB, E. Nass, S. Ganz, P.A. Delivering High-Quality Cancer Care: Charting a New Course for a System in Crisis. Committee on Improving the Quality of Cancer Care: Addressing the Challenges of an Aging Population, Institute of Medicine. Washington, DC: The National Academies Press; 2013.

[9] Burdett N, Vincent AD, O’Callaghan M, Kichenadasse G. Competing Risks in Older Patients With Cancer: A Systematic Review of Geriatric Oncology Trials. J Natl Cancer Inst. 2018;110:825–30.

[10] Eloranta S, Smedby KE, Dickman PW, Andersson TM. Cancer survival statistics for patients and healthcare professionals - a tutorial of real-world data analysis. J Intern Med. 2021;289:12–28.

[11] Rotenstein LS, Zhang Y, Jacobson JO. Chronic Comorbidity Among Patients With Cancer: An Impetus for Oncology and Primary Care Collaboration. JAMA Oncol. 2019;5:1099–100.

[12] Chan RJ, Nekhlyudov L, Duijts SFA, Hudson SV, Jones JM, Keogh J, et al. Future research in cancer survivorship. J Cancer Surviv. 2021;15:659–67.

[13] Chan RJ, Hollingdrake O, Bui U, Nekhlyudov L, Hart NH, Lui CW, et al. Evolving landscape of cancer survivorship research: an analysis of the Journal of Cancer Survivorship, 2007-2020. J Cancer Surviv. 2021;15:651–8.

[14] Klabunde CN, Legler JM, Warren JL, Baldwin LM, Schrag D. A refined comorbidity measurement algorithm for claims-based studies of breast, prostate, colorectal, and lung cancer patients. Annals of epidemiology. 2007;17:584–90.

[15] Mariotto AB, Wang Z, Klabunde CN, Cho H, Das B, Feuer EJ. Life tables adjusted for comorbidity more accurately estimate noncancer survival for recently diagnosed cancer patients. Journal of clinical epidemiology. 2013;66:1376–85.

[16] Stedman MR, Doria-Rose, P., Warren, J.L., Klabunde, C.N., Mariotto, A. The impact of different SEER-Medicare claims-based comorbidity indexes on predicting non-cancer mortality for cancer patients. 2018.

[17] Chiang CJ, Chen YC, Chen CJ, You SL, Lai MS, Taiwan Cancer Registry Task F. Cancer trends in Taiwan. Japanese journal of clinical oncology. 2010;40:897–904.

[18] Chiang CJ, Lo WC, Yang YW, You SL, Chen CJ, Lai MS. Incidence and survival of adult cancer patients in Taiwan, 2002-2012. J Formos Med Assoc. 2016;115:1076–88.

[19] Council TND. Taiwan Statistical Data Book: Taiwan National Developement Council; 2018.

[20] Chang CM, Yin WY, Wei CK, Wu CC, Su YC, Yu CH, et al. Adjusted Age-Adjusted Charlson Comorbidity Index Score as a Risk Measure of Perioperative Mortality before Cancer Surgery. PLoS One. 2016;11:e0148076.

[21] Yang CC, Chen PC, Hsu CW, Chang SL, Lee CC. Validity of the age-adjusted charlson comorbidity index on clinical outcomes for patients with nasopharyngeal cancer post radiation treatment: a 5-year nationwide cohort study. PLoS One. 2015;10:e0117323.

[22] Hsu CL, Chen JH, Chen KY, Shih JY, Yang JC, Yu CJ, et al. Advanced non-small cell lung cancer in the elderly: the impact of age and comorbidities on treatment modalities and patient prognosis. J Geriatr Oncol. 2015;6:38–45.

[23] Chien LC, Wu YJ, Hsiung CA, Wang LH, Chang IS. Smoothed Lexis Diagrams With Applications to Lung and Breast Cancer Trends in Taiwan. J Am Stat Assoc. 2015;110:1000–12.

[24] Chien LH, Tseng TJ, Chen CH, Jiang HF, Tsai FY, Liu TW, et al. Comparison of annual percentage change in breast cancer incidence rate between Taiwan and the United States-A smoothed Lexis diagram approach. Cancer Med. 2017;6:1762–75.

[25] Chien LH, Tseng TJ, Tsai FY, Wang JH, Hsiung CA, Liu TW, et al. Patterns of age-specific socioeconomic inequalities in net survival for common cancers in Taiwan, a country with universal health coverage. Cancer epidemiology. 2018;53:42–8.

[26] Hastie T, Tibshirani, R., Friedman, J. The Elements of Statistical Learning. Second ed: Springer; 2009.

[27] Heagerty PJ, Zheng Y. Survival model predictive accuracy and ROC curves. Biometrics. 2005;61:92–105.

[28] Quan H, Li B, Couris CM, Fushimi K, Graham P, Hider P, et al. Updating and validating the Charlson comorbidity index and score for risk adjustment in hospital discharge abstracts using data from 6 countries. American journal of epidemiology. 2011;173:676–82.

[29] Maringe C, Fowler H, Rachet B, Luque-Fernandez MA. Reproducibility, reliability and validity of population-based administrative health data for the assessment of cancer non-related comorbidities. PLoS One. 2017;12:e0172814.

[30] Chiang CJ, You SL, Chen CJ, Yang YW, Lo WC, Lai MS. Quality assessment and improvement of nationwide cancer registration system in Taiwan: a review. Japanese journal of clinical oncology. 2015;45:291–6.

[31] Hsing AW, Ioannidis JP. Nationwide Population Science: Lessons From the Taiwan National Health Insurance Research Database. JAMA Intern Med. 2015;175:1527–9.

[32] Lu TH, Lee MC, Chou MC. Accuracy of cause-of-death coding in Taiwan: types of miscoding and effects on mortality statistics. International journal of epidemiology. 2000;29:336–43.

[33] Howlader N, Ries LA, Mariotto AB, Reichman ME, Ruhl J, Cronin KA. Improved estimates of cancer-specific survival rates from population-based data. J Natl Cancer Inst. 2010;102:1584–98.

[34] Charlson ME, Pompei P, Ales KL, MacKenzie CR. A new method of classifying prognostic comorbidity in longitudinal studies: development and validation. J Chronic Dis. 1987;40:373–83.

[35] Parry C, Kent EE, Mariotto AB, Alfano CM, Rowland JH. Cancer survivors: a booming population. Cancer Epidemiol Biomarkers Prev. 2011;20:1996–2005.

[36] Fowler H, Belot A, Ellis L, Maringe C, Luque-Fernandez MA, Njagi EN, et al. Comorbidity prevalence among cancer patients: a population-based cohort study of four cancers. BMC Cancer. 2020;20:2.

[37] Tseng CH, Tsuang BJ, Chiang CJ, Ku KC, Tseng JS, Yang TY, et al. The Relationship Between Air Pollution and Lung Cancer in Nonsmokers in Taiwan. Journal of thoracic oncology: official publication of the International Association for the Study of Lung Cancer. 2019;14:784–92.

[38] Steyerberg EW. Clinical Prediction Models, A Practical Approach to Development, Validation, and Updating. Second ed: Springer; 2019.

[39] Harrell FE, Jr. Regression Modeling Strategies. Second ed: Springer; 2015.

[40] Feuer EJ, Lee M, Mariotto AB, Cronin KA, Scoppa S, Penson DF, et al. The Cancer Survival Query System: making survival estimates from the Surveillance, Epidemiology, and End Results program more timely and relevant for recently diagnosed patients. Cancer. 2012;118:5652–62.

[41] Feuer EJ, Rabin BA, Zou Z, Wang Z, Xiong X, Ellis JL, et al. The Surveillance, Epidemiology, and End Results Cancer Survival Calculator SEER*CSC: validation in a managed care setting. Journal of the National Cancer Institute Monographs. 2014;2014:265–74.

[42] Wasif N, Neville M, Gray R, Cronin P, Pockaj BA. Competing Risk of Death in Elderly Patients with Newly Diagnosed Stage I Breast Cancer. Journal of the American College of Surgeons. 2019;229:30–6 e1.

[43] Tsai MH, Hsu CY, Lin MY, Yen MF, Chen HH, Chiu YH, et al. Incidence, Prevalence, and Duration of Chronic Kidney Disease in Taiwan: Results from a Community-Based Screening Program of 106,094 Individuals. Nephron. 2018;140:175–84.

[44] Luo YH, Chiu CH, Scott Kuo CH, Chou TY, Yeh YC, Hsu HS, et al. Lung Cancer in Republic of China. Journal of thoracic oncology : official publication of the International Association for the Study of Lung Cancer. 2021;16:519–27.

[45] Lewis DR, Check DP, Caporaso NE, Travis WD, Devesa SS. US lung cancer trends by histologic type. Cancer. 2014;120:2883–92.

[46] Howlader N, Forjaz G, Mooradian MJ, Meza R, Kong CY, Cronin KA, et al. The Effect of Advances in Lung-Cancer Treatment on Population Mortality. The New England journal of medicine. 2020;383:640–9.

[47] Chiang CJ, Yang YW, Chen JD, You SL, Yang HI, Lee MH, et al. Significant reduction in end-stage liver diseases burden through the national viral hepatitis therapy program in Taiwan. Hepatology. 2015;61:1154–62.

