## supplementary materials for "Prevalence of comorbidities and their impact on survival among older adults with the five most common cancers in Taiwan: A population study"

### *Comparison of intervals defining the comorbidity*

Assessing comorbid conditions using population-based administrative data requires a specification of the time interval for the assessment. To evaluate the effect of the interval for comorbidity assessment and to avoid the possibility of comorbid conditions due to cancer complications, we followed Maringe and colleagues to decide the interval for comorbidity assessment in Taiwan.(Maringe et al. 2017)

Given a cancer patient, we considered the disjoint and consecutive six-month periods from the cancer diagnosis date backward to 2000. We ordered these 6-month periods as follows; the most recent 6-month period was called period 1, the second most recent period was called period 2, …, etc. Given a comorbidity, we claim that this patient had this comorbidity in periods 2—K if there was a diagnosis from his/her impatient file in periods 2—K or if there were two diagnoses from his/her outpatient file in periods 2—K with the gap between them longer than one month.

Given any one of the 18 comorbidities, for K=5, 9, and 13, we obtained the hazard ratios of the comorbidity among the colorectal cancer patients, diagnosed between 2006 and 2014, by fitting three Cox regression models with the comorbidity as the only covariate of interest and with time from cancer diagnosis to noncancer death as the event. The first model considered all colorectal cancer patients and adjusted for age and sex; the second (third) model considered all the male (female) patients and adjusted for age only. Because the hazard ratios for K=9 often lie between those for K=5 and those for K=13, we report only the hazard ratios for K=5 and K=13 in Tables S4A and S4B with Table S4A for the age group 15—64 and Table S4B for 65—94. It follows from Table S4B that the hazard ratios from K=5 and those from K=13 were similar; hence, we decided to consider K=5 in this study. Because Table S4B also shows that the hazard ratios for diabetes with chronic complications, chronic renal failure, and moderate/severe liver disease were different between males and females, we decided to explore sex-specific comorbidity indexes in this study.

Together, Tables S4A and S4B show that sex-specific hazard ratios for younger adults were generally higher than those for older adults.

***Datasets for training, validation, and test***

In developing comorbidity indexes, we considered all the cancer patients collected in the TCR diagnosed from 2004—2014 and at age between 15 and 94. Tables S5A and S5B report the numbers and percentages of patients who had any of the 18 comorbidities and the number of patients alive before the end of 2016. Model development, selection and assessment all used this dataset. Six subsets of this dataset were considered: the dataset consisting of all the patients of any of the five cancers, referred to as Five-Cancer; Five-Cancer restricted to those ages 65—94 (Five-Cancer.65); Five-Cancer restricted to male (Five-Cancer.M); Five-Cancer.M restricted to 65—94 (Five-Cancer.M.65); Five-Cancer restricted to female (Five-Cancer.F); Five-Cancer.F restricted to 65—94 (Five-Cancer.F.65). We also considered 4 datasets of patients for each cancer of the oral, colon and rectum, liver, and lung. For example, all male oral cancer patients (Oral.M) and Oral.M restricted to ages 65—94 (Oral.M.65); similarly, Oral.F, Oral.F.65, CRC.M, CRC.M.65, CRC.F, CRC.F.65, Liver.M, Liver.M.65, Liver.F, Liver.F.65, Lung.M, Lung.M.65, Lung.F, Lung.F.65. We also considered 2 datasets of breast cancer patients: all female breast cancer patients (Breast.F), and Breast.F restricted to ages 65—94 (Breast.F.65).

We followed the suggestion from Hastie and colleagues (Hastie 2009) to divide each of the above 24 datasets randomly into 3 disjoint parts: one-half as a training set, one quarter as a validation set, and the remaining one quarter as a test set. This helps in dealing with the overfitting issue. This division was carried out for groups specified by cancer site, age at diagnosis (less than 65 or not), year of diagnosis, and gender and then merged them properly for each of the 24 datasets. The training sets were used to train the models; the validation sets were used to estimate prediction error for model selection; and the test sets were used for assessment of the generalization error of the selected models.

***Training the models***

For each of the 24 training sets, we fitted Cox's regression models with time from diagnosis to noncancer death as the outcome. Censoring events includes death due to the cancer or loss to follow-up based on the linkage of the TCR, TCOD, and NHIRD.
Because some of the training sets had few patients with HIV, we considered 17 main effects without HIV in this situation. Three sets of comorbidities were used in fitting the Cox models. The first considered only the main effects (Main17 or Main18), the second the main effects together with the interactions of the most common 6 comorbidities (Main17&6 or Main18&6)), the third the main effects together with the interactions of the most common 11 comorbidities (Main17&11 or Main18&11). In fitting the models, we adjusted for age and sex when the dataset included both male and female patients and for age only when consisting of patients of the same sex. Thus, we obtained 3 Cox's regression models for each training set. These models are summarily referred to as original models (OMs).

***More intuitive models and others***

Table S6A presents the estimated coefficients of the original model Main18&11 using the dataset Five-Cancer. Table S6A shows that some of the estimated coefficients of the comorbidities were negative, which is unintuitive. Thus, we deleted all the comorbidities with negative coefficients and fitted the model again until all the main effects were positive. The resulting model is termed Main18&11.ND and the hazard ratios are presented in Table S6B. In fact, for each original model, we obtained the corresponding model with no negative coefficient; denote them accordingly by Main17.ND, Main18.ND, Main17&6.ND, Main18&6.ND, Main17&11.ND, or Main18&11.ND. Note that we did not mind if the coefficient of an interaction term was negative and that when a comorbidity was deleted due to its negative coefficient, we deleted all the interaction terms involving this comorbidity. These models are referred to as negative deletion models (NDMs).

Based on the original models, we also applied backward variable selection to eliminate variables with p-values larger than 0.05. These models are denoted by Main17.VS, Main18.VS, Main17&6.VS, Main18&6.VS, Main17&11.VS, or Main18&11.VS. They are referred to as variable selection models (VSM). Table S6C presents the estimated coefficients of Main18&11.VS using Five-Cancer.

***Comorbidity indexes for each cancer patients cohort***

For each of the 24 training sets, we obtained 9 Cox's regression models as described in the last 3 paragraphs, where the coefficients of the comorbidities and the interaction terms provided the weights for defining the comorbidity indexes. For each of these Cox's regression models, we defined its comorbidity index for a cancer patient to be the sum of the coefficients in the model corresponding to the comorbid conditions and the interaction terms this patient had.

Considering older breast cancer patients, for example, we could use the comorbidity weights estimated in any of the 9 Cox's models trained by any of the training sets in Five-Cancer, Five-Cancer.65, Five-Cancer.F, Five-Cancer.F.65, Breast.F, or Breast.F.65. Thus, we had 54 sets of weights to consider and each of them defines a comorbidity index. Table S7A reports the AUCs, regarding 5-year survival, for these 54 indexes based on the validation set in Breast.F.65. Those based on other cancer validation sets from the older patients are shown in Table S7B through Table S7I.

***Model selection and assessment: performance of negative deletion models***

Based on the validation sets from each of Breast.F.65, CRC.M.65, CRC.F.65, Liver.M.65, Liver.F.65, Lung.M.65, Lung.F.65, Oral.M.65, Oral.F.65, Table S7 reports the AUCs using original models, negative deletion models, and variable selection models and their differences. Table S7 shows that negative deletion models perform similarly to original models and better than variable selection models, suggesting the use of NDMs to define the TCI in this study. For each of the 9 validation sets, we marked in red the largest AUC among the 18 AUCs from OM, NDM and VSM. It follows from these tables that the index defined by the Cox's model Main18&11.ND trained by Five-Cancer had a high AUC in each of the 9 validation sets. Because of this excellent performance and also because of simplicity, we chose this index in this study and referred to it as the Taiwan Comorbidity Index. The performance of the Taiwan Comorbidity Index was evaluated in the respective test sets in Table S7J.

**Comorbidity in noncancer cohort**

We constructed a cohort that represents the 2004—2014 Taiwan population without any cancer diagnosis, using the TCR, TCOD, NHIRD, and Monthly Bulletin of Interior Statistics (MBIS). The NHIRD has information only on the birth year of each beneficiary; we randomly assigned a birth month and date to each beneficiary. For each calendar year from 2004 to 2014, we took samples from the NHIRD so that the sample size was 1% of the population of that year, its age distribution was similar to that of the population of that same year, and each individual sampled in that year had no cancer diagnosis up to his/her "assigned" birth day of that year, which is referred to as the enrollment time, and was not included in the samples from any earlier year. Based on the NHIRD, we report the comorbidity of each individual at the enrollment date in the same way as we did for cancer patients; specifically, we considered the 24-months period for assessment without skipping the first 6-month period.

References

Hastie, T., Tibshirani, R.,, Friedman, J. (2009), *The Elements of Statistical Learning* (Second ed.): Springer.

Maringe, C., Fowler, H., Rachet, B., and Luque-Fernandez, M. A. (2017), "Reproducibility, reliability and validity of population-based administrative health data for the assessment of cancer non-related comorbidities," *PLoS One*, 12 (3), e0172814. DOI: 10.1371/journal.pone.0172814.

**Supplementary Tables and Figures**

**Prevalence of comorbidity and impact on survival among older persons with the five most common cancers in Taiwan: A population study**

**Authors**: Li-Hsin Chien, PhD+, Tzu-Jui Tseng, Dr.PH+, Tzu-Yu Chen, PhD, Chung-Hsing Chen, PhD, Chia-Yu Chen, MS, Fang-Yu Tsai, MS, Hsiu-Ying Ku, PhD, Shih Sheng Jiang, PhD, Chao A. Hsiung, PhD, Tsang-Wu Liu, MD*, I-Shou Chang, PhD*

Table S13A. Number of patients and probabilities of dying from cancer, dying from other-causes, and survival among patients who were diagnosed with distant lung cancer at ages 30—94 in 2004—2014 by subtypes and comorbidity level. 65
Figure S2B. Probabilities of dying from cancer, dying from other causes, and survival stratified by stage, comorbidity level, and age for **colorectal cancer**...87

Table S1. Definitions of the cancers in this study

| Cancer | ICD-9 | ICD-10 |
|---|---|---|
| All cancer | 140-199 | C00-C97 |
| Breast | 174,175 | C50 |
| Colorectal | 153,154.0-154.1,159.0 | C18-C20 |
| Lung | 162 | C33, C34 |
| Oral | 140-149 | C00-C14 |
| Liver | 155 | C22 |

In Taiwan Cancer Registry, cancer sites were supplied with the anatomical site coded to the ninth revision of the International Classification of Disease (ICD-9) and in Taiwan Cancer Registry Long Form, they were coded to the tenth revision of the International Classification of Disease (ICD-10).

Table S2. Numbers of patients diagnosed with cancer of the breast, colon and rectum, liver, lung, and oral reported in the TCR from 2004 to 2014 by gender and age of diagnosis.

| | Breast | Colorectal | Liver | Lung | Oral |
|---|---|---|---|---|---|
| All-TCR[1] | 92615 | 119315 | 113028 | 101760 | 76941 |
| All-NHIRD[2] | 92343 | 118755 | 112487 | 101244 | 76743 |
| NHIRD/TCR (%) | 99.7 | 99.5 | 99.5 | 99.5 | 99.7 |
| Gender | | | | | |
| Male | | 68391 | 79563 | 64273 | 66998 |
| Female | 92343 | 50364 | 32924 | 36971 | 9745 |
| Age | | | | | |
| 15-64 | | | | | |
| Male | | 30385 | 44505 | 21089 | 55393 |
| Female | 75609 | 22599 | 10781 | 15959 | 6999 |
| 65-94 | | | | | |
| Male | | 38006 | 35058 | 43184 | 11605 |
| Female | 16734 | 27765 | 22143 | 21012 | 2746 |

[1]All-TCR means the total number of cases in the 2004-2014 TCR ages 15-94; in this paper, only the first primary cancer of each patient was considered.
[2]All-NHIRD is the number of cases in All-TCR and linked to Taiwan NHIRD.

Table S3. Definitions of comorbidities+

| No. | Disease | ICD-9-CM |
|---|---|---|
| 1 | Acute Myocardial Infraction, AMI | 410.x |
| 2 | Old Myocardial Infraction, Old MI | 412.x |
| 3 | Congestive heart failure, CHF | 428.x |
| 4 | Peripheral vascular disease, PVD | 441.x,443.9,785.4,V43.4, Procedure 38.48 |
| 5 | Cerebrovascular disease, CVD | 430.x-438.x |
| 6 | Chronic obstructive pulmonary disease, COPD | 490.x-505x,506.4 |
| 7 | Dementia | 290.x |
| 8 | Paralysis | 342.x,344.1 |
| 9 | Diabetes without chronic complication, DM W/O CC | 250.0-250.3,250.7 |
| 10 | Diabetes with chronic complication, DM W CC | 250.4-250.6,250.8-250.9 |
| 11 | Chronic renal failure (renal disease), CRF | 582.x, 583.x, 585.x, 586.x, 588.x |
| 12* | Cirrhosis/chronic hepatitis (Mild liver disease), HBV, HCV, Mild LD | 571.2,571.4,571.5,571.6 070.2-070.7(excluded 070.41,070.51),070.9, V026.1 |
| 13 | Moderate-Server Liver Disease, M S LD | 572.2-572.8,456.0-456.21 |
| 14 | Ulcer (Peptic ulcer disease) | 531.x-534.x |
| 15 | Rheumatic disease, RD | 710.0,710.1,710.4,714.0-714.2,714.81, 725.x |
| 16 | AIDS | 042.x-044x |
| 17** | Hypertension, uncomplicated, HT UC | 401.1, 401.9, 642.0 |
| 18** | Hypertension, complicated, HT C | 401.0, 402.x-405.x, 642.1, 642.2, 642.7, 642.9 |

+Deyo RA, Cherkin DC, Ciol MA. Adapting a clinical comorbidity index for use with ICD-8-CM administrative databases. J Clin Epidemiol.1992; 45:613-9
Quan H, Sundararajan V, Halfon P, et al. Coding algorithms for defining Comorbidities in ICD-9-CM and ICD-10 administrative data. Med Care. 2005 Nov; 43(11): 1130-9.
*The definition of the Cirrhosis/chronic hepatitis (Mild liver disease) comorbidity in this study includes viral hepatitis B and viral hepatitis C, which differs from the one that used by Charlson in 1987.
**Elixhauser AHRQ-Web ICD-9-CM

Table S4A. The hazard ratio of each comorbidity with respect to death due to other causes, among colorectal cancer patients, ages 15-64.

| | Age 15-64 | | | | | | | | | | | | | |
|---|---|---|---|---|---|---|---|---|---|---|---|---|---|---|
| | | | | | | | | K=5 | | | | | | |
| | K+=5 | | | K=13 | | | | Male | | | Female | | | |
| Comorbidity | HR* | 95CI | | HR | 95CI | | Comparison# | HR | 95CI | | HR | 95CI | | Comparison |
| Condition1 | 2.37 | 1.37 | 4.10 | 2.18 | 1.43 | 3.33 | 4 | 1.73 | 0.90 | 3.34 | 15.14 | 5.64 | 40.65 | 1 |
| Condition2 | 2.71 | 1.60 | 4.59 | 2.24 | 1.39 | 3.62 | 4 | 2.28 | 1.26 | 4.14 | 9.53 | 3.05 | 29.73 | 2 |
| Condition3 | 5.16 | 4.07 | 6.53 | 5.01 | 4.14 | 6.07 | 4 | 4.34 | 3.22 | 5.84 | 7.64 | 5.15 | 11.33 | 2 |
| Condition4 | 3.23 | 1.97 | 5.29 | 2.96 | 2.11 | 4.14 | 4 | 2.75 | 1.48 | 5.14 | 4.59 | 2.05 | 10.28 | 4 |
| Condition5 | 2.99 | 2.49 | 3.59 | 2.82 | 2.42 | 3.30 | 4 | 2.83 | 2.29 | 3.50 | 3.58 | 2.50 | 5.13 | 3 |
| Condition6 | 1.63 | 1.33 | 1.98 | 1.39 | 1.20 | 1.62 | 3 | 1.70 | 1.35 | 2.14 | 1.45 | 0.99 | 2.13 | 4 |
| Condition7 | 2.65 | 0.37 | 18.86 | 1.19 | 0.17 | 8.48 | 4 | 3.79 | 0.53 | 26.94 | 0.00 | 0.00 | Inf | 3 |
| Condition8 | 4.47 | 2.77 | 7.22 | 3.92 | 2.80 | 5.49 | 4 | 4.89 | 2.94 | 8.15 | 2.74 | 0.68 | 11.00 | 3 |
| Condition9 | 2.33 | 2.06 | 2.63 | 2.37 | 2.11 | 2.66 | 4 | 2.08 | 1.80 | 2.40 | 3.08 | 2.46 | 3.84 | 1 |
| Condition10 | 3.36 | 2.84 | 3.98 | 3.00 | 2.59 | 3.48 | 4 | 2.89 | 2.34 | 3.55 | 4.79 | 3.58 | 6.40 | 1 |
| Condition11 | 5.07 | 4.22 | 6.09 | 4.16 | 3.51 | 4.93 | 2 | 4.34 | 3.46 | 5.44 | 7.29 | 5.33 | 9.98 | 2 |
| Condition12 | 1.74 | 1.49 | 2.04 | 1.62 | 1.42 | 1.84 | 4 | 1.73 | 1.44 | 2.08 | 1.75 | 1.27 | 2.40 | 4 |
| Condition13 | 12.76 | 8.77 | 18.56 | 11.03 | 7.92 | 15.38 | 4 | 14.36 | 9.86 | 20.92 | 0.00 | 0.00 | Inf | 3 |
| Condition14 | 1.56 | 1.33 | 1.83 | 1.41 | 1.24 | 1.60 | 4 | 1.68 | 1.40 | 2.02 | 1.27 | 0.92 | 1.75 | 3 |
| Condition15 | 2.64 | 1.61 | 4.32 | 2.10 | 1.41 | 3.13 | 4 | 1.46 | 0.47 | 4.54 | 3.24 | 1.86 | 5.63 | 3 |
| Condition16 | 6.23 | 1.56 | 24.93 | 5.19 | 1.30 | 20.77 | 4 | 7.77 | 1.94 | 31.12 | 0.00 | 0.00 | Inf | 3 |
| Condition17 | 1.47 | 1.31 | 1.65 | 1.70 | 1.52 | 1.89 | 2 | 1.41 | 1.23 | 1.62 | 1.63 | 1.31 | 2.03 | 3 |
| Condition18 | 2.22 | 1.92 | 2.56 | 2.08 | 1.83 | 2.36 | 4 | 1.99 | 1.67 | 2.36 | 2.85 | 2.22 | 3.67 | 2 |

*Obtained by fitting a Cox's regression model with only one comorbidity as the covariate of interest, adjusted for age and sex.

#1 if there was no overlap between the two confidence intervals; 2 if there was overlap but neither of the two point estimates was in the confidence interval of the other; 3 if exactly one of the point estimates was in the confidence interval of the other; 4 if each point estimate was in the confidence interval of the other.

+The number of half-years used in assessing the comorbidities.

Table S4B. The hazard ratio of each comorbidity with respect to death due to other causes, among colorectal cancer patients, ages 65-94

| | Age 65-94 | | | | | | | | | | | | | |
|---|---|---|---|---|---|---|---|---|---|---|---|---|---|---|
| | | | | | | | | K=5 | | | | | | |
| | K[+]=5 | | | K=13 | | | | Male | | | Female | | | |
| Comorbidity | HR* | 95CI | | HR | 95CI | | Comparison[#] | HR | 95CI | | HR | 95CI | | Comparison[#] |
| Condition1 | 2.16 | 1.82 | 2.58 | 1.99 | 1.76 | 2.25 | 4 | 2.05 | 1.66 | 2.52 | 2.50 | 1.81 | 3.46 | 4 |
| Condition2 | 1.56 | 1.27 | 1.93 | 1.67 | 1.43 | 1.96 | 4 | 1.64 | 1.29 | 2.09 | 1.36 | 0.89 | 2.07 | 4 |
| Condition3 | 2.23 | 2.07 | 2.39 | 2.10 | 1.97 | 2.23 | 3 | 2.22 | 2.02 | 2.44 | 2.22 | 1.99 | 2.47 | 4 |
| Condition4 | 1.65 | 1.43 | 1.91 | 1.61 | 1.45 | 1.78 | 4 | 1.45 | 1.19 | 1.75 | 2.09 | 1.66 | 2.62 | 2 |
| Condition5 | 1.80 | 1.70 | 1.90 | 1.74 | 1.66 | 1.83 | 4 | 1.73 | 1.62 | 1.86 | 1.92 | 1.76 | 2.10 | 2 |
| Condition6 | 1.47 | 1.39 | 1.55 | 1.37 | 1.30 | 1.43 | 2 | 1.54 | 1.44 | 1.64 | 1.36 | 1.24 | 1.50 | 2 |
| Condition7 | 2.24 | 2.05 | 2.44 | 2.15 | 1.98 | 2.32 | 4 | 2.35 | 2.09 | 2.65 | 2.08 | 1.83 | 2.36 | 3 |
| Condition8 | 2.23 | 1.86 | 2.68 | 2.55 | 2.26 | 2.89 | 3 | 2.14 | 1.70 | 2.70 | 2.40 | 1.79 | 3.23 | 4 |
| Condition9 | 1.73 | 1.64 | 1.81 | 1.73 | 1.65 | 1.81 | 4 | 1.66 | 1.56 | 1.77 | 1.84 | 1.70 | 1.99 | 2 |
| Condition10 | 2.07 | 1.93 | 2.21 | 2.08 | 1.96 | 2.20 | 4 | 1.89 | 1.72 | 2.07 | 2.35 | 2.12 | 2.61 | 1 |
| Condition11 | 2.60 | 2.42 | 2.79 | 2.16 | 2.02 | 2.29 | 1 | 2.26 | 2.07 | 2.47 | 3.44 | 3.07 | 3.87 | 1 |
| Condition12 | 1.25 | 1.15 | 1.37 | 1.14 | 1.06 | 1.22 | 2 | 1.16 | 1.03 | 1.30 | 1.41 | 1.23 | 1.63 | 2 |
| Condition13 | 5.93 | 4.50 | 7.81 | 3.88 | 3.08 | 4.87 | 2 | 4.12 | 2.78 | 6.11 | 10.19 | 6.92 | 15.02 | 1 |
| Condition14 | 1.26 | 1.19 | 1.33 | 1.19 | 1.14 | 1.25 | 3 | 1.30 | 1.21 | 1.39 | 1.20 | 1.09 | 1.31 | 3 |
| Condition15 | 1.48 | 1.19 | 1.83 | 1.30 | 1.11 | 1.52 | 4 | 1.40 | 1.01 | 1.93 | 1.57 | 1.17 | 2.10 | 4 |
| Condition16 | 1.63 | 0.23 | 11.55 | 1.54 | 0.22 | 10.94 | 4 | 1.67 | 0.23 | 11.83 | - | - | - | - |
| Condition17 | 1.26 | 1.21 | 1.32 | 1.38 | 1.31 | 1.44 | 2 | 1.28 | 1.21 | 1.36 | 1.23 | 1.14 | 1.32 | 4 |
| Condition18 | 1.30 | 1.24 | 1.37 | 1.38 | 1.32 | 1.45 | 2 | 1.25 | 1.17 | 1.33 | 1.40 | 1.29 | 1.51 | 2 |

[*]Obtained by fitting a Cox's regression model with only one comorbidity as the covariate of interest, adjusted for age and sex.

[#]1 if there was no overlap between the two confidence intervals; 2 if there was overlap but neither of the two point estimates was in the confidence interval of the other; 3 if exactly one of the point estimates was in the confidence interval of the other; 4 if each point estimate was in the confidence interval of the other.

[+]The number of half-years used in assessing the comorbidities.

Table S5A. The number and percentage of patients with comorbid conditions for each of the 5 cancer cohorts and non-cancer cohorts, ages 15-64

| Ages 15-64 | **Breast** | | **Colorectal** | | | | **Liver** | | | | **Lung** | | | | **Oral** | | | | **Noncancer cohort** | | | |
|---|---|---|---|---|---|---|---|---|---|---|---|---|---|---|---|---|---|---|---|---|---|---|
| | Female | | Male | | Female | | Male | | Female | | Male | | Female | | Male | | Female | | Male | | Female | |
| | N | % | N | % | N | % | N | % | N | % | N | % | N | % | N | % | N | % | N | % | N | % |
| **Number Alive*** | 63551 | 84.1 | 19069 | 62.8 | 14947 | 66.1 | 11528 | 25.9 | 3444 | 32.0 | 4201 | 19.9 | 5229 | 32.8 | 30012 | 54.2 | 5137 | 73.4 | NA | NA | NA | NA |
| **Cancer Deaths** | 10729 | 14.2 | 10057 | 33.1 | 7114 | 31.5 | 31294 | 70.3 | 6879 | 63.8 | 16191 | 76.8 | 10407 | 65.2 | 22299 | 40.3 | 1619 | 23.1 | NA | NA | NA | NA |
| **Other-Cause Deaths** | 1329 | 1.8 | 1259 | 4.1 | 538 | 2.4 | 1683 | 3.8 | 458 | 4.3 | 697 | 3.3 | 323 | 2.0 | 3082 | 5.6 | 243 | 3.5 | NA | NA | NA | NA |
| **Number of Comorbid conditions (%)** | | | | | | | | | | | | | | | | | | | | | | |
| **Comorbid conditions (%)** | N | % | N | % | N | % | N | % | N | % | N | % | N | % | N | % | N | % | N | % | N | % |
| 0 | 55560 | 73.5 | 18146 | 59.7 | 14826 | 65.6 | 19057 | 42.8 | 3513 | 32.6 | 12852 | 60.9 | 10095 | 63.3 | 37733 | 68.1 | 4910 | 70.2 | 785729 | 84.3 | 798789 | 86.7 |
| 1 | 12363 | 16.4 | 6179 | 20.3 | 4358 | 19.3 | 9185 | 20.6 | 2459 | 22.8 | 4094 | 19.4 | 3362 | 21.1 | 8699 | 15.7 | 1182 | 16.9 | 90586 | 9.7 | 80235 | 8.7 |
| 2 | 5050 | 6.7 | 3585 | 11.8 | 2079 | 9.2 | 7611 | 17.1 | 2156 | 20.0 | 2326 | 11.0 | 1535 | 9.6 | 5144 | 9.3 | 579 | 8.3 | 35578 | 3.8 | 27788 | 3.0 |
| 3 | 1726 | 2.3 | 1484 | 4.9 | 838 | 3.7 | 4787 | 10.8 | 1389 | 12.9 | 1130 | 5.4 | 632 | 4.0 | 2371 | 4.3 | 213 | 3.0 | 12988 | 1.4 | 9705 | 1.1 |
| 4+ | 910 | 1.2 | 991 | 3.3 | 498 | 2.2 | 3865 | 8.7 | 1264 | 11.7 | 687 | 3.3 | 335 | 2.1 | 1446 | 2.6 | 115 | 1.6 | 6986 | 0.7 | 4896 | 0.5 |
| **Comorbid Conditions** | | | | | | | | | | | | | | | | | | | | | | |
| **AMI** | 44 | 0.06 | 131 | 0.43 | 17 | 0.08 | 138 | 0.31 | 7 | 0.06 | 104 | 0.49 | 16 | 0.1 | 165 | 0.30 | <5 | NA | 1469 | 0.16 | 266 | 0.03 |
| **OLD MI** | 27 | 0.04 | 110 | 0.36 | 19 | 0.08 | 91 | 0.2 | 7 | 0.06 | 92 | 0.44 | 9 | 0.06 | 129 | 0.23 | <5 | NA | 986 | 0.11 | 162 | 0.02 |
| **CHF** | 296 | 0.39 | 347 | 1.14 | 173 | 0.77 | 500 | 1.12 | 194 | 1.8 | 243 | 1.15 | 103 | 0.65 | 470 | 0.85 | 29 | 0.41 | 2968 | 0.32 | 1886 | 0.20 |
| **PVD** | 208 | 0.28 | 115 | 0.38 | 73 | 0.32 | 244 | 0.55 | 65 | 0.6 | 108 | 0.51 | 73 | 0.46 | 196 | 0.35 | 28 | 0.4 | 1412 | 0.15 | 1264 | 0.14 |
| **CVD** | 928 | 1.23 | 1071 | 3.52 | 486 | 2.15 | 1400 | 3.15 | 344 | 3.19 | 736 | 3.49 | 335 | 2.1 | 1581 | 2.85 | 123 | 1.76 | 9963 | 1.07 | 6062 | 0.66 |
| **COPD** | 2585 | 3.42 | 1297 | 4.27 | 954 | 4.22 | 2213 | 4.97 | 682 | 6.33 | 1702 | 8.07 | 1098 | 6.88 | 2057 | 3.71 | 322 | 4.6 | 18761 | 2.01 | 18842 | 2.04 |
| **Dementia** | 11 | 0.01 | 20 | 0.07 | <5 | NA | 23 | 0.05 | 5 | 0.05 | 15 | 0.07 | <5 | NA | 16 | 0.03 | 0 | 0 | 131 | 0.01 | 73 | 0.01 |
| **Paralysis** | 59 | 0.08 | 106 | 0.35 | 40 | 0.18 | 151 | 0.34 | 24 | 0.22 | 83 | 0.39 | 20 | 0.13 | 202 | 0.36 | 12 | 0.17 | 1326 | 0.14 | 489 | 0.05 |
| **DM W/O CC** | 4322 | 5.72 | 3847 | 12.66 | 2226 | 9.85 | 8176 | 18.37 | 2197 | 20.38 | 2270 | 10.76 | 1289 | 8.08 | 5330 | 9.62 | 520 | 7.43 | 31686 | 3.40 | 24493 | 2.66 |
| **DM W CC** | 1160 | 1.53 | 1119 | 3.68 | 621 | 2.75 | 2408 | 5.41 | 767 | 7.11 | 660 | 3.13 | 338 | 2.12 | 1493 | 2.7 | 140 | 2.00 | 8510 | 0.91 | 6383 | 0.69 |
| **CRF** | 729 | 0.96 | 648 | 2.13 | 362 | 1.60 | 1464 | 3.29 | 460 | 4.27 | 404 | 1.92 | 179 | 1.12 | 714 | 1.29 | 162 | 2.31 | 5217 | 0.56 | 3897 | 0.42 |
| **Mild LD** | 3512 | 4.64 | 2308 | 7.6 | 1188 | 5.26 | 17960 | 40.36 | 5182 | 48.07 | 1619 | 7.68 | 1023 | 6.41 | 4713 | 8.51 | 365 | 5.22 | 36265 | 3.89 | 21749 | 2.36 |

| | | | | | | | | | | | | | | | | | | | | | | |
|---|---|---|---|---|---|---|---|---|---|---|---|---|---|---|---|---|---|---|---|---|---|---|
| **M S LD** | 34 | 0.04 | 98 | 0.32 | 11 | 0.05 | 3091 | 6.95 | 762 | 7.07 | 74 | 0.35 | 11 | 0.07 | 615 | 1.11 | 11 | 0.16 | 1130 | 0.12 | 228 | 0.02 |
| **Ulcer** | 4513 | 5.97 | 2234 | 7.35 | 1608 | 7.12 | 6978 | 15.68 | 2291 | 21.25 | 1751 | 8.3 | 1414 | 8.86 | 3967 | 7.16 | 444 | 6.34 | 30792 | 3.3 | 30716 | 3.33 |
| **RD** | 628 | 0.83 | 63 | 0.21 | 219 | 0.97 | 134 | 0.3 | 171 | 1.59 | 83 | 0.39 | 211 | 1.32 | 150 | 0.27 | 85 | 1.21 | 1157 | 0.12 | 4244 | 0.46 |
| **AIDS** | <5** | NA | 15 | 0.05 | <5 | NA | 53 | 0.12 | <5 | NA | 18 | 0.09 | <5 | NA | 53 | 0.1 | 0 | 0 | 1155 | 0.12 | 101 | 0.01 |
| **HT UC** | 9303 | 12.3 | 6523 | 21.47 | 3874 | 17.14 | 8636 | 19.4 | 2666 | 24.73 | 3902 | 18.5 | 2775 | 17.39 | 8198 | 14.8 | 1033 | 14.76 | 59287 | 6.36 | 49400 | 5.36 |
| **HT C** | 3323 | 4.39 | 2286 | 7.52 | 1463 | 6.47 | 3071 | 6.9 | 1028 | 9.54 | 1402 | 6.65 | 938 | 5.88 | 2657 | 4.8 | 330 | 4.71 | 20263 | 2.17 | 16833 | 1.83 |

* Death due to the cancer or other causes was decided by the NCI classification algorithm using the TCOD and TCR from 2004 until Dec. 31, 2016; survival information of patients not included in the TCOD were obtained from the beneficiary registry of NHIRD at Dec. 31, 2015; the latter were all considered alive.

**The percentage is calculated by deleting the cells whose case number <5.

Table S5B. The number and percentage of patients with comorbid conditions for each of the 5 cancer cohorts and non-cancer cohorts, ages 65-94

| Age:65-94 | Breast | | Colorectal | | | | Liver | | | | Lung | | | | Oral | | | | Noncancer cohort | | | |
|---|---|---|---|---|---|---|---|---|---|---|---|---|---|---|---|---|---|---|---|---|---|---|
| | Female | | Male | | Female | | Male | | Female | | Male | | Female | | Male | | Female | | Male | | Female | |
| | N | % | N | % | N | % | N | % | N | % | N | % | N | % | N | % | N | % | N | % | N | % |
| **Number Alive**** | 11231 | 67.1 | 15083 | 39.7 | 12156 | 43.8 | 5638 | 16.1 | 3778 | 17.1 | 3370 | 7.8 | 3389 | 16.1 | 3867 | 33.3 | 1155 | 42.1 | NA | NA | NA | NA |
| **Cancer Deaths** | 3454 | 20.6 | 16801 | 44.2 | 11922 | 42.9 | 26484 | 75.5 | 16680 | 75.3 | 36896 | 85.4 | 16297 | 77.6 | 5860 | 50.5 | 1183 | 43.1 | NA | NA | NA | NA |
| **Other-Cause Deaths** | 2049 | 12.2 | 6122 | 16.1 | 3687 | 13.3 | 2936 | 8.4 | 1685 | 7.6 | 2918 | 6.8 | 1326 | 6.3 | 1878 | 16.2 | 408 | 14.9 | NA | NA | NA | NA |
| **Number of Comorbid conditions (%)** | | | | | | | | | | | | | | | | | | | | | | |
| **Comorbid conditions (%)** | N | % | N | % | N | % | N | % | N | % | N | % | N | % | N | % | N | % | N | % | N | % |
| 0 | 4466 | 26.7 | 10664 | 28.1 | 7032 | 25.3 | 6986 | 19.9 | 2862 | 12.9 | 12241 | 28.3 | 5392 | 25.7 | 3957 | 34.1 | 707 | 25.7 | 45773 | 35.4 | 44662 | 32.1 |
| 1 | 4405 | 26.3 | 9246 | 24.3 | 6995 | 25.2 | 6979 | 19.9 | 4109 | 18.6 | 10277 | 23.8 | 5490 | 26.1 | 2671 | 23 | 739 | 26.9 | 28781 | 22.3 | 33589 | 24.2 |
| 2 | 3902 | 23.3 | 8304 | 21.8 | 6250 | 22.5 | 8004 | 22.8 | 5223 | 23.6 | 9304 | 21.5 | 4788 | 22.8 | 2303 | 19.8 | 604 | 22 | 24380 | 18.9 | 28437 | 20.5 |
| 3 | 2177 | 13 | 4966 | 13.1 | 3839 | 13.8 | 6143 | 17.5 | 4362 | 19.7 | 5721 | 13.2 | 2818 | 13.4 | 1391 | 12 | 379 | 13.8 | 15186 | 11.7 | 16508 | 11.9 |
| 4+ | 1784 | 10.7 | 4826 | 12.7 | 3649 | 13.1 | 6946 | 19.8 | 5587 | 25.2 | 5641 | 13.1 | 2524 | 12 | 1283 | 11.1 | 317 | 11.5 | 15212 | 11.8 | 15851 | 11.4 |
| **Comorbid Conditions** | | | | | | | | | | | | | | | | | | | | | | |
| **AMI** | 58 | 0.35 | 400 | 1.05 | 167 | 0.60 | 277 | 0.79 | 112 | 0.51 | 446 | 1.03 | 132 | 0.63 | 85 | 0.73 | 13 | 0.47 | 1386 | 1.07 | 813 | 0.58 |
| **OLD MI** | 48 | 0.29 | 345 | 0.91 | 138 | 0.50 | 290 | 0.83 | 84 | 0.38 | 512 | 1.19 | 99 | 0.47 | 94 | 0.81 | 15 | 0.55 | 1248 | 0.96 | 545 | 0.39 |
| **CHF** | 689 | 4.12 | 2004 | 5.27 | 1776 | 6.40 | 1785 | 5.09 | 1573 | 7.10 | 2502 | 5.79 | 1180 | 5.62 | 464 | 4.00 | 143 | 5.21 | 6477 | 5.01 | 7189 | 5.17 |
| **PVD** | 213 | 1.27 | 594 | 1.56 | 368 | 1.33 | 539 | 1.54 | 334 | 1.51 | 749 | 1.73 | 304 | 1.45 | 176 | 1.52 | 32 | 1.17 | 1903 | 1.47 | 1896 | 1.36 |
| **CVD** | 1909 | 11.41 | 5699 | 15.00 | 3661 | 13.19 | 4729 | 13.49 | 2612 | 11.80 | 6542 | 15.15 | 2617 | 12.45 | 1550 | 13.36 | 315 | 11.47 | 18669 | 14.43 | 16942 | 12.18 |
| **COPD** | 1758 | 10.51 | 7056 | 18.57 | 3498 | 12.60 | 6574 | 18.75 | 3077 | 13.90 | 12483 | 28.91 | 3562 | 16.95 | 2108 | 18.16 | 392 | 14.28 | 22515 | 17.41 | 16181 | 11.64 |
| **Dementia** | 558 | 3.33 | 1246 | 3.28 | 1287 | 4.64 | 967 | 2.76 | 898 | 4.06 | 1356 | 3.14 | 850 | 4.05 | 269 | 2.32 | 95 | 3.46 | 4661 | 3.60 | 5902 | 4.24 |
| **Paralysis** | 116 | 0.69 | 365 | 0.96 | 238 | 0.86 | 298 | 0.85 | 142 | 0.64 | 390 | 0.90 | 156 | 0.74 | 125 | 1.08 | 21 | 0.76 | 1342 | 1.04 | 1114 | 0.80 |

| | | | | | | | | | | | | | | | | | | | | | | |
|---|---|---|---|---|---|---|---|---|---|---|---|---|---|---|---|---|---|---|---|---|---|---|
| **DM W/O CC** | 4211 | 25.16 | 8092 | 21.29 | 6988 | 25.17 | 9323 | 26.59 | 7159 | 32.33 | 7403 | 17.14 | 4448 | 21.17 | 2311 | 19.91 | 682 | 24.84 | 22686 | 17.54 | 29347 | 21.11 |
| **DM W CC** | 1399 | 8.36 | 2744 | 7.22 | 2301 | 8.29 | 3224 | 9.20 | 2510 | 11.34 | 2495 | 5.78 | 1406 | 6.69 | 745 | 6.42 | 210 | 7.65 | 7708 | 5.96 | 9559 | 6.87 |
| **CRF** | 679 | 4.06 | 2430 | 6.39 | 1417 | 5.10 | 2647 | 7.55 | 1499 | 6.77 | 2392 | 5.54 | 814 | 3.87 | 575 | 4.95 | 116 | 4.22 | 7245 | 5.60 | 5657 | 4.07 |
| **Mild LD** | 1194 | 7.14 | 2281 | 6.00 | 1740 | 6.27 | 13314 | 37.98 | 10620 | 47.96 | 2435 | 5.64 | 1332 | 6.34 | 893 | 7.69 | 191 | 6.96 | 7774 | 6.01 | 8638 | 6.21 |
| **M S LD** | 32 | 0.19 | 80 | 0.21 | 62 | 0.22 | 1256 | 3.58 | 1308 | 5.91 | 60 | 0.14 | 25 | 0.12 | 58 | 0.50 | 7 | 0.25 | 279 | 0.22 | 305 | 0.22 |
| **Ulcer** | 2384 | 14.25 | 5978 | 15.73 | 4587 | 16.52 | 7666 | 21.87 | 6041 | 27.28 | 7224 | 16.73 | 3408 | 16.22 | 1616 | 13.93 | 408 | 14.86 | 18517 | 14.32 | 20357 | 14.64 |
| **RD** | 191 | 1.14 | 205 | 0.54 | 346 | 1.25 | 221 | 0.63 | 366 | 1.65 | 321 | 0.74 | 279 | 1.33 | 67 | 0.58 | 59 | 2.15 | 743 | 0.57 | 1910 | 1.37 |
| **AIDS** | 0 | 0.00 | <5** | NA | 0 | 0.00 | 13 | 0.04 | <5 | NA | 5 | 0.01 | <5 | NA | <5 | NA | 0 | 0.00 | 16 | 0.01 | 6 | 0.00 |
| **HT UC** | 7993 | 47.77 | 16559 | 43.57 | 13425 | 48.35 | 15393 | 43.91 | 11457 | 51.74 | 17588 | 40.73 | 10209 | 48.59 | 4509 | 38.85 | 1326 | 48.29 | 50294 | 38.89 | 61740 | 44.40 |
| **HT C** | 3619 | 21.63 | 7499 | 19.73 | 6284 | 22.63 | 6437 | 18.36 | 5118 | 23.11 | 7847 | 18.17 | 4540 | 21.61 | 1811 | 15.61 | 568 | 20.68 | 22414 | 17.33 | 27321 | 19.65 |

*Death due to the cancer or other causes was decided by the NCI classification algorithm using the TCOD and TCR from 2004 until Dec. 31, 2016; survival information of patients not included in the TCOD were obtained from the beneficiary registry of NHIRD at Dec. 31, 2015; the latter were all considered alive.

**The percentage is calculated by deleting the cells whose case number <5.

Table S6A. Hazard Ratios from Original Model Main18&11 using Five-Cancer.

| Comorbid condition | Coef | HR | Comorbid condition | Coef | HR |
|---|---|---|---|---|---|
| Age | 0.06 | 1.07 | CVD*Mild LD | -0.13 | 0.88 |
| Sex | 0.49 | 1.63 | COPD*Dementia | 0.06 | 1.07 |
| AMI | 0.23 | 1.26 | COPD*DM W/O CC | -0.11 | 0.90 |
| Old MI | 0.07 | 1.08 | COPD*DM W CC | -0.03 | 0.97 |
| CHF | 0.77 | 2.16 | COPD*CRF | -0.19 | 0.83 |
| PVD | 0.23 | 1.26 | COPD*Ulcer | -0.02 | 0.98 |
| CVD | 0.40 | 1.49 | COPD*HT UC | 0.00 | 1.00 |
| COPD | 0.28 | 1.33 | COPD*HT C | 0.09 | 1.09 |
| Dementia | 0.71 | 2.03 | COPD*Mild LD | -0.14 | 0.87 |
| Paralysis | 0.34 | 1.41 | Dementia*DM W/O CC | -0.03 | 0.97 |
| DM W/O CC | 0.24 | 1.27 | Dementia*DM W CC | -0.05 | 0.95 |
| DM W CC | 0.39 | 1.48 | Dementia*CRF | -0.33 | 0.72 |
| CRF | 0.78 | 2.18 | Dementia*Ulcer | -0.09 | 0.91 |
| Mild LD | -0.07 | 0.94 | Dementia*HT UC | -0.04 | 0.96 |
| M S LD | 0.93 | 2.52 | Dementia*HT C | -0.05 | 0.95 |
| Ulcer | -0.01 | 0.99 | Dementia*Mild LD | -0.17 | 0.85 |
| RD | 0.30 | 1.35 | DM W/O CC*DM W CC | -0.12 | 0.88 |
| AIDS | 1.66 | 5.28 | DM W/O CC*CRF | 0.03 | 1.03 |
| HT UC | 0.03 | 1.03 | DM W/O CC*Ulcer | 0.09 | 1.09 |
| HT C | -0.01 | 0.99 | DM W/O CC*HT UC | 0.04 | 1.04 |
| CHF*CVD | -0.11 | 0.90 | DM W/O CC*HT C | 0.12 | 1.12 |
| CHF*COPD | -0.12 | 0.89 | DM W/O CC*Mild LD | 0.02 | 1.03 |
| CHF*Dementia | -0.31 | 0.73 | DM W CC*CRF | 0.03 | 1.03 |
| CHF*DM W/O CC | -0.02 | 0.98 | DM W CC*Ulcer | -0.01 | 0.99 |
| CHF*DM W CC | -0.09 | 0.91 | DM W CC*HT UC | -0.01 | 0.99 |
| CHF*CRF | -0.05 | 0.95 | DM W CC*HT C | 0.03 | 1.03 |
| CHF*Ulcer | -0.03 | 0.97 | DM W CC*Mild LD | -0.09 | 0.92 |
| CHF*HT UC | -0.17 | 0.85 | CRF*Ulcer | -0.02 | 0.98 |
| CHF*HT C | 0.03 | 1.03 | CRF*HT UC | -0.07 | 0.93 |
| CHF*Mild LD | -0.04 | 0.96 | CRF*HT C | 0.30 | 1.34 |
| CVD*COPD | 0.00 | 1.00 | CRF*Mild LD | -0.19 | 0.83 |
| CVD*Dementia | -0.06 | 0.94 | Ulcer*HT UC | 0.01 | 1.01 |
| CVD*DM W/O CC | 0.11 | 1.12 | Ulcer*HT C | -0.08 | 0.93 |
| CVD*DM W CC | -0.09 | 0.91 | Ulcer*Mild LD | -0.01 | 0.99 |
| CVD*CRF | -0.16 | 0.85 | HT UC*HT C | 0.00 | 1.00 |
| CVD*Ulcer | 0.06 | 1.06 | HT UC*Mild LD | -0.01 | 0.99 |
| CVD*HT UC | 0.05 | 1.06 | HT C*Mild LD | 0.03 | 1.03 |
| CVD*HT C | -0.08 | 0.92 | | | |

Table S6B. Hazard ratios from Negative Deleted Model Main18&11.ND using Five-Cancer.

| Comorbid condition | Coef | HR | Comorbid condition | Coef | HR |
|---|---|---|---|---|---|
| Age | 0.06 | 1.07 | CHF*HT UC | -0.17 | 0.85 |
| Sex | 0.49 | 1.63 | CVD*COPD | 0.01 | 1.01 |
| AMI | 0.27 | 1.3 | CVD*Dementia | -0.07 | 0.93 |
| Old MI | 0.08 | 1.08 | CVD* DM W/O CC | 0.12 | 1.13 |
| CHF | 0.75 | 2.11 | CVD* DM W CC | -0.08 | 0.92 |
| PVD | 0.24 | 1.27 | CVD*CRF | -0.16 | 0.85 |
| CVD | 0.37 | 1.45 | CVD*HT UC | 0.06 | 1.06 |
| COPD | 0.26 | 1.3 | COPD*Dementia | 0.05 | 1.05 |
| Dementia | 0.66 | 1.94 | COPD* DM W/O CC | -0.1 | 0.9 |
| Paralysis | 0.34 | 1.4 | COPD* DM W CC | -0.02 | 0.98 |
| DM W/O CC | 0.28 | 1.32 | COPD*CRF | -0.17 | 0.84 |
| DM W CC | 0.38 | 1.46 | COPD*HT UC | 0.01 | 1.01 |
| CRF | 0.8 | 2.23 | Dementia* DM W/O CC | -0.02 | 0.98 |
| M S LD | 0.81 | 2.24 | Dementia* DM W CC | -0.05 | 0.95 |
| RD | 0.29 | 1.33 | Dementia*CRF | -0.35 | 0.71 |
| AIDS | 1.65 | 5.22 | Dementia*HT UC | -0.04 | 0.96 |
| HT UC | 0.02 | 1.02 | DM W/O CC * DM W CC | -0.11 | 0.89 |
| CHF*CVD | -0.12 | 0.88 | DM W/O CC *CRF | 0.07 | 1.07 |
| CHF*COPD | -0.11 | 0.9 | DM W/O CC *HT UC | 0.05 | 1.05 |
| CHF*Dementia | -0.31 | 0.74 | DM W CC *CRF | 0.11 | 1.12 |
| CHF* DM W/O CC | 0.02 | 1.02 | DM W CC *HT UC | 0 | 1 |
| CHF* DM W CC | -0.08 | 0.93 | CRF*HT UC | -0.05 | 0.95 |
| CHF*CRF | 0.03 | 1.03 | | | |

Table S6C. Hazard ratios from Variable Selected Model Main18&11.VS using Five-Cancer.

| Comorbid condition | Coef | HR | Comorbid condition | Coef | HR |
|---|---|---|---|---|---|
| Diagnosis age | 0.06 | 1.07 | HT C | -0.07 | 0.93 |
| Sex | 0.49 | 1.64 | CHF*Dementia | -0.38 | 0.69 |
| AMI | 0.25 | 1.28 | CHF*HT UC | -0.20 | 0.82 |
| CHF | 0.68 | 1.98 | CVD*DM W/O CC | 0.10 | 1.10 |
| PVD | 0.23 | 1.26 | CVD*CRF | -0.19 | 0.82 |
| CVD | 0.43 | 1.54 | CVD*HT C | -0.09 | 0.91 |
| COPD | 0.29 | 1.34 | CVD*Mild LD | -0.13 | 0.87 |
| Dementia | 0.60 | 1.83 | COPD*DM W/O CC | -0.11 | 0.90 |
| Paralysis | 0.35 | 1.42 | COPD*CRF | -0.21 | 0.81 |
| DM W/O CC | 0.29 | 1.34 | COPD*Mild LD | -0.15 | 0.86 |
| DM W CC | 0.36 | 1.43 | Dementia*CRF | -0.37 | 0.69 |
| CRF | 0.76 | 2.13 | DM W/O CC*DM W CC | -0.14 | 0.87 |
| M S LD | 0.93 | 2.52 | DM W/O CC*Mild LD | 0.13 | 1.14 |
| RD | 0.30 | 1.35 | CRF*HT C | 0.30 | 1.35 |
| AIDS | 1.66 | 5.26 | CRF*Mild LD | -0.20 | 0.82 |
| HT UC | 0.04 | 1.04 | | | |

Table S7A. The AUCs using OM, NDM, and VSM and the differences based on the **Breast.F.65** validation set

| Training set | Covariates | OM | NDM | VSM | Difference OM-NDM | Difference NDM-VSM |
|---|---|---|---|---|---|---|
| Five-Cancer | Main18 | 0.7593 | 0.7598 | 0.7133 | -0.0004 | 0.0465 |
| Five-Cancer.65 | Main18 | 0.7595 | 0.7601 | 0.7126 | -0.0007 | 0.0476 |
| Five-Cancer.F | Main18 | 0.7585 | 0.7584 | 0.7133 | 0.0000 | 0.0451 |
| Five-Cancer.F.65 | Main18 | 0.7592 | 0.7592 | 0.7126 | 0.0000 | 0.0466 |
| Breast.F | Main17 | 0.7608 | 0.7617 | 0.7135 | -0.0009 | 0.0482 |
| Breast.F.65 | Main17 | 0.7623 | 0.7628 | 0.7237 | -0.0005 | 0.0390 |
| Five-Cancer | Main18&6 | 0.7590 | 0.7592 | 0.7137 | -0.0002 | 0.0455 |
| Five-Cancer.65 | Main18&6 | 0.7586 | 0.7593 | 0.7183 | -0.0007 | 0.0410 |
| Five-Cancer.F | Main18&6 | 0.7565 | 0.7574 | 0.7158 | -0.0009 | 0.0416 |
| Five-Cancer.F.65 | Main18&6 | 0.7563 | 0.7579 | 0.7126 | -0.0015 | 0.0453 |
| Breast.F | Main17&6 | 0.7578 | 0.7605 | 0.7154 | -0.0027 | 0.0451 |
| Breast.F.65 | Main17&6 | 0.7579 | 0.7616 | 0.7248 | -0.0037 | 0.0368 |
| Five-Cancer | Main18&11 | **0.7594** | **0.7609** | **0.7261** | -0.0015 | 0.0348 |
| Five-Cancer.65 | Main18&11 | 0.7579 | 0.7609 | 0.7243 | -0.0030 | 0.0366 |
| Five-Cancer.F | Main18&11 | 0.7563 | 0.7568 | 0.7261 | -0.0005 | 0.0307 |
| Five-Cancer.F.65 | Main18&11 | 0.7550 | 0.7588 | 0.7241 | -0.0038 | 0.0347 |
| Breast.F | Main17&11 | 0.7490 | 0.7514 | 0.7273 | -0.0024 | 0.0241 |
| Breast.F.65 | Main17&11 | 0.7445 | 0.7480 | 0.7197 | -0.0035 | 0.0283 |
| Difference | | 0.0178 | 0.0148 | 0.0147 | | |
| | | | | Average | -0.0015 | 0.0399 |
| | | | | SD | 0.0013 | 0.0072 |

Table S7B. The AUCs using OM, NDM, and VSM and the differences based on the **CRC.M.65** validation set

| Training set | Covariates | OM | NDM | VSM | Difference OM-NDM | Difference NDM-VSM |
|---|---|---|---|---|---|---|
| Five-Cancer | Main18 | 0.7122 | 0.7125 | 0.6794 | -0.0003 | 0.0330 |
| Five-Cancer.65 | Main18 | 0.7118 | 0.7123 | 0.6786 | -0.0004 | 0.0336 |
| Five-Cancer.M | Main18 | 0.7110 | 0.7114 | 0.6791 | -0.0004 | 0.0323 |
| Five-Cancer.M.65 | Main18 | 0.7106 | 0.7115 | 0.6788 | -0.0009 | 0.0327 |
| CRC.M | Main17 | 0.7126 | 0.7126 | 0.6786 | 0.0000 | 0.0340 |
| CRC.M.65 | Main17 | 0.7121 | 0.7120 | 0.6852 | 0.0000 | 0.0268 |
| Five-Cancer | Main18&6 | 0.7117 | 0.7116 | 0.6816 | 0.0001 | 0.0300 |
| Five-Cancer.65 | Main18&6 | 0.7111 | 0.7112 | 0.6837 | -0.0001 | 0.0274 |
| Five-Cancer.M | Main18&6 | 0.7100 | 0.7119 | 0.6799 | -0.0019 | 0.0320 |
| Five-Cancer.M.65 | Main18&6 | 0.7093 | 0.7112 | 0.6790 | -0.0019 | 0.0322 |
| CRC.M | Main17&6 | 0.7111 | 0.7113 | 0.6792 | -0.0002 | 0.0321 |
| CRC.M.65 | Main17&6 | 0.7105 | 0.7108 | 0.6849 | -0.0003 | 0.0259 |
| Five-Cancer | Main18&11 | **0.7128** | **0.7139** | **0.6907** | -0.0011 | 0.0232 |
| Five-Cancer.65 | Main18&11 | 0.7123 | 0.7136 | 0.6864 | -0.0013 | 0.0272 |
| Five-Cancer.M | Main18&11 | 0.7097 | 0.7123 | 0.6813 | -0.0026 | 0.0310 |
| Five-Cancer.M.65 | Main18&11 | 0.7093 | 0.7112 | 0.6805 | -0.0020 | 0.0307 |
| CRC.M | Main17&11 | 0.7098 | 0.7108 | 0.6851 | -0.0010 | 0.0258 |
| CRC.M.65 | Main17&11 | 0.7093 | 0.7111 | 0.6844 | -0.0018 | 0.0267 |
| Difference | | 0.0035 | 0.0031 | 0.0121 | | |
| | | | | Average | -0.0009 | 0.0298 |
| | | | | SD | 0.0009 | 0.0033 |

Table S7C. The AUCs using OM, NDM, and VSM the differences based on the **CRC.F.65** validation set

| Training set | Covariates | OM | NDM | VSM | Difference OM-NDM | Difference NDM-VSM |
|---|---|---|---|---|---|---|
| Five-Cancer | Main18 | 0.7433 | 0.7436 | 0.7125 | -0.0003 | 0.0311 |
| Five-Cancer.65 | Main18 | 0.7431 | 0.7436 | 0.7096 | -0.0005 | 0.0340 |
| Five-Cancer.F | Main18 | 0.7441 | 0.7441 | 0.7110 | 0.0000 | 0.0331 |
| Five-Cancer.F.65 | Main18 | 0.7442 | 0.7443 | 0.7103 | 0.0000 | 0.0340 |
| CRC.F | Main17 | 0.7429 | 0.7428 | 0.7090 | 0.0001 | 0.0338 |
| CRC.F.65 | Main17 | 0.7430 | 0.7429 | 0.7102 | 0.0001 | 0.0327 |
| Five-Cancer | Main18&6 | 0.7430 | 0.7434 | 0.7096 | -0.0004 | 0.0338 |
| Five-Cancer.65 | Main18&6 | 0.7426 | 0.7432 | 0.7110 | -0.0006 | 0.0322 |
| Five-Cancer.F | Main18&6 | 0.7430 | 0.7433 | 0.7133 | -0.0003 | 0.0300 |
| Five-Cancer.F.65 | Main18&6 | 0.7429 | 0.7434 | 0.7088 | -0.0005 | 0.0346 |
| CRC.F | Main17&6 | 0.7412 | 0.7416 | 0.7121 | -0.0003 | 0.0295 |
| CRC.F.65 | Main17&6 | 0.7408 | 0.7417 | 0.7082 | -0.0009 | 0.0335 |
| Five-Cancer | Main18&11 | **0.7450** | **0.7460** | **0.7228** | -0.0010 | 0.0232 |
| Five-Cancer.65 | Main18&11 | 0.7445 | 0.7458 | 0.7195 | -0.0014 | 0.0263 |
| Five-Cancer.F | Main18&11 | 0.7436 | 0.7443 | 0.7198 | -0.0006 | 0.0244 |
| Five-Cancer.F.65 | Main18&11 | 0.7428 | 0.7453 | 0.7185 | -0.0025 | 0.0268 |
| CRC.F | Main17&11 | 0.7412 | 0.7423 | 0.7115 | -0.0011 | 0.0308 |
| CRC.F.65 | Main17&11 | 0.7403 | 0.7420 | 0.7094 | -0.0017 | 0.0326 |
| Difference | | 0.0047 | 0.0044 | 0.0146 | | |
| | | | | Average | -0.0007 | 0.0309 |
| | | | | SD | 0.0007 | 0.0035 |

Table S7D. The AUCs using OM, NDM, and VSM and the differences based on the **Liver.M.65** validation set

| Training set | Covariates | OM | NDM | VSM | Difference OM-NDM | Difference NDM-VSM |
|---|---|---|---|---|---|---|
| Five-Cancer | Main18 | 0.6636 | 0.6601 | 0.6415 | 0.0035 | 0.0186 |
| Five-Cancer.65 | Main18 | 0.6654 | 0.6608 | 0.6376 | 0.0046 | 0.0232 |
| Five-Cancer.M | Main18 | 0.6662 | 0.6611 | 0.6405 | 0.0052 | 0.0206 |
| Five-Cancer.M.65 | Main18 | 0.6677 | 0.6615 | 0.6380 | 0.0062 | 0.0235 |
| Liver.M | Main17 | 0.6697 | 0.6604 | 0.6398 | 0.0093 | 0.0206 |
| Liver.M.65 | Main17 | 0.6709 | 0.6617 | 0.6418 | 0.0092 | 0.0199 |
| Five-Cancer | Main18&6 | 0.6621 | 0.6602 | 0.6346 | 0.0019 | 0.0256 |
| Five-Cancer.65 | Main18&6 | 0.6639 | 0.6607 | 0.6360 | 0.0032 | 0.0247 |
| Five-Cancer.M | Main18&6 | 0.6651 | 0.6609 | 0.6401 | 0.0042 | 0.0208 |
| Five-Cancer.M.65 | Main18&6 | 0.6670 | 0.6606 | 0.6383 | 0.0064 | 0.0223 |
| Liver.M | Main17&6 | 0.6687 | 0.6581 | 0.6357 | 0.0106 | 0.0224 |
| Liver.M.65 | Main17&6 | 0.6690 | 0.6597 | 0.6416 | 0.0093 | 0.0181 |
| Five-Cancer | Main18&11 | **0.6652** | **0.6634** | **0.6364** | 0.0017 | 0.0270 |
| Five-Cancer.65 | Main18&11 | 0.6660 | 0.6639 | 0.6357 | 0.0020 | 0.0282 |
| Five-Cancer.M | Main18&11 | 0.6684 | 0.6633 | 0.6394 | 0.0051 | 0.0239 |
| Five-Cancer.M.65 | Main18&11 | 0.6691 | 0.6631 | 0.6324 | 0.0060 | 0.0307 |
| Liver.M | Main17&11 | 0.6676 | 0.6577 | 0.6359 | 0.0099 | 0.0218 |
| Liver.M.65 | Main17&11 | 0.6664 | 0.6576 | 0.6376 | 0.0089 | 0.0199 |
| Difference | | 0.0088 | 0.0063 | 0.0094 | | |
| | | | | Average | 0.0060 | 0.0229 |
| | | | | SD | 0.0029 | 0.0034 |

Table S7E. The AUCs using OM, NDM, and VSM and the differences based on the **Liver.F.65** validation set

| Training set | Covariates | OM | NDM | VSM | Difference OM-NDM | Difference NDM-VSM |
|---|---|---|---|---|---|---|
| Five-Cancer | Main18 | 0.6926 | 0.6901 | 0.6688 | 0.0025 | 0.0212 |
| Five-Cancer.65 | Main18 | 0.6918 | 0.6886 | 0.6698 | 0.0032 | 0.0188 |
| Five-Cancer.F | Main18 | 0.6933 | 0.6925 | 0.6700 | 0.0008 | 0.0225 |
| Five-Cancer.F.65 | Main18 | 0.6937 | 0.6927 | 0.6697 | 0.0010 | 0.0230 |
| Liver.F | Main17 | 0.7001 | 0.6922 | 0.6715 | 0.0079 | 0.0208 |
| Liver.F.65 | Main17 | 0.7005 | 0.6927 | 0.6704 | 0.0077 | 0.0223 |
| Five-Cancer | Main18&6 | 0.6923 | 0.6901 | 0.6677 | 0.0022 | 0.0224 |
| Five-Cancer.65 | Main18&6 | 0.6926 | 0.6887 | 0.6685 | 0.0039 | 0.0202 |
| Five-Cancer.F | Main18&6 | 0.6918 | 0.6920 | 0.6698 | -0.0002 | 0.0222 |
| Five-Cancer.F.65 | Main18&6 | 0.6933 | 0.6925 | 0.6684 | 0.0008 | 0.0241 |
| Liver.F | Main17&6 | 0.6994 | 0.6734 | 0.6714 | 0.0260 | 0.0020 |
| Liver.F.65 | Main17&6 | 0.6997 | 0.6735 | 0.6717 | 0.0262 | 0.0018 |
| Five-Cancer | Main18&11 | **0.6985** | **0.6946** | **0.6727** | 0.0039 | 0.0219 |
| Five-Cancer.65 | Main18&11 | 0.6974 | 0.6928 | 0.6717 | 0.0046 | 0.0211 |
| Five-Cancer.F | Main18&11 | 0.6950 | 0.6957 | 0.6735 | -0.0007 | 0.0222 |
| Five-Cancer.F.65 | Main18&11 | 0.6954 | 0.6981 | 0.6722 | -0.0027 | 0.0259 |
| Liver.F | Main17&11 | 0.7003 | 0.6767 | 0.6744 | 0.0236 | 0.0023 |
| Liver.F.65 | Main17&11 | 0.6995 | 0.6739 | 0.6744 | 0.0255 | -0.0005 |
| Difference | | 0.0087 | 0.0247 | 0.0067 | | |
| | | | | Average | 0.0076 | 0.0175 |
| | | | | SD | 0.0101 | 0.0090 |

Table S7F. The AUCs using OM, NDM, and VSM and the differences based on the **Lung.M.65** validation set

| Training set | Covariates | OM | NDM | VSM | Difference OM-NDM | Difference NDM-VSM |
|---|---|---|---|---|---|---|
| Five-Cancer | Main18 | 0.6527 | 0.6527 | 0.6351 | 0.0000 | 0.0176 |
| Five-Cancer.65 | Main18 | 0.6525 | 0.6525 | 0.6339 | 0.0000 | 0.0186 |
| Five-Cancer.M | Main18 | 0.6532 | 0.6533 | 0.6353 | -0.0001 | 0.0180 |
| Five-Cancer.M.65 | Main18 | 0.6528 | 0.6532 | 0.6337 | -0.0005 | 0.0195 |
| Lung.M | Main17 | 0.6519 | 0.6520 | 0.6346 | -0.0001 | 0.0174 |
| Lung.M.65 | Main17 | 0.6508 | 0.6510 | 0.6337 | -0.0002 | 0.0173 |
| Five-Cancer | Main18&6 | 0.6524 | 0.6525 | 0.6343 | -0.0001 | 0.0182 |
| Five-Cancer.65 | Main18&6 | 0.6521 | 0.6524 | 0.6366 | -0.0003 | 0.0158 |
| Five-Cancer.M | Main18&6 | 0.6531 | 0.6532 | 0.6356 | -0.0001 | 0.0176 |
| Five-Cancer.M.65 | Main18&6 | 0.6525 | 0.6532 | 0.6339 | -0.0007 | 0.0193 |
| Lung.M | Main17&6 | 0.6510 | 0.6510 | 0.6346 | 0.0001 | 0.0164 |
| Lung.M.65 | Main17&6 | 0.6499 | 0.6523 | 0.6396 | -0.0024 | 0.0127 |
| Five-Cancer | Main18&11 | **0.6547** | **0.6561** | **0.6390** | -0.0014 | 0.0171 |
| Five-Cancer.65 | Main18&11 | 0.6540 | 0.6540 | 0.6389 | 0.0000 | 0.0151 |
| Five-Cancer.M | Main18&11 | 0.6551 | 0.6539 | 0.6359 | 0.0012 | 0.0180 |
| Five-Cancer.M.65 | Main18&11 | 0.6542 | 0.6561 | 0.6340 | -0.0019 | 0.0220 |
| Lung.M | Main17&11 | 0.6477 | 0.6495 | 0.6340 | -0.0018 | 0.0155 |
| Lung.M.65 | Main17&11 | 0.6461 | 0.6479 | 0.6410 | -0.0018 | 0.0069 |
| Difference | | 0.0090 | 0.0082 | 0.0073 | | |
| | | | | Average | -0.0006 | 0.0168 |
| | | | | SD | 0.0009 | 0.0032 |

Table S7G. The AUCs using OM, NDM, and VSM and the differences based on the **Lung.F.65** validation set

| Training set | Covariates | OM | NDM | VSM | Difference OM-NDM | Difference NDM-VSM |
|---|---|---|---|---|---|---|
| Five-Cancer | Main18 | 0.6914 | 0.6914 | 0.6782 | 0.0001 | 0.0132 |
| Five-Cancer.65 | Main18 | 0.6914 | 0.6914 | 0.6784 | 0.0000 | 0.0131 |
| Five-Cancer.F | Main18 | 0.6918 | 0.6918 | 0.6784 | 0.0000 | 0.0134 |
| Five-Cancer.F.65 | Main18 | 0.6919 | 0.6919 | 0.6784 | 0.0000 | 0.0135 |
| Lung.F | Main17 | 0.6916 | 0.6917 | 0.6829 | 0.0000 | 0.0088 |
| Lung.F.65 | Main17 | 0.6925 | 0.6919 | 0.6850 | 0.0006 | 0.0069 |
| Five-Cancer | Main18&6 | 0.6915 | 0.6915 | 0.6786 | 0.0000 | 0.0128 |
| Five-Cancer.65 | Main18&6 | 0.6913 | 0.6914 | 0.6789 | -0.0001 | 0.0125 |
| Five-Cancer.F | Main18&6 | 0.6920 | 0.6921 | 0.6796 | -0.0001 | 0.0125 |
| Five-Cancer.F.65 | Main18&6 | 0.6920 | 0.6918 | 0.6783 | 0.0002 | 0.0136 |
| Lung.F | Main17&6 | 0.6895 | 0.6915 | 0.6829 | -0.0020 | 0.0086 |
| Lung.F.65 | Main17&6 | 0.6907 | 0.6894 | 0.6846 | 0.0013 | 0.0048 |
| Five-Cancer | Main18&11 | **0.6905** | **0.6903** | **0.6826** | 0.0002 | 0.0077 |
| Five-Cancer.65 | Main18&11 | 0.6901 | 0.6904 | 0.6812 | -0.0003 | 0.0091 |
| Five-Cancer.F | Main18&11 | 0.6903 | 0.6908 | 0.6820 | -0.0005 | 0.0088 |
| Five-Cancer.F.65 | Main18&11 | 0.6900 | 0.6908 | 0.6809 | -0.0008 | 0.0099 |
| Lung.F | Main17&11 | 0.6851 | 0.6886 | 0.6783 | -0.0035 | 0.0103 |
| Lung.F.65 | Main17&11 | 0.6864 | 0.6880 | 0.6800 | -0.0016 | 0.0080 |
| Difference | | 0.0074 | 0.0041 | 0.0068 | | |
| | | | | Average | -0.0004 | 0.0104 |
| | | | | SD | 0.0011 | 0.0027 |

Table S7H. The AUCs using OM, NDM, and VSM and the differences based on the **Oral.M.65** validation set

| Training set | Covariates | OM | NDM | VSM | Difference OM-NDM | Difference NDM-VSM |
|---|---|---|---|---|---|---|
| Five-Cancer | Main18 | 0.6607 | 0.6614 | 0.6306 | -0.0007 | 0.0307 |
| Five-Cancer.65 | Main18 | 0.6585 | 0.6594 | 0.6296 | -0.0009 | 0.0299 |
| Five-Cancer.M | Main18 | 0.6578 | 0.6587 | 0.6306 | -0.0009 | 0.0281 |
| Five-Cancer.M.65 | Main18 | 0.6548 | 0.6568 | 0.6260 | -0.0020 | 0.0308 |
| Oral.M | Main17 | 0.6638 | 0.6641 | 0.6262 | -0.0003 | 0.0379 |
| Oral.M.65 | Main17 | 0.6582 | 0.6636 | 0.6343 | -0.0053 | 0.0293 |
| Five-Cancer | Main18&6 | 0.6602 | 0.6611 | 0.6266 | -0.0009 | 0.0344 |
| Five-Cancer.65 | Main18&6 | 0.6576 | 0.6593 | 0.6339 | -0.0017 | 0.0255 |
| Five-Cancer.M | Main18&6 | 0.6573 | 0.6582 | 0.6329 | -0.0008 | 0.0253 |
| Five-Cancer.M.65 | Main18&6 | 0.6545 | 0.6569 | 0.6264 | -0.0025 | 0.0306 |
| Oral.M | Main17&6 | 0.6622 | 0.6639 | 0.6282 | -0.0017 | 0.0357 |
| Oral.M.65 | Main17&6 | 0.6521 | 0.6621 | 0.6445 | -0.0100 | 0.0176 |
| Five-Cancer | Main18&11 | **0.6626** | **0.6627** | **0.6387** | -0.0001 | 0.0240 |
| Five-Cancer.65 | Main18&11 | 0.6590 | 0.6593 | 0.6346 | -0.0003 | 0.0247 |
| Five-Cancer.M | Main18&11 | 0.6588 | 0.6595 | 0.6282 | -0.0007 | 0.0313 |
| Five-Cancer.M.65 | Main18&11 | 0.6548 | 0.6563 | 0.6270 | -0.0016 | 0.0294 |
| Oral.M | Main17&11 | 0.6531 | 0.6490 | 0.6293 | 0.0041 | 0.0197 |
| Oral.M.65 | Main17&11 | 0.6400 | 0.6531 | 0.6498 | -0.0130 | 0.0033 |
| Difference | | 0.0238 | 0.0151 | 0.0238 | | |
| | | | | Average | -0.0022 | 0.0271 |
| | | | | SD | 0.0039 | 0.0079 |

Table S7I. The AUCs using OM, NDM, and VSM and the differences based on the **Oral.F.65** validation set

| Training set | Covariates | OM | NDM | VSM | Difference OM-NDM | Difference NDM-VSM |
|---|---|---|---|---|---|---|
| Five-Cancer | Main18 | 0.6952 | 0.6975 | 0.6846 | -0.0022 | 0.0128 |
| Five-Cancer.65 | Main18 | 0.6949 | 0.6978 | 0.6848 | -0.0029 | 0.0131 |
| Five-Cancer.F | Main18 | 0.6962 | 0.6971 | 0.6870 | -0.0009 | 0.0102 |
| Five-Cancer.F.65 | Main18 | 0.6957 | 0.6965 | 0.6853 | -0.0008 | 0.0112 |
| Oral.F | Main17 | 0.6892 | 0.6957 | 0.6925 | -0.0065 | 0.0031 |
| Oral.F.65 | Main17 | 0.6893 | 0.6924 | 0.6915 | -0.0030 | 0.0009 |
| Five-Cancer | Main18&6 | 0.6957 | 0.6976 | 0.6851 | -0.0020 | 0.0125 |
| Five-Cancer.65 | Main18&6 | 0.6954 | 0.6981 | 0.6910 | -0.0027 | 0.0071 |
| Five-Cancer.F | Main18&6 | 0.6952 | 0.6968 | 0.6903 | -0.0016 | 0.0064 |
| Five-Cancer.F.65 | Main18&6 | 0.6953 | 0.6974 | 0.6849 | -0.0021 | 0.0124 |
| Oral.F | Main17&6 | 0.6866 | 0.6882 | 0.6923 | -0.0015 | -0.0041 |
| Oral.F.65 | Main17&6 | 0.6887 | 0.6913 | 0.6879 | -0.0026 | 0.0033 |
| Five-Cancer | Main18&11 | **0.6961** | **0.6970** | **0.6857** | -0.0009 | 0.0113 |
| Five-Cancer.65 | Main18&11 | 0.6966 | 0.7003 | 0.6837 | -0.0037 | 0.0166 |
| Five-Cancer.F | Main18&11 | 0.6936 | 0.6953 | 0.6893 | -0.0017 | 0.0060 |
| Five-Cancer.F.65 | Main18&11 | 0.6939 | 0.6938 | 0.6844 | 0.0001 | 0.0094 |
| Oral.F | Main17&11 | 0.6846 | 0.6924 | 0.6939 | -0.0078 | -0.0015 |
| Oral.F.65 | Main17&11 | 0.6843 | 0.6842 | 0.7132 | 0.0002 | -0.0291 |
| Difference | | 0.0123 | 0.0161 | 0.0295 | | |
| | | | | Average | -0.0024 | 0.0057 |
| | | | | SD | 0.0020 | 0.0103 |

Table S7J. Comparison of the AUCs based on the validation and test sets for the model Main18&11.ND.

| Five-Cancer | Main18&11 | NDM | Validation set | Test set |
|---|---|---|---|---|
| | | Breast.F.65 | 0.7609 | 0.7316 |
| | | CRC.M.65 | 0.7139 | 0.7129 |
| | | CRC.F.65 | 0.7460 | 0.7512 |
| | | Liver.M.65 | 0.6634 | 0.6818 |
| | | Liver.F.65 | 0.6946 | 0.6907 |
| | | Lung.M.65 | 0.6561 | 0.6440 |
| | | Lung.F.65 | 0.6903 | 0.7156 |
| | | Oral.M.65 | 0.6627 | 0.6511 |
| | | Oral.F.65 | 0.6970 | 0.7133 |

Table S8. Algorithm for mapping stage at diagnosis from TNM to SEER Summary Stage (localized, regional, and distant):

| | 6th edition | 7th edition |
|---|---|---|
| Cancer /SEER stages | | |
| **Breast** | | |
| Localized | T1, T2, T3 | T1mi-1c, T2, T3 |
| Regional | N1-3, T4 | N1-3b, T4 |
| Distant | N3c, M1 | N3c, M1, MC |
| Non-Distant* | M0, MB | M0, MB |
| **Colorectal** | | |
| Localized | T1, T2, T3 | T1, T2, T3 |
| Regional | N1, N2, T4 | N1, N2, T4 |
| Distant | M1 | M1, MC |
| Non-Distant* | M0, MB | M0, MB |
| **Lung**, | | |
| Localized | T1, T2 | T1, T2 |
| Regional | N1, N2, T3, T4 | N1, N2, T3, T4 |
| Distant | M1 | N3, M1, MC |
| Non-Distant* | M0, MB | M0, MB |
| **Oral-** | | |
| Localized | T1, T2 | T1, T2 |
| Regional | N1-3, T3, T4 | N1-3, T3, T4 |
| Distant | M1 | M1, MC |
| Non-Distant* | M0, MB | M0, MB |
| **Liver-**Liver cell Carcinoma | | |
| Localized | T1, T2, T3 | T1, T2, T3(a) |
| Regional | N1, T4 | N1, T3b, T4 |
| Distant | M1 | M1, MC |
| Non-Distant* | M0, MB | M0, MB |
| **Liver**-Intrahepatic bile duct carcinoma | | |
| Localized | T1, T2, T3 | T1, T2 |
| Regional | N1, T4 | N1, T3, T4 |
| Distant | M1 | M1, MC |
| Non-Distant* | M0, MB | M0, MB |

The cancer stage information for cancer cases was obtained from The Taiwan Cancer Registry Long Form Database, which are based on the TNM stage information derived from the American Joint Committee on Cancer (AJCC) and the International Union Against Cancer (UICC). AJCC 6th edition was used from 2004 to 2009, 7th was used since 2010. The algorithms for mapping stage at diagnosis from TNM to SEER

Summary Stage in Table S8 follows that of Japan National Cancer Control and Information Service (https://ganjoho.jp/data/reg_stat/cancer_reg/hospital/info/toroku08.pdf) and clinical advice.

* For patients whose cancer stage cannot be classified into localized, regional and distant stages, we categorize their cancer stage as non-distant stage if the metastatic information can be obtained from TCR-LF.

Table S9A. Number of patients by age, stage, and comorbidity level: **Breast Cancer**

| Breast cancer | | | | | | | | | | | | | |
|---|---|---|---|---|---|---|---|---|---|---|---|---|---|
| Age | | 15-64 | | | 65-74 | | | 75-84 | | | 85-94 | | |
| Comorbidity level* | | 0 | 1 | 2 | 0 | 1 | 2 | 0 | 1 | 2 | 0 | 1 | 2 |
| Stage** | NA+ | 6113 | 1790 | 543 | 459 | 680 | 376 | 144 | 333 | 237 | 40 | 78 | 86 |
| | Localized | 27105 | 8104 | 2092 | 1592 | 2688 | 1043 | 356 | 1034 | 696 | 58 | 161 | 172 |
| | Regional | 18393 | 5180 | 1353 | 1059 | 1704 | 722 | 244 | 665 | 435 | 40 | 116 | 105 |
| | Distant | 2846 | 542 | 147 | 268 | 268 | 119 | 84 | 136 | 104 | 14 | 45 | 20 |
| | | X-squared = 103.86, p-value < 2.2e-16 | | | X-squared = 36.705, p-value = 2.072e-07 | | | X-squared = 15.865, p-value = 0.003206 | | | X-squared = 9.7462, p-value = 0.04493 | | |
| | Non-distant | 423 | 101 | 29 | 35 | 42 | 15 | <5 | 29 | 21 | <5 | <5 | <5 |
| | Unknown++ | 680 | 122 | 46 | 46 | 54 | 23 | 10 | 28 | 20 | 9 | <5 | 5 |

*comorbidity level: 0 refers to individuals with no comorbid conditions. 2 refers to conditions that include comorbid conditions 3, 6, 7, 11, 13, 16 or the comorbid index exceeded 0.66. 1 refers to individuals not classified into 0 nor 2 of comorbidity level.

** For patients whose cancer stage cannot be classified into localized, regional and distant stages, we categorize their cancer stage as non-distant stage if the metastatic information can be obtained from TCR-LF. Patients whose cancer stage cannot be classified into localized, regional, distant and non-distant stage is categorized as unknown stage.

+ Cancer patients in TCR but not in TCRLF.

++ Cancer patients in TCRLF stage information missing.

Table S9B. Number of patients by age, stage, and comorbidity level: **Colorectal Cancer**

| Colorectal cancer | | | | | | | | | | | | | | | | | | | | | | | | | |
|---|---|---|---|---|---|---|---|---|---|---|---|---|---|---|---|---|---|---|---|---|---|---|---|---|---|
| Age | | 15-64 | | | | | | 65-74 | | | | | | 75-84 | | | | | | 85-94 | | | | | |
| Sex | | Male | | | Female | | | Male | | | Female | | | Male | | | Female | | | Male | | | Female | | |
| Comorbidity level* | | 0 | 1 | 2 | 0 | 1 | 2 | 0 | 1 | 2 | 0 | 1 | 2 | 0 | 1 | 2 | 0 | 1 | 2 | 0 | 1 | 2 | 0 | 1 | 2 |
| Stage** | NA+ | 2134 | 1097 | 426 | 1568 | 674 | 274 | 908 | 1067 | 834 | 560 | 805 | 471 | 712 | 1071 | 1242 | 404 | 853 | 800 | 222 | 265 | 451 | 203 | 309 | 417 |
| | Localized | 5896 | 3492 | 1117 | 4673 | 2184 | 674 | 1975 | 2657 | 1629 | 1257 | 2021 | 998 | 1181 | 2058 | 2025 | 698 | 1732 | 1172 | 254 | 446 | 628 | 222 | 458 | 500 |
| | Regional | 5689 | 2713 | 817 | 4818 | 1816 | 499 | 1777 | 2036 | 1149 | 1190 | 1651 | 709 | 1073 | 1671 | 1489 | 746 | 1537 | 1069 | 236 | 384 | 494 | 199 | 466 | 394 |
| | Distant | 3398 | 1382 | 439 | 2971 | 921 | 290 | 1023 | 1095 | 626 | 612 | 803 | 392 | 625 | 1032 | 925 | 448 | 854 | 600 | 152 | 222 | 322 | 160 | 281 | 260 |
| | | X-squared = 135.1, p-value < 2.2e-16 | | | X-squared = 111.15, p-value < 2.2e-16 | | | X-squared = 40.276, p-value = 3.795e-08 | | | X-squared = 25.809, p-value = 3.457e-05 | | | X-squared = 16.571, p-value = 0.002341 | | | X-squared = 15.891, p-value = 0.003169 | | | X-squared = 4.0896, p-value = 0.394 | | | X-squared = 13.113, p-value = 0.01074 | | |
| | Non-distant | 457 | 284 | 125 | 353 | 164 | 54 | 118 | 193 | 124 | 68 | 114 | 71 | 61 | 90 | 161 | 24 | 101 | 118 | 26 | 49 | 85 | 28 | 63 | 91 |
| | Unknown++ | 572 | 249 | 98 | 443 | 168 | 55 | 169 | 181 | 137 | 103 | 137 | 78 | 118 | 145 | 226 | 67 | 148 | 109 | 34 | 56 | 77 | 43 | 62 | 89 |

*comorbidity level: 0 refers to individuals with no comorbid conditions. 2 refers to conditions that include comorbid conditions 3, 6, 7, 11, 13, 16 or the comorbid score exceeded 0.66. 1 refers to individuals not classified into 0 nor 2 of comorbidity level.

** For patients whose cancer stage cannot be classified into localized, regional and distant stages, we categorize their cancer stage as non-distant stage if the metastatic information can be obtained from TCR-LF. Patients whose cancer stage cannot be classified into localized, regional, distant and non-distant stage is categorized as unknown stage.

+ Cancer patients in TCR but not in TCRLF.

++ Cancer patients in TCRLF with stage information missing.

Table S9C. Number of patients by age, stage, and comorbidity level: **Liver Cancer**

| Liver cancer | | | | | | | | | | | | | | | | | | | | | | | | | |
|---|---|---|---|---|---|---|---|---|---|---|---|---|---|---|---|---|---|---|---|---|---|---|---|---|---|
| Age | | 15-64 | | | | | | 65-74 | | | | | | 75-84 | | | | | | 85-94 | | | | | |
| Sex | | Male | | | Female | | | Male | | | Female | | | Male | | | Female | | | Male | | | Female | | |
| Comorbidity level* | | 0 | 1 | 2 | 0 | 1 | 2 | 0 | 1 | 2 | 0 | 1 | 2 | 0 | 1 | 2 | 0 | 1 | 2 | 0 | 1 | 2 | 0 | 1 | 2 |
| Stage** | NA+ | 1627 | 1289 | 928 | 306 | 354 | 266 | 536 | 800 | 799 | 191 | 542 | 493 | 337 | 720 | 953 | 183 | 530 | 618 | 125 | 173 | 295 | 83 | 194 | 266 |
| | Localized | 9176 | 11912 | 5352 | 1673 | 3560 | 1598 | 2180 | 5852 | 3942 | 764 | 4032 | 2395 | 984 | 3095 | 3107 | 404 | 2581 | 2060 | 167 | 465 | 628 | 108 | 381 | 370 |
| | Regional | 3940 | 2209 | 833 | 641 | 461 | 163 | 801 | 1110 | 646 | 241 | 591 | 343 | 356 | 645 | 608 | 153 | 532 | 385 | 55 | 137 | 158 | 50 | 145 | 132 |
| | Distant | 3320 | 1293 | 478 | 636 | 352 | 147 | 674 | 750 | 509 | 260 | 461 | 244 | 269 | 472 | 488 | 156 | 394 | 277 | 63 | 107 | 115 | 50 | 90 | 93 |
| | | X-squared = 2303.7, p-value < 2.2e-16 | | | X-squared = 686.46, p-value < 2.2e-16 | | | X-squared = 420.4, p-value < 2.2e-16 | | | X-squared = 250.5, p-value < 2.2e-16 | | | X-squared = 106.06, p-value < 2.2e-16 | | | X-squared = 116.53, p-value < 2.2e-16 | | | X-squared = 17.53, p-value = 0.001524 | | | X-squared = 12.144, p-value = 0.01631 | | |
| | Non-distant | 113 | 83 | 39 | 27 | 17 | 11 | 25 | 60 | 41 | 15 | 42 | 21 | 16 | 36 | 54 | 11 | 44 | 57 | 7 | 23 | 40 | 6 | 29 | 32 |
| | Unknown++ | 881 | 661 | 371 | 230 | 223 | 116 | 248 | 395 | 248 | 98 | 264 | 156 | 115 | 245 | 246 | 60 | 213 | 162 | 28 | 44 | 66 | 29 | 52 | 60 |

*comorbidity level: 0 refers to individuals with no comorbid conditions. 2 refers to conditions that include comorbid conditions 3, 6, 7, 11, 13, 16 or the comorbid score exceeded 0.66. 1 refers to individuals not classified into 0 nor 2 of comorbidity level.

** For patients whose cancer stage cannot be classified into localized, regional and distant stages, we categorize their cancer stage as non-distant stage if the metastatic information can be obtained from TCR-LF. Patients whose cancer stage cannot be classified into localized, regional, distant and non-distant stage is categorized as unknown stage.

+ Cancer patient in TCR but not in TCRLF.

++ Cancer patient in TCRLF stage information missing.

Table S9D. Number of patients by age, stage, and comorbidity level: **Lung Cancer**

| Lung cancer | | | | | | | | | | | | | | | | | | | | | | | | | |
|---|---|---|---|---|---|---|---|---|---|---|---|---|---|---|---|---|---|---|---|---|---|---|---|---|---|
| Age | | 15-64 | | | | | | 65-74 | | | | | | 75-84 | | | | | | 85-94 | | | | | |
| Sex | | Male | | | Female | | | Male | | | Female | | | Male | | | Female | | | Male | | | Female | | |
| Comorbidity level* | | 0 | 1 | 2 | 0 | 1 | 2 | 0 | 1 | 2 | 0 | 1 | 2 | 0 | 1 | 2 | 0 | 1 | 2 | 0 | 1 | 2 | 0 | 1 | 2 |
| Stage** | NA[+] | 945 | 328 | 258 | 521 | 201 | 91 | 694 | 504 | 686 | 276 | 326 | 241 | 791 | 797 | 1558 | 252 | 447 | 539 | 235 | 279 | 641 | 151 | 232 | 300 |
| | Localized | 1458 | 908 | 420 | 2035 | 1079 | 395 | 584 | 834 | 723 | 468 | 788 | 430 | 385 | 577 | 1005 | 156 | 440 | 349 | 58 | 103 | 242 | 19 | 64 | 70 |
| | Regional | 2393 | 1068 | 612 | 1420 | 663 | 275 | 1322 | 1208 | 1228 | 454 | 638 | 363 | 955 | 1227 | 1862 | 212 | 520 | 418 | 178 | 221 | 470 | 72 | 125 | 157 |
| | Distant | 7678 | 3027 | 1350 | 5915 | 2274 | 753 | 3463 | 3302 | 2494 | 1839 | 2426 | 1127 | 2489 | 3342 | 4174 | 1042 | 2261 | 1582 | 566 | 865 | 1330 | 302 | 680 | 592 |
| | | X-squared = 151.4, p-value < 2.2e-16 | | | X-squared = 91.334, p-value < 2.2e-16 | | | X-squared = 116.3, p-value < 2.2e-16 | | | X-squared = 34.998, p-value = 4.649e-07 | | | X-squared = 71.589, p-value = 1.048e-14 | | | X-squared = 19.346, p-value = 0.0006719 | | | X-squared = 29.34, p-value = 6.666e-06 | | | X-squared = 13.221, p-value = 0.01024 | | |
| | Non-distant | 46 | 18 | 13 | 28 | 10 | 6 | 14 | 21 | 30 | 7 | <5 | 11 | 27 | 23 | 65 | 11 | 18 | 31 | 18 | 20 | 38 | <5 | 17 | 19 |
| | Unknown[++] | 332 | 145 | 90 | 176 | 69 | 48 | 238 | 181 | 205 | 54 | 75 | 67 | 180 | 196 | 344 | 51 | 101 | 81 | 44 | 44 | 104 | 23 | 33 | 48 |

*comorbidity level: 0 refers to individuals with no comorbid conditions. 2 refers to conditions that include comorbid conditions 3, 6, 7, 11, 13, 16 or the comorbid score exceeded 0.66. 1 refers to individuals not classified into 0 nor 2 of comorbidity level.

** For patients whose cancer stage cannot be classified into localized, regional and distant stages, we categorize their cancer stage as non-distant stage if the metastatic information can be obtained from TCR-LF. Patients whose cancer stage cannot be classified into localized, regional, distant and non-distant stage is categorized as unknown stage.

[+] Cancer patient in TCR but not in TCRLF.

[++] Cancer patient in TCRLF stage information missing.

Table S9E. Number of patients by age, stage, and comorbidity level: **Oral Cancer**

| Oral cancer | | | | | | | | | | | | | | | | | | | | | | | | | |
|---|---|---|---|---|---|---|---|---|---|---|---|---|---|---|---|---|---|---|---|---|---|---|---|---|---|
| Age | | 15-64 | | | | | | 65-74 | | | | | | 75-84 | | | | | | 85-94 | | | | | |
| Sex | | Male | | | Female | | | Male | | | Female | | | Male | | | Female | | | Male | | | Female | | |
| Comorbidity level* | | 0 | 1 | 2 | 0 | 1 | 2 | 0 | 1 | 2 | 0 | 1 | 2 | 0 | 1 | 2 | 0 | 1 | 2 | 0 | 1 | 2 | 0 | 1 | 2 |
| Stage** | NA[+] | 6013 | 1722 | 624 | 1589 | 424 | 122 | 473 | 424 | 353 | 113 | 145 | 77 | 170 | 172 | 237 | 56 | 77 | 66 | 20 | 32 | 35 | 9 | 19 | 21 |
| | Localized | 10510 | 4528 | 1534 | 1129 | 480 | 178 | 839 | 996 | 619 | 135 | 256 | 125 | 180 | 339 | 327 | 53 | 159 | 118 | 36 | 43 | 72 | 9 | 32 | 32 |
| | Regional | 19379 | 6168 | 2284 | 1966 | 614 | 192 | 1461 | 1298 | 875 | 163 | 264 | 141 | 412 | 467 | 611 | 95 | 156 | 118 | 77 | 93 | 146 | 26 | 42 | 47 |
| | Distant | 878 | 254 | 115 | 92 | 23 | 9 | 107 | 63 | 62 | 9 | 19 | 10 | 35 | 48 | 42 | 6 | 10 | 10 | 7 | 8 | 16 | <5 | <5 | <5 |
| | | X-squared = 197.34, p-value < 2.2e-16 | | | X-squared = 34.364, p-value = 6.274e-07 | | | X-squared = 35.159, p-value = 4.308e-07 | | | X-squared = 1.4998, p-value = 0.8267 | | | X-squared = 23.333, p-value = 0.0001086 | | | X-squared = 10.23, p-value = 0.03672 | | | X-squared = 0.38399, p-value = 0.9838 | | | X-squared = NaN, p-value = NA | | |
| | Non-distant | 254 | 77 | 41 | 43 | 11 | <5 | 16 | 10 | 15 | <5 | <5 | <5 | 13 | 20 | 10 | <5 | 7 | 8 | <5 | 5 | 7 | 0 | 5 | <5 |
| | Unknown[++] | 699 | 220 | 93 | 91 | 26 | 6 | 70 | 53 | 53 | 18 | 18 | 14 | 31 | 34 | 39 | 6 | 10 | 8 | 6 | 12 | 12 | <5 | <5 | 7 |

*comorbidity level: 0 refers to individuals with no comorbid conditions. 2 refers to conditions that include comorbid conditions 3, 6, 7, 11, 13, 16 or the comorbid score exceeded 0.66. 1 refers to individuals not classified into 0 nor 2 of comorbidity level.

** For patients whose cancer stage cannot be classified into localized, regional and distant stages, we categorize their cancer stage as non-distant stage if the metastatic information can be obtained from TCR-LF. Patients whose cancer stage cannot be classified into localized, regional, distant and non-distant stage is categorized as unknown stage.

[+] Cancer patient in TCR but not in TCRLF.

[++] Cancer patient in TCRLF stage information missing.

Table S10A. Numbers of cancer patients in TCR by year of diagnosis and age of diagnosis and the number and percentage of them in TCRLF: **breast cancer**

| Year of diagnosis | TCR | TCRLF | TCRLF/TCR | 15-64 | 65-94 |
|---|---|---|---|---|---|
| sex | Female | Female | (%) | Female | Female |
| 2004 | 6066 | 4337 | 71.5 | 5123 | 943 |
| 2005 | 6465 | 4911 | 76.0 | 5447 | 1018 |
| 2006 | 6762 | 5271 | 78.0 | 5679 | 1083 |
| 2007 | 7372 | 6344 | 86.1 | 6089 | 1283 |
| 2008 | 7825 | 6660 | 85.1 | 6501 | 1324 |
| 2009 | 8429 | 7406 | 87.9 | 6884 | 1545 |
| 2010 | 9115 | 8586 | 94.2 | 7466 | 1649 |
| 2011 | 9394 | 8840 | 94.1 | 7639 | 1755 |
| 2012 | 9789 | 9298 | 95.0 | 7955 | 1834 |
| 2013 | 10377 | 9936 | 95.8 | 8295 | 2082 |
| 2014 | 10749 | 10398 | 96.7 | 8531 | 2218 |
| Age of diagnosis | | | | | |
| 15-34 | 3555 | | | | |
| 35-49 | 33565 | | | | |
| 50-64 | 38489 | | | | |
| 65-74 | 11193 | | | | |
| 75-84 | 4580 | | | | |
| 85-94 | 961 | | | | |

Table S10B. Numbers of cancer patients in TCR by year of diagnosis, age of diagnosis and sex and the number and percentage of them in TCRLF: **colorectal cancer**

| Year of diagnosis | TCR | | | TCRLF | TCRLF/TCR | 15-64 | | 65-94 | |
|---|---|---|---|---|---|---|---|---|---|
| sex | All | Male | Female | All | (%) | Male | Female | Male | Female |
| 2004 | 8455 | 4839 | 3616 | 5693 | 67.3 | 1993 | 1570 | 2846 | 2046 |
| 2005 | 8638 | 4960 | 3678 | 6087 | 70.5 | 1958 | 1625 | 3002 | 2053 |
| 2006 | 9094 | 5169 | 3925 | 6548 | 72.0 | 2160 | 1789 | 3009 | 2136 |
| 2007 | 9634 | 5557 | 4077 | 7937 | 82.4 | 2276 | 1806 | 3281 | 2271 |
| 2008 | 9826 | 5656 | 4170 | 8217 | 83.6 | 2354 | 1776 | 3302 | 2394 |
| 2009 | 10633 | 6105 | 4528 | 9242 | 86.9 | 2626 | 1923 | 3479 | 2605 |
| 2010 | 11936 | 6944 | 4992 | 11052 | 92.6 | 3273 | 2260 | 3671 | 2732 |
| 2011 | 11987 | 6972 | 5015 | 11236 | 93.7 | 3259 | 2317 | 3713 | 2698 |
| 2012 | 12626 | 7264 | 5362 | 11855 | 93.9 | 3503 | 2495 | 3761 | 2867 |
| 2013 | 12783 | 7343 | 5440 | 12005 | 93.9 | 3403 | 2482 | 3940 | 2958 |
| 2014 | 13143 | 7582 | 5561 | 12474 | 94.9 | 3580 | 2556 | 4002 | 3005 |
| Age of diagnosis | | | | | | | | | |
| 15-34 | 2025 | 1085 | 940 | | | | | | |
| 35-49 | 12362 | 6521 | 5841 | | | | | | |
| 50-64 | 38597 | 22779 | 15818 | | | | | | |
| 65-74 | 29738 | 17698 | 12040 | | | | | | |
| 75-84 | 27385 | 15905 | 11480 | | | | | | |
| 85-94 | 8648 | 4403 | 4245 | | | | | | |

Table S10C. Numbers of cancer patients in TCR by year of diagnosis, age of diagnosis and sex and the number and percentage of them in TCRLF: **liver cancer**

| Year of diagnosis | TCR | | | TCRLF | TCRLF/TCR | 15-64 | | 65-94 | |
|---|---|---|---|---|---|---|---|---|---|
| sex | All | Male | Female | All | % | Male | Female | Male | Female |
| 2004 | 9597 | 6929 | 2668 | 7118 | 74.2 | 3944 | 975 | 2985 | 1693 |
| 2005 | 9772 | 7064 | 2708 | 7476 | 76.5 | 4027 | 974 | 3037 | 1734 |
| 2006 | 9985 | 7107 | 2878 | 7956 | 79.7 | 3876 | 1003 | 3231 | 1875 |
| 2007 | 10285 | 7353 | 2932 | 9099 | 88.5 | 4088 | 1015 | 3265 | 1917 |
| 2008 | 10245 | 7194 | 3051 | 9163 | 89.4 | 4019 | 989 | 3175 | 2062 |
| 2009 | 10538 | 7414 | 3124 | 9706 | 92.1 | 4104 | 989 | 3310 | 2135 |
| 2010 | 10424 | 7375 | 3049 | 9839 | 94.4 | 4182 | 964 | 3193 | 2085 |
| 2011 | 10480 | 7403 | 3077 | 9987 | 95.3 | 4152 | 953 | 3251 | 2124 |
| 2012 | 10483 | 7331 | 3152 | 9999 | 95.4 | 4107 | 966 | 3224 | 2186 |
| 2013 | 10542 | 7348 | 3194 | 10246 | 97.2 | 4051 | 1001 | 3297 | 2193 |
| 2014 | 10136 | 7045 | 3091 | 9889 | 97.6 | 3955 | 952 | 3090 | 2139 |
| Age of diagnosis | | | | | | | | | |
| 15-34 | 1682 | 1323 | 359 | | | | | | |
| 35-49 | 13794 | 12105 | 1689 | | | | | | |
| 50-64 | 39810 | 31077 | 8733 | | | | | | |
| 65-74 | 30769 | 19616 | 11153 | | | | | | |
| 75-84 | 21566 | 12746 | 8820 | | | | | | |
| 85-94 | 4866 | 2696 | 2170 | | | | | | |

Table S10D. Numbers of cancer patients in TCR by year of diagnosis, age of diagnosis and sex and the number and percentage of them in TCRLF: **lung cancer**

| Year of diagnosis | TCR | | | TCRLF | TCRLF/TCR | 15-64 | | 65-94 | |
|---|---|---|---|---|---|---|---|---|---|
| sex | All | Male | Female | All | % | Male | Female | Male | Female |
| 2004 | 7587 | 5147 | 2440 | 5666 | 74.7 | 1443 | 971 | 3704 | 1469 |
| 2005 | 7929 | 5318 | 2611 | 6047 | 76.3 | 1575 | 1035 | 3743 | 1576 |
| 2006 | 8266 | 5451 | 2815 | 6456 | 78.1 | 1568 | 1129 | 3883 | 1686 |
| 2007 | 8639 | 5629 | 3010 | 7534 | 87.2 | 1701 | 1190 | 3928 | 1820 |
| 2008 | 8690 | 5673 | 3017 | 7721 | 88.8 | 1720 | 1273 | 3953 | 1744 |
| 2009 | 9533 | 6068 | 3465 | 8657 | 90.8 | 1932 | 1460 | 4136 | 2005 |
| 2010 | 9474 | 5963 | 3511 | 8787 | 92.7 | 1954 | 1496 | 4009 | 2015 |
| 2011 | 9818 | 6178 | 3640 | 9263 | 94.3 | 2070 | 1635 | 4108 | 2005 |
| 2012 | 10284 | 6247 | 4037 | 9865 | 95.9 | 2279 | 1805 | 3968 | 2232 |
| 2013 | 10300 | 6248 | 4052 | 9998 | 97.1 | 2433 | 1875 | 3815 | 2177 |
| 2014 | 10724 | 6351 | 4373 | 10487 | 97.8 | 2414 | 2090 | 3937 | 2283 |
| Age of diagnosis | | | | | | | | | |
| 15-34 | 656 | 333 | 323 | | | | | | |
| 35-49 | 8182 | 4302 | 3880 | | | | | | |
| 50-64 | 28210 | 16454 | 11756 | | | | | | |
| 65-74 | 27325 | 17731 | 9594 | | | | | | |
| 75-84 | 28508 | 19997 | 8511 | | | | | | |
| 85-94 | 8363 | 5456 | 2907 | | | | | | |

Table S10E. Numbers of cancer patients in TCR by year of diagnosis, age of diagnosis and sex and the number and percentage of them in TCRLF: **oral cancer**

| Year of diagnosis | TCR | | | TCRLF | TCRLF/TCR | 15-64 | | 65-94 | |
|---|---|---|---|---|---|---|---|---|---|
| sex | All | Male | Female | All | % | Male | Female | Male | Female |
| 2004 | 5845 | 5070 | 775 | 3452 | 59.1 | 4191 | 582 | 879 | 193 |
| 2005 | 5862 | 5055 | 807 | 3562 | 60.8 | 4250 | 578 | 805 | 229 |
| 2006 | 6316 | 5508 | 808 | 4028 | 63.8 | 4516 | 591 | 992 | 217 |
| 2007 | 6700 | 5820 | 880 | 4629 | 69.1 | 4837 | 628 | 983 | 252 |
| 2008 | 6778 | 5935 | 843 | 4755 | 70.2 | 4876 | 617 | 1059 | 226 |
| 2009 | 7261 | 6355 | 906 | 6766 | 93.2 | 5263 | 633 | 1092 | 273 |
| 2010 | 7387 | 6481 | 906 | 6973 | 94.4 | 5314 | 645 | 1167 | 261 |
| 2011 | 7522 | 6571 | 951 | 7333 | 97.5 | 5495 | 686 | 1076 | 265 |
| 2012 | 7635 | 6688 | 947 | 7437 | 97.4 | 5546 | 690 | 1142 | 257 |
| 2013 | 7581 | 6677 | 904 | 7463 | 98.4 | 5509 | 619 | 1168 | 285 |
| 2014 | 7856 | 6838 | 1018 | 7760 | 98.8 | 5596 | 730 | 1242 | 288 |
| Age of diagnosis | | | | | | | | | |
| 15-34 | | 2592 | 823 | | | | | | |
| 35-49 | | 23569 | 2761 | | | | | | |
| 50-64 | | 29232 | 3415 | | | | | | |
| 65-74 | | 7787 | 1516 | | | | | | |
| 75-84 | | 3187 | 966 | | | | | | |
| 85-94 | | 631 | 264 | | | | | | |

Table S11A. Number and percentage of patients by stage and year of diagnosis: **breast cancer.**

| Stage | 2004 | | 2005 | | 2006 | | 2007 | | 2008 | | 2009 | | 2010 | | 2011 | | 2012 | | 2013 | | 2014 | |
|---|---|---|---|---|---|---|---|---|---|---|---|---|---|---|---|---|---|---|---|---|---|---|
| | N | % | N | % | N | % | N | % | N | % | N | % | N | % | N | % | N | % | N | % | N | % |
| Localized | 2313 | 53.3 | 2658 | 54.1 | 2741 | 52.0 | 3409 | 53.7 | 3556 | 53.4 | 4050 | 54.7 | 4786 | 55.7 | 4844 | 54.8 | 5267 | 56.6 | 5704 | 57.4 | 6009 | 57.8 |
| regional | 1686 | 38.9 | 1910 | 38.9 | 2073 | 39.3 | 2392 | 37.7 | 2494 | 37.4 | 2716 | 36.7 | 3184 | 37.1 | 3317 | 37.5 | 3300 | 35.5 | 3490 | 35.1 | 3600 | 34.6 |
| Distant | 199 | 4.6 | 210 | 4.3 | 312 | 5.9 | 414 | 6.5 | 478 | 7.2 | 480 | 6.5 | 461 | 5.4 | 475 | 5.4 | 501 | 5.4 | 514 | 5.2 | 587 | 5.6 |
| Non-distant | 22 | 0.5 | 30 | 0.6 | 24 | 0.5 | 22 | 0.3 | 22 | 0.3 | 43 | 0.6 | 92 | 1.1 | 95 | 1.1 | 106 | 1.1 | 138 | 1.4 | 132 | 1.3 |
| Unknown | 117 | 2.7 | 103 | 2.1 | 121 | 2.3 | 107 | 1.7 | 110 | 1.7 | 117 | 1.6 | 63 | 0.7 | 109 | 1.2 | 124 | 1.3 | 90 | 0.9 | 70 | 0.7 |

For patients whose cancer stage cannot be classified into localized, regional and distant stages, we categorize their cancer stage as non-distant stage if the metastatic information can be obtained from TCR-LF. Patients whose cancer stage cannot be classified into localized, regional, distant and non-distant stage is categorized as unknown stage.

Table S11B. Number and percentage of patients by stage and year of diagnosis: **colorectal cancer**.

| Stage | 2004 | | 2005 | | 2006 | | 2007 | | 2008 | | 2009 | | 2010 | | 2011 | | 2012 | | 2013 | | 2014 | |
|---|---|---|---|---|---|---|---|---|---|---|---|---|---|---|---|---|---|---|---|---|---|---|
| | N | % | N | % | N | % | N | % | N | % | N | % | N | % | N | % | N | % | N | % | N | % |
| Localized | 2082 | 36.6 | 2283 | 37.5 | 2484 | 37.9 | 3031 | 38.2 | 3031 | 36.9 | 3476 | 37.6 | 4521 | 40.9 | 4557 | 40.6 | 4904 | 41.4 | 4917 | 41.0 | 5136 | 41.2 |
| regional | 1918 | 33.7 | 2077 | 34.1 | 2129 | 32.5 | 2490 | 31.4 | 2725 | 33.2 | 3080 | 33.3 | 3946 | 35.7 | 4085 | 36.4 | 4099 | 34.6 | 4130 | 34.4 | 4351 | 34.9 |
| Distant | 1143 | 20.1 | 1228 | 20.2 | 1445 | 22.1 | 1905 | 24.0 | 1993 | 24.3 | 2113 | 22.9 | 1949 | 17.6 | 1964 | 17.5 | 2088 | 17.6 | 2113 | 17.6 | 2128 | 17.1 |
| Non-distant | 38 | 0.7 | 14 | 0.2 | 28 | 0.4 | 43 | 0.5 | 33 | 0.4 | 43 | 0.5 | 524 | 4.7 | 468 | 4.2 | 592 | 5.0 | 671 | 5.6 | 705 | 5.7 |
| Unknown | 512 | 9.0 | 485 | 8.0 | 462 | 7.1 | 468 | 5.9 | 435 | 5.3 | 530 | 5.7 | 112 | 1.0 | 162 | 1.4 | 172 | 1.5 | 174 | 1.4 | 154 | 1.2 |

For patients whose cancer stage cannot be classified into localized, regional and distant stages, we categorize their cancer stage as non-distant stage if the metastatic information can be obtained from TCR-LF. Patients whose cancer stage cannot be classified into localized, regional, distant and non-distant stage is categorized as unknown stage. The somewhat large differences in the percentages between 2009 and 2010 might be caused by the difference between AJCC 6$^{th}$ edition and 7$^{th}$ edition; see Table S8.

Table S11C. Number and percentage of patients by stage and year of diagnosis: **liver cancer**

| Stage | 2004 | | 2005 | | 2006 | | 2007 | | 2008 | | 2009 | | 2010 | | 2011 | | 2012 | | 2013 | | 2014 | |
|---|---|---|---|---|---|---|---|---|---|---|---|---|---|---|---|---|---|---|---|---|---|---|
| | N | % | N | % | N | % | N | % | N | % | N | % | N | % | N | % | N | % | N | % | N | % |
| Localized | 5071 | 71.2 | 5580 | 74.6 | 5707 | 71.7 | 6556 | 72.1 | 6785 | 74.0 | 7275 | 75.0 | 5898 | 59.9 | 6007 | 60.1 | 6046 | 60.5 | 6120 | 59.7 | 6001 | 60.7 |
| regional | 585 | 8.2 | 548 | 7.3 | 667 | 8.4 | 847 | 9.3 | 805 | 8.8 | 924 | 9.5 | 2268 | 23.1 | 2233 | 22.4 | 2214 | 22.1 | 2201 | 21.5 | 2110 | 21.3 |
| Distant | 737 | 10.4 | 735 | 9.8 | 910 | 11.4 | 1167 | 12.8 | 1149 | 12.5 | 1160 | 12.0 | 1205 | 12.2 | 1160 | 11.6 | 1101 | 11.0 | 1215 | 11.9 | 1216 | 12.3 |
| Non-distant | 5 | 0.1 | <5 | 0.0 | - | - | 5 | 0.1 | <5 | 0.0 | <5 | 0.0 | 145 | 1.5 | 150 | 1.5 | 164 | 1.6 | 209 | 2.0 | 187 | 1.9 |
| Unknown | 720 | 10.1 | 610 | 8.2 | 672 | 8.4 | 524 | 5.8 | 422 | 4.6 | 343 | 3.5 | 323 | 3.3 | 437 | 4.4 | 474 | 4.7 | 501 | 4.9 | 375 | 3.8 |

For patients whose cancer stage cannot be classified into localized, regional and distant stages, we categorize their cancer stage as non-distant stage if the metastatic information can be obtained from TCR-LF. Patients whose cancer stage cannot be classified into localized, regional, distant and non-distant stage is categorized as unknown stage. The somewhat large differences in the percentages between 2009 and 2010 might be caused by the difference between AJCC 6th edition and 7th edition; see Table S8.

Table S11D. Number and percentage of patients by stage and year of diagnosis: **lung cancer.**

| Stage | 2004 | | 2005 | | 2006 | | 2007 | | 2008 | | 2009 | | 2010 | | 2011 | | 2012 | | 2013 | | 2014 | |
|---|---|---|---|---|---|---|---|---|---|---|---|---|---|---|---|---|---|---|---|---|---|---|
| | N | % | N | % | N | % | N | % | N | % | N | % | N | % | N | % | N | % | N | % | N | % |
| Localized | 628 | 11.1 | 736 | 12.2 | 776 | 12.0 | 983 | 13.0 | 1070 | 13.9 | 1408 | 16.3 | 1287 | 14.6 | 1302 | 14.1 | 1656 | 16.8 | 1788 | 17.9 | 2012 | 19.2 |
| regional | 1665 | 29.4 | 1631 | 27.0 | 1758 | 27.2 | 1971 | 26.2 | 1990 | 25.8 | 2099 | 24.2 | 1410 | 16.0 | 1384 | 14.9 | 1432 | 14.5 | 1383 | 13.8 | 1436 | 13.7 |
| Distant | 3030 | 53.5 | 3223 | 53.3 | 3521 | 54.5 | 4259 | 56.5 | 4381 | 56.7 | 4925 | 56.9 | 5851 | 66.6 | 6287 | 67.9 | 6475 | 65.6 | 6467 | 64.7 | 6743 | 64.3 |
| Non-distant | <5 | 0.0 | | 0.0 | <5 | - | <5 | 0.0 | <5 | 0.0 | | 0.0 | 103 | 1.2 | 86 | 0.9 | 101 | 1.0 | 110 | 1.1 | 113 | 1.1 |
| Unknown | 341 | 6.0 | 457 | 7.6 | 400 | 6.2 | 320 | 4.2 | 278 | 3.6 | 225 | 2.6 | 136 | 1.5 | 204 | 2.2 | 201 | 2.0 | 250 | 2.5 | 183 | 1.7 |

For patients whose cancer stage cannot be classified into localized, regional and distant stages, we categorize their cancer stage as non-distant stage if the metastatic information can be obtained from TCR-LF. Patients whose cancer stage cannot be classified into localized, regional, distant and non-distant stage is categorized as unknown stage. The somewhat large differences in the percentages between 2009 and 2010 might have something to do with the difference between AJCC 6th edition and 7th edition; see Table S8.

Table S11E. Number and percentage of patients by stage and year of diagnosis: **oral cancer.**

| Stage | 2004 | | 2005 | | 2006 | | 2007 | | 2008 | | 2009 | | 2010 | | 2011 | | 2012 | | 2013 | | 2014 | |
|---|---|---|---|---|---|---|---|---|---|---|---|---|---|---|---|---|---|---|---|---|---|---|
| | N | % | N | % | N | % | N | % | N | % | N | % | N | % | N | % | N | % | N | % | N | % |
| Localized | 1312 | 38.0 | 1382 | 38.8 | 1484 | 36.8 | 1779 | 38.4 | 1804 | 37.9 | 2396 | 35.4 | 2394 | 34.3 | 2514 | 34.3 | 2581 | 34.7 | 2573 | 34.5 | 2649 | 34.1 |
| regional | 1886 | 54.6 | 1938 | 54.4 | 2293 | 56.9 | 2592 | 56.0 | 2707 | 56.9 | 3961 | 58.5 | 4146 | 59.5 | 4387 | 59.8 | 4362 | 58.7 | 4402 | 59.0 | 4638 | 59.8 |
| Distant | 83 | 2.4 | 64 | 1.8 | 78 | 1.9 | 117 | 2.5 | 112 | 2.4 | 223 | 3.3 | 242 | 3.5 | 207 | 2.8 | 227 | 3.1 | 243 | 3.3 | 240 | 3.1 |
| Non-distant | <5 | 0.1 | <5 | 0.1 | <5 | - | <5 | 0.0 | <5 | 0.1 | <5 | 0.1 | 80 | 1.1 | 106 | 1.4 | 113 | 1.5 | 132 | 1.8 | 132 | 1.7 |
| Unknown | 167 | 4.8 | 175 | 4.9 | 170 | 4.2 | 140 | 3.0 | 129 | 2.7 | 182 | 2.7 | 111 | 1.6 | 119 | 1.6 | 154 | 2.1 | 113 | 1.5 | 101 | 1.3 |

For patients whose cancer stage cannot be classified into localized, regional and distant stages, we categorize their cancer stage as non-distant stage if the metastatic information can be obtained from TCR-LF. Patients whose cancer stage cannot be classified into localized, regional, distant and non-distant stage is categorized as unknown stage.

Table S12A1. Number of patients and five-year probabilities and the confidence intervals of dying from cancer, dying from other-causes, and survival by stage, age and comorbidity level: **female breast cancer**

| Stage | Age at diagnosis | Comorbidity | N | Survival (%) | | | Cancer deaths (%) | | | Other-cause deaths (%) | | |
|---|---|---|---|---|---|---|---|---|---|---|---|---|
| | | | | Estimate | 95%CI. | | Estimate | 95%CI. | | Estimate | 95%CI. | |
| Localized | 65-74 | 0 | 1592 | 92.0 | 89.5 | 94.0 | 5.7 | 4.4 | 7.2 | 2.3 | 1.6 | 3.3 |
| | | 1 | 2688 | 90.4 | 88.2 | 92.2 | 5.8 | 4.8 | 6.9 | 3.9 | 3.0 | 4.8 |
| | | 2 | 1043 | 80.3 | 75.7 | 84.4 | 8.6 | 6.8 | 10.8 | 11.0 | 8.9 | 13.5 |
| | 75-84 | 0 | 356 | 80.9 | 72.7 | 87.4 | 11.4 | 7.8 | 15.8 | 7.7 | 4.8 | 11.5 |
| | | 1 | 1034 | 81.3 | 76.8 | 85.3 | 8.4 | 6.5 | 10.6 | 10.3 | 8.2 | 12.7 |
| | | 2 | 696 | 57.6 | 50.3 | 64.5 | 16.3 | 13.2 | 19.6 | 26.1 | 22.3 | 30.1 |
| | 85-94 | 0 | 58 | 63.0 | 37.2 | 82.9 | 17.6 | 7.9 | 30.4 | 19.4 | 9.2 | 32.4 |
| | | 1 | 161 | 50.5 | 34.4 | 65.1 | 18.4 | 12.0 | 25.9 | 31.2 | 23.0 | 39.7 |
| | | 2 | 172 | 38.8 | 23.2 | 53.7 | 23.5 | 16.7 | 31.0 | 37.7 | 29.6 | 45.8 |
| Regional | 65-74 | 0 | 1059 | 72.0 | 67.4 | 76.1 | 24.1 | 21.2 | 27.2 | 3.9 | 2.7 | 5.4 |
| | | 1 | 1704 | 73.9 | 70.2 | 77.2 | 20.6 | 18.4 | 22.8 | 5.6 | 4.4 | 6.9 |
| | | 2 | 722 | 66.1 | 59.8 | 71.9 | 20.9 | 17.7 | 24.3 | 13.0 | 10.4 | 15.9 |
| | 75-84 | 0 | 244 | 55.8 | 43.6 | 66.7 | 32.1 | 25.5 | 38.9 | 12.1 | 7.8 | 17.4 |
| | | 1 | 665 | 57.3 | 50.1 | 64.0 | 31.5 | 27.4 | 35.7 | 11.3 | 8.6 | 14.3 |
| | | 2 | 435 | 42.1 | 32.0 | 51.9 | 36.5 | 31.2 | 41.7 | 21.4 | 16.9 | 26.3 |
| | 85-94 | 0 | 40 | 24.0 | -8.6 | 56.7 | 42.4 | 25.4 | 58.4 | 33.6 | 17.8 | 50.3 |
| | | 1 | 116 | 32.0 | 12.7 | 50.8 | 46.3 | 35.7 | 56.3 | 21.6 | 13.5 | 31.0 |
| | | 2 | 105 | 20.8 | -0.8 | 42.8 | 39.9 | 29.1 | 50.4 | 39.4 | 28.1 | 50.4 |
| Distant | 65-74 | 0 | 268 | 17.0 | 9.0 | 25.1 | 79.7 | 73.5 | 84.6 | 3.3 | 1.4 | 6.4 |
| | | 1 | 268 | 17.0 | 8.0 | 26.2 | 76.0 | 69.7 | 81.2 | 7.0 | 4.1 | 10.9 |

| | | | | | | | | | | | | |
|---|---|---|---|---|---|---|---|---|---|---|---|---|
| | | 2 | 119 | 13.7 | -1.4 | 29.5 | 72.6 | 62.5 | 80.4 | 13.8 | 8.0 | 21.0 |
| | 75-84 | 0 | 84 | 9.6 | -7.0 | 27.7 | 78.5 | 66.6 | 86.6 | 11.8 | 5.7 | 20.4 |
| | | 1 | 136 | 12.6 | -0.9 | 27.1 | 77.9 | 68.0 | 85.1 | 9.4 | 4.9 | 15.8 |
| | | 2 | 104 | 17.6 | 0.8 | 34.7 | 69.8 | 58.9 | 78.4 | 12.6 | 6.4 | 20.8 |
| | 85-94 | 0 | 14 | 25.0 | -20.1 | 70.8 | 60.7 | 27.1 | 82.6 | 14.3 | 2.1 | 37.5 |
| | | 1 | 45 | 9.6 | -14.9 | 36.7 | 75.6 | 57.6 | 86.7 | 14.9 | 5.7 | 28.2 |
| | | 2 | 20 | 34.3 | -7.7 | 73.0 | 35.7 | 15.2 | 56.9 | 30.0 | 11.8 | 50.8 |

Table S12A2. One and two-year probabilities and their confidence intervals of dying from cancer, dying from other-causes, and survival by stage, age, and comorbidity level: **female breast cancer**

| Stage | Age at diagnosis | Comorbidity | One-year (%) | | | | | | | | | Two-year (%) | | | | | | | | |
|---|---|---|---|---|---|---|---|---|---|---|---|---|---|---|---|---|---|---|---|---|
| | | | Survival | | | Cancer deaths | | | Other-cause deaths | | | Survival | | | Cancer deaths | | | Other-cause deaths | | |
| | | | Est. | 95%CI. | | Est. | 95%CI. | | Est. | 95%CI. | | Est. | 95%CI. | | Est. | 95%CI. | | Est. | 95%CI. | |
| Localized | 65-74 | 0 | 98.9 | 97.9 | 99.4 | 0.9 | 0.5 | 1.4 | 0.3 | 0.1 | 0.6 | 97.8 | 96.6 | 98.7 | 1.5 | 1.0 | 2.2 | 0.7 | 0.3 | 1.2 |
| | | 1 | 98.8 | 98.1 | 99.3 | 0.6 | 0.3 | 0.9 | 0.6 | 0.4 | 1.0 | 97.3 | 96.3 | 98.1 | 1.3 | 0.9 | 1.8 | 1.4 | 1.0 | 1.9 |
| | | 2 | 97.5 | 95.9 | 98.6 | 1.0 | 0.5 | 1.7 | 1.5 | 0.9 | 2.4 | 94.0 | 91.6 | 95.9 | 2.5 | 1.7 | 3.7 | 3.4 | 2.4 | 4.7 |
| | 75-84 | 0 | 97.5 | 94.2 | 99.1 | 1.7 | 0.7 | 3.5 | 0.8 | 0.2 | 2.3 | 94.5 | 90.2 | 97.2 | 3.2 | 1.7 | 5.5 | 2.3 | 1.1 | 4.3 |
| | | 1 | 98.1 | 96.6 | 99.0 | 0.5 | 0.2 | 1.1 | 1.5 | 0.9 | 2.3 | 95.3 | 93.1 | 96.9 | 2.2 | 1.4 | 3.3 | 2.5 | 1.7 | 3.6 |
| | | 2 | 94.0 | 91.0 | 96.1 | 3.4 | 2.3 | 5.0 | 2.6 | 1.6 | 4.0 | 87.3 | 83.2 | 90.6 | 5.8 | 4.2 | 7.7 | 6.9 | 5.2 | 9.1 |
| | 85-94 | 0 | 93.1 | 78.7 | 98.7 | 3.4 | 0.6 | 10.7 | 3.4 | 0.6 | 10.7 | 85.4 | 67.7 | 95.3 | 5.2 | 1.4 | 13.2 | 9.4 | 3.4 | 19.1 |
| | | 1 | 91.9 | 84.2 | 96.5 | 3.7 | 1.5 | 7.5 | 4.4 | 1.9 | 8.3 | 86.7 | 77.5 | 93.1 | 5.7 | 2.8 | 10.1 | 7.6 | 4.1 | 12.4 |
| | | 2 | 83.7 | 74.6 | 90.4 | 4.1 | 1.8 | 7.8 | 12.2 | 7.8 | 17.6 | 69.4 | 57.9 | 79.0 | 8.4 | 4.8 | 13.3 | 22.2 | 16.2 | 28.8 |
| Regional | 65-74 | 0 | 97.1 | 95.4 | 98.2 | 2.3 | 1.5 | 3.3 | 0.7 | 0.3 | 1.3 | 91.5 | 88.9 | 93.6 | 7.1 | 5.7 | 8.8 | 1.4 | 0.8 | 2.2 |
| | | 1 | 97.5 | 96.3 | 98.3 | 1.8 | 1.3 | 2.5 | 0.7 | 0.4 | 1.2 | 92.8 | 91.0 | 94.4 | 5.9 | 4.8 | 7.1 | 1.3 | 0.8 | 1.9 |

| | | | | | | | | | | | | | | | | | | | | |
|---|---|---|---|---|---|---|---|---|---|---|---|---|---|---|---|---|---|---|---|---|
| | | 2 | 94.5 | 91.7 | 96.5 | 3.2 | 2.1 | 4.7 | 2.4 | 1.4 | 3.7 | 86.8 | 82.8 | 90.1 | 8.6 | 6.7 | 10.9 | 4.6 | 3.2 | 6.3 |
| | 75-84 | 0 | 93.0 | 87.7 | 96.1 | 6.6 | 3.9 | 10.1 | 0.4 | 0.0 | 2.1 | 81.9 | 74.0 | 87.8 | 15.1 | 10.8 | 20.0 | 3.1 | 1.4 | 6.0 |
| | | 1 | 94.9 | 92.1 | 96.8 | 3.8 | 2.5 | 5.4 | 1.4 | 0.7 | 2.5 | 85.5 | 81.2 | 89.0 | 10.7 | 8.5 | 13.3 | 3.8 | 2.5 | 5.5 |
| | | 2 | 87.4 | 82.4 | 91.2 | 9.7 | 7.1 | 12.7 | 3.0 | 1.7 | 4.9 | 73.2 | 66.5 | 79.1 | 19.7 | 16.0 | 23.7 | 7.1 | 4.8 | 9.8 |
| | 85-94 | 0 | 87.4 | 66.6 | 96.7 | 10.1 | 3.2 | 21.9 | 2.5 | 0.2 | 11.4 | 65.2 | 38.4 | 84.3 | 26.7 | 13.6 | 41.6 | 8.2 | 2.0 | 20.0 |
| | | 1 | 88.8 | 78.4 | 95.1 | 7.8 | 3.8 | 13.6 | 3.4 | 1.1 | 8.0 | 74.5 | 61.2 | 84.7 | 20.1 | 13.2 | 28.2 | 5.3 | 2.2 | 10.5 |
| | | 2 | 74.3 | 60.0 | 85.3 | 16.2 | 9.9 | 23.9 | 9.5 | 4.9 | 16.1 | 54.2 | 37.3 | 69.3 | 28.2 | 19.8 | 37.1 | 17.6 | 10.9 | 25.6 |
| Distant | 65-74 | 0 | 74.2 | 68.8 | 79.3 | 25.8 | 20.7 | 31.2 | 0.0 | NA | NA | 53.0 | 47.0 | 59.3 | 47.0 | 40.7 | 53.0 | 0.0 | NA | NA |
| | | 1 | 75.3 | 68.2 | 81.0 | 23.6 | 18.7 | 28.8 | 1.1 | 0.3 | 3.0 | 54.2 | 45.9 | 61.6 | 43.5 | 37.5 | 49.4 | 2.3 | 0.9 | 4.7 |
| | | 2 | 63.7 | 49.9 | 75.0 | 30.4 | 22.4 | 38.8 | 5.9 | 2.6 | 11.2 | 38.0 | 22.9 | 52.2 | 52.5 | 42.7 | 61.3 | 9.5 | 5.0 | 15.8 |
| | 75-84 | 0 | 59.2 | 42.8 | 72.7 | 36.0 | 25.8 | 46.3 | 4.8 | 1.5 | 10.9 | 29.7 | 12.5 | 46.3 | 61.6 | 49.9 | 71.4 | 8.7 | 3.8 | 16.2 |
| | | 1 | 52.6 | 40.0 | 63.7 | 42.9 | 34.5 | 51.1 | 4.4 | 1.8 | 8.9 | 40.6 | 27.5 | 52.8 | 53.4 | 44.4 | 61.5 | 6.0 | 2.8 | 11.0 |
| | | 2 | 64.4 | 50.7 | 75.4 | 32.7 | 23.9 | 41.8 | 2.9 | 0.8 | 7.6 | 36.3 | 20.6 | 51.0 | 55.7 | 45.2 | 64.9 | 8.1 | 3.7 | 14.6 |
| | 85-94 | 0 | 50.0 | 1.9 | 85.8 | 35.7 | 12.0 | 60.6 | 14.3 | 2.1 | 37.5 | 25.0 | -20.1 | 70.8 | 60.7 | 27.1 | 82.6 | 14.3 | 2.1 | 37.5 |
| | | 1 | 42.2 | 19.8 | 61.6 | 53.3 | 37.6 | 66.7 | 4.4 | 0.8 | 13.5 | 30.6 | 6.8 | 53.0 | 60.3 | 44.1 | 73.1 | 9.2 | 2.8 | 20.1 |
| | | 2 | 40.0 | -1.5 | 76.4 | 30.0 | 11.8 | 50.7 | 30.0 | 11.8 | 50.8 | 34.3 | -7.7 | 73.0 | 35.7 | 15.2 | 56.9 | 30.0 | 11.8 | 50.8 |

Table S12B1. Number of patients and five-year probabilities of dying from cancer, dying from other-causes, and survival by stage, age and comorbidity level: **male colorectal cancer**

| Stage | Age at diagnosis | Comorbidity | N | Survival (%) | | | Cancer deaths (%) | | | Other-cause deaths (%) | | |
|---|---|---|---|---|---|---|---|---|---|---|---|---|
| | | | | Estimate | 95%CI. | | Estimate | 95%CI. | | Estimate | 95%CI. | |
| Localized | 65-74 | 0 | 1975 | 80.8 | 77.8 | 83.5 | 14.0 | 12.3 | 15.8 | 5.2 | 4.2 | 6.4 |
| | | 1 | 2657 | 79.7 | 76.9 | 82.2 | 12.2 | 10.8 | 13.7 | 8.1 | 6.9 | 9.4 |
| | | 2 | 1629 | 66.3 | 62.1 | 70.3 | 16.1 | 14.2 | 18.2 | 17.6 | 15.5 | 19.7 |
| | 75-84 | 0 | 1181 | 63.7 | 58.8 | 68.4 | 23.2 | 20.6 | 25.9 | 13.1 | 11.0 | 15.3 |
| | | 1 | 2058 | 61.4 | 57.5 | 65.2 | 22.2 | 20.3 | 24.3 | 16.4 | 14.6 | 18.2 |
| | | 2 | 2025 | 46.0 | 41.7 | 50.2 | 26.1 | 24.1 | 28.2 | 27.8 | 25.7 | 30.0 |
| | 85-94 | 0 | 254 | 34.3 | 21.6 | 46.8 | 38.9 | 32.4 | 45.4 | 26.7 | 20.8 | 33.0 |
| | | 1 | 446 | 43.9 | 34.3 | 53.2 | 30.0 | 25.3 | 34.8 | 26.1 | 21.5 | 30.9 |
| | | 2 | 628 | 25.9 | 17.7 | 34.1 | 38.8 | 34.7 | 42.9 | 35.3 | 31.2 | 39.4 |
| Regional | 65-74 | 0 | 1777 | 53.6 | 49.8 | 57.2 | 41.5 | 39.0 | 44.1 | 4.9 | 3.9 | 6.1 |
| | | 1 | 2036 | 55.8 | 52.2 | 59.3 | 37.3 | 34.9 | 39.6 | 6.9 | 5.8 | 8.2 |
| | | 2 | 1149 | 46.5 | 41.1 | 51.8 | 40.2 | 37.0 | 43.4 | 13.2 | 11.1 | 15.5 |
| | 75-84 | 0 | 1073 | 39.9 | 34.7 | 45.0 | 50.2 | 46.9 | 53.4 | 9.9 | 8.0 | 11.9 |
| | | 1 | 1671 | 39.0 | 34.6 | 43.3 | 49.1 | 46.4 | 51.7 | 11.9 | 10.3 | 13.7 |
| | | 2 | 1489 | 30.2 | 25.3 | 35.0 | 52.3 | 49.5 | 55.0 | 17.6 | 15.5 | 19.7 |
| | 85-94 | 0 | 236 | 25.7 | 13.9 | 37.4 | 58.9 | 51.7 | 65.3 | 15.4 | 10.8 | 20.8 |
| | | 1 | 384 | 17.4 | 7.8 | 27.1 | 63.7 | 58.0 | 68.8 | 18.9 | 14.8 | 23.4 |
| | | 2 | 494 | 13.5 | 4.8 | 22.4 | 61.2 | 56.4 | 65.7 | 25.2 | 21.2 | 29.5 |
| Distant | 65-74 | 0 | 1023 | 9.3 | 5.9 | 12.9 | 86.1 | 83.7 | 88.2 | 4.6 | 3.4 | 6.0 |
| | | 1 | 1095 | 9.3 | 6.0 | 12.6 | 87.1 | 84.8 | 89.1 | 3.6 | 2.6 | 4.9 |

| | | | | | | | | | | | | |
|---|---|---|---|---|---|---|---|---|---|---|---|---|
| | | 2 | 626 | 5.9 | 1.2 | 10.8 | 86.8 | 83.8 | 89.3 | 7.3 | 5.4 | 9.5 |
| | 75-84 | 0 | 625 | 4.2 | 0.2 | 8.3 | 91.2 | 88.6 | 93.3 | 4.6 | 3.1 | 6.5 |
| | | 1 | 1032 | 4.1 | 1.0 | 7.3 | 91.0 | 89.0 | 92.6 | 4.9 | 3.7 | 6.4 |
| | | 2 | 925 | 3.7 | -0.2 | 7.8 | 87.9 | 85.6 | 89.9 | 8.4 | 6.7 | 10.3 |
| | 85-94 | 0 | 152 | 0.7 | -7.0 | 9.3 | 93.4 | 87.8 | 96.5 | 6.0 | 2.9 | 10.5 |
| | | 1 | 222 | 1.5 | -6.7 | 10.3 | 88.2 | 83.0 | 91.9 | 10.3 | 6.6 | 14.8 |
| | | 2 | 322 | 0.9 | -5.7 | 7.8 | 89.5 | 85.6 | 92.4 | 9.6 | 6.7 | 13.3 |

Table S12B2. One and Two-year probabilities of dying from cancer, dying from other-causes, and survival by stage, age and comorbidity level: **male colorectal cancer**

| Stage | Age at diagnosis | Comorbidity | One-year (%) | | | | | | | | | Two-year (%) | | | | | | | | |
|---|---|---|---|---|---|---|---|---|---|---|---|---|---|---|---|---|---|---|---|---|
| | | | Survival | | | Cancer deaths | | | Other-cause deaths | | | Survival | | | Cancer deaths | | | Other-cause deaths | | |
| | | | Est. | 95%CI. | | Est. | 95%CI. | | Est. | 95%CI. | | Est. | 95%CI. | | Est. | 95%CI. | | Est. | 95%CI. | |
| Localized | 65-74 | 0 | 97.1 | 95.9 | 98.0 | 2.1 | 1.6 | 2.8 | 0.8 | 0.4 | 1.2 | 93.3 | 91.6 | 94.7 | 4.9 | 4.0 | 6.0 | 1.8 | 1.3 | 2.5 |
| | | 1 | 96.3 | 95.2 | 97.3 | 2.2 | 1.7 | 2.8 | 1.4 | 1.0 | 1.9 | 93.1 | 91.6 | 94.4 | 4.3 | 3.5 | 5.1 | 2.6 | 2.1 | 3.3 |
| | | 2 | 91.3 | 89.1 | 93.1 | 5.1 | 4.1 | 6.2 | 3.6 | 2.8 | 4.6 | 85.1 | 82.4 | 87.6 | 8.4 | 7.1 | 9.8 | 6.5 | 5.3 | 7.8 |
| | 75-84 | 0 | 91.9 | 89.5 | 93.8 | 6.1 | 4.8 | 7.6 | 2.0 | 1.3 | 3.0 | 85.2 | 82.0 | 88.0 | 10.6 | 8.9 | 12.5 | 4.2 | 3.2 | 5.5 |
| | | 1 | 91.4 | 89.6 | 93.0 | 5.8 | 4.9 | 6.9 | 2.7 | 2.1 | 3.5 | 85.8 | 83.4 | 87.9 | 9.6 | 8.4 | 11.0 | 4.6 | 3.7 | 5.6 |
| | | 2 | 83.1 | 80.6 | 85.4 | 10.2 | 8.9 | 11.5 | 6.7 | 5.7 | 7.9 | 72.9 | 69.8 | 75.8 | 15.2 | 13.6 | 16.8 | 11.9 | 10.5 | 13.4 |
| | 85-94 | 0 | 81.1 | 73.2 | 87.4 | 12.6 | 8.9 | 17.0 | 6.3 | 3.8 | 9.7 | 69.3 | 60.0 | 77.4 | 21.5 | 16.6 | 26.8 | 9.2 | 6.0 | 13.3 |
| | | 1 | 82.7 | 77.2 | 87.3 | 12.8 | 9.9 | 16.1 | 4.5 | 2.8 | 6.7 | 73.9 | 67.2 | 79.8 | 16.7 | 13.4 | 20.4 | 9.4 | 6.9 | 12.4 |
| | | 2 | 73.1 | 67.5 | 78.1 | 16.9 | 14.1 | 19.9 | 10.0 | 7.8 | 12.5 | 54.6 | 47.8 | 61.1 | 27.7 | 24.1 | 31.3 | 17.7 | 14.8 | 20.9 |
| Regional | 65-74 | 0 | 89.9 | 87.9 | 91.6 | 9.0 | 7.7 | 10.4 | 1.1 | 0.7 | 1.7 | 78.3 | 75.7 | 80.7 | 19.8 | 17.9 | 21.7 | 1.9 | 1.3 | 2.6 |
| | | 1 | 90.5 | 88.7 | 92.1 | 8.0 | 6.8 | 9.2 | 1.5 | 1.1 | 2.1 | 79.7 | 77.2 | 82.1 | 17.4 | 15.8 | 19.1 | 2.8 | 2.2 | 3.6 |

| | | | | | | | | | | | | | | | | | | | | |
|---|---|---|---|---|---|---|---|---|---|---|---|---|---|---|---|---|---|---|---|---|
| | | 2 | 86.3 | 83.3 | 88.9 | 10.5 | 8.8 | 12.3 | 3.2 | 2.3 | 4.4 | 74.0 | 70.1 | 77.5 | 20.5 | 18.2 | 22.9 | 5.6 | 4.3 | 7.0 |
| | 75-84 | 0 | 81.8 | 78.6 | 84.7 | 16.0 | 13.9 | 18.3 | 2.1 | 1.4 | 3.1 | 67.3 | 63.3 | 71.1 | 28.6 | 25.9 | 31.3 | 4.1 | 3.0 | 5.4 |
| | | 1 | 81.0 | 78.2 | 83.4 | 15.6 | 13.9 | 17.4 | 3.4 | 2.6 | 4.4 | 66.6 | 63.2 | 69.8 | 27.9 | 25.7 | 30.1 | 5.5 | 4.5 | 6.7 |
| | | 2 | 74.5 | 71.2 | 77.5 | 20.6 | 18.6 | 22.7 | 4.8 | 3.8 | 6.0 | 58.2 | 54.3 | 62.0 | 33.8 | 31.3 | 36.2 | 8.0 | 6.7 | 9.5 |
| | 85-94 | 0 | 66.5 | 57.4 | 74.3 | 28.8 | 23.2 | 34.7 | 4.7 | 2.5 | 7.9 | 53.5 | 43.5 | 62.5 | 39.7 | 33.4 | 45.9 | 6.9 | 4.1 | 10.6 |
| | | 1 | 65.6 | 58.3 | 72.1 | 28.1 | 23.7 | 32.7 | 6.3 | 4.1 | 9.0 | 48.5 | 40.2 | 56.3 | 41.2 | 36.2 | 46.2 | 10.3 | 7.5 | 13.6 |
| | | 2 | 57.3 | 50.3 | 63.8 | 32.9 | 28.8 | 37.1 | 9.8 | 7.3 | 12.6 | 40.2 | 32.5 | 47.7 | 44.8 | 40.3 | 49.2 | 14.9 | 11.9 | 18.3 |
| Distant | 65-74 | 0 | 56.2 | 52.1 | 60.0 | 41.4 | 38.4 | 44.5 | 2.4 | 1.5 | 3.4 | 32.7 | 28.6 | 36.8 | 63.7 | 60.7 | 66.6 | 3.5 | 2.5 | 4.8 |
| | | 1 | 54.3 | 50.5 | 58.0 | 43.7 | 40.7 | 46.6 | 2.0 | 1.3 | 3.0 | 30.4 | 26.5 | 34.2 | 66.6 | 63.7 | 69.3 | 3.1 | 2.2 | 4.2 |
| | | 2 | 44.2 | 38.5 | 49.7 | 51.0 | 47.0 | 54.8 | 4.8 | 3.3 | 6.7 | 22.1 | 16.6 | 27.6 | 71.3 | 67.6 | 74.7 | 6.6 | 4.8 | 8.7 |
| | 75-84 | 0 | 38.5 | 33.1 | 43.7 | 58.2 | 54.2 | 61.9 | 3.4 | 2.2 | 5.0 | 18.5 | 13.8 | 23.3 | 77.8 | 74.3 | 80.9 | 3.7 | 2.4 | 5.4 |
| | | 1 | 41.0 | 36.9 | 45.1 | 55.8 | 52.7 | 58.7 | 3.2 | 2.3 | 4.4 | 20.2 | 16.3 | 24.0 | 75.9 | 73.2 | 78.5 | 3.9 | 2.8 | 5.2 |
| | | 2 | 32.6 | 28.0 | 37.2 | 61.6 | 58.4 | 64.7 | 5.7 | 4.4 | 7.4 | 14.9 | 10.6 | 19.3 | 78.1 | 75.3 | 80.6 | 7.0 | 5.4 | 8.7 |
| | 85-94 | 0 | 21.9 | 11.4 | 32.3 | 73.5 | 65.6 | 79.8 | 4.6 | 2.0 | 8.8 | 9.3 | 0.2 | 18.9 | 85.4 | 78.7 | 90.2 | 5.3 | 2.5 | 9.7 |
| | | 1 | 24.4 | 15.0 | 33.6 | 69.3 | 62.7 | 74.9 | 6.4 | 3.6 | 10.1 | 12.0 | 3.3 | 21.1 | 80.2 | 74.2 | 84.9 | 7.8 | 4.7 | 11.8 |
| | | 2 | 21.7 | 14.0 | 29.4 | 71.4 | 66.2 | 76.0 | 6.9 | 4.5 | 10.0 | 7.6 | 0.7 | 14.6 | 84.3 | 79.9 | 87.9 | 8.1 | 5.5 | 11.4 |

Table S12C1. Number of patients and five-year probabilities of dying from cancer, dying from other-causes, and survival by stage, age and comorbidity level: **female colorectal cancer**

| Stage | Age at diagnosis | Comorbidity | N | Survival (%) | | | Cancer deaths (%) | | | Other-cause deaths (%) | | |
|---|---|---|---|---|---|---|---|---|---|---|---|---|
| | | | | Estimate | 95%CI. | | Estimate | 95%CI. | | Estimate | 95%CI. | |
| Localized | 65-74 | 0 | 1257 | 86.5 | 83.3 | 89.3 | 9.8 | 8.0 | 11.8 | 3.7 | 2.6 | 5.0 |
| | | 1 | 2021 | 85.2 | 82.5 | 87.6 | 9.9 | 8.5 | 11.5 | 4.9 | 3.9 | 6.0 |
| | | 2 | 998 | 70.4 | 65.2 | 75.2 | 13.0 | 10.8 | 15.5 | 16.6 | 14.0 | 19.3 |
| | 75-84 | 0 | 698 | 71.8 | 66.1 | 76.9 | 21.7 | 18.4 | 25.1 | 6.5 | 4.7 | 8.8 |
| | | 1 | 1732 | 71.0 | 67.1 | 74.6 | 17.5 | 15.5 | 19.6 | 11.5 | 9.9 | 13.3 |
| | | 2 | 1172 | 52.4 | 46.9 | 57.7 | 27.0 | 24.2 | 29.8 | 20.6 | 18.1 | 23.3 |
| | 85-94 | 0 | 222 | 35.9 | 23.1 | 48.4 | 45.5 | 38.4 | 52.4 | 18.5 | 13.2 | 24.5 |
| | | 1 | 458 | 47.5 | 38.1 | 56.4 | 31.0 | 26.3 | 35.8 | 21.5 | 17.3 | 26.1 |
| | | 2 | 500 | 27.0 | 17.4 | 36.5 | 37.1 | 32.5 | 41.8 | 35.9 | 31.0 | 40.8 |
| Regional | 65-74 | 0 | 1190 | 60.9 | 56.6 | 64.9 | 35.6 | 32.6 | 38.6 | 3.5 | 2.5 | 4.8 |
| | | 1 | 1651 | 63.7 | 60.0 | 67.2 | 32.1 | 29.6 | 34.6 | 4.2 | 3.2 | 5.4 |
| | | 2 | 709 | 51.7 | 45.1 | 57.9 | 38.4 | 34.5 | 42.3 | 9.9 | 7.6 | 12.5 |
| | 75-84 | 0 | 746 | 45.3 | 39.3 | 51.0 | 47.7 | 43.8 | 51.6 | 7.0 | 5.1 | 9.1 |
| | | 1 | 1537 | 46.0 | 41.6 | 50.2 | 46.0 | 43.2 | 48.7 | 8.0 | 6.6 | 9.7 |
| | | 2 | 1069 | 35.8 | 30.1 | 41.3 | 49.5 | 46.2 | 52.8 | 14.7 | 12.5 | 17.1 |
| | 85-94 | 0 | 199 | 19.0 | 6.9 | 31.3 | 67.0 | 59.4 | 73.5 | 14.0 | 9.3 | 19.6 |
| | | 1 | 466 | 22.4 | 14.0 | 30.9 | 61.0 | 56.0 | 65.7 | 16.5 | 13.1 | 20.4 |
| | | 2 | 394 | 16.1 | 6.4 | 25.9 | 58.5 | 53.1 | 63.4 | 25.5 | 21.0 | 30.2 |
| Distant | 65-74 | 0 | 612 | 12.7 | 8.5 | 16.9 | 85.3 | 82.0 | 88.1 | 2.0 | 1.1 | 3.4 |
| | | 1 | 803 | 9.4 | 5.6 | 13.2 | 87.1 | 84.4 | 89.4 | 3.5 | 2.4 | 5.0 |

| | | | | | | | | | | | | |
|---|---|---|---|---|---|---|---|---|---|---|---|---|
| | | 2 | 392 | 8.0 | 2.1 | 14.2 | 85.7 | 81.7 | 89.0 | 6.2 | 4.1 | 8.9 |
| | 75-84 | 0 | 448 | 8.2 | 3.3 | 13.4 | 87.9 | 84.3 | 90.7 | 3.9 | 2.4 | 6.1 |
| | | 1 | 854 | 6.5 | 2.9 | 10.2 | 89.1 | 86.6 | 91.1 | 4.4 | 3.2 | 6.0 |
| | | 2 | 600 | 2.0 | -2.5 | 6.6 | 90.4 | 87.7 | 92.5 | 7.6 | 5.7 | 10.0 |
| | 85-94 | 0 | 160 | 1.2 | -9.7 | 12.8 | 85.5 | 78.9 | 90.2 | 13.3 | 8.3 | 19.5 |
| | | 1 | 281 | 4.2 | -2.8 | 11.6 | 88.0 | 83.4 | 91.5 | 7.8 | 5.0 | 11.4 |
| | | 2 | 260 | 1.6 | -7.0 | 10.9 | 84.0 | 78.6 | 88.1 | 14.4 | 10.5 | 18.9 |

Table S12C2. One and two-year probabilities of dying from cancer, dying from other-causes, and survival by stage, age and comorbidity level: **female colorectal cancer**

| Stage | Age at diagnosis | Comorbidity | One-year (%) | | | | | | | | | Two-year (%) | | | | | | | | |
|---|---|---|---|---|---|---|---|---|---|---|---|---|---|---|---|---|---|---|---|---|
| | | | Survival | | | Cancer deaths | | | Other-cause deaths | | | Survival | | | Cancer deaths | | | Other-cause deaths | | |
| | | | Est. | 95%CI. | | Est. | 95%CI. | | Est. | 95%CI. | | Est. | 95%CI. | | Est. | 95%CI. | | Est. | 95%CI. | |
| Localized | 65-74 | 0 | 97.9 | 96.6 | 98.8 | 1.3 | 0.8 | 2.0 | 0.8 | 0.4 | 1.4 | 95.2 | 93.3 | 96.7 | 3.1 | 2.3 | 4.2 | 1.6 | 1.0 | 2.5 |
| | | 1 | 97.7 | 96.7 | 98.5 | 1.5 | 1.0 | 2.1 | 0.8 | 0.5 | 1.3 | 95.4 | 94.0 | 96.6 | 3.1 | 2.4 | 3.9 | 1.5 | 1.0 | 2.1 |
| | | 2 | 91.9 | 89.2 | 94.1 | 4.8 | 3.6 | 6.3 | 3.3 | 2.3 | 4.6 | 87.8 | 84.6 | 90.6 | 6.9 | 5.4 | 8.6 | 5.3 | 4.0 | 6.9 |
| | 75-84 | 0 | 93.2 | 90.3 | 95.4 | 5.3 | 3.8 | 7.2 | 1.4 | 0.7 | 2.5 | 88.4 | 84.7 | 91.3 | 9.4 | 7.4 | 11.8 | 2.2 | 1.3 | 3.5 |
| | | 1 | 94.2 | 92.5 | 95.6 | 4.0 | 3.1 | 5.0 | 1.8 | 1.2 | 2.5 | 88.2 | 85.8 | 90.3 | 7.4 | 6.3 | 8.8 | 4.3 | 3.4 | 5.4 |
| | | 2 | 86.1 | 83.0 | 88.8 | 8.2 | 6.7 | 9.9 | 5.7 | 4.5 | 7.1 | 76.8 | 73.0 | 80.3 | 14.1 | 12.2 | 16.2 | 9.1 | 7.5 | 10.8 |
| | 85-94 | 0 | 73.3 | 64.3 | 80.8 | 22.1 | 16.9 | 27.8 | 4.5 | 2.3 | 7.8 | 58.4 | 47.9 | 67.9 | 32.8 | 26.6 | 39.1 | 8.8 | 5.5 | 13.0 |
| | | 1 | 83.4 | 78.0 | 87.9 | 11.6 | 8.8 | 14.7 | 5.0 | 3.3 | 7.3 | 73.8 | 67.3 | 79.6 | 17.6 | 14.3 | 21.3 | 8.5 | 6.2 | 11.4 |
| | | 2 | 69.5 | 63.0 | 75.4 | 19.0 | 15.7 | 22.6 | 11.4 | 8.8 | 14.4 | 57.0 | 49.5 | 64.0 | 26.4 | 22.6 | 30.4 | 16.6 | 13.4 | 20.0 |
| Regional | 65-74 | 0 | 92.9 | 90.8 | 94.6 | 6.5 | 5.2 | 8.0 | 0.6 | 0.3 | 1.2 | 82.2 | 79.1 | 84.9 | 16.2 | 14.1 | 18.4 | 1.6 | 1.0 | 2.5 |
| | | 1 | 92.9 | 91.1 | 94.4 | 6.3 | 5.2 | 7.6 | 0.8 | 0.4 | 1.3 | 83.1 | 80.5 | 85.3 | 15.3 | 13.6 | 17.1 | 1.6 | 1.1 | 2.3 |

| | | | | | | | | | | | | | | | | | | | | |
|---|---|---|---|---|---|---|---|---|---|---|---|---|---|---|---|---|---|---|---|---|
| | | 2 | 86.4 | 82.7 | 89.6 | 11.2 | 9.0 | 13.6 | 2.4 | 1.5 | 3.7 | 74.7 | 69.9 | 78.9 | 21.9 | 18.9 | 25.1 | 3.5 | 2.3 | 5.0 |
| | | 0 | 84.2 | 80.5 | 87.4 | 14.0 | 11.6 | 16.6 | 1.7 | 1.0 | 2.9 | 69.1 | 64.4 | 73.5 | 27.4 | 24.2 | 30.7 | 3.4 | 2.3 | 4.9 |
| | 75-84 | 1 | 83.2 | 80.6 | 85.5 | 14.9 | 13.2 | 16.7 | 1.9 | 1.3 | 2.7 | 68.2 | 64.9 | 71.3 | 28.5 | 26.2 | 30.8 | 3.3 | 2.5 | 4.3 |
| | | 2 | 76.4 | 72.7 | 79.8 | 19.1 | 16.8 | 21.5 | 4.5 | 3.4 | 5.9 | 60.7 | 56.1 | 65.0 | 31.9 | 29.0 | 34.7 | 7.4 | 6.0 | 9.1 |
| | | 0 | 58.8 | 48.5 | 67.8 | 36.7 | 30.0 | 43.4 | 4.5 | 2.2 | 8.1 | 39.5 | 28.5 | 49.9 | 52.9 | 45.6 | 59.6 | 7.7 | 4.5 | 11.9 |
| | 85-94 | 1 | 63.9 | 57.3 | 70.0 | 29.8 | 25.7 | 34.0 | 6.2 | 4.3 | 8.7 | 42.6 | 35.1 | 49.9 | 46.6 | 42.0 | 51.1 | 10.7 | 8.1 | 13.8 |
| | | 2 | 55.0 | 47.1 | 62.4 | 35.1 | 30.4 | 39.8 | 9.9 | 7.2 | 13.1 | 38.1 | 29.4 | 46.6 | 46.0 | 41.0 | 50.9 | 15.9 | 12.4 | 19.7 |
| | | 0 | 56.1 | 51.4 | 60.5 | 43.4 | 39.4 | 47.3 | 0.5 | 0.1 | 1.4 | 32.6 | 27.9 | 37.1 | 66.4 | 62.5 | 70.0 | 1.0 | 0.4 | 2.1 |
| | 65-74 | 1 | 52.7 | 48.0 | 57.1 | 44.7 | 41.2 | 48.1 | 2.6 | 1.7 | 3.9 | 29.0 | 24.5 | 33.5 | 67.7 | 64.3 | 70.9 | 3.3 | 2.2 | 4.7 |
| | | 2 | 49.2 | 42.2 | 55.8 | 47.2 | 42.2 | 52.0 | 3.6 | 2.0 | 5.8 | 23.3 | 16.5 | 30.0 | 71.3 | 66.6 | 75.6 | 5.4 | 3.4 | 7.9 |
| | | 0 | 40.6 | 34.5 | 46.4 | 57.2 | 52.5 | 61.6 | 2.2 | 1.1 | 3.9 | 21.8 | 16.1 | 27.3 | 75.3 | 71.0 | 79.1 | 2.9 | 1.6 | 4.8 |
| Distant | 75-84 | 1 | 37.7 | 33.2 | 42.1 | 59.5 | 56.1 | 62.7 | 2.8 | 1.9 | 4.1 | 19.6 | 15.4 | 23.7 | 76.6 | 73.5 | 79.3 | 3.9 | 2.7 | 5.3 |
| | | 2 | 28.7 | 23.0 | 34.2 | 65.8 | 61.9 | 69.5 | 5.5 | 3.9 | 7.5 | 13.0 | 7.9 | 18.1 | 80.9 | 77.5 | 83.8 | 6.2 | 4.4 | 8.3 |
| | | 0 | 21.4 | 9.5 | 33.4 | 67.9 | 60.1 | 74.5 | 10.7 | 6.5 | 16.0 | 9.1 | -1.8 | 20.6 | 79.5 | 72.4 | 85.0 | 11.3 | 7.0 | 16.8 |
| | 85-94 | 1 | 23.0 | 15.3 | 30.7 | 72.3 | 66.7 | 77.2 | 4.7 | 2.6 | 7.6 | 9.2 | 2.0 | 16.6 | 84.3 | 79.4 | 88.1 | 6.5 | 4.0 | 9.9 |
| | | 2 | 17.5 | 8.2 | 27.0 | 70.5 | 64.6 | 75.6 | 12.0 | 8.4 | 16.2 | 5.2 | -3.6 | 14.5 | 80.4 | 75.1 | 84.7 | 14.4 | 10.5 | 18.9 |

Table S12D1. Number of patients and five-year probabilities of dying from cancer, dying from other-causes, and survival by stage, age and comorbidity level: **male liver cancer**

| Stage | Age at diagnosis | Comorbidity | N | Survival (%) | | | Cancer deaths (%) | | | Other-cause deaths (%) | | |
|---|---|---|---|---|---|---|---|---|---|---|---|---|
| | | | | Estimate | 95%CI. | | Estimate | 95%CI. | | Estimate | 95%CI. | |
| Localized | 65-74 | 0 | 2180 | 23.6 | 20.6 | 26.6 | 71.2 | 69.1 | 73.2 | 5.2 | 4.3 | 6.2 |
| | | 1 | 5852 | 37.0 | 35.0 | 39.0 | 57.9 | 56.5 | 59.3 | 5.2 | 4.6 | 5.8 |
| | | 2 | 3942 | 25.5 | 22.9 | 28.1 | 65.2 | 63.5 | 66.8 | 9.3 | 8.4 | 10.3 |
| | 75-84 | 0 | 984 | 16.4 | 11.9 | 21.0 | 76.6 | 73.6 | 79.3 | 7.0 | 5.4 | 8.8 |
| | | 1 | 3095 | 25.1 | 22.2 | 28.0 | 66.1 | 64.3 | 67.9 | 8.8 | 7.8 | 9.9 |
| | | 2 | 3107 | 15.5 | 12.4 | 18.6 | 69.8 | 68.0 | 71.5 | 14.7 | 13.4 | 16.1 |
| | 85-94 | 0 | 167 | 6.8 | -3.4 | 17.6 | 84.8 | 77.7 | 89.8 | 8.4 | 4.7 | 13.6 |
| | | 1 | 465 | 12.2 | 4.9 | 19.7 | 75.7 | 71.2 | 79.7 | 12.1 | 9.2 | 15.4 |
| | | 2 | 628 | 5.1 | -1.5 | 11.9 | 75.8 | 72.0 | 79.1 | 19.2 | 16.1 | 22.5 |
| Regional | 65-74 | 0 | 801 | 5.2 | 2.2 | 8.2 | 93.0 | 90.8 | 94.7 | 1.8 | 1.0 | 3.0 |
| | | 1 | 1110 | 6.6 | 3.4 | 9.8 | 89.3 | 87.1 | 91.1 | 4.2 | 3.1 | 5.5 |
| | | 2 | 646 | 4.7 | 0.8 | 8.9 | 91.3 | 88.5 | 93.4 | 4.0 | 2.7 | 5.8 |
| | 75-84 | 0 | 356 | 3.7 | -2.4 | 10.1 | 88.7 | 84.8 | 91.7 | 7.6 | 5.1 | 10.8 |
| | | 1 | 645 | 5.2 | 1.2 | 9.5 | 90.1 | 87.3 | 92.3 | 4.7 | 3.2 | 6.5 |
| | | 2 | 608 | 2.3 | -2.2 | 6.9 | 90.7 | 87.9 | 92.9 | 7.0 | 5.1 | 9.3 |
| | 85-94 | 0 | 55 | 1.8 | -7.8 | 16.7 | 96.4 | 83.2 | 99.3 | 1.8 | 0.1 | 8.6 |
| | | 1 | 137 | 0.9 | -7.2 | 10.2 | 93.3 | 87.1 | 96.5 | 5.9 | 2.7 | 10.7 |
| | | 2 | 158 | 1.1 | -9.1 | 12.5 | 88.1 | 81.2 | 92.7 | 10.7 | 6.3 | 16.4 |
| Distant | 65-74 | 0 | 674 | 0.2 | -2.4 | 2.9 | 97.0 | 95.3 | 98.0 | 2.9 | 1.8 | 4.3 |
| | | 1 | 750 | 1.8 | -1.1 | 4.9 | 94.7 | 92.8 | 96.1 | 3.5 | 2.4 | 5.0 |

| | | | | | | | | | | | | |
|---|---|---|---|---|---|---|---|---|---|---|---|---|
| | | 2 | 509 | 1.7 | -2.2 | 5.9 | 93.5 | 90.9 | 95.4 | 4.8 | 3.1 | 6.8 |
| | 75-84 | 0 | 269 | 1.5 | -3.9 | 7.4 | 93.6 | 89.8 | 96.0 | 4.9 | 2.7 | 7.9 |
| | | 1 | 472 | 1.2 | -2.9 | 5.5 | 94.1 | 91.4 | 96.0 | 4.7 | 3.1 | 6.9 |
| | | 2 | 488 | 0.8 | -3.6 | 5.5 | 93.0 | 90.2 | 95.0 | 6.2 | 4.3 | 8.6 |
| | 85-94 | 0 | 63 | 5.0 | -8.2 | 19.8 | 90.1 | 79.0 | 95.5 | 4.8 | 1.2 | 12.6 |
| | | 1 | 107 | 1.9 | -7.3 | 12.2 | 93.3 | 86.1 | 96.8 | 4.8 | 1.7 | 10.5 |
| | | 2 | 115 | 0.9 | -7.5 | 10.6 | 93.9 | 87.3 | 97.1 | 5.2 | 2.1 | 10.4 |

Table S12D2. One and two-year probabilities of dying from cancer, dying from other-causes, and survival by stage, age and comorbidity level for **male liver cancer**

| Stage | Age at diagnosis | Comorbidity | One-year (%) | | | | | | | | | Two-year (%) | | | | | | | | |
|---|---|---|---|---|---|---|---|---|---|---|---|---|---|---|---|---|---|---|---|---|
| | | | Survival | | | Cancer deaths | | | Other-cause deaths | | | Survival | | | Cancer deaths | | | Other-cause deaths | | |
| | | | Est. | 95%CI. | | Est. | 95%CI. | | Est. | 95%CI. | | Est. | 95%CI. | | Est. | 95%CI. | | Est. | 95%CI. | |
| Localized | 65-74 | 0 | 59.9 | 57.2 | 62.5 | 38.1 | 36.0 | 40.1 | 2.0 | 1.5 | 2.7 | 45.7 | 42.8 | 48.5 | 51.0 | 48.9 | 53.1 | 3.3 | 2.6 | 4.1 |
| | | 1 | 77.5 | 76.1 | 78.9 | 20.9 | 19.8 | 21.9 | 1.6 | 1.3 | 1.9 | 64.4 | 62.7 | 66.0 | 33.1 | 31.9 | 34.3 | 2.6 | 2.2 | 3.0 |
| | | 2 | 71.2 | 69.3 | 73.1 | 25.9 | 24.6 | 27.3 | 2.9 | 2.4 | 3.4 | 54.8 | 52.5 | 57.0 | 40.3 | 38.8 | 41.9 | 4.9 | 4.3 | 5.6 |
| | 75-84 | 0 | 50.9 | 46.6 | 55.1 | 45.9 | 42.7 | 49.0 | 3.2 | 2.2 | 4.4 | 37.8 | 33.4 | 42.1 | 58.0 | 54.8 | 61.0 | 4.2 | 3.1 | 5.6 |
| | | 1 | 68.6 | 66.4 | 70.7 | 28.7 | 27.1 | 30.3 | 2.7 | 2.2 | 3.4 | 53.0 | 50.5 | 55.5 | 42.7 | 40.9 | 44.4 | 4.3 | 3.6 | 5.1 |
| | | 2 | 59.6 | 57.0 | 62.1 | 34.0 | 32.3 | 35.6 | 6.4 | 5.6 | 7.3 | 42.7 | 39.9 | 45.4 | 48.4 | 46.6 | 50.1 | 8.9 | 8.0 | 10.0 |
| | 85-94 | 0 | 35.0 | 23.8 | 45.7 | 59.5 | 51.6 | 66.6 | 5.4 | 2.7 | 9.6 | 23.4 | 12.8 | 34.0 | 70.5 | 62.9 | 76.9 | 6.0 | 3.1 | 10.4 |
| | | 1 | 52.9 | 46.2 | 59.2 | 41.7 | 37.2 | 46.2 | 5.4 | 3.6 | 7.7 | 34.1 | 27.0 | 41.1 | 57.8 | 53.1 | 62.2 | 8.1 | 5.8 | 10.8 |
| | | 2 | 47.4 | 41.1 | 53.4 | 43.3 | 39.4 | 47.1 | 9.3 | 7.2 | 11.7 | 26.9 | 20.4 | 33.4 | 59.8 | 55.8 | 63.5 | 13.3 | 10.8 | 16.1 |
| Regional | 65-74 | 0 | 24.2 | 20.4 | 27.9 | 74.7 | 71.5 | 77.6 | 1.1 | 0.6 | 2.1 | 14.0 | 10.6 | 17.3 | 84.8 | 82.1 | 87.1 | 1.3 | 0.6 | 2.2 |
| | | 1 | 28.8 | 25.2 | 32.4 | 68.8 | 66.0 | 71.5 | 2.3 | 1.6 | 3.4 | 15.7 | 12.3 | 19.1 | 80.9 | 78.5 | 83.1 | 3.4 | 2.4 | 4.6 |

| | | | | | | | | | | | | | | | | | | | | |
|---|---|---|---|---|---|---|---|---|---|---|---|---|---|---|---|---|---|---|---|---|
| | | 2 | 26.6 | 21.8 | 31.2 | 71.0 | 67.3 | 74.3 | 2.5 | 1.5 | 3.9 | 13.8 | 9.4 | 18.3 | 82.5 | 79.3 | 85.3 | 3.6 | 2.4 | 5.3 |
| | 75-84 | 0 | 21.9 | 15.0 | 28.8 | 72.8 | 67.8 | 77.0 | 5.3 | 3.3 | 8.0 | 10.2 | 4.0 | 16.7 | 83.9 | 79.5 | 87.3 | 5.9 | 3.8 | 8.7 |
| | | 1 | 25.6 | 20.6 | 30.5 | 70.8 | 67.1 | 74.1 | 3.6 | 2.3 | 5.2 | 12.6 | 8.2 | 17.1 | 83.3 | 80.2 | 86.0 | 4.1 | 2.7 | 5.8 |
| | | 2 | 19.5 | 14.5 | 24.4 | 76.3 | 72.7 | 79.4 | 4.3 | 2.9 | 6.1 | 10.6 | 6.0 | 15.3 | 84.2 | 81.1 | 86.9 | 5.2 | 3.6 | 7.1 |
| | 85-94 | 0 | 18.2 | 2.9 | 33.4 | 80.0 | 66.5 | 88.5 | 1.8 | 0.1 | 8.6 | 7.3 | -4.9 | 21.2 | 90.9 | 78.7 | 96.3 | 1.8 | 0.1 | 8.6 |
| | | 1 | 15.9 | 5.1 | 26.9 | 78.2 | 70.3 | 84.2 | 5.9 | 2.7 | 10.7 | 7.8 | -1.9 | 18.2 | 86.3 | 79.1 | 91.2 | 5.9 | 2.7 | 10.7 |
| | | 2 | 14.7 | 3.8 | 25.8 | 76.4 | 69.1 | 82.2 | 8.9 | 5.1 | 14.0 | 5.5 | -4.5 | 16.3 | 84.9 | 78.1 | 89.8 | 9.6 | 5.6 | 14.7 |
| Distant | 65-74 | 0 | 9.7 | 6.1 | 13.5 | 87.7 | 85.0 | 90.0 | 2.5 | 1.5 | 3.9 | 2.7 | -0.3 | 5.8 | 94.5 | 92.4 | 96.0 | 2.9 | 1.8 | 4.3 |
| | | 1 | 14.1 | 10.2 | 17.9 | 83.1 | 80.3 | 85.6 | 2.8 | 1.8 | 4.2 | 5.4 | 2.1 | 8.7 | 91.4 | 89.2 | 93.2 | 3.2 | 2.1 | 4.7 |
| | | 2 | 14.9 | 9.8 | 20.1 | 80.9 | 77.3 | 84.1 | 4.2 | 2.7 | 6.1 | 7.3 | 2.6 | 12.0 | 88.0 | 84.8 | 90.5 | 4.8 | 3.1 | 6.8 |
| | 75-84 | 0 | 6.6 | 0.6 | 12.9 | 89.3 | 85.0 | 92.5 | 4.1 | 2.2 | 7.0 | 3.5 | -2.2 | 9.5 | 92.0 | 88.0 | 94.7 | 4.5 | 2.5 | 7.4 |
| | | 1 | 10.6 | 5.8 | 15.5 | 85.8 | 82.4 | 88.6 | 3.6 | 2.2 | 5.6 | 4.1 | -0.2 | 8.5 | 91.7 | 88.8 | 93.9 | 4.2 | 2.7 | 6.3 |
| | | 2 | 8.7 | 3.9 | 13.6 | 86.6 | 83.3 | 89.3 | 4.7 | 3.1 | 6.8 | 3.2 | -1.2 | 7.8 | 91.4 | 88.5 | 93.6 | 5.4 | 3.6 | 7.6 |
| | 85-94 | 0 | 5.0 | -8.2 | 19.8 | 90.1 | 79.0 | 95.5 | 4.8 | 1.2 | 12.6 | 5.0 | -8.2 | 19.8 | 90.1 | 79.0 | 95.5 | 4.8 | 1.2 | 12.6 |
| | | 1 | 5.8 | -3.1 | 15.4 | 91.4 | 83.9 | 95.5 | 2.9 | 0.7 | 7.6 | 1.9 | -7.3 | 12.2 | 93.3 | 86.1 | 96.8 | 4.8 | 1.7 | 10.5 |
| | | 2 | 5.2 | -4.3 | 15.5 | 89.6 | 82.3 | 93.9 | 5.2 | 2.1 | 10.4 | 0.9 | -7.5 | 10.6 | 93.9 | 87.3 | 97.1 | 5.2 | 2.1 | 10.4 |

Table S12E1. Number of patients and five-year probabilities of dying from cancer, dying from other-causes, and survival by stage, age and comorbidity level: **female liver cancer**

| Stage | Age at diagnosis | Comorbidity | N | Survival (%) | | | Cancer deaths (%) | | | Other-cause deaths (%) | | |
|---|---|---|---|---|---|---|---|---|---|---|---|---|
| | | | | Estimate | 95%CI. | | Estimate | 95%CI. | | Estimate | 95%CI. | |
| Localized | 65-74 | 0 | 764 | 33.0 | 27.9 | 38.0 | 63.6 | 59.8 | 67.2 | 3.3 | 2.2 | 4.9 |
| | | 1 | 4032 | 39.7 | 37.3 | 42.0 | 56.1 | 54.4 | 57.8 | 4.2 | 3.6 | 5.0 |
| | | 2 | 2395 | 24.0 | 20.7 | 27.3 | 67.4 | 65.2 | 69.4 | 8.6 | 7.5 | 9.9 |
| | 75-84 | 0 | 404 | 16.9 | 10.2 | 23.7 | 77.7 | 72.9 | 81.7 | 5.4 | 3.4 | 8.1 |
| | | 1 | 2581 | 24.1 | 20.8 | 27.3 | 67.4 | 65.3 | 69.4 | 8.5 | 7.3 | 9.7 |
| | | 2 | 2060 | 14.8 | 11.3 | 18.5 | 73.4 | 71.3 | 75.5 | 11.7 | 10.3 | 13.3 |
| | 85-94 | 0 | 108 | 5.7 | -8.8 | 21.4 | 79.0 | 69.5 | 85.8 | 15.3 | 9.1 | 23.1 |
| | | 1 | 381 | 6.2 | -1.5 | 14.4 | 81.1 | 76.3 | 85.1 | 12.6 | 9.3 | 16.5 |
| | | 2 | 370 | 3.9 | -4.6 | 12.8 | 77.2 | 72.3 | 81.4 | 18.9 | 14.9 | 23.2 |
| Regional | 65-74 | 0 | 241 | 4.8 | -0.7 | 10.8 | 92.7 | 88.1 | 95.6 | 2.5 | 1.0 | 5.1 |
| | | 1 | 591 | 6.9 | 2.8 | 11.2 | 90.0 | 86.9 | 92.3 | 3.1 | 1.9 | 4.9 |
| | | 2 | 343 | 5.0 | -1.4 | 11.7 | 88.3 | 84.0 | 91.5 | 6.7 | 4.2 | 9.9 |
| | 75-84 | 0 | 153 | 2.8 | -4.6 | 11.4 | 93.1 | 87.0 | 96.4 | 4.1 | 1.6 | 8.3 |
| | | 1 | 532 | 2.3 | -2.2 | 7.1 | 92.1 | 89.1 | 94.2 | 5.6 | 3.8 | 7.9 |
| | | 2 | 385 | 4.2 | -1.5 | 10.2 | 88.9 | 85.1 | 91.7 | 6.9 | 4.7 | 9.8 |
| | 85-94 | 0 | 50 | 4.0 | -12.5 | 23.1 | 88.0 | 74.5 | 94.6 | 8.0 | 2.4 | 17.9 |
| | | 1 | 145 | 0.0 | -5.1 | 8.7 | 97.9 | 90.7 | 99.5 | 2.1 | 0.6 | 5.6 |
| | | 2 | 132 | 1.9 | -9.0 | 14.7 | 89.2 | 80.7 | 94.1 | 8.9 | 4.6 | 14.9 |
| Distant | 65-74 | 0 | 260 | 0.4 | -2.9 | 4.4 | 98.0 | 95.1 | 99.2 | 1.6 | 0.5 | 3.7 |
| | | 1 | 461 | 1.1 | -2.3 | 4.8 | 95.8 | 93.4 | 97.4 | 3.0 | 1.7 | 4.9 |

| | | | | | | | | | | | | |
|---|---|---|---|---|---|---|---|---|---|---|---|---|
| | | 2 | 244 | 0.9 | -5.2 | 7.6 | 93.2 | 89.0 | 95.8 | 5.9 | 3.4 | 9.4 |
| | 75-84 | 0 | 156 | 1.4 | -4.7 | 8.6 | 95.4 | 90.2 | 97.8 | 3.2 | 1.2 | 6.9 |
| | | 1 | 394 | 0.8 | -3.4 | 5.3 | 94.8 | 92.0 | 96.6 | 4.4 | 2.6 | 6.7 |
| | | 2 | 277 | 2.1 | -2.7 | 7.2 | 94.6 | 91.2 | 96.7 | 3.3 | 1.6 | 6.0 |
| | 85-94 | 0 | 50 | 2.0 | -10.6 | 19.1 | 96.0 | 80.8 | 99.2 | 2.0 | 0.1 | 11.3 |
| | | 1 | 90 | 2.2 | -6.3 | 12.3 | 94.4 | 86.8 | 97.7 | 3.3 | 0.9 | 8.6 |
| | | 2 | 93 | 2.2 | -7.7 | 13.6 | 92.4 | 84.4 | 96.4 | 5.4 | 2.0 | 11.4 |

Table S12E2. One and two-year probabilities of dying from cancer, dying from other-causes, and survival by stage, age and comorbidity level: **female liver cancer**

| Stage | Age at diagnosis | Comorbidity | One-year (%) | | | | | | | | | Two-year (%) | | | | | | | | |
|---|---|---|---|---|---|---|---|---|---|---|---|---|---|---|---|---|---|---|---|---|
| | | | Survival | | | Cancer deaths | | | Other-cause deaths | | | Survival | | | Cancer deaths | | | Other-cause deaths | | |
| | | | Est. | 95%CI. | | Est. | 95%CI. | | Est. | 95%CI. | | Est. | 95%CI. | | Est. | 95%CI. | | Est. | 95%CI. | |
| Localized | 65-74 | 0 | 67.4 | 63.1 | 71.4 | 31.1 | 27.9 | 34.4 | 1.4 | 0.8 | 2.5 | 54.7 | 50.0 | 59.1 | 43.4 | 39.9 | 47.0 | 1.8 | 1.1 | 3.0 |
| | | 1 | 81.7 | 80.1 | 83.1 | 17.2 | 16.0 | 18.4 | 1.2 | 0.9 | 1.5 | 69.2 | 67.3 | 71.0 | 28.7 | 27.3 | 30.2 | 2.1 | 1.6 | 2.5 |
| | | 2 | 72.8 | 70.4 | 75.1 | 24.7 | 23.0 | 26.4 | 2.5 | 1.9 | 3.2 | 56.2 | 53.3 | 58.9 | 39.4 | 37.4 | 41.4 | 4.4 | 3.7 | 5.3 |
| | 75-84 | 0 | 51.4 | 44.7 | 57.5 | 46.2 | 41.2 | 50.9 | 2.5 | 1.3 | 4.4 | 37.7 | 30.8 | 44.2 | 58.8 | 53.8 | 63.5 | 3.5 | 2.0 | 5.7 |
| | | 1 | 72.2 | 69.9 | 74.4 | 25.6 | 23.9 | 27.3 | 2.2 | 1.7 | 2.9 | 55.8 | 53.1 | 58.5 | 40.1 | 38.2 | 42.0 | 4.1 | 3.3 | 4.9 |
| | | 2 | 62.0 | 59.0 | 64.9 | 33.4 | 31.4 | 35.4 | 4.6 | 3.7 | 5.5 | 43.7 | 40.4 | 47.0 | 49.5 | 47.3 | 51.7 | 6.8 | 5.7 | 7.9 |
| | 85-94 | 0 | 29.6 | 14.8 | 44.0 | 61.1 | 51.2 | 69.6 | 9.3 | 4.7 | 15.6 | 15.7 | 0.9 | 31.1 | 70.4 | 60.8 | 78.0 | 13.9 | 8.2 | 21.1 |
| | | 1 | 50.9 | 43.4 | 57.9 | 43.6 | 38.5 | 48.5 | 5.5 | 3.5 | 8.1 | 32.4 | 24.6 | 39.9 | 60.2 | 55.0 | 65.0 | 7.5 | 5.1 | 10.4 |
| | | 2 | 45.7 | 37.5 | 53.4 | 45.4 | 40.3 | 50.4 | 8.9 | 6.3 | 12.1 | 25.1 | 16.4 | 33.7 | 60.4 | 55.1 | 65.2 | 14.6 | 11.2 | 18.4 |
| Regional | 65-74 | 0 | 21.8 | 14.2 | 29.2 | 75.7 | 69.8 | 80.7 | 2.5 | 1.0 | 5.1 | 10.2 | 3.9 | 16.6 | 87.3 | 82.3 | 91.0 | 2.5 | 1.0 | 5.1 |
| | | 1 | 30.4 | 25.3 | 35.4 | 67.2 | 63.2 | 70.8 | 2.4 | 1.4 | 3.9 | 17.2 | 12.7 | 21.8 | 80.4 | 76.9 | 83.4 | 2.4 | 1.4 | 3.9 |

| | | | | | | | | | | | | | | | | | | | | |
|---|---|---|---|---|---|---|---|---|---|---|---|---|---|---|---|---|---|---|---|---|
| | | 2 | 27.4 | 20.1 | 34.5 | 67.7 | 62.5 | 72.3 | 5.0 | 3.0 | 7.6 | 15.4 | 8.8 | 22.0 | 79.7 | 75.0 | 83.6 | 5.0 | 3.0 | 7.6 |
| | 75-84 | 0 | 15.3 | 6.6 | 24.2 | 82.0 | 74.9 | 87.3 | 2.6 | 0.9 | 6.1 | 8.6 | 0.6 | 17.1 | 88.1 | 81.7 | 92.4 | 3.3 | 1.2 | 7.1 |
| | | 1 | 20.9 | 15.7 | 26.0 | 75.8 | 71.9 | 79.2 | 3.4 | 2.1 | 5.2 | 10.3 | 5.5 | 15.3 | 84.9 | 81.6 | 87.7 | 4.7 | 3.2 | 6.8 |
| | | 2 | 18.3 | 11.8 | 24.7 | 76.5 | 72.0 | 80.4 | 5.2 | 3.3 | 7.7 | 8.9 | 2.9 | 15.1 | 84.8 | 80.8 | 88.0 | 6.3 | 4.2 | 9.0 |
| | 85-94 | 0 | 8.0 | -8.4 | 26.0 | 86.0 | 72.5 | 93.2 | 6.0 | 1.5 | 15.2 | 4.0 | -12.5 | 23.1 | 88.0 | 74.5 | 94.6 | 8.0 | 2.4 | 17.9 |
| | | 1 | 10.1 | 2.2 | 18.4 | 87.8 | 81.0 | 92.2 | 2.1 | 0.6 | 5.6 | 5.6 | -1.3 | 13.3 | 92.3 | 86.2 | 95.7 | 2.1 | 0.6 | 5.6 |
| | | 2 | 15.2 | 4.2 | 26.3 | 78.8 | 70.8 | 84.8 | 6.1 | 2.8 | 11.0 | 8.0 | -2.6 | 19.2 | 84.3 | 76.9 | 89.6 | 7.7 | 3.9 | 13.0 |
| Distant | 65-74 | 0 | 11.3 | 5.6 | 17.1 | 87.2 | 82.4 | 90.7 | 1.6 | 0.5 | 3.7 | 5.3 | 0.6 | 10.3 | 93.1 | 89.2 | 95.6 | 1.6 | 0.5 | 3.7 |
| | | 1 | 11.5 | 6.9 | 16.2 | 85.9 | 82.4 | 88.8 | 2.6 | 1.4 | 4.4 | 4.1 | 0.3 | 8.1 | 93.1 | 90.3 | 95.1 | 2.8 | 1.6 | 4.6 |
| | | 2 | 13.2 | 5.7 | 20.9 | 81.8 | 76.4 | 86.1 | 4.9 | 2.7 | 8.2 | 6.2 | -0.4 | 13.1 | 88.9 | 84.2 | 92.2 | 4.9 | 2.7 | 8.2 |
| | 75-84 | 0 | 6.4 | -1.0 | 14.3 | 90.4 | 84.5 | 94.1 | 3.2 | 1.2 | 6.9 | 1.4 | -4.7 | 8.6 | 95.4 | 90.2 | 97.8 | 3.2 | 1.2 | 6.9 |
| | | 1 | 11.6 | 6.2 | 17.1 | 84.6 | 80.7 | 87.8 | 3.8 | 2.2 | 6.0 | 3.6 | -1.0 | 8.4 | 92.3 | 89.2 | 94.6 | 4.1 | 2.4 | 6.4 |
| | | 2 | 6.5 | 1.1 | 12.1 | 90.6 | 86.5 | 93.5 | 2.9 | 1.4 | 5.4 | 2.1 | -2.7 | 7.2 | 94.6 | 91.2 | 96.7 | 3.3 | 1.6 | 6.0 |
| | 85-94 | 0 | 6.0 | 1.8 | 19.1 | 94.0 | 80.9 | 98.2 | 0.0 | NA | NA | 2.0 | -10.6 | 19.1 | 96.0 | 80.8 | 99.2 | 2.0 | 0.1 | 11.3 |
| | | 1 | 2.2 | -6.3 | 12.3 | 94.4 | 86.8 | 97.7 | 3.3 | 0.9 | 8.6 | 2.2 | -6.3 | 12.3 | 94.4 | 86.8 | 97.7 | 3.3 | 0.9 | 8.6 |
| | | 2 | 6.5 | -4.6 | 18.5 | 88.0 | 79.4 | 93.2 | 5.4 | 2.0 | 11.4 | 2.2 | -7.7 | 13.6 | 92.4 | 84.4 | 96.4 | 5.4 | 2.0 | 11.4 |

Table S12F1. Number of patients and five-year probabilities of dying from cancer, dying from other-causes, and survival by stage, age and comorbidity level: **male lung cancer**

| Stage | Age at diagnosis | Comorbidity | N | Survival (%) | | | Cancer deaths (%) | | | Other-cause deaths (%) | | |
|---|---|---|---|---|---|---|---|---|---|---|---|---|
| | | | | Estimate | 95%CI. | | Estimate | 95%CI. | | Estimate | 95%CI. | |
| Localized | 65-74 | 0 | 584 | 51.3 | 44.5 | 57.7 | 43.0 | 38.5 | 47.5 | 5.7 | 3.9 | 8.0 |
| | | 1 | 834 | 58.7 | 52.8 | 64.2 | 34.2 | 30.5 | 37.9 | 7.1 | 5.3 | 9.3 |
| | | 2 | 723 | 42.4 | 35.8 | 48.8 | 47.6 | 43.4 | 51.6 | 10.0 | 7.7 | 12.6 |
| | 75-84 | 0 | 385 | 33.8 | 25.2 | 42.1 | 56.7 | 51.2 | 61.8 | 9.6 | 6.7 | 13.0 |
| | | 1 | 577 | 37.9 | 30.4 | 45.3 | 51.2 | 46.5 | 55.7 | 10.9 | 8.2 | 13.9 |
| | | 2 | 1005 | 24.6 | 19.1 | 30.1 | 62.9 | 59.5 | 66.1 | 12.5 | 10.4 | 14.8 |
| | 85-94 | 0 | 58 | 6.3 | -12.0 | 28.1 | 83.0 | 67.6 | 91.5 | 10.7 | 4.3 | 20.5 |
| | | 1 | 103 | 23.6 | 6.9 | 40.8 | 66.8 | 54.6 | 76.5 | 9.5 | 4.6 | 16.6 |
| | | 2 | 242 | 9.5 | -2.2 | 21.7 | 67.7 | 60.9 | 73.5 | 22.8 | 17.4 | 28.7 |
| Regional | 65-74 | 0 | 1322 | 11.5 | 8.3 | 14.8 | 84.0 | 81.8 | 86.0 | 4.5 | 3.5 | 5.7 |
| | | 1 | 1208 | 13.7 | 10.0 | 17.5 | 81.0 | 78.5 | 83.3 | 5.2 | 4.0 | 6.7 |
| | | 2 | 1228 | 11.3 | 7.6 | 15.1 | 81.5 | 79.0 | 83.7 | 7.2 | 5.8 | 8.8 |
| | 75-84 | 0 | 955 | 4.9 | 1.7 | 8.2 | 90.8 | 88.7 | 92.5 | 4.4 | 3.2 | 5.8 |
| | | 1 | 1227 | 7.3 | 3.9 | 10.7 | 86.6 | 84.5 | 88.5 | 6.1 | 4.8 | 7.6 |
| | | 2 | 1862 | 4.7 | 2.0 | 7.5 | 88.1 | 86.5 | 89.6 | 7.2 | 6.0 | 8.4 |
| | 85-94 | 0 | 178 | 0.8 | -7.6 | 10.2 | 90.2 | 84.4 | 93.9 | 9.0 | 5.4 | 13.7 |
| | | 1 | 221 | 2.7 | -5.3 | 11.2 | 88.7 | 83.5 | 92.4 | 8.6 | 5.3 | 13.0 |
| | | 2 | 470 | 1.4 | -4.0 | 7.1 | 89.4 | 86.1 | 91.9 | 9.2 | 6.8 | 12.1 |
| Distant | 65-74 | 0 | 3463 | 1.9 | 0.5 | 3.4 | 94.4 | 93.5 | 95.1 | 3.7 | 3.1 | 4.4 |
| | | 1 | 3302 | 2.8 | 1.2 | 4.4 | 93.5 | 92.5 | 94.3 | 3.7 | 3.1 | 4.4 |

| | | | | | | | | | | | | |
|---|---|---|---|---|---|---|---|---|---|---|---|---|
| | | 2 | 2494 | 2.5 | 0.8 | 4.4 | 93.6 | 92.5 | 94.5 | 3.9 | 3.2 | 4.7 |
| | 75-84 | 0 | 2489 | 1.1 | -0.5 | 2.7 | 95.2 | 94.3 | 96.0 | 3.7 | 3.0 | 4.6 |
| | | 1 | 3342 | 1.6 | 0.0 | 3.1 | 93.9 | 93.0 | 94.7 | 4.6 | 3.9 | 5.3 |
| | | 2 | 4174 | 0.9 | -0.6 | 2.5 | 92.8 | 92.0 | 93.6 | 6.3 | 5.6 | 7.0 |
| | 85-94 | 0 | 566 | 0.4 | -3.4 | 4.5 | 93.8 | 91.5 | 95.6 | 5.7 | 4.0 | 7.9 |
| | | 1 | 865 | 0.5 | -2.8 | 4.0 | 93.0 | 91.0 | 94.5 | 6.6 | 5.0 | 8.3 |
| | | 2 | 1330 | 0.7 | -2.0 | 3.6 | 92.4 | 90.8 | 93.7 | 6.9 | 5.6 | 8.3 |

Table S12F2. One and two-year probabilities of dying from cancer, dying from other-causes, and survival by stage, age and comorbidity level: **male lung cancer**

| Stage | Age at diagnosis | Comorbidity | One-year (%) | | | | | | | | | Two-year (%) | | | | | | | | |
|---|---|---|---|---|---|---|---|---|---|---|---|---|---|---|---|---|---|---|---|---|
| | | | Survival | | | Cancer deaths | | | Other-cause deaths | | | Survival | | | Cancer deaths | | | Other-cause deaths | | |
| | | | Est. | 95%CI. | | Est. | 95%CI. | | Est. | 95%CI. | | Est. | 95%CI. | | Est. | 95%CI. | | Est. | 95%CI. | |
| Localized | 65-74 | 0 | 86.4 | 82.3 | 89.8 | 11.5 | 9.1 | 14.2 | 2.1 | 1.1 | 3.5 | 74.7 | 69.5 | 79.2 | 22.2 | 18.9 | 25.7 | 3.1 | 1.9 | 4.8 |
| | | 1 | 87.9 | 84.7 | 90.5 | 10.6 | 8.6 | 12.8 | 1.6 | 0.9 | 2.6 | 78.0 | 73.9 | 81.6 | 19.2 | 16.5 | 22.0 | 2.8 | 1.8 | 4.1 |
| | | 2 | 83.7 | 79.8 | 87.0 | 14.3 | 11.8 | 16.9 | 2.1 | 1.2 | 3.3 | 70.4 | 65.4 | 74.9 | 25.0 | 21.8 | 28.3 | 4.6 | 3.2 | 6.3 |
| | 75-84 | 0 | 71.8 | 65.1 | 77.6 | 24.0 | 19.9 | 28.4 | 4.2 | 2.5 | 6.5 | 54.0 | 46.4 | 61.0 | 40.5 | 35.4 | 45.4 | 5.6 | 3.6 | 8.2 |
| | | 1 | 77.3 | 72.3 | 81.6 | 19.6 | 16.5 | 22.9 | 3.1 | 1.9 | 4.8 | 67.1 | 61.3 | 72.4 | 27.8 | 24.1 | 31.5 | 5.2 | 3.5 | 7.2 |
| | | 2 | 69.9 | 65.8 | 73.6 | 26.3 | 23.6 | 29.1 | 3.8 | 2.7 | 5.1 | 50.4 | 45.6 | 54.9 | 43.2 | 40.1 | 46.3 | 6.4 | 5.0 | 8.0 |
| | 85-94 | 0 | 51.3 | 31.5 | 67.5 | 45.2 | 31.9 | 57.5 | 3.5 | 0.6 | 11.0 | 23.8 | 2.9 | 44.5 | 65.5 | 51.2 | 76.6 | 10.7 | 4.3 | 20.5 |
| | | 1 | 65.0 | 50.6 | 76.7 | 30.1 | 21.5 | 39.1 | 4.9 | 1.8 | 10.3 | 47.4 | 32.2 | 61.0 | 46.6 | 36.6 | 56.1 | 5.9 | 2.4 | 11.7 |
| | | 2 | 56.5 | 46.6 | 65.4 | 36.1 | 30.1 | 42.1 | 7.5 | 4.6 | 11.2 | 34.1 | 23.1 | 44.8 | 51.4 | 44.8 | 57.5 | 14.6 | 10.4 | 19.4 |
| Regional | 65-74 | 0 | 52.9 | 49.3 | 56.3 | 44.6 | 41.9 | 47.3 | 2.5 | 1.8 | 3.5 | 30.4 | 26.8 | 33.9 | 66.2 | 63.5 | 68.7 | 3.4 | 2.5 | 4.5 |
| | | 1 | 57.2 | 53.4 | 60.8 | 40.1 | 37.3 | 42.8 | 2.7 | 1.9 | 3.8 | 35.8 | 32.0 | 39.5 | 61.0 | 58.2 | 63.7 | 3.2 | 2.3 | 4.3 |

| | | | | | | | | | | | | | | | | | | | | |
|---|---|---|---|---|---|---|---|---|---|---|---|---|---|---|---|---|---|---|---|---|
| | | 2 | 48.3 | 44.4 | 52.1 | 47.9 | 45.0 | 50.6 | 3.8 | 2.9 | 5.0 | 28.8 | 24.7 | 32.8 | 65.3 | 62.5 | 67.9 | 5.9 | 4.7 | 7.4 |
| | 75-84 | 0 | 35.8 | 31.6 | 39.8 | 61.7 | 58.5 | 64.7 | 2.5 | 1.7 | 3.7 | 16.6 | 12.9 | 20.4 | 79.9 | 77.2 | 82.3 | 3.5 | 2.5 | 4.8 |
| | | 1 | 43.9 | 40.0 | 47.5 | 53.2 | 50.4 | 55.9 | 2.9 | 2.1 | 4.0 | 22.3 | 18.7 | 25.9 | 73.5 | 70.9 | 75.9 | 4.2 | 3.2 | 5.4 |
| | | 2 | 35.5 | 32.3 | 38.6 | 60.1 | 57.8 | 62.3 | 4.5 | 3.6 | 5.5 | 17.0 | 14.1 | 20.0 | 77.5 | 75.5 | 79.4 | 5.5 | 4.5 | 6.6 |
| | 85-94 | 0 | 18.0 | 7.4 | 28.7 | 73.0 | 65.9 | 78.9 | 9.0 | 5.4 | 13.7 | 3.4 | -5.4 | 12.8 | 87.6 | 81.8 | 91.7 | 9.0 | 5.4 | 13.7 |
| | | 1 | 31.3 | 21.9 | 40.3 | 64.1 | 57.4 | 70.1 | 4.6 | 2.4 | 8.0 | 10.7 | 2.3 | 19.3 | 82.8 | 76.9 | 87.3 | 6.5 | 3.7 | 10.3 |
| | | 2 | 28.6 | 22.1 | 35.0 | 65.4 | 60.9 | 69.5 | 6.0 | 4.1 | 8.4 | 8.4 | 2.6 | 14.5 | 83.1 | 79.4 | 86.2 | 8.5 | 6.2 | 11.2 |
| Distant | 65-74 | 0 | 30.6 | 28.5 | 32.7 | 66.7 | 65.1 | 68.2 | 2.7 | 2.2 | 3.3 | 12.9 | 11.1 | 14.8 | 83.9 | 82.6 | 85.1 | 3.2 | 2.6 | 3.8 |
| | | 1 | 33.9 | 31.7 | 36.1 | 63.4 | 61.7 | 65.0 | 2.7 | 2.2 | 3.3 | 15.3 | 13.3 | 17.2 | 81.5 | 80.2 | 82.8 | 3.2 | 2.6 | 3.8 |
| | | 2 | 27.2 | 24.7 | 29.7 | 69.9 | 68.1 | 71.6 | 2.9 | 2.3 | 3.6 | 12.0 | 9.8 | 14.1 | 84.7 | 83.2 | 86.0 | 3.4 | 2.7 | 4.2 |
| | 75-84 | 0 | 21.2 | 18.9 | 23.6 | 75.9 | 74.2 | 77.5 | 2.9 | 2.3 | 3.6 | 7.6 | 5.7 | 9.6 | 89.0 | 87.7 | 90.2 | 3.4 | 2.7 | 4.1 |
| | | 1 | 23.8 | 21.7 | 25.9 | 73.2 | 71.7 | 74.7 | 2.9 | 2.4 | 3.6 | 8.4 | 6.6 | 10.2 | 87.6 | 86.4 | 88.6 | 4.0 | 3.4 | 4.7 |
| | | 2 | 19.3 | 17.3 | 21.2 | 76.0 | 74.7 | 77.3 | 4.7 | 4.1 | 5.4 | 7.3 | 5.6 | 9.0 | 87.0 | 86.0 | 88.0 | 5.7 | 5.0 | 6.4 |
| | 85-94 | 0 | 12.7 | 7.8 | 17.7 | 82.2 | 78.8 | 85.1 | 5.1 | 3.5 | 7.2 | 4.0 | -0.2 | 8.4 | 90.6 | 87.9 | 92.8 | 5.3 | 3.7 | 7.4 |
| | | 1 | 15.2 | 11.1 | 19.3 | 79.7 | 76.9 | 82.2 | 5.1 | 3.8 | 6.7 | 5.0 | 1.3 | 8.8 | 88.7 | 86.4 | 90.6 | 6.3 | 4.8 | 8.0 |
| | | 2 | 14.0 | 10.7 | 17.4 | 80.1 | 77.9 | 82.1 | 5.9 | 4.7 | 7.2 | 5.0 | 2.0 | 8.1 | 88.5 | 86.7 | 90.1 | 6.5 | 5.3 | 7.9 |

Table S12G1. Number of patients and five-year probabilities of dying from cancer, dying from other-causes, and survival by stage, age and comorbidity level: **female lung cancer**

| Stage | Age at diagnosis | Comorbidity | N | Survival (%) | | | Cancer deaths (%) | | | Other-cause deaths (%) | | |
|---|---|---|---|---|---|---|---|---|---|---|---|---|
| | | | | Estimate | 95%CI. | | Estimate | 95%CI. | | Estimate | 95%CI. | |
| Localized | 65-74 | 0 | 468 | 76.4 | 69.8 | 81.9 | 21.1 | 16.9 | 25.6 | 2.5 | 1.2 | 4.5 |
| | | 1 | 788 | 76.5 | 71.2 | 81.1 | 20.3 | 16.9 | 23.9 | 3.2 | 2.0 | 4.9 |
| | | 2 | 430 | 69.5 | 61.7 | 76.4 | 24.7 | 20.1 | 29.6 | 5.7 | 3.5 | 8.7 |
| | 75-84 | 0 | 156 | 56.7 | 41.7 | 69.3 | 36.4 | 27.7 | 45.1 | 6.9 | 2.9 | 13.2 |
| | | 1 | 440 | 59.8 | 51.4 | 67.3 | 35.0 | 29.7 | 40.3 | 5.2 | 3.0 | 8.3 |
| | | 2 | 349 | 43.7 | 33.7 | 53.3 | 44.0 | 38.0 | 49.8 | 12.2 | 8.7 | 16.4 |
| | 85-94 | 0 | 19 | 20.4 | -19.8 | 62.7 | 63.0 | 33.5 | 82.2 | 16.6 | 3.7 | 37.5 |
| | | 1 | 64 | 25.7 | 4.5 | 47.7 | 66.0 | 49.3 | 78.4 | 8.3 | 3.0 | 17.1 |
| | | 2 | 70 | 5.8 | -24.2 | 40.5 | 68.7 | 47.6 | 82.7 | 25.4 | 11.9 | 41.5 |
| Regional | 65-74 | 0 | 454 | 30.1 | 23.6 | 36.4 | 67.4 | 62.3 | 71.9 | 2.6 | 1.4 | 4.5 |
| | | 1 | 638 | 23.9 | 17.9 | 29.8 | 70.7 | 66.5 | 74.5 | 5.4 | 3.7 | 7.6 |
| | | 2 | 363 | 18.7 | 10.9 | 26.7 | 74.5 | 68.9 | 79.3 | 6.8 | 4.4 | 9.8 |
| | 75-84 | 0 | 212 | 13.7 | 5.3 | 22.4 | 81.9 | 75.5 | 86.8 | 4.4 | 2.2 | 7.9 |
| | | 1 | 520 | 14.3 | 8.2 | 20.5 | 78.8 | 74.7 | 82.4 | 6.8 | 4.8 | 9.4 |
| | | 2 | 418 | 11.5 | 4.6 | 18.6 | 80.7 | 76.0 | 84.5 | 7.8 | 5.4 | 10.9 |
| | 85-94 | 0 | 72 | 1.4 | -12.2 | 16.9 | 90.1 | 79.8 | 95.3 | 8.5 | 3.2 | 16.9 |
| | | 1 | 125 | 2.7 | -7.0 | 13.5 | 90.0 | 83.0 | 94.3 | 7.3 | 3.5 | 12.8 |
| | | 2 | 157 | 3.0 | -6.5 | 13.5 | 88.4 | 81.7 | 92.8 | 8.5 | 4.8 | 13.7 |
| Distant | 65-74 | 0 | 1839 | 6.4 | 4.1 | 8.8 | 90.6 | 89.0 | 92.0 | 3.0 | 2.3 | 3.9 |
| | | 1 | 2426 | 6.9 | 4.8 | 9.0 | 89.9 | 88.4 | 91.1 | 3.2 | 2.6 | 4.0 |

| | | | | | | | | | | | | |
|---|---|---|---|---|---|---|---|---|---|---|---|---|
| | | 2 | 1127 | 5.4 | 2.1 | 8.8 | 89.1 | 86.9 | 90.9 | 5.5 | 4.3 | 7.0 |
| | 75-84 | 0 | 1042 | 3.0 | 0.1 | 6.1 | 92.5 | 90.6 | 94.1 | 4.5 | 3.3 | 5.9 |
| | | 1 | 2261 | 3.5 | 1.2 | 5.8 | 90.7 | 89.3 | 91.9 | 5.8 | 4.9 | 6.9 |
| | | 2 | 1582 | 2.5 | -0.2 | 5.3 | 90.9 | 89.3 | 92.3 | 6.6 | 5.4 | 7.9 |
| | 85-94 | 0 | 302 | 1.7 | -4.3 | 8.3 | 91.3 | 87.3 | 94.1 | 7.0 | 4.5 | 10.2 |
| | | 1 | 680 | 1.5 | -2.4 | 5.7 | 92.0 | 89.6 | 93.9 | 6.4 | 4.7 | 8.5 |
| | | 2 | 592 | 0.3 | -4.3 | 5.1 | 91.5 | 88.8 | 93.5 | 8.2 | 6.1 | 10.7 |

Table S12G2. One and Two -year probabilities of dying from cancer, dying from other-causes, and survival by stage, age and comorbidity level: **female lung cancer**

| Stage | Age at diagnosis | Comorbidity | One-year (%) | | | | | | | | | Two-year (%) | | | | | | | | |
|---|---|---|---|---|---|---|---|---|---|---|---|---|---|---|---|---|---|---|---|---|
| | | | Survival | | | Cancer deaths | | | Other-cause deaths | | | Survival | | | Cancer deaths | | | Other-cause deaths | | |
| | | | Est. | 95%CI. | | Est. | 95%CI. | | Est. | 95%CI. | | Est. | 95%CI. | | Est. | 95%CI. | | Est. | 95%CI. | |
| Localized | 65-74 | 0 | 97.4 | 94.7 | 98.9 | 1.7 | 0.8 | 3.2 | 0.9 | 0.3 | 2.1 | 92.3 | 88.5 | 95.0 | 6.8 | 4.7 | 9.4 | 0.9 | 0.3 | 2.1 |
| | | 1 | 97.1 | 95.1 | 98.3 | 2.3 | 1.4 | 3.5 | 0.6 | 0.2 | 1.4 | 93.9 | 91.2 | 95.9 | 4.9 | 3.5 | 6.6 | 1.2 | 0.6 | 2.2 |
| | | 2 | 92.8 | 88.9 | 95.4 | 6.3 | 4.2 | 8.8 | 0.9 | 0.3 | 2.2 | 86.7 | 81.6 | 90.7 | 10.9 | 8.1 | 14.1 | 2.4 | 1.2 | 4.3 |
| | 75-84 | 0 | 90.4 | 85.1 | 94.4 | 9.6 | 5.6 | 14.9 | 0.0 | NA | NA | 79.7 | 69.8 | 86.8 | 18.8 | 13.0 | 25.6 | 1.4 | 0.3 | 4.7 |
| | | 1 | 93.0 | 89.3 | 95.4 | 6.6 | 4.5 | 9.2 | 0.5 | 0.1 | 1.5 | 83.0 | 77.8 | 87.2 | 15.3 | 12.1 | 18.9 | 1.7 | 0.7 | 3.3 |
| | | 2 | 83.7 | 77.4 | 88.7 | 11.5 | 8.4 | 15.1 | 4.9 | 2.9 | 7.5 | 69.9 | 62.4 | 76.5 | 24.3 | 19.8 | 29.1 | 5.8 | 3.6 | 8.6 |
| | 85-94 | 0 | 62.3 | 23.8 | 87.0 | 32.4 | 12.7 | 54.0 | 5.3 | 0.3 | 22.1 | 45.3 | 4.6 | 78.0 | 43.7 | 20.3 | 65.1 | 10.9 | 1.6 | 30.4 |
| | | 1 | 70.3 | 52.0 | 83.6 | 25.0 | 15.1 | 36.1 | 4.7 | 1.2 | 11.9 | 54.9 | 34.8 | 71.5 | 38.6 | 26.4 | 50.6 | 6.5 | 2.0 | 14.5 |
| | | 2 | 69.8 | 51.5 | 83.8 | 21.5 | 12.7 | 31.8 | 8.6 | 3.5 | 16.7 | 49.3 | 28.4 | 67.8 | 37.2 | 25.6 | 48.8 | 13.4 | 6.5 | 22.8 |
| Regional | 65-74 | 0 | 74.1 | 68.7 | 78.7 | 24.8 | 20.9 | 28.9 | 1.1 | 0.4 | 2.4 | 58.8 | 52.6 | 64.4 | 39.2 | 34.6 | 43.7 | 2.0 | 1.0 | 3.7 |
| | | 1 | 75.5 | 70.9 | 79.5 | 22.6 | 19.4 | 25.9 | 1.9 | 1.0 | 3.2 | 59.4 | 54.2 | 64.3 | 38.0 | 34.2 | 41.9 | 2.5 | 1.5 | 4.0 |

| | | | | | | | | | | | | | | | | | | | | |
|---|---|---|---|---|---|---|---|---|---|---|---|---|---|---|---|---|---|---|---|---|
| | | 2 | 63.3 | 56.0 | 69.9 | 32.2 | 27.5 | 37.1 | 4.4 | 2.6 | 6.9 | 49.7 | 41.9 | 56.9 | 45.1 | 39.8 | 50.2 | 5.3 | 3.3 | 7.9 |
| | 75-84 | 0 | 53.8 | 44.6 | 61.8 | 44.3 | 37.6 | 50.9 | 1.9 | 0.6 | 4.5 | 33.2 | 23.7 | 42.2 | 63.0 | 56.0 | 69.2 | 3.8 | 1.8 | 7.1 |
| | | 1 | 60.1 | 54.3 | 65.5 | 36.8 | 32.6 | 40.9 | 3.1 | 1.8 | 4.8 | 38.8 | 32.7 | 44.7 | 57.1 | 52.7 | 61.3 | 4.1 | 2.6 | 6.0 |
| | | 2 | 52.6 | 45.8 | 58.9 | 43.8 | 39.0 | 48.5 | 3.6 | 2.1 | 5.7 | 30.9 | 23.9 | 37.7 | 63.3 | 58.4 | 67.8 | 5.8 | 3.8 | 8.3 |
| | 85-94 | 0 | 24.0 | 7.0 | 40.7 | 69.0 | 56.7 | 78.4 | 7.0 | 2.6 | 14.6 | 11.3 | -3.6 | 27.1 | 81.7 | 70.3 | 89.0 | 7.0 | 2.6 | 14.6 |
| | | 1 | 35.6 | 23.0 | 47.4 | 60.4 | 51.1 | 68.4 | 4.0 | 1.5 | 8.6 | 15.4 | 4.1 | 26.9 | 79.0 | 70.6 | 85.2 | 5.6 | 2.5 | 10.7 |
| | | 2 | 38.9 | 27.4 | 49.6 | 56.6 | 48.4 | 64.0 | 4.5 | 2.0 | 8.6 | 12.2 | 1.9 | 22.9 | 80.6 | 73.3 | 86.0 | 7.2 | 3.8 | 12.0 |
| Distant | 65-74 | 0 | 54.8 | 51.9 | 57.5 | 43.7 | 41.4 | 45.9 | 1.6 | 1.1 | 2.2 | 29.7 | 26.8 | 32.5 | 67.9 | 65.7 | 70.0 | 2.4 | 1.8 | 3.2 |
| | | 1 | 54.9 | 52.4 | 57.3 | 43.8 | 41.8 | 45.8 | 1.3 | 0.9 | 1.8 | 31.2 | 28.7 | 33.7 | 66.4 | 64.5 | 68.3 | 2.3 | 1.8 | 3.0 |
| | | 2 | 46.6 | 42.6 | 50.5 | 50.0 | 47.0 | 52.9 | 3.4 | 2.4 | 4.5 | 24.1 | 20.3 | 28.0 | 71.4 | 68.7 | 74.0 | 4.5 | 3.4 | 5.8 |
| | 75-84 | 0 | 38.3 | 34.3 | 42.3 | 59.0 | 55.9 | 61.9 | 2.7 | 1.8 | 3.8 | 18.4 | 14.7 | 22.1 | 78.1 | 75.4 | 80.5 | 3.5 | 2.5 | 4.8 |
| | | 1 | 42.7 | 39.9 | 45.4 | 54.2 | 52.1 | 56.2 | 3.1 | 2.5 | 3.9 | 20.8 | 18.1 | 23.4 | 74.6 | 72.7 | 76.4 | 4.6 | 3.8 | 5.6 |
| | | 2 | 32.9 | 29.4 | 36.3 | 62.4 | 60.0 | 64.7 | 4.8 | 3.8 | 5.9 | 16.3 | 13.1 | 19.5 | 78.0 | 75.9 | 80.0 | 5.7 | 4.6 | 6.9 |
| | 85-94 | 0 | 19.9 | 12.4 | 27.4 | 74.5 | 69.2 | 79.0 | 5.6 | 3.4 | 8.6 | 7.2 | 0.6 | 14.0 | 86.2 | 81.8 | 89.6 | 6.6 | 4.2 | 9.8 |
| | | 1 | 26.6 | 21.5 | 31.5 | 69.0 | 65.4 | 72.3 | 4.4 | 3.1 | 6.2 | 10.8 | 6.3 | 15.3 | 83.7 | 80.7 | 86.3 | 5.5 | 3.9 | 7.4 |
| | | 2 | 20.7 | 15.4 | 25.9 | 74.2 | 70.5 | 77.6 | 5.1 | 3.5 | 7.0 | 8.2 | 3.4 | 13.1 | 85.4 | 82.2 | 88.0 | 6.5 | 4.7 | 8.6 |

Table S12H1. Number of patients and five-year probabilities of dying from cancer, dying from other-causes, and survival by stage, age and comorbidity level: **male oral cancer**

| Stage | Age at diagnosis | Comorbidity | N | Survival (%) | | | Cancer deaths (%) | | | Other-cause deaths (%) | | |
|---|---|---|---|---|---|---|---|---|---|---|---|---|
| | | | | Estimate | 95%CI. | | Estimate | 95%CI. | | Estimate | 95%CI. | |
| Localized | 65-74 | 0 | 839 | 72.7 | 67.4 | 77.4 | 20.4 | 17.5 | 23.5 | 6.9 | 5.1 | 9.0 |
| | | 1 | 996 | 66.7 | 61.3 | 71.7 | 21.8 | 19.0 | 24.7 | 11.5 | 9.3 | 14.0 |
| | | 2 | 619 | 58.6 | 51.2 | 65.5 | 21.8 | 18.2 | 25.5 | 19.7 | 16.2 | 23.3 |
| | 75-84 | 0 | 180 | 56.7 | 43.5 | 68.5 | 28.2 | 21.5 | 35.3 | 15.1 | 10.0 | 21.2 |
| | | 1 | 339 | 54.3 | 44.1 | 63.7 | 28.8 | 23.6 | 34.2 | 16.9 | 12.6 | 21.8 |
| | | 2 | 327 | 38.8 | 27.6 | 49.7 | 34.2 | 28.6 | 39.9 | 27.0 | 21.7 | 32.6 |
| | 85-94 | 0 | 36 | 38.5 | 6.0 | 68.1 | 39.3 | 22.4 | 55.8 | 22.2 | 9.5 | 38.2 |
| | | 1 | 43 | 46.2 | 16.2 | 72.4 | 26.9 | 14.2 | 41.4 | 27.0 | 13.5 | 42.5 |
| | | 2 | 72 | 16.3 | -11.0 | 45.0 | 35.3 | 22.2 | 48.6 | 48.4 | 32.8 | 62.4 |
| Regional | 65-74 | 0 | 1461 | 33.4 | 29.1 | 37.6 | 58.8 | 56.0 | 61.5 | 7.8 | 6.4 | 9.4 |
| | | 1 | 1298 | 37.8 | 33.0 | 42.6 | 51.9 | 48.9 | 54.8 | 10.3 | 8.6 | 12.3 |
| | | 2 | 875 | 25.3 | 19.3 | 31.2 | 60.7 | 57.1 | 64.1 | 14.0 | 11.7 | 16.6 |
| | 75-84 | 0 | 412 | 20.6 | 12.5 | 28.8 | 67.7 | 62.6 | 72.3 | 11.6 | 8.6 | 15.2 |
| | | 1 | 467 | 20.7 | 13.0 | 28.5 | 67.8 | 63.0 | 72.1 | 11.4 | 8.5 | 14.8 |
| | | 2 | 611 | 13.5 | 6.4 | 20.9 | 65.8 | 61.8 | 69.6 | 20.6 | 17.4 | 24.1 |
| | 85-94 | 0 | 77 | 8.6 | -9.0 | 27.4 | 75.5 | 63.9 | 83.8 | 15.9 | 8.7 | 25.1 |
| | | 1 | 93 | 6.8 | -12.1 | 27.2 | 74.3 | 62.3 | 82.9 | 18.9 | 10.5 | 29.2 |
| | | 2 | 146 | 5.6 | -8.3 | 20.3 | 75.0 | 66.6 | 81.5 | 19.4 | 13.1 | 26.8 |
| Distant | 65-74 | 0 | 107 | 8.8 | -1.7 | 20.2 | 86.5 | 78.1 | 91.9 | 4.7 | 1.7 | 9.9 |
| | | 1 | 63 | 9.5 | -8.3 | 29.0 | 80.4 | 67.0 | 88.8 | 10.1 | 4.0 | 19.5 |

| | | | | | | | | | | | | |
|---|---|---|---|---|---|---|---|---|---|---|---|---|
| | | 2 | 62 | 0.0 | -16.5 | 21.3 | 89.6 | 74.8 | 96.0 | 10.4 | 3.9 | 20.5 |
| | 75-84 | 0 | 35 | 2.9 | -17.1 | 27.7 | 88.6 | 70.2 | 95.9 | 8.6 | 2.1 | 21.2 |
| | | 1 | 48 | 4.8 | -12.5 | 24.6 | 88.7 | 74.0 | 95.3 | 6.5 | 1.5 | 17.1 |
| | | 2 | 42 | 6.0 | -12.5 | 28.2 | 86.9 | 70.1 | 94.6 | 7.1 | 1.7 | 17.9 |
| | 85-94 | 0 | 7 | 14.3 | 1.5 | 80.1 | 85.7 | 19.9 | 98.5 | 0.0 | NA | NA |
| | | 1 | 8 | 25.0 | 5.5 | 76.7 | 75.0 | 23.3 | 94.5 | 0.0 | NA | NA |
| | | 2 | 16 | 18.8 | -20.1 | 59.5 | 68.8 | 38.7 | 86.3 | 12.5 | 1.8 | 33.9 |

Table S12H2. One and two-year probabilities of dying from cancer, dying from other-causes, and survival by stage, age and comorbidity level: **male oral cancer**

| Stage | Age at diagnosis | Comorbidity | One-year (%) | | | | | | | | | Two-year (%) | | | | | | | | |
|---|---|---|---|---|---|---|---|---|---|---|---|---|---|---|---|---|---|---|---|---|
| | | | Survival | | | Cancer deaths | | | Other-cause deaths | | | Survival | | | Cancer deaths | | | Other-cause deaths | | |
| | | | Est. | 95%CI. | | Est. | 95%CI. | | Est. | 95%CI. | | Est. | 95%CI. | | Est. | 95%CI. | | Est. | 95%CI. | |
| Localized | 65-74 | 0 | 95.0 | 92.6 | 96.7 | 3.9 | 2.8 | 5.4 | 1.1 | 0.5 | 2.0 | 88.9 | 85.6 | 91.6 | 8.9 | 7.1 | 11.0 | 2.2 | 1.4 | 3.4 |
| | | 1 | 94.2 | 91.9 | 95.9 | 4.6 | 3.4 | 6.1 | 1.2 | 0.7 | 2.0 | 85.3 | 81.9 | 88.2 | 11.1 | 9.3 | 13.2 | 3.6 | 2.5 | 4.9 |
| | | 2 | 91.0 | 87.3 | 93.8 | 5.3 | 3.8 | 7.3 | 3.7 | 2.4 | 5.4 | 82.7 | 77.9 | 86.9 | 9.9 | 7.7 | 12.5 | 7.4 | 5.5 | 9.7 |
| | 75-84 | 0 | 87.8 | 79.7 | 93.2 | 9.4 | 5.7 | 14.3 | 2.8 | 1.0 | 6.0 | 76.9 | 67.0 | 84.8 | 18.0 | 12.7 | 24.0 | 5.1 | 2.5 | 9.0 |
| | | 1 | 85.0 | 78.9 | 89.7 | 11.2 | 8.1 | 14.8 | 3.8 | 2.1 | 6.3 | 74.5 | 66.9 | 81.1 | 17.9 | 14.0 | 22.3 | 7.5 | 5.0 | 10.8 |
| | | 2 | 79.5 | 72.5 | 85.4 | 13.8 | 10.3 | 17.7 | 6.7 | 4.4 | 9.8 | 68.6 | 60.3 | 76.1 | 19.9 | 15.7 | 24.5 | 11.4 | 8.2 | 15.2 |
| | 85-94 | 0 | 77.6 | 52.5 | 92.3 | 16.8 | 6.7 | 30.8 | 5.7 | 1.0 | 16.8 | 62.5 | 34.5 | 83.0 | 28.8 | 14.8 | 44.5 | 8.6 | 2.2 | 21.0 |
| | | 1 | 81.4 | 60.4 | 92.8 | 16.3 | 7.1 | 28.8 | 2.3 | 0.2 | 10.7 | 64.5 | 39.0 | 83.7 | 23.6 | 12.1 | 37.2 | 11.9 | 4.3 | 23.8 |
| | | 2 | 76.4 | 59.3 | 88.5 | 15.3 | 8.1 | 24.6 | 8.3 | 3.4 | 16.2 | 56.3 | 35.2 | 74.2 | 24.5 | 15.0 | 35.2 | 19.3 | 10.8 | 29.6 |
| Regional | 65-74 | 0 | 67.1 | 63.7 | 70.2 | 29.8 | 27.4 | 32.1 | 3.2 | 2.3 | 4.1 | 50.8 | 47.0 | 54.4 | 44.4 | 41.8 | 47.0 | 4.8 | 3.8 | 6.0 |
| | | 1 | 72.1 | 68.7 | 75.2 | 24.9 | 22.6 | 27.3 | 3.0 | 2.2 | 4.0 | 55.9 | 51.9 | 59.8 | 38.6 | 36.0 | 41.3 | 5.4 | 4.3 | 6.8 |

| | | | | | | | | | | | | | | | | | | | | |
|---|---|---|---|---|---|---|---|---|---|---|---|---|---|---|---|---|---|---|---|---|
| | | 2 | 61.8 | 57.0 | 66.3 | 32.8 | 29.7 | 35.9 | 5.4 | 4.0 | 7.0 | 44.3 | 39.0 | 49.4 | 47.7 | 44.3 | 51.0 | 8.0 | 6.4 | 10.0 |
| | 75-84 | 0 | 56.4 | 49.3 | 62.9 | 38.7 | 34.0 | 43.4 | 4.9 | 3.1 | 7.3 | 38.6 | 31.0 | 46.0 | 53.6 | 48.6 | 58.4 | 7.7 | 5.4 | 10.6 |
| | | 1 | 57.6 | 51.2 | 63.5 | 38.5 | 34.1 | 42.9 | 3.9 | 2.4 | 5.9 | 37.0 | 30.1 | 43.7 | 56.6 | 51.9 | 61.0 | 6.3 | 4.4 | 8.8 |
| | | 2 | 49.1 | 42.7 | 55.2 | 41.0 | 37.1 | 44.9 | 9.9 | 7.7 | 12.4 | 30.0 | 23.2 | 36.7 | 55.8 | 51.8 | 59.7 | 14.2 | 11.6 | 17.1 |
| | 85-94 | 0 | 35.8 | 17.7 | 52.9 | 55.0 | 43.1 | 65.4 | 9.2 | 4.0 | 17.0 | 21.2 | 3.1 | 39.6 | 65.6 | 53.7 | 75.1 | 13.2 | 6.7 | 21.8 |
| | | 1 | 48.4 | 32.7 | 62.2 | 46.2 | 35.8 | 56.0 | 5.4 | 2.0 | 11.3 | 25.6 | 9.3 | 41.6 | 64.4 | 53.5 | 73.4 | 10.0 | 4.9 | 17.3 |
| | | 2 | 35.6 | 22.2 | 48.4 | 53.5 | 45.1 | 61.2 | 11.0 | 6.6 | 16.6 | 19.0 | 5.8 | 32.4 | 67.1 | 58.8 | 74.2 | 13.8 | 8.8 | 20.0 |
| Distant | 65-74 | 0 | 39.3 | 25.6 | 51.7 | 57.0 | 47.0 | 65.8 | 3.7 | 1.2 | 8.6 | 17.8 | 5.7 | 29.9 | 77.6 | 68.3 | 84.4 | 4.7 | 1.7 | 9.9 |
| | | 1 | 46.0 | 27.0 | 62.5 | 49.2 | 36.3 | 60.9 | 4.8 | 1.2 | 12.1 | 24.8 | 5.6 | 43.8 | 67.0 | 53.2 | 77.6 | 8.2 | 2.9 | 16.8 |
| | | 2 | 32.3 | 13.3 | 50.1 | 61.3 | 47.9 | 72.2 | 6.5 | 2.0 | 14.5 | 12.9 | -4.0 | 30.7 | 79.0 | 66.4 | 87.4 | 8.1 | 2.9 | 16.6 |
| | 75-84 | 0 | 22.9 | -1.0 | 46.2 | 71.4 | 52.8 | 83.7 | 5.7 | 1.0 | 17.2 | 2.9 | -17.1 | 27.7 | 88.6 | 70.2 | 95.9 | 8.6 | 2.1 | 21.2 |
| | | 1 | 39.6 | 18.2 | 58.3 | 56.3 | 40.9 | 69.0 | 4.2 | 0.7 | 12.7 | 16.7 | -1.1 | 35.0 | 79.2 | 64.3 | 88.4 | 4.2 | 0.7 | 12.7 |
| | | 2 | 14.3 | -4.7 | 34.2 | 81.0 | 65.0 | 90.2 | 4.8 | 0.8 | 14.5 | 11.9 | -8.1 | 33.3 | 81.0 | 65.0 | 90.2 | 7.1 | 1.7 | 17.9 |
| | 85-94 | 0 | 14.3 | 1.5 | 80.1 | 85.7 | 19.9 | 98.5 | 0.0 | NA | NA | 14.3 | 1.5 | 80.1 | 85.7 | 19.9 | 98.5 | 0.0 | NA | NA |
| | | 1 | 25.0 | 5.5 | 76.7 | 75.0 | 23.3 | 94.5 | 0.0 | NA | NA | 25.0 | 5.5 | 76.7 | 75.0 | 23.3 | 94.5 | 0.0 | NA | NA |
| | | 2 | 18.8 | -20.1 | 59.5 | 68.8 | 38.7 | 86.3 | 12.5 | 1.8 | 33.9 | 18.8 | -20.1 | 59.5 | 68.8 | 38.7 | 86.3 | 12.5 | 1.8 | 33.9 |

Table S12I1. Number pf patients and five-year probabilities of dying from cancer, dying from other-causes, and survival by stage, age and comorbidity level: **female oral cancer**

| Stage | Age at diagnosis | Comorbidity | N | Survival (%) | | | Cancer deaths (%) | | | Other-cause deaths (%) | | |
|---|---|---|---|---|---|---|---|---|---|---|---|---|
| | | | | Estimate | 95%CI. | | Estimate | 95%CI. | | Estimate | 95%CI. | |
| Localized | 65-74 | 0 | 135 | 78.1 | 65.8 | 86.9 | 18.6 | 12.0 | 26.3 | 3.4 | 1.1 | 7.9 |
| | | 1 | 256 | 72.0 | 61.5 | 80.7 | 18.9 | 13.8 | 24.5 | 9.2 | 5.5 | 14.0 |
| | | 2 | 125 | 69.3 | 54.2 | 81.5 | 19.8 | 12.8 | 27.8 | 10.9 | 5.7 | 18.0 |
| | 75-84 | 0 | 53 | 70.1 | 46.8 | 86.6 | 21.4 | 10.8 | 34.3 | 8.5 | 2.6 | 18.9 |
| | | 1 | 159 | 63.0 | 48.3 | 75.6 | 22.5 | 15.8 | 30.0 | 14.5 | 8.7 | 21.7 |
| | | 2 | 118 | 53.8 | 36.7 | 69.1 | 24.6 | 16.7 | 33.4 | 21.6 | 14.2 | 29.9 |
| | 85-94 | 0 | 9 | 62.2 | -2.7 | 96.9 | 26.7 | 2.6 | 61.8 | 11.1 | 0.5 | 40.9 |
| | | 1 | 32 | 57.7 | 23.5 | 83.7 | 24.4 | 10.1 | 42.1 | 17.9 | 6.2 | 34.4 |
| | | 2 | 32 | 35.5 | -2.6 | 70.0 | 37.1 | 19.7 | 54.5 | 27.5 | 10.3 | 48.1 |
| Regional | 65-74 | 0 | 163 | 48.5 | 34.4 | 61.1 | 43.0 | 34.4 | 51.2 | 8.6 | 4.4 | 14.4 |
| | | 1 | 264 | 46.0 | 34.9 | 56.5 | 44.7 | 37.8 | 51.4 | 9.2 | 5.7 | 13.7 |
| | | 2 | 141 | 34.6 | 18.6 | 50.0 | 47.7 | 38.7 | 56.2 | 17.7 | 11.4 | 25.2 |
| | 75-84 | 0 | 95 | 29.3 | 11.3 | 46.6 | 59.5 | 48.1 | 69.2 | 11.2 | 5.4 | 19.4 |
| | | 1 | 156 | 22.8 | 8.3 | 37.5 | 59.6 | 50.9 | 67.2 | 17.5 | 11.6 | 24.5 |
| | | 2 | 118 | 25.8 | 10.6 | 40.9 | 62.0 | 52.2 | 70.5 | 12.2 | 7.0 | 18.9 |
| | 85-94 | 0 | 26 | 6.2 | -31.1 | 49.2 | 70.0 | 43.3 | 85.9 | 23.8 | 7.5 | 45.2 |
| | | 1 | 42 | 20.7 | -4.4 | 46.1 | 71.0 | 52.0 | 83.6 | 8.3 | 1.9 | 20.8 |
| | | 2 | 47 | 9.1 | -12.3 | 32.1 | 79.3 | 64.0 | 88.7 | 11.6 | 3.9 | 23.7 |
| Distant | 65-74 | 0 | 9 | 22.2 | -31.3 | 77.2 | 66.7 | 22.4 | 89.6 | 11.1 | 0.4 | 41.7 |
| | | 1 | 19 | 17.8 | -13.5 | 52.7 | 77.0 | 47.0 | 91.3 | 5.3 | 0.3 | 22.2 |

| | | | | | | | | | | | | |
|---|---|---|---|---|---|---|---|---|---|---|---|---|
| | | 2 | 10 | 26.7 | -25.3 | 79.5 | 63.3 | 20.0 | 87.8 | 10.0 | 0.5 | 37.4 |
| | 75-84 | 0 | 6 | 33.3 | 7.5 | 87.8 | 66.7 | 12.2 | 92.5 | 0.0 | NA | NA |
| | | 1 | 10 | 10.0 | -40.6 | 67.2 | 80.0 | 32.6 | 95.7 | 10.0 | 0.1 | 44.9 |
| | | 2 | 10 | 20.0 | -28.4 | 71.8 | 70.0 | 27.8 | 90.5 | 10.0 | 0.4 | 37.9 |
| | 85-94 | 0 | <5 | 100.0 | 100.0 | 100.0 | 0.0 | NA | NA | 0.0 | NA | NA |
| | | 1 | <5 | 25.0 | 2.0 | 98.3 | 75.0 | 1.7 | 98.0 | 0.0 | NA | NA |
| | | 2 | <5 | 50.0 | 4.0 | 100.0 | 50.0 | 0.0 | 96.0 | 0.0 | NA | NA |

Table S12I2. One and two-year probabilities of dying from cancer, dying from other-causes, and survival by stage, age and comorbidity level: **female oral cancer**

| Stage | Age at diagnosis | Comorbidity | One-year (%) | | | | | | | | | Two-year (%) | | | | | | | | |
|---|---|---|---|---|---|---|---|---|---|---|---|---|---|---|---|---|---|---|---|---|
| | | | Survival | | | Cancer deaths | | | Other-cause deaths | | | Survival | | | Cancer deaths | | | Other-cause deaths | | |
| | | | Est. | 95%CI. | | Est. | 95%CI. | | Est. | 95%CI. | | Est. | 95%CI. | | Est. | 95%CI. | | Est. | 95%CI. | |
| Localized | 65-74 | 0 | 94.1 | 86.3 | 97.9 | 4.4 | 1.8 | 8.9 | 1.5 | 0.3 | 4.8 | 86.4 | 76.6 | 92.7 | 11.4 | 6.7 | 17.5 | 2.2 | 0.6 | 5.9 |
| | | 1 | 93.4 | 88.1 | 96.5 | 5.5 | 3.1 | 8.7 | 1.2 | 0.3 | 3.2 | 86.5 | 79.5 | 91.5 | 11.0 | 7.5 | 15.3 | 2.5 | 1.0 | 5.2 |
| | | 2 | 92.0 | 83.2 | 96.7 | 6.4 | 3.0 | 11.6 | 1.6 | 0.3 | 5.2 | 81.5 | 69.7 | 89.8 | 14.2 | 8.7 | 21.2 | 4.3 | 1.6 | 9.1 |
| | 75-84 | 0 | 94.3 | 79.6 | 99.2 | 3.8 | 0.7 | 11.6 | 1.9 | 0.1 | 8.9 | 82.4 | 63.0 | 93.8 | 11.8 | 4.7 | 22.4 | 5.8 | 1.5 | 14.6 |
| | | 1 | 91.2 | 83.2 | 95.9 | 6.3 | 3.2 | 10.9 | 2.5 | 0.8 | 5.9 | 81.5 | 71.5 | 88.9 | 14.6 | 9.5 | 20.8 | 3.8 | 1.6 | 7.7 |
| | | 2 | 88.1 | 77.6 | 94.8 | 6.8 | 3.2 | 12.3 | 5.1 | 2.1 | 10.1 | 73.1 | 59.2 | 84.1 | 14.7 | 9.0 | 21.8 | 12.2 | 7.0 | 18.9 |
| | 85-94 | 0 | 88.9 | 59.4 | 99.5 | 11.1 | 0.5 | 40.6 | 0.0 | NA | NA | 77.8 | 18.5 | 99.1 | 11.1 | 0.5 | 40.6 | 11.1 | 0.5 | 40.9 |
| | | 1 | 81.3 | 55.1 | 95.1 | 12.5 | 3.9 | 26.5 | 6.3 | 1.1 | 18.4 | 67.5 | 37.1 | 88.3 | 19.2 | 7.6 | 34.7 | 13.4 | 4.1 | 28.2 |
| | | 2 | 81.3 | 66.0 | 92.5 | 18.8 | 7.5 | 34.0 | 0.0 | NA | NA | 60.0 | 29.5 | 81.9 | 33.1 | 16.9 | 50.2 | 6.9 | 1.2 | 20.3 |
| Regional | 65-74 | 0 | 79.8 | 70.4 | 86.6 | 18.4 | 12.9 | 24.7 | 1.8 | 0.5 | 4.9 | 66.9 | 56.1 | 75.8 | 30.0 | 23.0 | 37.2 | 3.1 | 1.2 | 6.7 |
| | | 1 | 77.6 | 70.3 | 83.4 | 20.5 | 15.9 | 25.6 | 1.9 | 0.7 | 4.1 | 62.8 | 53.6 | 70.8 | 31.7 | 26.0 | 37.4 | 5.6 | 3.2 | 8.9 |

| | | | | | | | | | | | | | | | | | | | | |
|---|---|---|---|---|---|---|---|---|---|---|---|---|---|---|---|---|---|---|---|---|
| | | 2 | 64.5 | 51.6 | 75.6 | 26.2 | 19.3 | 33.7 | 9.2 | 5.2 | 14.7 | 50.1 | 35.9 | 63.0 | 37.6 | 29.5 | 45.6 | 12.3 | 7.5 | 18.4 |
| | 75-84 | 0 | 66.3 | 51.8 | 77.6 | 30.6 | 21.6 | 40.0 | 3.2 | 0.8 | 8.2 | 47.5 | 31.7 | 61.4 | 47.1 | 36.6 | 56.9 | 5.4 | 2.0 | 11.4 |
| | | 1 | 58.1 | 46.1 | 68.6 | 36.1 | 28.6 | 43.6 | 5.8 | 2.8 | 10.2 | 40.3 | 27.0 | 52.8 | 49.0 | 40.8 | 56.7 | 10.7 | 6.4 | 16.3 |
| | | 2 | 46.6 | 32.0 | 59.9 | 44.9 | 35.8 | 53.6 | 8.5 | 4.3 | 14.4 | 37.2 | 22.4 | 51.2 | 52.6 | 43.2 | 61.2 | 10.2 | 5.6 | 16.5 |
| | 85-94 | 0 | 30.8 | -1.4 | 61.0 | 57.7 | 36.2 | 74.3 | 11.5 | 2.8 | 27.1 | 30.8 | -1.4 | 61.0 | 57.7 | 36.2 | 74.3 | 11.5 | 2.8 | 27.1 |
| | | 1 | 40.5 | 17.2 | 60.8 | 54.8 | 38.4 | 68.5 | 4.8 | 0.8 | 14.4 | 28.3 | 5.9 | 50.1 | 66.9 | 49.0 | 79.7 | 4.8 | 0.8 | 14.4 |
| | | 2 | 19.1 | -1.7 | 40.3 | 72.3 | 57.0 | 83.0 | 8.5 | 2.7 | 18.7 | 12.2 | -7.4 | 33.3 | 79.3 | 64.0 | 88.7 | 8.5 | 2.7 | 18.7 |
| Distant | 65-74 | 0 | 33.3 | -24.0 | 82.6 | 55.6 | 17.0 | 82.3 | 11.1 | 0.4 | 41.7 | 22.2 | -31.3 | 77.2 | 66.7 | 22.4 | 89.6 | 11.1 | 0.4 | 41.7 |
| | | 1 | 41.4 | 4.6 | 71.7 | 53.3 | 28.0 | 73.3 | 5.3 | 0.3 | 22.2 | 29.6 | -5.2 | 62.4 | 65.1 | 37.3 | 83.0 | 5.3 | 0.3 | 22.2 |
| | | 2 | 40.0 | -14.4 | 83.6 | 50.0 | 16.0 | 77.0 | 10.0 | 0.5 | 37.4 | 26.7 | -25.3 | 79.5 | 63.3 | 20.0 | 87.8 | 10.0 | 0.5 | 37.4 |
| | 75-84 | 0 | 33.3 | 7.5 | 87.8 | 66.7 | 12.2 | 92.5 | 0.0 | NA | NA | 33.3 | 7.5 | 87.8 | 66.7 | 12.2 | 92.5 | 0.0 | NA | NA |
| | | 1 | 50.0 | 23.2 | 83.7 | 50.0 | 16.3 | 76.8 | 0.0 | NA | NA | 30.0 | 9.6 | 71.8 | 70.0 | 28.2 | 90.4 | 0.0 | NA | NA |
| | | 2 | 20.0 | -28.4 | 71.8 | 70.0 | 27.8 | 90.5 | 10.0 | 0.4 | 37.9 | 20.0 | -28.4 | 71.8 | 70.0 | 27.8 | 90.5 | 10.0 | 0.4 | 37.9 |
| | 85-94 | 0 | 100.0 | 100.0 | 100.0 | 0.0 | NA | NA | 0.0 | NA | NA | 100.0 | 100.0 | 100.0 | 0.0 | NA | NA | 0.0 | NA | NA |
| | | 1 | 50.0 | 11.9 | 97.7 | 50.0 | 2.3 | 88.1 | 0.0 | NA | NA | 25.0 | 2.0 | 98.3 | 75.0 | 1.7 | 98.0 | 0.0 | NA | NA |
| | | 2 | 50.0 | 4.0 | 100.0 | 50.0 | 0.0 | 96.0 | 0.0 | NA | NA | 50.0 | 4.0 | 100.0 | 50.0 | 0.0 | 96.0 | 0.0 | NA | NA |

Table S13A. Number of patients and probabilities of dying from cancer, dying from other-causes, and survival among patients who were diagnosed with distant lung cancer at ages 30—94 in 2004—2014 by subtypes and comorbidity level

| Subtype | Comorbidity | N | One-year | | | | | | | | |
|---|---|---|---|---|---|---|---|---|---|---|---|
| | | | Survival (%) | | | Cancer deaths (%) | | | Other-cause deaths (%) | | |
| | | | Estimate | 95%CI | | Estimate | 95%CI | | Estimate | 95%%CI | |
| SCC | 0 | 2907 | 30.5 | 28.2 | 32.8 | 66.7 | 64.9 | 68.4 | 2.8 | 2.2 | 3.4 |
| | 1 | 2285 | 25.9 | 23.4 | 28.3 | 71.6 | 69.7 | 73.4 | 2.5 | 1.9 | 3.2 |
| | 2 | 2501 | 22.6 | 20.0 | 25.1 | 73.3 | 71.5 | 75.0 | 4.1 | 3.4 | 5.0 |
| ADC | 0 | 15277 | 53.8 | 52.8 | 54.7 | 44.8 | 44.0 | 45.6 | 1.4 | 1.2 | 1.6 |
| | 1 | 11655 | 50.9 | 49.8 | 52.1 | 47.1 | 46.2 | 48.0 | 2.0 | 1.7 | 2.2 |
| | 2 | 7001 | 37.2 | 35.6 | 38.8 | 59.5 | 58.4 | 60.6 | 3.3 | 2.9 | 3.7 |
| SCLC | 0 | 403 | 35.0 | 28.7 | 41.0 | 63.0 | 58.1 | 67.5 | 2.0 | 0.9 | 3.7 |
| | 1 | 272 | 29.8 | 21.8 | 37.5 | 66.5 | 60.6 | 71.8 | 3.7 | 1.9 | 6.4 |
| | 2 | 203 | 26.1 | 17.3 | 34.6 | 70.9 | 64.2 | 76.7 | 3.0 | 1.2 | 6.0 |
| Subtype | Comorbidity | N | Two-year | | | | | | | | |
| SCC | 0 | 2907 | 12.3 | 10.4 | 14.3 | 84.6 | 83.2 | 85.8 | 3.1 | 2.5 | 3.8 |
| | 1 | 2285 | 9.2 | 7.1 | 11.4 | 87.2 | 85.8 | 88.5 | 3.6 | 2.8 | 4.4 |
| | 2 | 2501 | 8.5 | 6.3 | 10.6 | 87.1 | 85.7 | 88.3 | 4.5 | 3.7 | 5.3 |
| ADC | 0 | 15277 | 28.8 | 27.8 | 29.7 | 69.3 | 68.5 | 70.0 | 1.9 | 1.7 | 2.2 |
| | 1 | 11655 | 27.4 | 26.2 | 28.5 | 69.7 | 68.8 | 70.5 | 2.9 | 2.6 | 3.3 |
| | 2 | 7001 | 18.9 | 17.4 | 20.4 | 76.9 | 75.8 | 77.8 | 4.2 | 3.8 | 4.7 |
| SCLC | 0 | 403 | 17.5 | 12.0 | 22.9 | 80.3 | 75.9 | 83.9 | 2.3 | 1.1 | 4.1 |

| | | | | | | | | | | | |
|---|---|---|---|---|---|---|---|---|---|---|---|
| | 1 | 272 | 12.9 | 6.1 | 19.9 | 83.1 | 78.0 | 87.1 | 4.0 | 2.1 | 6.9 |
| | 2 | 203 | 11.1 | 3.9 | 18.6 | 85.9 | 80.2 | 90.1 | 3.0 | 1.2 | 6.0 |
| Subtype | Comorbidity | N | Five-year | | | | | | | | |
| SCC | 0 | 2907 | 3.2 | 1.6 | 4.8 | 93.4 | 92.4 | 94.3 | 3.4 | 2.8 | 4.1 |
| | 1 | 2285 | 2.1 | 0.3 | 4.0 | 94.0 | 92.9 | 95.0 | 3.9 | 3.1 | 4.7 |
| | 2 | 2501 | 0.9 | -0.9 | 2.8 | 93.9 | 92.9 | 94.8 | 5.2 | 4.3 | 6.1 |
| ADC | 0 | 15277 | 5.1 | 4.3 | 5.8 | 92.5 | 92.0 | 92.9 | 2.5 | 2.2 | 2.7 |
| | 1 | 11655 | 5.2 | 4.2 | 6.1 | 91.0 | 90.4 | 91.6 | 3.8 | 3.5 | 4.2 |
| | 2 | 7001 | 3.6 | 2.4 | 4.9 | 91.3 | 90.6 | 92.0 | 5.1 | 4.5 | 5.6 |
| SCLC | 0 | 403 | 4.0 | -0.2 | 8.7 | 93.4 | 90.0 | 95.7 | 2.5 | 1.3 | 4.4 |
| | 1 | 272 | 1.2 | -4.5 | 7.5 | 93.7 | 89.7 | 96.2 | 5.1 | 2.8 | 8.3 |
| | 2 | 203 | 2.8 | -3.0 | 9.4 | 94.3 | 89.3 | 97.0 | 3.0 | 1.2 | 6.0 |

Table S13B. Number of patients and probabilities of dying from cancer, dying from other-causes, and survival among patients who were diagnosed with distant lung ADC at ages 30—94 by comorbidity level for period 2004—2010 and 2011—2014

| Period | Comorbidity | N | One-year | | | | | | | | |
|---|---|---|---|---|---|---|---|---|---|---|---|
| | | | Survival (%) | | | Cancer deaths (%) | | | Other-cause deaths (%) | | |
| | | | Estimate | 95%CI. | | Estimate | 95%CI. | | Estimate | 95%CI. | |
| 2004-2010 | 0 | 7994 | 50.5 | 49.1 | 51.8 | 48.1 | 47.0 | 49.2 | 1.5 | 1.2 | 1.7 |
| | 1 | 5730 | 47.8 | 46.2 | 49.5 | 50.2 | 48.9 | 51.5 | 1.9 | 1.6 | 2.3 |
| | 2 | 3481 | 34.6 | 32.4 | 36.8 | 62.4 | 60.8 | 64.0 | 3.0 | 2.5 | 3.6 |
| 2011-2014 | 0 | 7283 | 57.3 | 55.9 | 58.7 | 41.3 | 40.1 | 42.4 | 1.4 | 1.1 | 1.7 |
| | 1 | 5925 | 54.0 | 52.3 | 55.6 | 44.1 | 42.8 | 45.3 | 2.0 | 1.7 | 2.4 |
| | 2 | 3520 | 39.8 | 37.5 | 42.0 | 56.7 | 55.0 | 58.3 | 3.6 | 3.0 | 4.2 |
| Period | Comorbidity | N | Two-year | | | | | | | | |
| 2004-2010 | 0 | 7994 | 27.2 | 25.9 | 28.5 | 70.8 | 69.8 | 71.8 | 2.0 | 1.7 | 2.3 |
| | 1 | 5730 | 26.0 | 24.4 | 27.6 | 71.3 | 70.1 | 72.5 | 2.7 | 2.3 | 3.1 |
| | 2 | 3481 | 17.5 | 15.5 | 19.5 | 78.7 | 77.3 | 80.0 | 3.8 | 3.2 | 4.4 |
| 2011-2014 | 0 | 7283 | 30.4 | 29.0 | 31.9 | 67.7 | 66.5 | 68.8 | 1.9 | 1.6 | 2.2 |
| | 1 | 5925 | 28.6 | 27.0 | 30.3 | 68.1 | 66.9 | 69.3 | 3.2 | 2.8 | 3.7 |
| | 2 | 3520 | 20.2 | 18.1 | 22.4 | 75.1 | 73.6 | 76.5 | 4.7 | 4.0 | 5.4 |
| Period | Comorbidity | N | Five-year | | | | | | | | |
| 2004-2010 | 0 | 7994 | 5.6 | 4.7 | 6.5 | 92.0 | 91.4 | 92.6 | 2.4 | 2.1 | 2.8 |
| | 1 | 5730 | 5.9 | 4.7 | 7.1 | 90.7 | 89.9 | 91.4 | 3.4 | 2.9 | 3.9 |
| | 2 | 3481 | 4.3 | 2.7 | 6.0 | 91.2 | 90.2 | 92.1 | 4.4 | 3.8 | 5.2 |

| | | | | | | | | | | | |
|---|---|---|---|---|---|---|---|---|---|---|---|
| 2011-2014 | 0 | 7283 | 2.6 | 1.4 | 3.9 | 94.9 | 94.0 | 95.7 | 2.5 | 2.1 | 2.9 |
| | 1 | 5925 | 2.4 | 0.8 | 4.2 | 93.2 | 92.0 | 94.2 | 4.3 | 3.8 | 4.9 |
| | 2 | 3520 | 1.2 | -0.8 | 3.2 | 93.1 | 91.8 | 94.2 | 5.7 | 5.0 | 6.6 |

Table S14A. Number of patients and **one**-year probabilities of dying from cancer, dying from other-causes, and survival among **males** who were diagnosed with squamous cell carcinoma lung cancer **(SCC)** by stage, age and comorbidity level (n>100)

| Stage | Age | Comorbidity | N | Survival (%) | | | Cancer deaths (%) | | | Other-Cause Deaths (%) | | |
|---|---|---|---|---|---|---|---|---|---|---|---|---|
| | | | | Estimate | 95%CI. | | Estimate | 95%CI. | | Estimate | 95%CI. | |
| Localized | 65-74 | 0 | 156 | 84.6 | 75.4 | 91.0 | 12.8 | 8.1 | 18.6 | 2.6 | 0.8 | 6.0 |
| | | 1 | 226 | 83.6 | 76.0 | 89.3 | 13.7 | 9.6 | 18.6 | 2.7 | 1.1 | 5.4 |
| | | 2 | 303 | 77.2 | 70.2 | 82.9 | 20.2 | 15.8 | 24.9 | 2.6 | 1.2 | 4.9 |
| | 75-84 | 0 | 135 | 70.4 | 58.2 | 80.2 | 24.5 | 17.6 | 32.0 | 5.2 | 2.3 | 9.9 |
| | | 1 | 188 | 71.8 | 61.8 | 80.0 | 23.4 | 17.6 | 29.7 | 4.8 | 2.4 | 8.5 |
| | | 2 | 387 | 68.0 | 61.1 | 74.0 | 27.9 | 23.5 | 32.4 | 4.1 | 2.5 | 6.5 |
| | 85-94 | 0 | 0 | 0.0 | 0.0 | 0.0 | 0.0 | 0.0 | 0.0 | 0.0 | 0.0 | 0.0 |
| | | 1 | 0 | 0.0 | 0.0 | 0.0 | 0.0 | 0.0 | 0.0 | 0.0 | 0.0 | 0.0 |
| | | 2 | 0 | 0.0 | 0.0 | 0.0 | 0.0 | 0.0 | 0.0 | 0.0 | 0.0 | 0.0 |
| Regional | 65-74 | 0 | 567 | 50.0 | 44.4 | 55.2 | 47.5 | 43.3 | 51.6 | 2.5 | 1.4 | 4.0 |
| | | 1 | 494 | 52.7 | 46.7 | 58.3 | 44.8 | 40.4 | 49.2 | 2.4 | 1.3 | 4.1 |
| | | 2 | 581 | 50.5 | 44.8 | 55.8 | 46.4 | 42.3 | 50.4 | 3.1 | 1.9 | 4.8 |
| | 75-84 | 0 | 410 | 36.0 | 29.7 | 42.0 | 61.8 | 56.9 | 66.3 | 2.2 | 1.1 | 4.0 |
| | | 1 | 490 | 39.3 | 33.2 | 45.1 | 57.4 | 52.9 | 61.7 | 3.3 | 2.0 | 5.1 |
| | | 2 | 855 | 35.7 | 31.0 | 40.4 | 59.5 | 56.1 | 62.7 | 4.8 | 3.5 | 6.4 |
| | 85-94 | 0 | 0 | 0.0 | 0.0 | 0.0 | 0.0 | 0.0 | 0.0 | 0.0 | 0.0 | 0.0 |
| | | 1 | 0 | 0.0 | 0.0 | 0.0 | 0.0 | 0.0 | 0.0 | 0.0 | 0.0 | 0.0 |
| | | 2 | 207 | 29.5 | 20.1 | 38.6 | 66.2 | 59.3 | 72.2 | 4.3 | 2.1 | 7.8 |

| | | | | | | | | | | | | |
|---|---|---|---|---|---|---|---|---|---|---|---|---|
| Distant | 65-74 | 0 | 707 | 24.2 | 19.6 | 28.8 | 72.5 | 69.1 | 75.7 | 3.3 | 2.1 | 4.8 |
| | | 1 | 611 | 24.9 | 20.1 | 29.5 | 73.0 | 69.3 | 76.4 | 2.1 | 1.2 | 3.5 |
| | | 2 | 644 | 26.9 | 22.0 | 31.7 | 70.1 | 66.4 | 73.5 | 3.0 | 1.8 | 4.5 |
| | 75-84 | 0 | 606 | 18.7 | 14.0 | 23.5 | 77.8 | 74.3 | 80.9 | 3.5 | 2.2 | 5.2 |
| | | 1 | 666 | 20.2 | 15.8 | 24.5 | 77.4 | 74.1 | 80.4 | 2.4 | 1.4 | 3.8 |
| | | 2 | 1036 | 18.2 | 14.4 | 22.0 | 77.3 | 74.6 | 79.7 | 4.5 | 3.3 | 5.9 |
| | 85-94 | 0 | 115 | 11.2 | 1.1 | 21.7 | 85.3 | 77.1 | 90.7 | 3.6 | 1.2 | 8.3 |
| | | 1 | 178 | 9.9 | 1.5 | 18.7 | 85.0 | 78.8 | 89.5 | 5.1 | 2.5 | 9.0 |
| | | 2 | 280 | 11.1 | 3.7 | 18.6 | 81.8 | 76.8 | 85.8 | 7.1 | 4.5 | 10.5 |

Table S14B. Number of patients and **two**-year probabilities of dying from cancer, dying from other-causes, and survival among **males** who were diagnosed with squamous cell carcinoma lung cancer **(SCC)** by stage, age and comorbidity level (n>100)

| Stage | Age | Comorbidity | N | Survival (%) | | | Cancer deaths (%) | | | Other-Cause Deaths (%) | | |
|---|---|---|---|---|---|---|---|---|---|---|---|---|
| | | | | Estimate | 95%CI. | | Estimate | 95%CI. | | Estimate | 95%CI. | |
| Localized | 65-74 | 0 | 156 | 68.3 | 57.2 | 77.3 | 28.5 | 21.5 | 35.9 | 3.2 | 1.2 | 6.9 |
| | | 1 | 226 | 72.4 | 63.3 | 79.9 | 23.1 | 17.8 | 28.9 | 4.5 | 2.3 | 7.8 |
| | | 2 | 303 | 65.6 | 57.6 | 72.6 | 30.3 | 25.2 | 35.6 | 4.0 | 2.2 | 6.7 |
| | 75-84 | 0 | 135 | 48.0 | 34.4 | 60.3 | 44.5 | 35.8 | 52.7 | 7.5 | 3.8 | 12.9 |
| | | 1 | 188 | 57.3 | 45.9 | 67.4 | 35.1 | 28.3 | 42.0 | 7.6 | 4.4 | 12.0 |
| | | 2 | 387 | 47.6 | 39.9 | 54.9 | 45.9 | 40.8 | 50.8 | 6.5 | 4.3 | 9.3 |
| | 85-94 | 0 | 0 | 0.0 | 0.0 | 0.0 | 0.0 | 0.0 | 0.0 | 0.0 | 0.0 | 0.0 |
| | | 1 | 0 | 0.0 | 0.0 | 0.0 | 0.0 | 0.0 | 0.0 | 0.0 | 0.0 | 0.0 |

| | | | | | | | | | | | | |
|---|---|---|---|---|---|---|---|---|---|---|---|---|
| | | 2 | 0 | 0.0 | 0.0 | 0.0 | 0.0 | 0.0 | 0.0 | 0.0 | 0.0 | 0.0 |
| Regional | 65-74 | 0 | 567 | 26.6 | 21.2 | 31.8 | 70.0 | 66.1 | 73.7 | 3.4 | 2.1 | 5.1 |
| | | 1 | 494 | 30.1 | 24.4 | 35.6 | 67.5 | 63.1 | 71.5 | 2.4 | 1.3 | 4.1 |
| | | 2 | 581 | 28.7 | 23.0 | 34.3 | 66.4 | 62.3 | 70.1 | 4.9 | 3.3 | 6.9 |
| | 75-84 | 0 | 410 | 14.7 | 9.3 | 20.1 | 82.4 | 78.3 | 85.8 | 2.9 | 1.6 | 4.9 |
| | | 1 | 490 | 18.0 | 12.6 | 23.5 | 78.1 | 74.1 | 81.5 | 3.9 | 2.4 | 5.9 |
| | | 2 | 855 | 14.8 | 10.5 | 19.0 | 79.7 | 76.8 | 82.3 | 5.5 | 4.1 | 7.2 |
| | 85-94 | 0 | 0 | 0.0 | 0.0 | 0.0 | 0.0 | 0.0 | 0.0 | 0.0 | 0.0 | 0.0 |
| | | 1 | 0 | 0.0 | 0.0 | 0.0 | 0.0 | 0.0 | 0.0 | 0.0 | 0.0 | 0.0 |
| | | 2 | 207 | 8.7 | -0.3 | 18.1 | 82.4 | 76.5 | 87.0 | 8.9 | 5.5 | 13.3 |
| Distant | 65-74 | 0 | 707 | 8.9 | 5.2 | 12.8 | 87.7 | 85.0 | 89.9 | 3.4 | 2.2 | 4.9 |
| | | 1 | 611 | 8.8 | 5.0 | 12.7 | 88.7 | 85.9 | 91.0 | 2.5 | 1.4 | 3.9 |
| | | 2 | 644 | 9.7 | 5.7 | 13.8 | 87.0 | 84.1 | 89.4 | 3.3 | 2.1 | 4.9 |
| | 75-84 | 0 | 606 | 5.5 | 1.7 | 9.5 | 90.6 | 88.0 | 92.7 | 3.8 | 2.5 | 5.6 |
| | | 1 | 666 | 5.5 | 1.9 | 9.3 | 90.7 | 88.2 | 92.7 | 3.8 | 2.5 | 5.5 |
| | | 2 | 1036 | 6.7 | 3.5 | 10.1 | 88.3 | 86.2 | 90.1 | 5.0 | 3.8 | 6.4 |
| | 85-94 | 0 | 115 | 3.7 | -4.6 | 13.1 | 92.7 | 85.7 | 96.4 | 3.6 | 1.2 | 8.3 |
| | | 1 | 178 | 2.3 | -5.6 | 10.9 | 90.8 | 85.4 | 94.3 | 6.8 | 3.7 | 11.3 |
| | | 2 | 280 | 3.2 | -3.3 | 10.1 | 89.3 | 85.1 | 92.4 | 7.5 | 4.8 | 10.9 |

Table S14C: Number of patients and **five**-year probabilities of dying from cancer, dying from other-causes, and survival among **males** who were diagnosed with squamous cell carcinoma lung cancer **(SCC)** by stage, age and comorbidity level (n>100)

| Stage | Age | Comorbidity | N | Survival (%) | | | Cancer deaths (%) | | | Other-Cause Deaths (%) | | |
|---|---|---|---|---|---|---|---|---|---|---|---|---|
| | | | | Estimate | 95%CI. | | Estimate | 95%CI. | | Estimate | 95%CI. | |
| Localized | 65-74 | 0 | 156 | 44.1 | 30.3 | 56.7 | 48.3 | 39.5 | 56.7 | 7.6 | 3.8 | 13.0 |
| | | 1 | 226 | 51.3 | 39.3 | 62.3 | 37.0 | 30.2 | 43.8 | 11.8 | 7.6 | 16.9 |
| | | 2 | 303 | 36.4 | 26.7 | 45.8 | 54.6 | 48.3 | 60.5 | 9.0 | 5.9 | 12.8 |
| | 75-84 | 0 | 135 | 29.9 | 16.5 | 43.0 | 61.7 | 52.6 | 69.6 | 8.4 | 4.4 | 13.9 |
| | | 1 | 188 | 29.9 | 16.2 | 43.3 | 54.1 | 46.0 | 61.5 | 16.0 | 10.7 | 22.2 |
| | | 2 | 387 | 22.1 | 13.4 | 30.8 | 66.2 | 60.6 | 71.1 | 11.8 | 8.6 | 15.6 |
| | 85-94 | 0 | 0 | 0.0 | 0.0 | 0.0 | 0.0 | 0.0 | 0.0 | 0.0 | 0.0 | 0.0 |
| | | 1 | 0 | 0.0 | 0.0 | 0.0 | 0.0 | 0.0 | 0.0 | 0.0 | 0.0 | 0.0 |
| | | 2 | 0 | 0.0 | 0.0 | 0.0 | 0.0 | 0.0 | 0.0 | 0.0 | 0.0 | 0.0 |
| Regional | 65-74 | 0 | 567 | 10.4 | 5.5 | 15.4 | 84.8 | 81.3 | 87.6 | 4.8 | 3.2 | 6.9 |
| | | 1 | 494 | 13.0 | 7.6 | 18.6 | 82.7 | 78.8 | 86.0 | 4.2 | 2.6 | 6.4 |
| | | 2 | 581 | 11.9 | 6.7 | 17.3 | 82.0 | 78.4 | 85.0 | 6.1 | 4.3 | 8.3 |
| | 75-84 | 0 | 410 | 6.4 | 1.6 | 11.4 | 90.0 | 86.5 | 92.7 | 3.6 | 2.0 | 5.8 |
| | | 1 | 490 | 6.7 | 1.6 | 12.0 | 87.5 | 84.1 | 90.2 | 5.8 | 3.9 | 8.2 |
| | | 2 | 855 | 3.8 | 0.0 | 7.6 | 89.6 | 87.3 | 91.6 | 6.6 | 5.1 | 8.4 |
| | 85-94 | 0 | 0 | 0.0 | 0.0 | 0.0 | 0.0 | 0.0 | 0.0 | 0.0 | 0.0 | 0.0 |
| | | 1 | 0 | 0.0 | 0.0 | 0.0 | 0.0 | 0.0 | 0.0 | 0.0 | 0.0 | 0.0 |
| | | 2 | 207 | 0.9 | -7.7 | 10.2 | 88.6 | 83.0 | 92.4 | 10.5 | 6.8 | 15.3 |

| | | | | | | | | | | | | |
|---|---|---|---|---|---|---|---|---|---|---|---|---|
| Distant | 65-74 | 0 | 707 | 1.6 | -1.4 | 4.8 | 94.8 | 92.8 | 96.3 | 3.6 | 2.4 | 5.2 |
| | | 1 | 611 | 1.8 | -1.2 | 5.1 | 95.8 | 93.5 | 97.3 | 2.5 | 1.4 | 3.9 |
| | | 2 | 644 | 0.8 | -2.4 | 4.3 | 95.1 | 93.1 | 96.6 | 4.0 | 2.7 | 5.8 |
| | 75-84 | 0 | 606 | 0.9 | -2.4 | 4.6 | 94.9 | 92.7 | 96.4 | 4.2 | 2.8 | 6.0 |
| | | 1 | 666 | 1.4 | -2.0 | 4.9 | 94.1 | 92.0 | 95.7 | 4.5 | 3.1 | 6.3 |
| | | 2 | 1036 | 0.4 | -2.5 | 3.3 | 93.9 | 92.3 | 95.2 | 5.7 | 4.4 | 7.2 |
| | 85-94 | 0 | 115 | 1.4 | -7.2 | 11.9 | 94.1 | 86.5 | 97.5 | 4.5 | 1.6 | 9.7 |
| | | 1 | 178 | 2.3 | -5.6 | 10.9 | 90.8 | 85.4 | 94.3 | 6.8 | 3.7 | 11.3 |
| | | 2 | 280 | 3.2 | -3.3 | 10.1 | 89.3 | 85.1 | 92.4 | 7.5 | 4.8 | 10.9 |

Table S14D: Number of patients and **one**-year probabilities of dying from cancer, dying from other-causes, and survival among **males** who were diagnosed with lung adenocarcinoma **(ADC)** by stage, age and comorbidity level (n>100)

| Stage | Age | Comorbidity | N | Survival (%) | | | Cancer deaths (%) | | | Other-Cause Deaths (%) | | |
|---|---|---|---|---|---|---|---|---|---|---|---|---|
| | | | | Estimate | 95CI. | | Estimate | 95CI. | | Estimate | 95CI. | |
| Localized | 65-74 | 0 | 336 | 92.5 | 87.9 | 95.7 | 6.0 | 3.8 | 8.9 | 1.5 | 0.6 | 3.3 |
| | | 1 | 508 | 92.9 | 89.4 | 95.3 | 6.3 | 4.4 | 8.6 | 0.8 | 0.3 | 1.9 |
| | | 2 | 314 | 91.4 | 86.4 | 94.8 | 7.3 | 4.8 | 10.6 | 1.3 | 0.4 | 3.1 |
| | 75-84 | 0 | 168 | 82.1 | 72.9 | 88.7 | 15.5 | 10.5 | 21.5 | 2.4 | 0.8 | 5.6 |
| | | 1 | 293 | 87.0 | 81.1 | 91.3 | 11.6 | 8.3 | 15.6 | 1.4 | 0.5 | 3.3 |
| | | 2 | 406 | 80.3 | 74.5 | 85.1 | 16.8 | 13.3 | 20.6 | 3.0 | 1.6 | 5.0 |
| | 85-94 | 0 | 0 | 0.0 | 0.0 | 0.0 | 0.0 | 0.0 | 0.0 | 0.0 | 0.0 | 0.0 |
| | | 1 | 0 | 0.0 | 0.0 | 0.0 | 0.0 | 0.0 | 0.0 | 0.0 | 0.0 | 0.0 |

| | | | | | | | | | | | | |
|---|---|---|---|---|---|---|---|---|---|---|---|---|
| | | 2 | 0 | 0.0 | 0.0 | 0.0 | 0.0 | 0.0 | 0.0 | 0.0 | 0.0 | 0.0 |
| Regional | 65-74 | 0 | 431 | 62.1 | 55.9 | 67.7 | 35.8 | 31.3 | 40.3 | 2.1 | 1.0 | 3.8 |
| | | 1 | 431 | 70.3 | 64.3 | 75.6 | 27.4 | 23.3 | 31.7 | 2.3 | 1.2 | 4.1 |
| | | 2 | 338 | 55.9 | 48.5 | 62.6 | 41.1 | 35.8 | 46.3 | 3.0 | 1.5 | 5.2 |
| | 75-84 | 0 | 285 | 45.3 | 37.3 | 52.8 | 51.9 | 45.9 | 57.5 | 2.8 | 1.3 | 5.2 |
| | | 1 | 432 | 54.5 | 48.4 | 60.2 | 43.8 | 39.1 | 48.5 | 1.6 | 0.7 | 3.2 |
| | | 2 | 513 | 44.8 | 38.7 | 50.5 | 51.7 | 47.3 | 56.0 | 3.5 | 2.2 | 5.4 |
| | 85-94 | 0 | 0 | 0.0 | 0.0 | 0.0 | 0.0 | 0.0 | 0.0 | 0.0 | 0.0 | 0.0 |
| | | 1 | 0 | 0.0 | 0.0 | 0.0 | 0.0 | 0.0 | 0.0 | 0.0 | 0.0 | 0.0 |
| | | 2 | 127 | 34.3 | 20.9 | 47.1 | 58.6 | 49.4 | 66.6 | 7.1 | 3.5 | 12.5 |
| Distant | 65-74 | 0 | 1669 | 40.4 | 37.3 | 43.4 | 57.5 | 55.1 | 59.9 | 2.0 | 1.4 | 2.8 |
| | | 1 | 1714 | 44.5 | 41.4 | 47.5 | 53.2 | 50.8 | 55.6 | 2.3 | 1.6 | 3.1 |
| | | 2 | 1064 | 34.7 | 30.9 | 38.5 | 62.9 | 59.9 | 65.7 | 2.4 | 1.6 | 3.4 |
| | 75-84 | 0 | 1074 | 28.3 | 24.6 | 32.0 | 69.1 | 66.3 | 71.8 | 2.5 | 1.7 | 3.6 |
| | | 1 | 1574 | 32.7 | 29.5 | 35.8 | 64.8 | 62.4 | 67.1 | 2.5 | 1.8 | 3.3 |
| | | 2 | 1729 | 26.8 | 23.7 | 29.9 | 69.1 | 66.9 | 71.2 | 4.1 | 3.2 | 5.1 |
| | 85-94 | 0 | 229 | 21.7 | 13.6 | 29.7 | 74.8 | 68.7 | 79.9 | 3.5 | 1.6 | 6.5 |
| | | 1 | 406 | 25.1 | 18.4 | 31.7 | 69.5 | 64.8 | 73.7 | 5.4 | 3.5 | 7.9 |
| | | 2 | 573 | 21.1 | 15.8 | 26.3 | 74.4 | 70.6 | 77.7 | 4.6 | 3.1 | 6.5 |

Table S14E: Number of patients and **two**-year probabilities of dying from cancer, dying from other-causes, and survival among **males** who were diagnosed with lung adenocarcinoma **(ADC)** by stage, age and comorbidity level (n>100)

| Stage | Age | Comorbidity | N | Survival (%) | | | Cancer deaths (%) | | | Other-Cause Deaths (%) | | |
|---|---|---|---|---|---|---|---|---|---|---|---|---|
| | | | | Estimate | 95%CI. | | Estimate | 95%CI. | | Estimate | 95%CI. | |
| Localized | 65-74 | 0 | 336 | 84.3 | 78.1 | 89.1 | 13.0 | 9.6 | 16.9 | 2.7 | 1.3 | 4.9 |
| | | 1 | 508 | 85.9 | 81.3 | 89.6 | 12.0 | 9.3 | 15.1 | 2.0 | 1.0 | 3.6 |
| | | 2 | 314 | 82.5 | 75.8 | 87.8 | 14.2 | 10.5 | 18.4 | 3.4 | 1.7 | 5.9 |
| | 75-84 | 0 | 168 | 69.2 | 58.6 | 77.8 | 27.8 | 21.0 | 34.8 | 3.1 | 1.1 | 6.6 |
| | | 1 | 293 | 81.1 | 74.4 | 86.4 | 16.8 | 12.7 | 21.4 | 2.1 | 0.9 | 4.3 |
| | | 2 | 406 | 63.8 | 56.7 | 70.2 | 31.1 | 26.6 | 35.8 | 5.1 | 3.2 | 7.6 |
| | 85-94 | 0 | 0 | 0.0 | 0.0 | 0.0 | 0.0 | 0.0 | 0.0 | 0.0 | 0.0 | 0.0 |
| | | 1 | 0 | 0.0 | 0.0 | 0.0 | 0.0 | 0.0 | 0.0 | 0.0 | 0.0 | 0.0 |
| | | 2 | 0 | 0.0 | 0.0 | 0.0 | 0.0 | 0.0 | 0.0 | 0.0 | 0.0 | 0.0 |
| Regional | 65-74 | 0 | 431 | 43.2 | 36.6 | 49.5 | 53.5 | 48.6 | 58.1 | 3.3 | 1.9 | 5.3 |
| | | 1 | 431 | 48.8 | 42.1 | 55.0 | 48.1 | 43.3 | 52.8 | 3.0 | 1.7 | 5.0 |
| | | 2 | 338 | 38.3 | 30.4 | 45.7 | 56.9 | 51.4 | 62.1 | 4.8 | 2.9 | 7.5 |
| | 75-84 | 0 | 285 | 28.5 | 20.6 | 36.2 | 67.6 | 61.7 | 72.7 | 3.9 | 2.1 | 6.7 |
| | | 1 | 432 | 31.5 | 25.0 | 37.7 | 65.0 | 60.2 | 69.3 | 3.5 | 2.1 | 5.6 |
| | | 2 | 513 | 27.1 | 21.1 | 33.0 | 68.0 | 63.7 | 71.9 | 4.9 | 3.3 | 7.0 |
| | 85-94 | 0 | 0 | 0.0 | 0.0 | 0.0 | 0.0 | 0.0 | 0.0 | 0.0 | 0.0 | 0.0 |
| | | 1 | 0 | 0.0 | 0.0 | 0.0 | 0.0 | 0.0 | 0.0 | 0.0 | 0.0 | 0.0 |
| | | 2 | 127 | 12.9 | 0.9 | 25.4 | 79.1 | 70.5 | 85.4 | 8.0 | 4.1 | 13.7 |

| | | | | | | | | | | | | |
|---|---|---|---|---|---|---|---|---|---|---|---|---|
| Distant | 65-74 | 0 | 1669 | 19.5 | 16.7 | 22.3 | 77.8 | 75.7 | 79.8 | 2.7 | 2.0 | 3.6 |
| | | 1 | 1714 | 23.1 | 20.2 | 26.0 | 74.0 | 71.9 | 76.1 | 2.9 | 2.2 | 3.8 |
| | | 2 | 1064 | 17.7 | 14.3 | 21.2 | 79.4 | 76.8 | 81.8 | 2.8 | 2.0 | 4.0 |
| | 75-84 | 0 | 1074 | 11.4 | 8.2 | 14.6 | 85.5 | 83.2 | 87.5 | 3.1 | 2.2 | 4.3 |
| | | 1 | 1574 | 13.2 | 10.4 | 16.0 | 83.0 | 81.0 | 84.7 | 3.9 | 3.0 | 4.9 |
| | | 2 | 1729 | 11.4 | 8.6 | 14.3 | 83.0 | 81.1 | 84.6 | 5.6 | 4.6 | 6.8 |
| | 85-94 | 0 | 229 | 6.7 | 0.1 | 13.7 | 89.3 | 84.4 | 92.7 | 4.0 | 2.0 | 7.1 |
| | | 1 | 406 | 8.6 | 2.7 | 14.8 | 84.4 | 80.5 | 87.6 | 6.9 | 4.7 | 9.7 |
| | | 2 | 573 | 8.4 | 3.8 | 13.2 | 86.1 | 83.0 | 88.7 | 5.5 | 3.8 | 7.5 |

Table S14F: Number of patients and **five**-year probabilities of dying from cancer, dying from other-causes, and survival among **males** who were diagnosed with lung adenocarcinoma **(ADC)** by stage, age and comorbidity level (n>100)

| Stage | Age | Comorbidity | N | Survival (%) | | | Cancer deaths (%) | | | Other-Cause Deaths (%) | | |
|---|---|---|---|---|---|---|---|---|---|---|---|---|
| | | | | Estimate | 95%CI. | | Estimate | 95%CI. | | Estimate | 95%CI. | |
| Localized | 65-74 | 0 | 336 | 58.9 | 50.1 | 66.9 | 36.8 | 30.7 | 42.8 | 4.3 | 2.4 | 7.1 |
| | | 1 | 508 | 65.8 | 58.3 | 72.4 | 29.4 | 24.6 | 34.2 | 4.9 | 3.0 | 7.4 |
| | | 2 | 314 | 55.0 | 44.8 | 64.4 | 36.4 | 30.1 | 42.7 | 8.6 | 5.5 | 12.6 |
| | 75-84 | 0 | 168 | 44.2 | 30.0 | 57.3 | 46.0 | 37.4 | 54.3 | 9.7 | 5.3 | 15.7 |
| | | 1 | 293 | 49.2 | 38.9 | 58.8 | 44.8 | 38.0 | 51.4 | 5.9 | 3.2 | 9.7 |
| | | 2 | 406 | 33.0 | 23.8 | 42.0 | 54.0 | 48.4 | 59.3 | 13.0 | 9.6 | 16.9 |
| | 85-94 | 0 | 0 | 0.0 | 0.0 | 0.0 | 0.0 | 0.0 | 0.0 | 0.0 | 0.0 | 0.0 |
| | | 1 | 0 | 0.0 | 0.0 | 0.0 | 0.0 | 0.0 | 0.0 | 0.0 | 0.0 | 0.0 |

| | | | | | | | | | | | | |
|---|---|---|---|---|---|---|---|---|---|---|---|---|
| | | 2 | 0 | 0.0 | 0.0 | 0.0 | 0.0 | 0.0 | 0.0 | 0.0 | 0.0 | 0.0 |
| Regional | 65-74 | 0 | 431 | 16.4 | 10.4 | 22.6 | 79.2 | 74.7 | 83.0 | 4.3 | 2.7 | 6.6 |
| | | 1 | 431 | 17.6 | 11.2 | 24.2 | 77.3 | 72.7 | 81.3 | 5.0 | 3.2 | 7.5 |
| | | 2 | 338 | 13.2 | 5.8 | 20.8 | 80.5 | 75.2 | 84.7 | 6.4 | 4.0 | 9.5 |
| | 75-84 | 0 | 285 | 6.1 | -0.3 | 12.8 | 88.8 | 84.2 | 92.1 | 5.2 | 2.9 | 8.3 |
| | | 1 | 432 | 10.3 | 4.1 | 16.7 | 83.3 | 79.2 | 86.7 | 6.4 | 4.2 | 9.2 |
| | | 2 | 513 | 8.9 | 3.2 | 14.8 | 83.5 | 79.8 | 86.6 | 7.5 | 5.4 | 10.2 |
| | 85-94 | 0 | 0 | 0.0 | 0.0 | 0.0 | 0.0 | 0.0 | 0.0 | 0.0 | 0.0 | 0.0 |
| | | 1 | 0 | 0.0 | 0.0 | 0.0 | 0.0 | 0.0 | 0.0 | 0.0 | 0.0 | 0.0 |
| | | 2 | 127 | 2.8 | -7.5 | 14.5 | 89.2 | 81.4 | 93.8 | 8.0 | 4.1 | 13.7 |
| Distant | 65-74 | 0 | 1669 | 2.8 | 0.7 | 5.0 | 93.7 | 92.3 | 94.8 | 3.5 | 2.7 | 4.5 |
| | | 1 | 1714 | 4.2 | 1.9 | 6.5 | 92.0 | 90.5 | 93.3 | 3.8 | 3.0 | 4.8 |
| | | 2 | 1064 | 3.6 | 0.9 | 6.4 | 93.1 | 91.3 | 94.6 | 3.3 | 2.3 | 4.5 |
| | 75-84 | 0 | 1074 | 1.4 | -1.0 | 3.9 | 95.2 | 93.6 | 96.3 | 3.4 | 2.4 | 4.6 |
| | | 1 | 1574 | 2.1 | -0.2 | 4.5 | 93.2 | 91.7 | 94.4 | 4.7 | 3.7 | 5.8 |
| | | 2 | 1729 | 1.5 | -0.9 | 4.1 | 92.1 | 90.7 | 93.3 | 6.4 | 5.3 | 7.6 |
| | 85-94 | 0 | 229 | 0.5 | -5.1 | 6.7 | 95.0 | 91.0 | 97.2 | 4.5 | 2.3 | 7.8 |
| | | 1 | 406 | 1.1 | -4.2 | 6.7 | 91.4 | 88.0 | 93.8 | 7.6 | 5.2 | 10.4 |
| | | 2 | 573 | 1.7 | -2.4 | 6.0 | 92.5 | 89.9 | 94.4 | 5.9 | 4.1 | 8.0 |

Table S14G. Number of patients and **one**-year probabilities of dying from cancer, dying from other-causes, and survival among **females** who were diagnosed with lung Adenocarcinoma **(ADC)** by stage, age and comorbidity level (n>100)

| Stage | Age | Comorbidity | N | Survival (%) | | | Cancer deaths (%) | | | Other-Cause Deaths (%) | | |
|---|---|---|---|---|---|---|---|---|---|---|---|---|
| | | | | Estimate | 95%CI. | | Estimate | 95%CI. | | Estimate | 95%CI. | |
| Localized | 65-74 | 0 | 429 | 98.1 | 95.5 | 99.3 | 1.4 | 0.6 | 2.9 | 0.5 | 0.1 | 1.6 |
| | | 1 | 720 | 97.8 | 95.9 | 98.8 | 1.9 | 1.1 | 3.2 | 0.3 | 0.1 | 1.0 |
| | | 2 | 381 | 94.7 | 91.0 | 97.1 | 4.5 | 2.7 | 6.9 | 0.8 | 0.2 | 2.2 |
| | 75-84 | 0 | 127 | 90.6 | 84.7 | 94.9 | 9.4 | 5.1 | 15.3 | 0.0 | NA | NA |
| | | 1 | 378 | 95.2 | 91.6 | 97.4 | 4.2 | 2.5 | 6.6 | 0.5 | 0.1 | 1.8 |
| | | 2 | 270 | 86.3 | 79.5 | 91.4 | 8.9 | 5.9 | 12.7 | 4.8 | 2.7 | 7.8 |
| | 85-94 | 0 | 0 | 0.0 | 0.0 | 0.0 | 0.0 | 0.0 | 0.0 | 0.0 | 0.0 | 0.0 |
| | | 1 | 0 | 0.0 | 0.0 | 0.0 | 0.0 | 0.0 | 0.0 | 0.0 | 0.0 | 0.0 |
| | | 2 | 0 | 0.0 | 0.0 | 0.0 | 0.0 | 0.0 | 0.0 | 0.0 | 0.0 | 0.0 |
| Regional | 65-74 | 0 | 327 | 80.7 | 74.7 | 85.3 | 18.4 | 14.4 | 22.8 | 0.9 | 0.3 | 2.5 |
| | | 1 | 503 | 80.3 | 75.4 | 84.3 | 18.3 | 15.1 | 21.8 | 1.4 | 0.6 | 2.7 |
| | | 2 | 265 | 72.1 | 63.9 | 79.0 | 23.8 | 18.8 | 29.1 | 4.2 | 2.2 | 7.1 |
| | 75-84 | 0 | 151 | 61.6 | 53.9 | 69.4 | 38.4 | 30.6 | 46.1 | 0.0 | NA | NA |
| | | 1 | 375 | 66.7 | 60.1 | 72.4 | 31.2 | 26.6 | 35.9 | 2.1 | 1.0 | 4.0 |
| | | 2 | 262 | 58.0 | 49.6 | 65.4 | 39.3 | 33.4 | 45.2 | 2.7 | 1.2 | 5.2 |
| | 85-94 | 0 | 0 | 0.0 | 0.0 | 0.0 | 0.0 | 0.0 | 0.0 | 0.0 | 0.0 | 0.0 |
| | | 1 | 0 | 0.0 | 0.0 | 0.0 | 0.0 | 0.0 | 0.0 | 0.0 | 0.0 | 0.0 |
| | | 2 | 107 | 38.3 | 24.7 | 50.8 | 57.9 | 48.0 | 66.7 | 3.7 | 1.2 | 8.6 |

| | | | | | | | | | | | | |
|---|---|---|---|---|---|---|---|---|---|---|---|---|
| Distant | 65-74 | 0 | 1489 | 59.0 | 55.8 | 62.1 | 39.4 | 36.9 | 41.8 | 1.6 | 1.1 | 2.4 |
| | | 1 | 1970 | 60.4 | 57.7 | 62.9 | 38.6 | 36.5 | 40.8 | 1.0 | 0.6 | 1.5 |
| | | 2 | 871 | 51.7 | 47.3 | 55.8 | 46.2 | 42.8 | 49.4 | 2.2 | 1.4 | 3.3 |
| | 75-84 | 0 | 779 | 43.5 | 38.8 | 47.9 | 54.2 | 50.7 | 57.6 | 2.3 | 1.4 | 3.6 |
| | | 1 | 1757 | 48.3 | 45.1 | 51.3 | 49.1 | 46.7 | 51.4 | 2.7 | 2.0 | 3.5 |
| | | 2 | 1099 | 39.3 | 35.1 | 43.4 | 56.4 | 53.4 | 59.3 | 4.3 | 3.2 | 5.6 |
| | 85-94 | 0 | 201 | 25.9 | 16.2 | 35.3 | 68.7 | 61.8 | 74.6 | 5.5 | 2.9 | 9.2 |
| | | 1 | 477 | 31.6 | 25.5 | 37.5 | 64.6 | 60.1 | 68.7 | 3.8 | 2.3 | 5.8 |
| | | 2 | 401 | 23.4 | 17.0 | 29.8 | 72.1 | 67.4 | 76.2 | 4.5 | 2.8 | 6.8 |

Table S14H: Number of patients and **two**-year probabilities of dying from cancer, dying from other-causes, and survival among **females** who were diagnosed with lung Adenocarcinoma **(ADC)** by stage, age and comorbidity level (n>100)

| Stage | Age | Comorbidity | N | Survival (%) | | | Cancer deaths (%) | | | Other-Cause Deaths (%) | | |
|---|---|---|---|---|---|---|---|---|---|---|---|---|
| | | | | Estimate | 95%CI. | | Estimate | 95%CI. | | Estimate | 95%CI. | |
| Localized | 65-74 | 0 | 429 | 93.6 | 89.8 | 96.0 | 6.0 | 3.9 | 8.6 | 0.5 | 0.1 | 1.6 |
| | | 1 | 720 | 95.2 | 92.7 | 97.0 | 4.1 | 2.8 | 5.7 | 0.7 | 0.3 | 1.6 |
| | | 2 | 381 | 89.6 | 84.6 | 93.3 | 8.2 | 5.7 | 11.4 | 2.1 | 1.0 | 4.0 |
| | 75-84 | 0 | 127 | 82.5 | 71.6 | 89.4 | 16.6 | 10.6 | 23.9 | 0.9 | 0.1 | 4.6 |
| | | 1 | 378 | 87.0 | 81.7 | 91.0 | 11.3 | 8.3 | 14.9 | 1.7 | 0.7 | 3.5 |
| | | 2 | 270 | 74.9 | 66.6 | 81.8 | 19.9 | 15.2 | 25.0 | 5.2 | 3.0 | 8.3 |
| | 85-94 | 0 | 0 | 0.0 | 0.0 | 0.0 | 0.0 | 0.0 | 0.0 | 0.0 | 0.0 | 0.0 |
| | | 1 | 0 | 0.0 | 0.0 | 0.0 | 0.0 | 0.0 | 0.0 | 0.0 | 0.0 | 0.0 |

| | | | | | | | | | | | | |
|---|---|---|---|---|---|---|---|---|---|---|---|---|
| | | 2 | 0 | 0.0 | 0.0 | 0.0 | 0.0 | 0.0 | 0.0 | 0.0 | 0.0 | 0.0 |
| Regional | 65-74 | 0 | 327 | 66.4 | 59.2 | 72.6 | 31.7 | 26.6 | 36.9 | 1.9 | 0.8 | 3.9 |
| | | 1 | 503 | 66.0 | 60.3 | 71.1 | 32.0 | 27.9 | 36.2 | 2.0 | 1.0 | 3.6 |
| | | 2 | 265 | 56.8 | 47.7 | 65.0 | 37.8 | 31.9 | 43.8 | 5.3 | 3.1 | 8.5 |
| | 75-84 | 0 | 151 | 39.8 | 29.0 | 49.7 | 58.2 | 49.7 | 65.6 | 2.0 | 0.5 | 5.4 |
| | | 1 | 375 | 43.7 | 36.5 | 50.5 | 52.8 | 47.6 | 57.8 | 3.5 | 2.0 | 5.7 |
| | | 2 | 262 | 34.6 | 25.9 | 42.9 | 61.1 | 54.8 | 66.9 | 4.3 | 2.3 | 7.2 |
| | 85-94 | 0 | 0 | 0.0 | 0.0 | 0.0 | 0.0 | 0.0 | 0.0 | 0.0 | 0.0 | 0.0 |
| | | 1 | 0 | 0.0 | 0.0 | 0.0 | 0.0 | 0.0 | 0.0 | 0.0 | 0.0 | 0.0 |
| | | 2 | 107 | 13.4 | 1.0 | 26.2 | 80.0 | 70.9 | 86.5 | 6.6 | 2.9 | 12.5 |
| Distant | 65-74 | 0 | 1489 | 32.7 | 29.4 | 35.9 | 64.8 | 62.3 | 67.3 | 2.5 | 1.8 | 3.4 |
| | | 1 | 1970 | 35.5 | 32.6 | 38.2 | 62.5 | 60.3 | 64.7 | 2.0 | 1.5 | 2.7 |
| | | 2 | 871 | 27.1 | 22.8 | 31.3 | 69.7 | 66.5 | 72.6 | 3.2 | 2.2 | 4.6 |
| | 75-84 | 0 | 779 | 20.8 | 16.5 | 25.1 | 76.0 | 72.8 | 78.9 | 3.1 | 2.1 | 4.5 |
| | | 1 | 1757 | 24.4 | 21.3 | 27.5 | 71.5 | 69.3 | 73.6 | 4.1 | 3.3 | 5.1 |
| | | 2 | 1099 | 20.0 | 16.1 | 23.9 | 75.0 | 72.3 | 77.4 | 5.0 | 3.9 | 6.5 |
| | 85-94 | 0 | 201 | 9.8 | 1.4 | 18.6 | 83.7 | 77.8 | 88.2 | 6.5 | 3.6 | 10.5 |
| | | 1 | 477 | 11.5 | 6.1 | 17.0 | 83.2 | 79.5 | 86.3 | 5.3 | 3.5 | 7.6 |
| | | 2 | 401 | 10.1 | 4.3 | 16.0 | 84.4 | 80.4 | 87.6 | 5.5 | 3.6 | 8.1 |

Table S14I. Number of patients and **five**-year probabilities of dying from cancer, dying from other-causes, and survival among **females** who were diagnosed with lung Adenocarcinoma **(ADC)** by stage, age and comorbidity level (n>100)

| Stage | Age | Comorbidity | N | Survival (%) | | | Cancer deaths (%) | | | Other-Cause Deaths (%) | | |
|---|---|---|---|---|---|---|---|---|---|---|---|---|
| | | | | Estimate | 95%CI. | | Estimate | 95%CI. | | Estimate | 95%CI. | |
| Localized | 65-74 | 0 | 429 | 78.2 | 71.5 | 83.7 | 19.9 | 15.6 | 24.6 | 1.9 | 0.8 | 3.9 |
| | | 1 | 720 | 79.4 | 74.0 | 83.9 | 18.2 | 14.7 | 21.9 | 2.4 | 1.3 | 4.1 |
| | | 2 | 381 | 72.0 | 63.8 | 79.1 | 22.5 | 17.8 | 27.7 | 5.4 | 3.2 | 8.5 |
| | 75-84 | 0 | 127 | 55.9 | 38.5 | 70.3 | 36.4 | 26.6 | 46.3 | 7.7 | 3.1 | 15.2 |
| | | 1 | 378 | 64.6 | 55.9 | 72.2 | 31.1 | 25.5 | 36.8 | 4.3 | 2.3 | 7.3 |
| | | 2 | 270 | 49.6 | 38.1 | 60.3 | 38.3 | 31.6 | 45.0 | 12.0 | 8.1 | 16.8 |
| | 85-94 | 0 | 0 | 0.0 | 0.0 | 0.0 | 0.0 | 0.0 | 0.0 | 0.0 | 0.0 | 0.0 |
| | | 1 | 0 | 0.0 | 0.0 | 0.0 | 0.0 | 0.0 | 0.0 | 0.0 | 0.0 | 0.0 |
| | | 2 | 0 | 0.0 | 0.0 | 0.0 | 0.0 | 0.0 | 0.0 | 0.0 | 0.0 | 0.0 |
| Regional | 65-74 | 0 | 327 | 34.6 | 26.8 | 42.1 | 63.1 | 56.9 | 68.7 | 2.3 | 1.0 | 4.5 |
| | | 1 | 503 | 26.1 | 19.4 | 32.7 | 69.2 | 64.3 | 73.5 | 4.7 | 3.0 | 7.1 |
| | | 2 | 265 | 22.6 | 13.0 | 32.4 | 70.5 | 63.5 | 76.4 | 6.9 | 4.2 | 10.6 |
| | 75-84 | 0 | 151 | 17.9 | 8.1 | 27.9 | 79.3 | 71.2 | 85.3 | 2.8 | 0.9 | 6.6 |
| | | 1 | 375 | 16.7 | 9.2 | 24.4 | 76.2 | 71.0 | 80.6 | 7.1 | 4.6 | 10.3 |
| | | 2 | 262 | 11.5 | 2.7 | 20.7 | 81.9 | 75.5 | 86.7 | 6.6 | 3.8 | 10.6 |
| | 85-94 | 0 | 0 | 0.0 | 0.0 | 0.0 | 0.0 | 0.0 | 0.0 | 0.0 | 0.0 | 0.0 |
| | | 1 | 0 | 0.0 | 0.0 | 0.0 | 0.0 | 0.0 | 0.0 | 0.0 | 0.0 | 0.0 |
| | | 2 | 107 | 1.7 | -9.1 | 14.8 | 90.7 | 81.7 | 95.4 | 7.6 | 3.5 | 13.7 |

| | | | | | | | | | | | | |
|---|---|---|---|---|---|---|---|---|---|---|---|---|
| Distant | 65-74 | 0 | 1489 | 7.2 | 4.5 | 9.9 | 89.7 | 87.8 | 91.3 | 3.1 | 2.3 | 4.1 |
| | | 1 | 1970 | 7.3 | 5.0 | 9.7 | 89.7 | 88.0 | 91.1 | 3.0 | 2.3 | 3.9 |
| | | 2 | 871 | 6.3 | 2.6 | 10.1 | 89.1 | 86.5 | 91.1 | 4.7 | 3.3 | 6.3 |
| | 75-84 | 0 | 779 | 3.3 | -0.1 | 6.9 | 92.6 | 90.3 | 94.4 | 4.1 | 2.8 | 5.7 |
| | | 1 | 1757 | 4.1 | 1.5 | 6.7 | 90.5 | 88.9 | 91.9 | 5.4 | 4.4 | 6.6 |
| | | 2 | 1099 | 3.0 | -0.2 | 6.3 | 91.0 | 89.0 | 92.7 | 6.0 | 4.7 | 7.6 |
| | 85-94 | 0 | 201 | 2.9 | -4.8 | 11.2 | 90.1 | 84.8 | 93.7 | 7.0 | 4.0 | 11.1 |
| | | 1 | 477 | 1.1 | -3.7 | 6.2 | 92.2 | 89.2 | 94.4 | 6.7 | 4.6 | 9.3 |
| | | 2 | 401 | 1.5 | -3.9 | 7.2 | 91.5 | 88.2 | 94.0 | 7.0 | 4.7 | 9.9 |

Figure S1A. Prevalence of cancer survivors by calendar year and years from diagnosis: **All cancer**

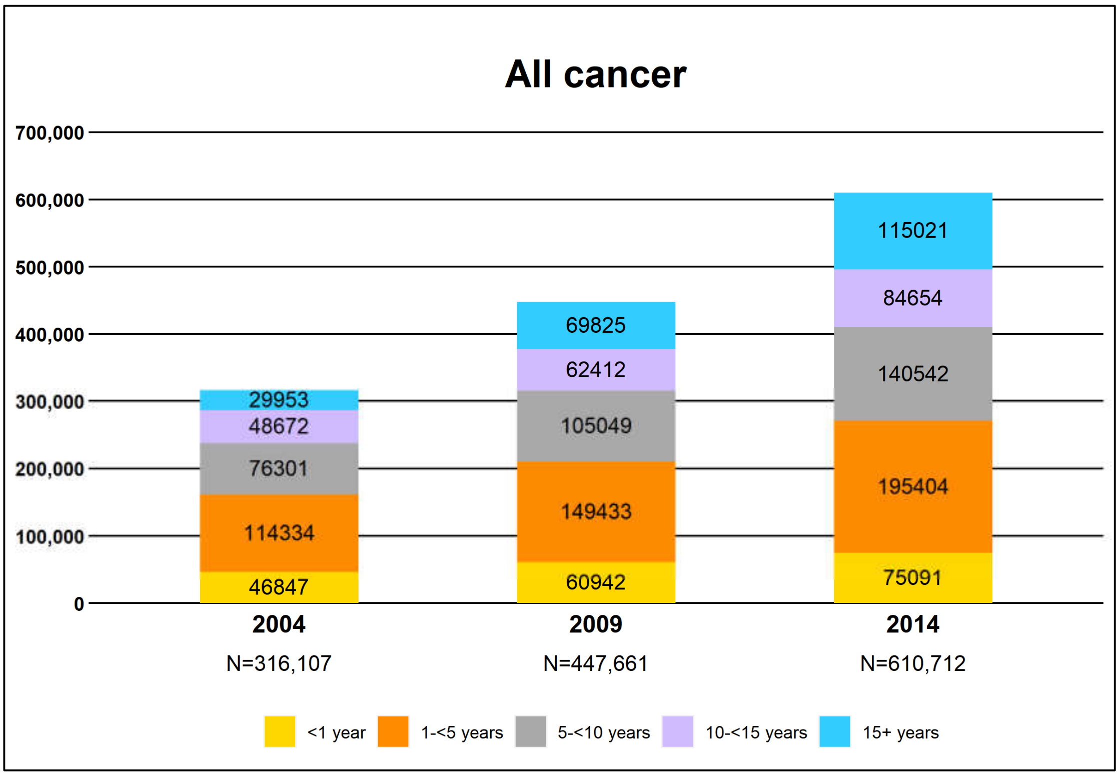

Figure S1B. Prevalence of cancer survivors by calendar year and years from diagnosis: **Breast Cancer**

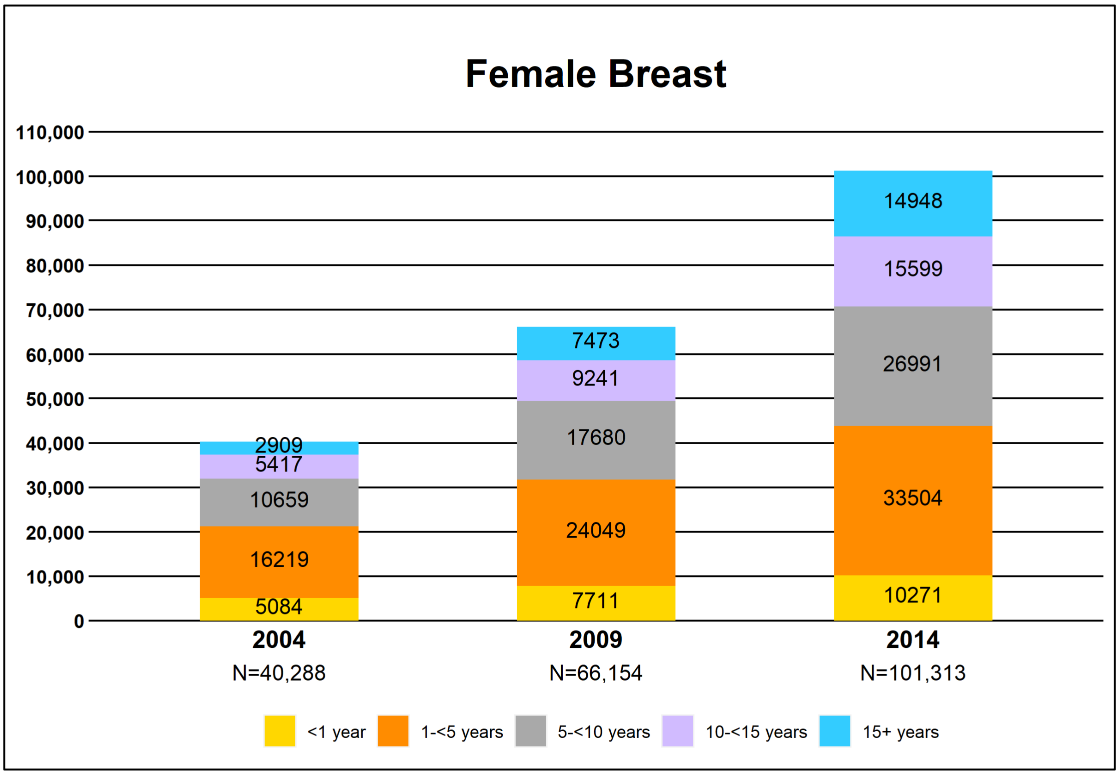

Figure S1C. Prevalence of cancer survivors by calendar year and years from diagnosis: **Colorectal Cancer**

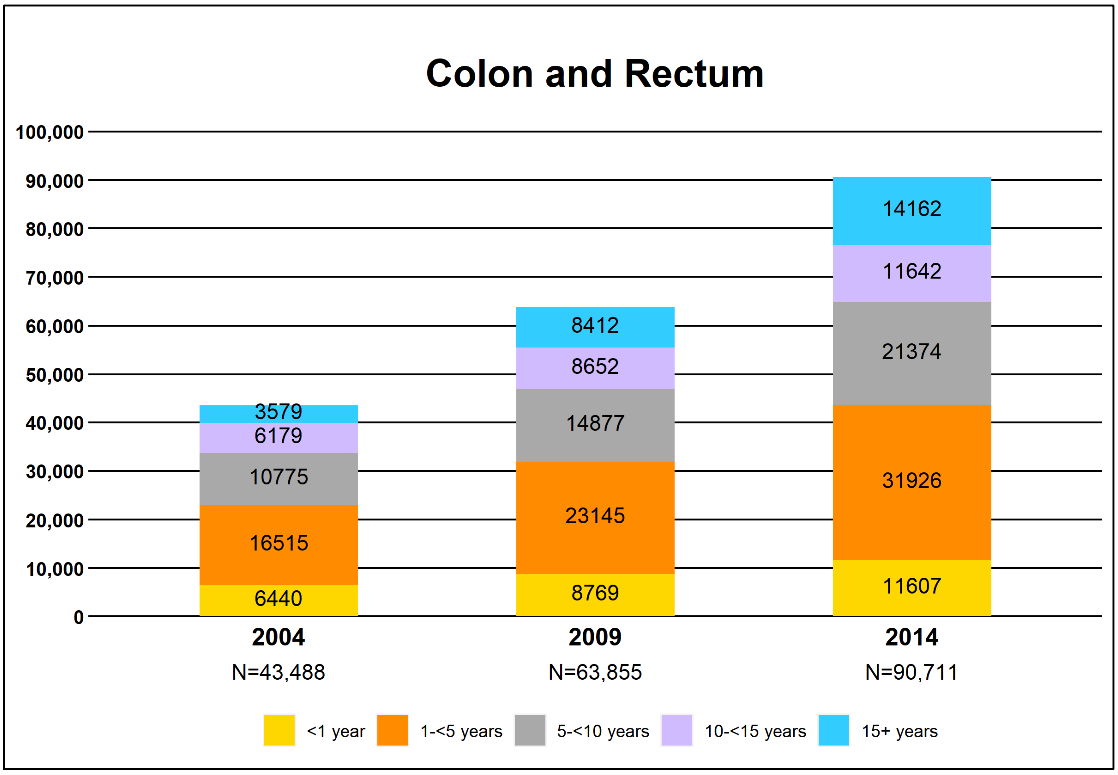

Figure S1D. Prevalence of cancer survivors by calendar year and years from diagnosis: **Liver Cancer**

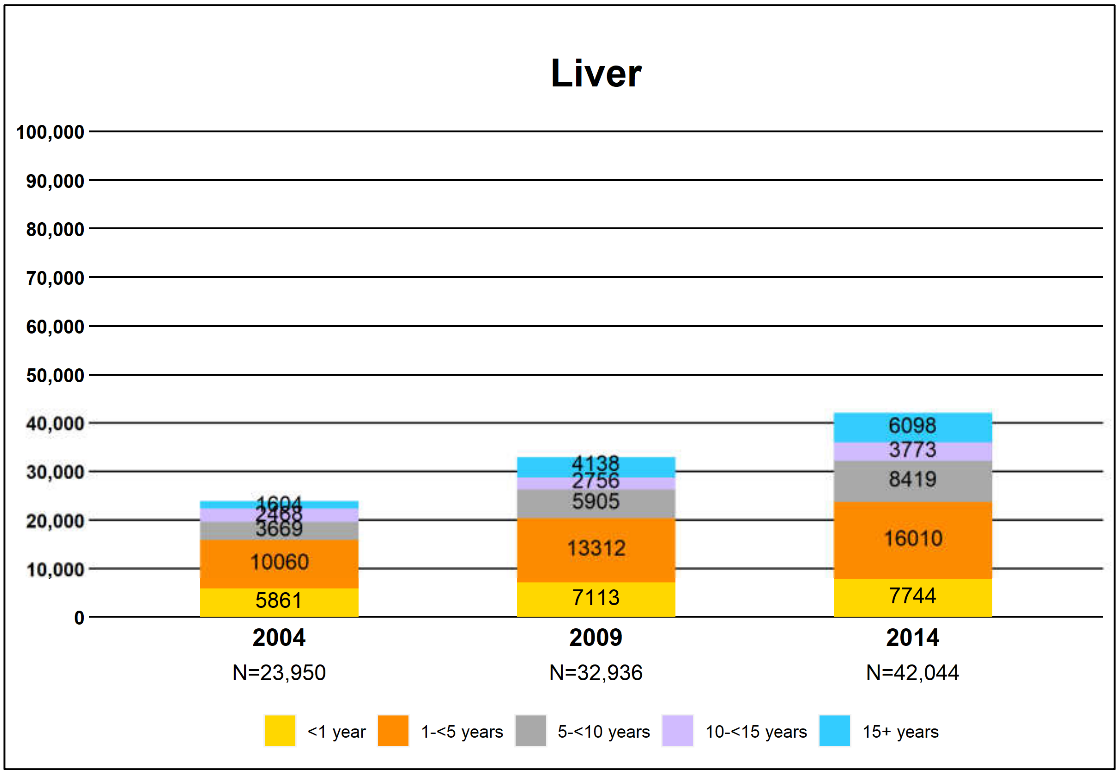

Figure S1E. Prevalence of cancer survivors by calendar year and years from diagnosis: **Lung Cancer**

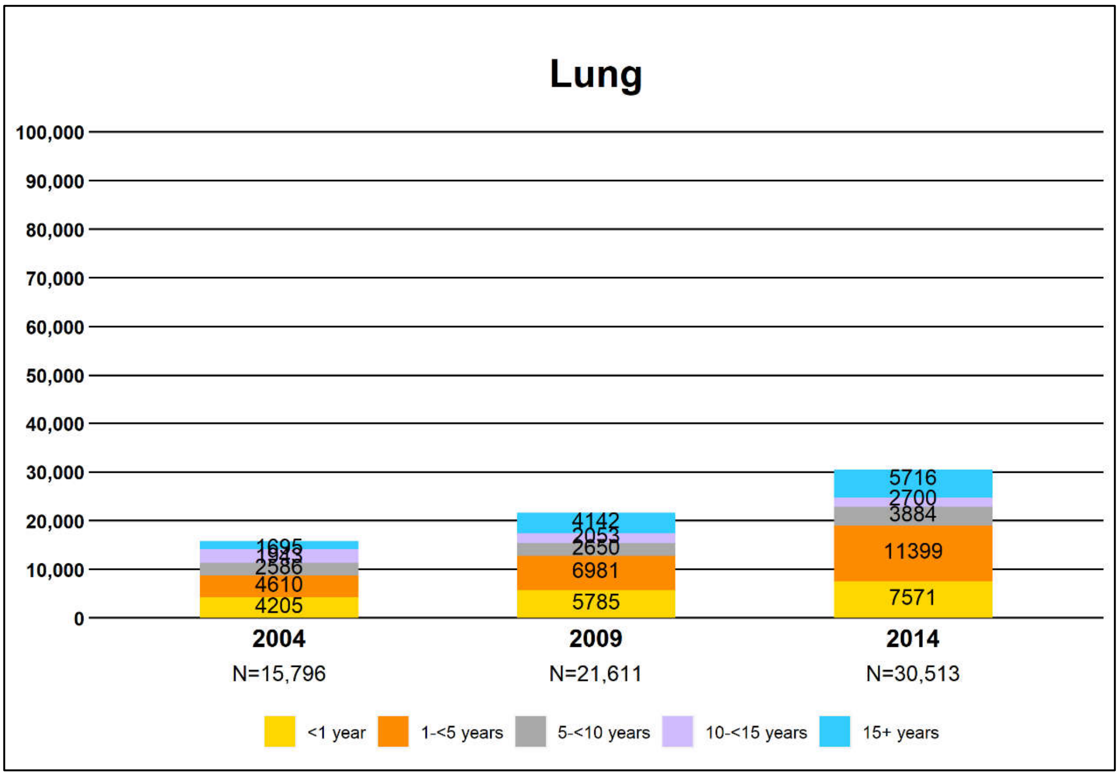

Figure S1F. Prevalence of cancer survivors by calendar year and years from diagnosis: **Oral Cancer**

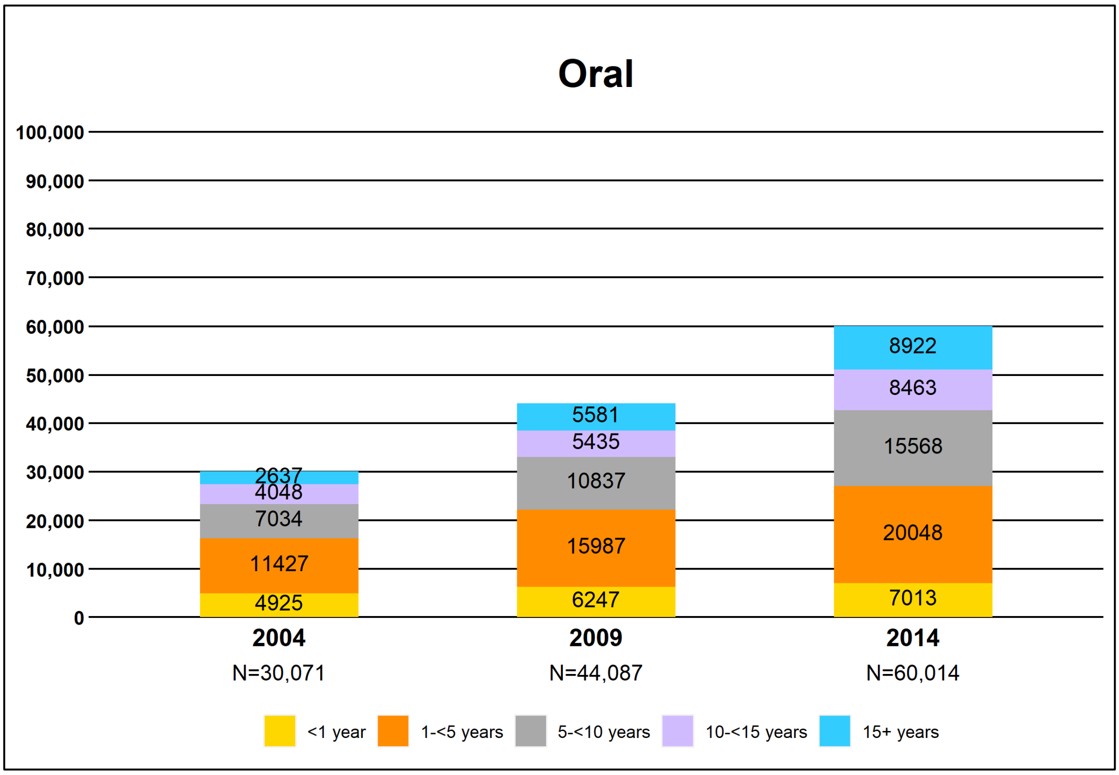

Figure S2A. Probabilities of dying from cancer, dying from other causes, and survival stratified by stage, comorbidity level, and age for **breast cancer**

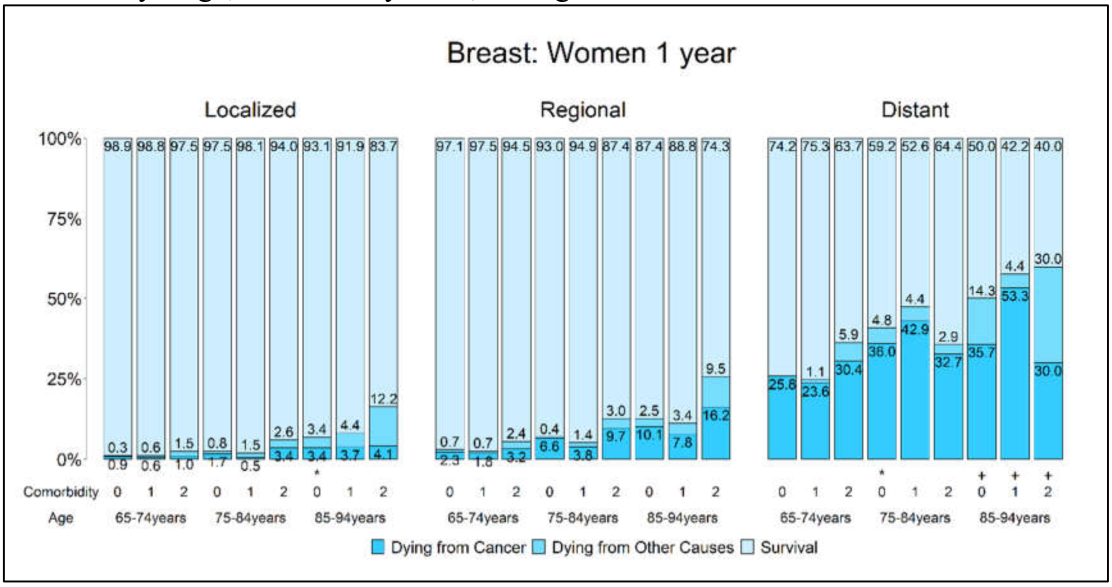

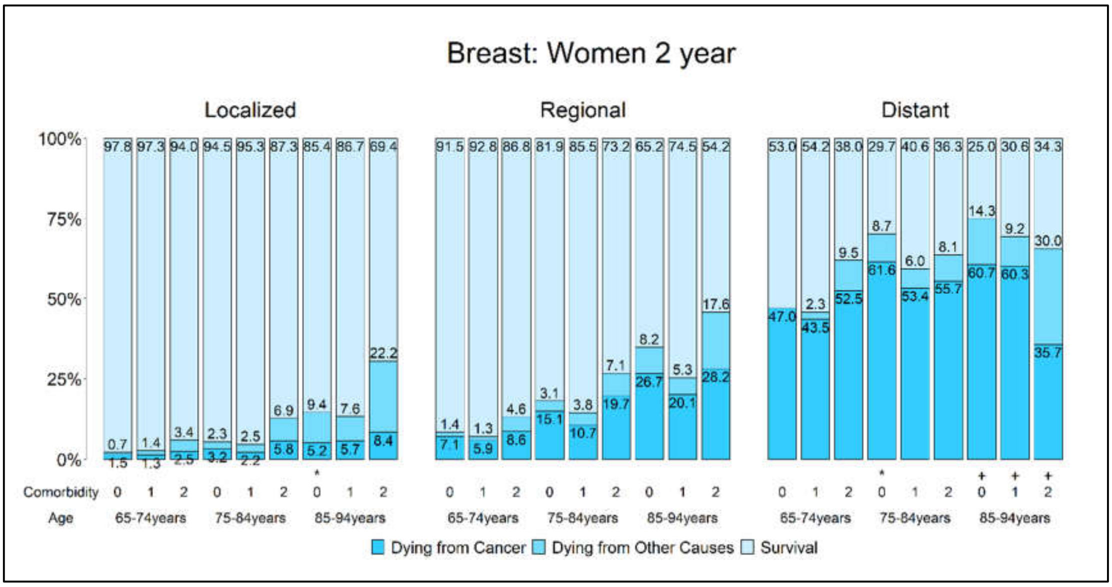

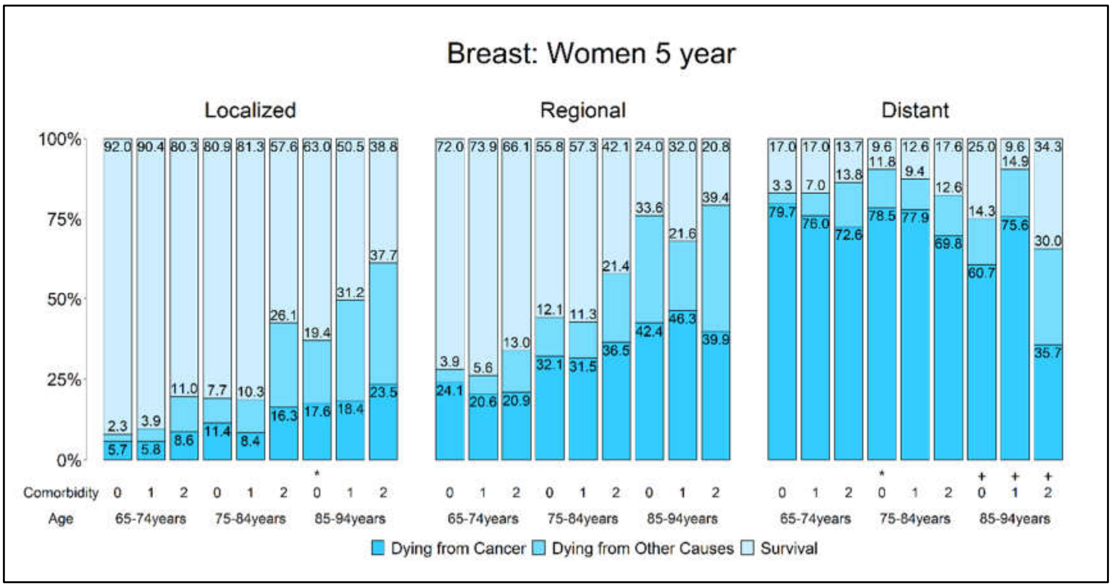

Figure S2B. Probabilities of dying from cancer, dying from other causes, and survival stratified by stage, comorbidity level, and age for **colorectal cancer**

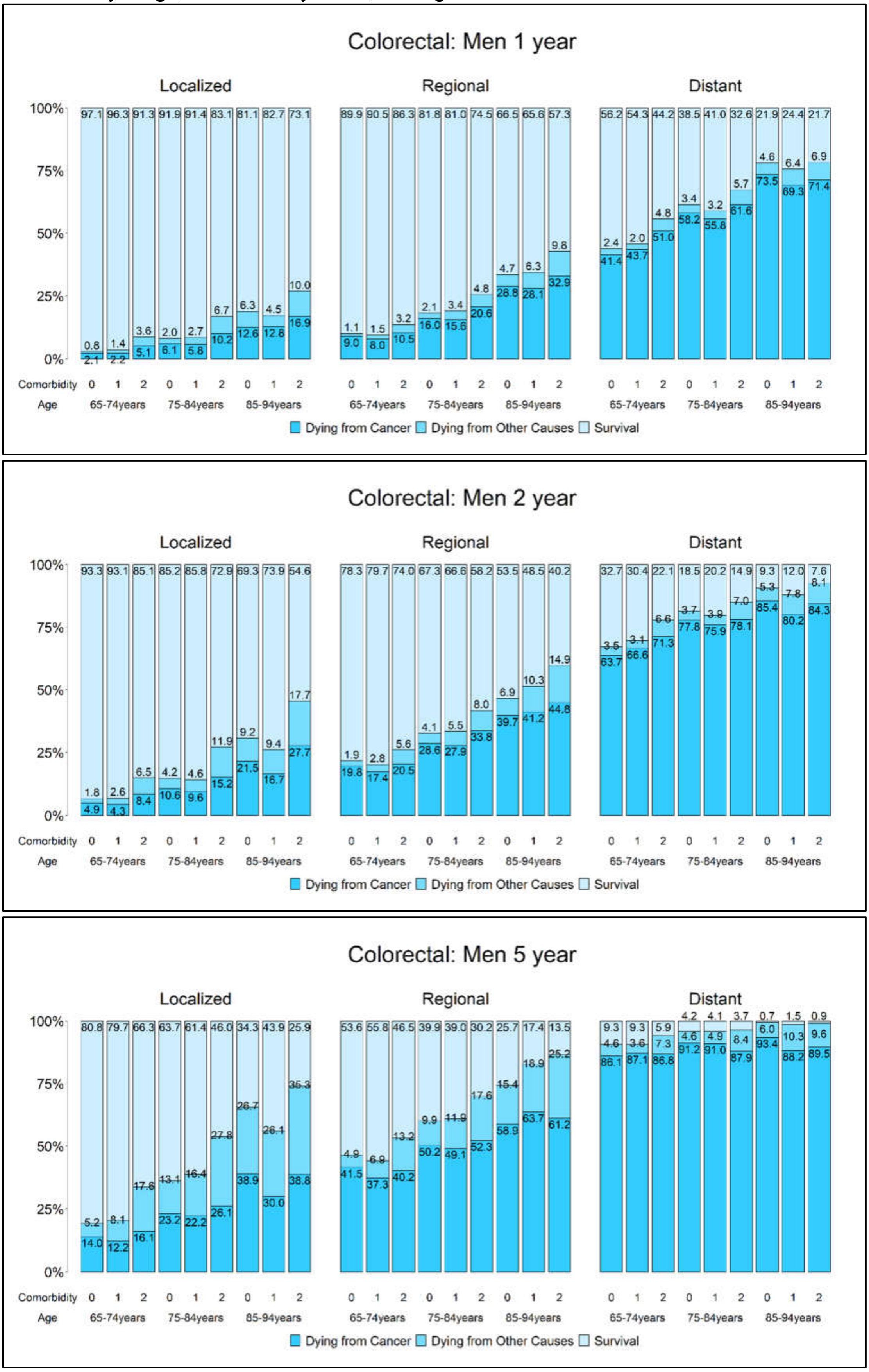

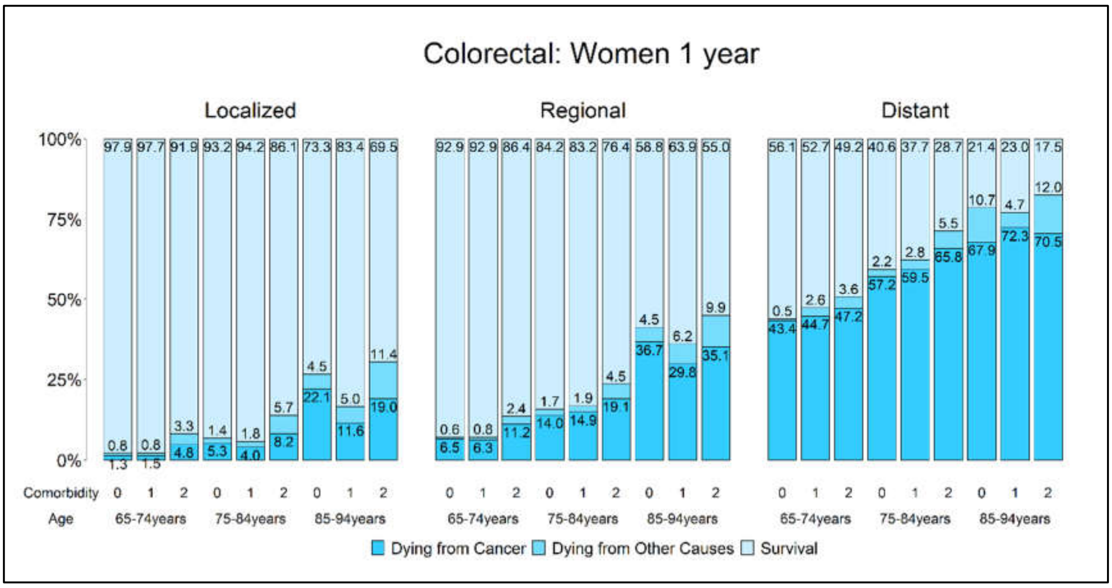
Colorectal: Women 1 year
Localized
Regional
Distant
100%
75%
50%
25%
0%
Comorbidity 0 1 2 0 1 2 0 1 2
Age 65-74years 75-84years 85-94years
Dying from Cancer
Dying from Other Causes
Survival

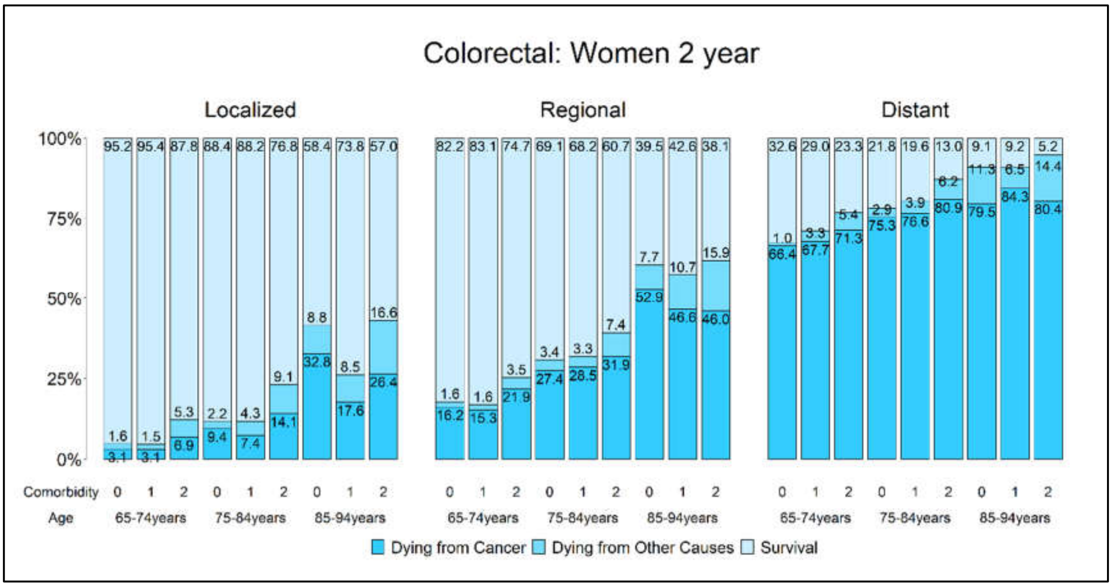
Colorectal: Women 2 year
Localized
Regional
Distant
100%
75%
50%
25%
0%
Comorbidity 0 1 2 0 1 2 0 1 2
Age 65-74years 75-84years 85-94years
Dying from Cancer
Dying from Other Causes
Survival

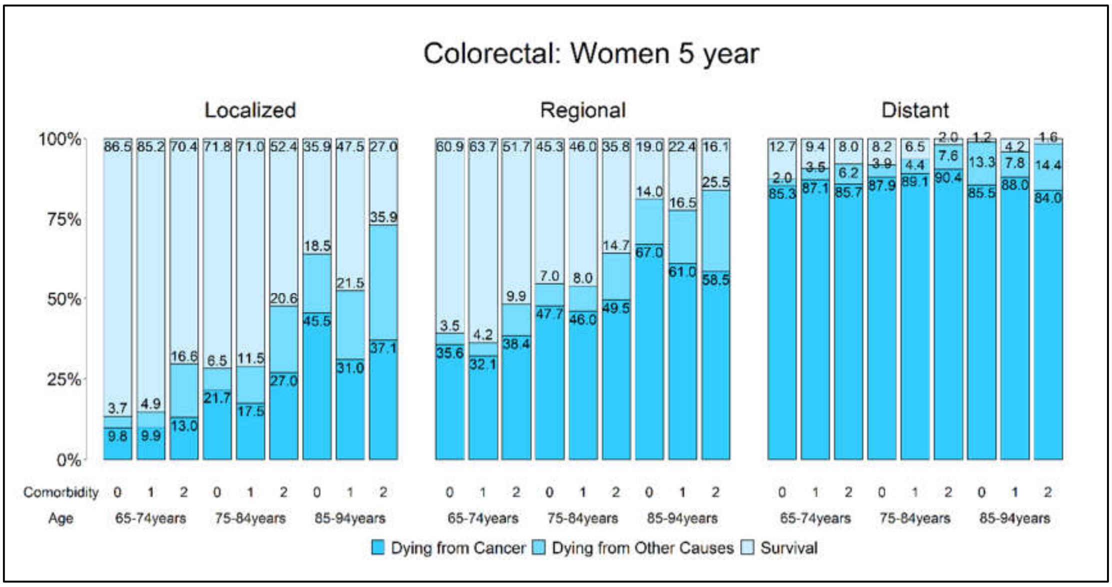
Colorectal: Women 5 year
Localized
Regional
Distant
100%
75%
50%
25%
0%
Comorbidity 0 1 2 0 1 2 0 1 2
Age 65-74years 75-84years 85-94years
Dying from Cancer
Dying from Other Causes
Survival

Figure S2C. Probabilities of dying from cancer, dying from other causes, and survival stratified by stage, comorbidity level, and age for **liver cancer**

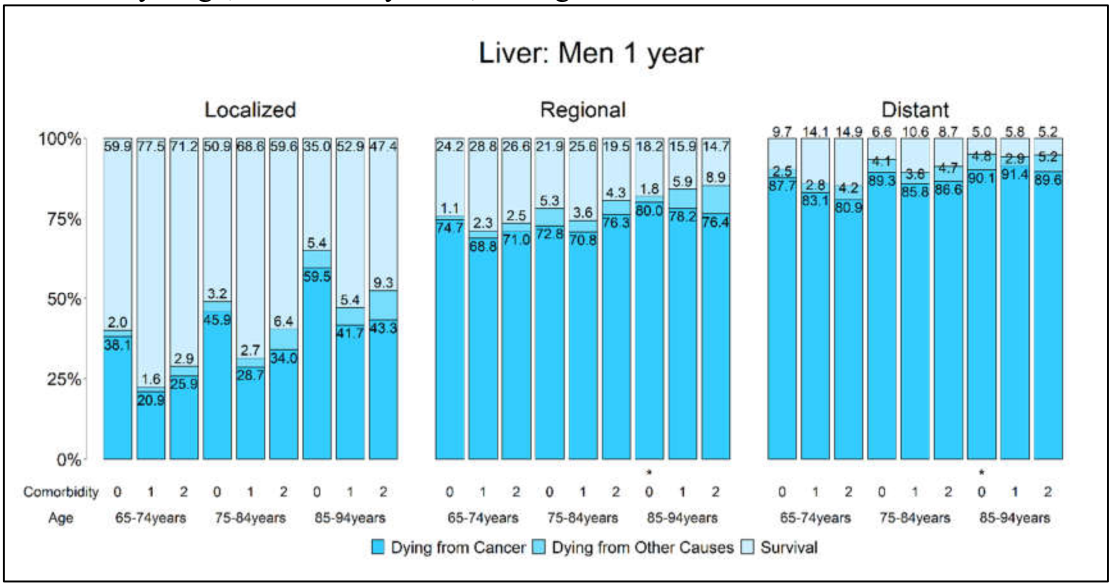

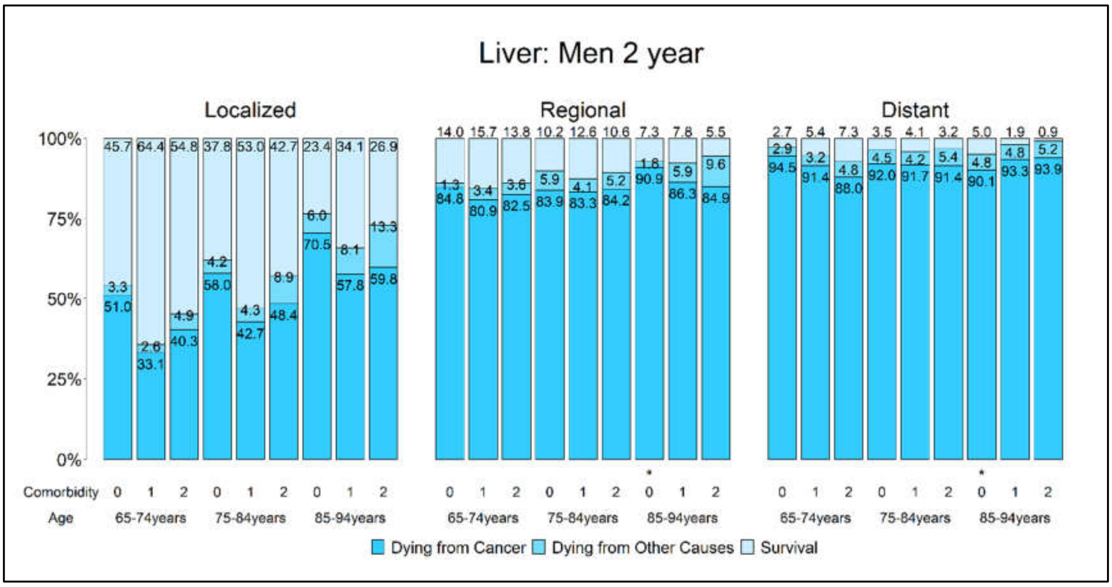

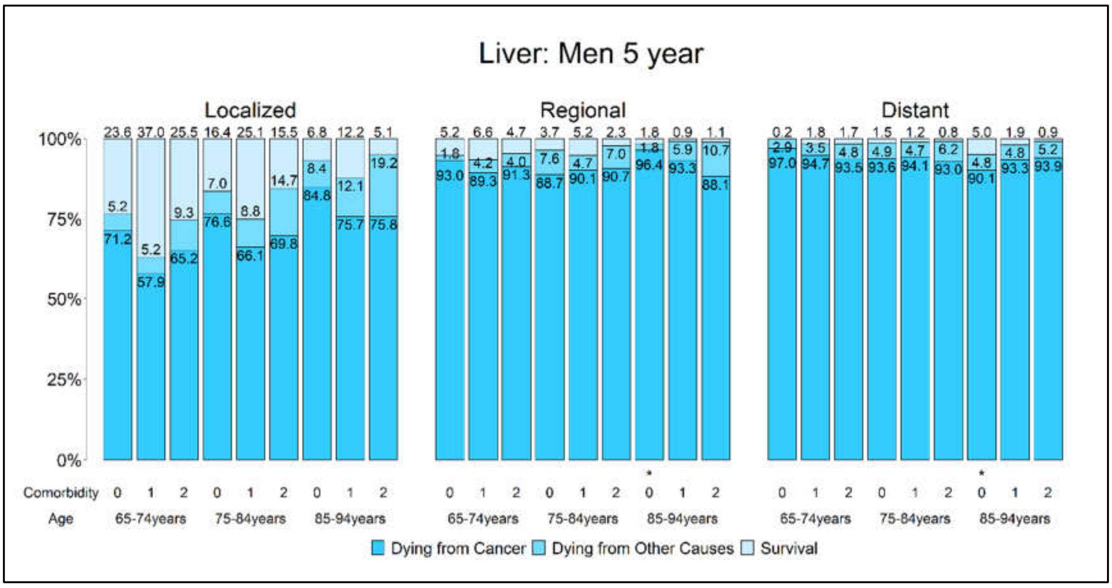

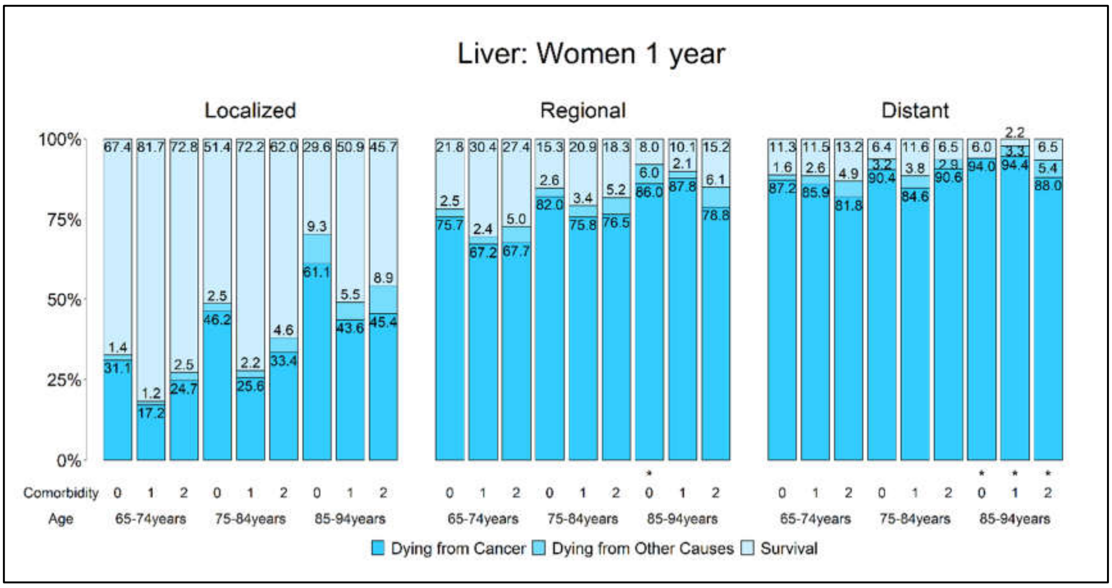
Liver: Women 1 year
Localized
Regional
Distant
100%
75%
50%
25%
0%
Comorbidity
Age
65-74years
75-84years
85-94years
Dying from Cancer
Dying from Other Causes
Survival

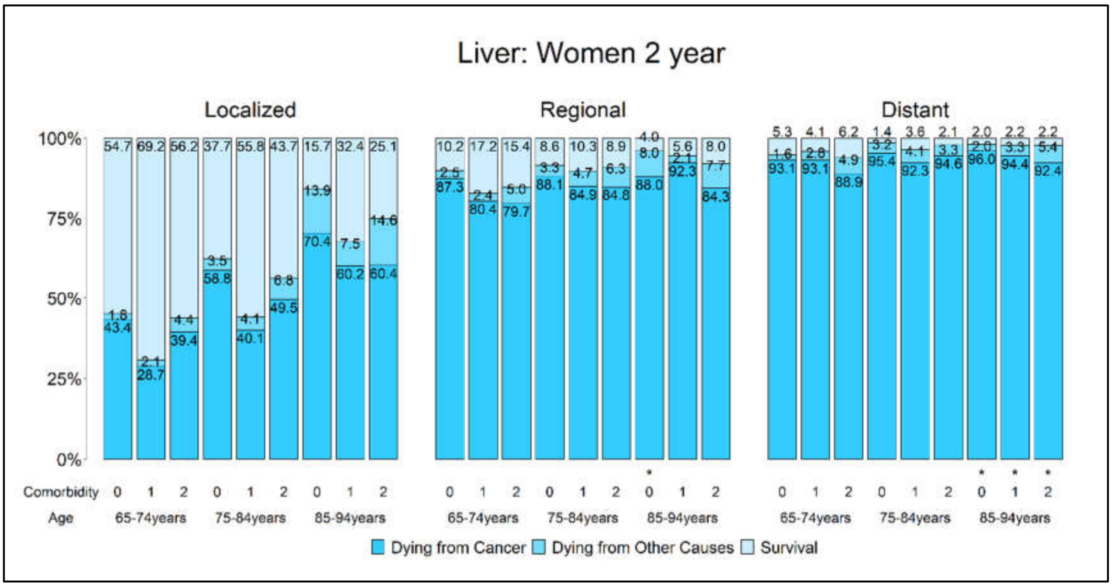
Liver: Women 2 year
Localized
Regional
Distant
100%
75%
50%
25%
0%
Comorbidity
Age
65-74years
75-84years
85-94years
Dying from Cancer
Dying from Other Causes
Survival

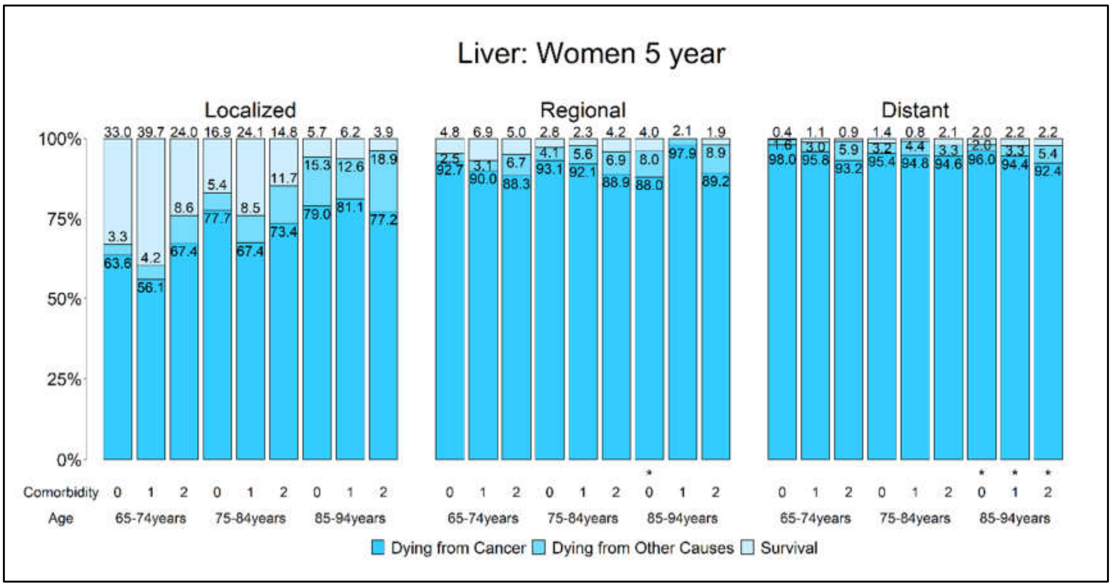
Liver: Women 5 year
Localized
Regional
Distant
100%
75%
50%
25%
0%
Comorbidity
Age
65-74years
75-84years
85-94years
Dying from Cancer
Dying from Other Causes
Survival

Figure S2D. Probabilities of dying from cancer, dying from other causes, and survival stratified by stage, comorbidity level, and age for **lung cancer**

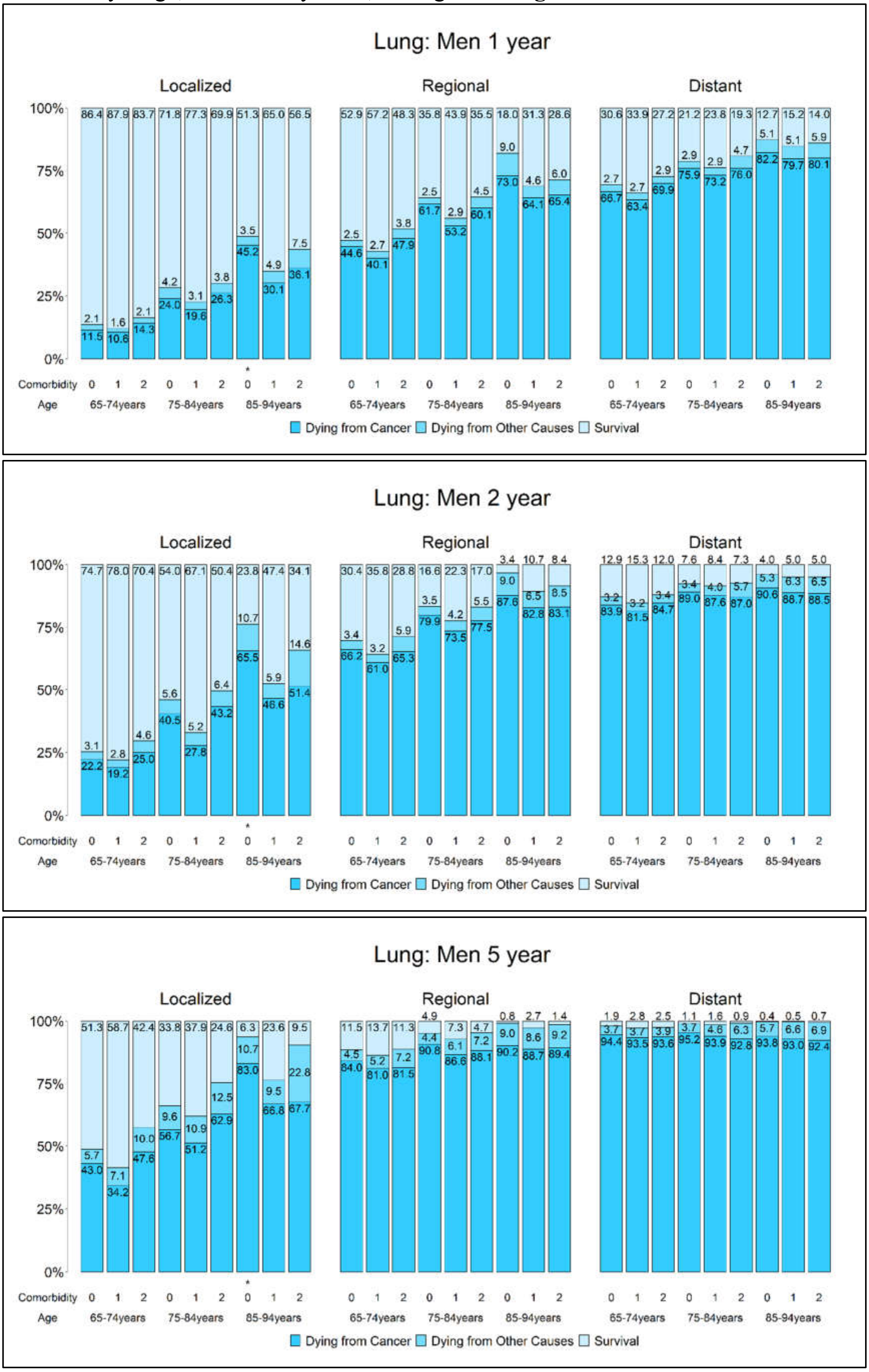

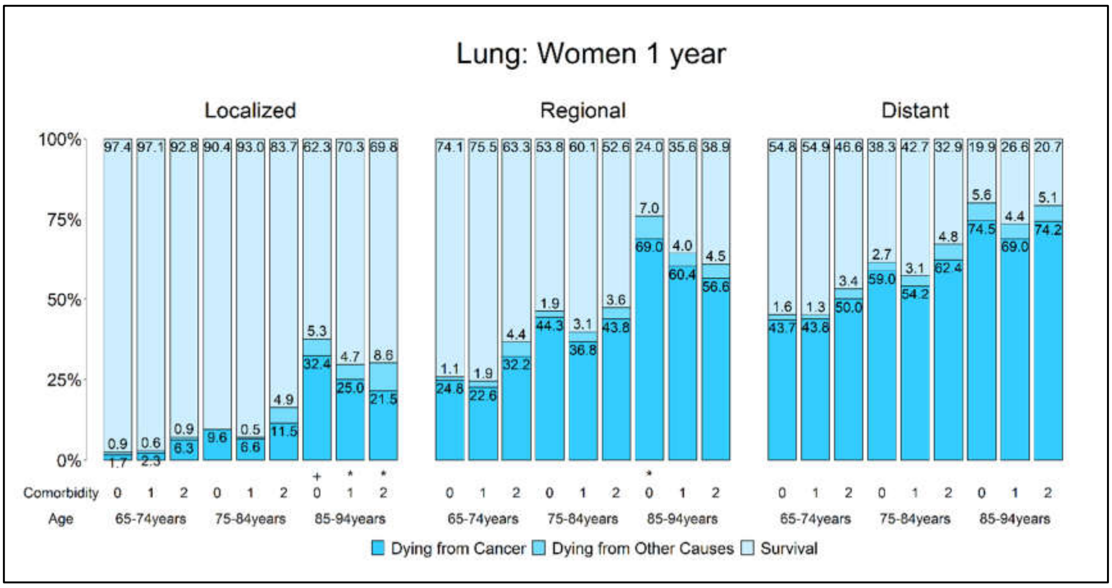
Lung: Women 1 year
Localized
Regional
Distant
100%
75%
50%
25%
0%
Comorbidity
Age
65-74years
75-84years
85-94years
Dying from Cancer
Dying from Other Causes
Survival

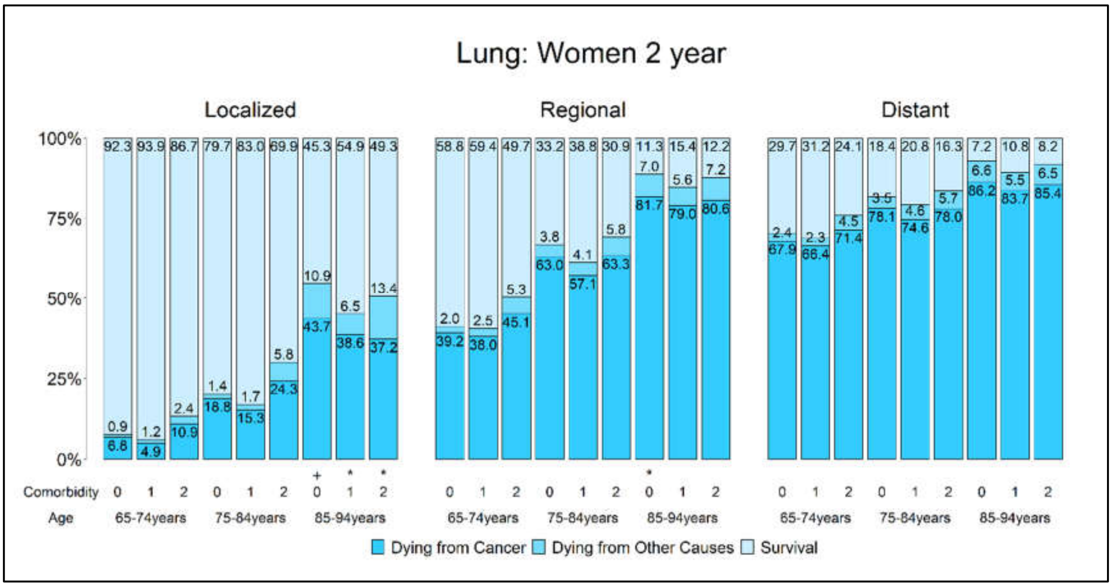
Lung: Women 2 year
Localized
Regional
Distant
100%
75%
50%
25%
0%
Comorbidity
Age
65-74years
75-84years
85-94years
Dying from Cancer
Dying from Other Causes
Survival

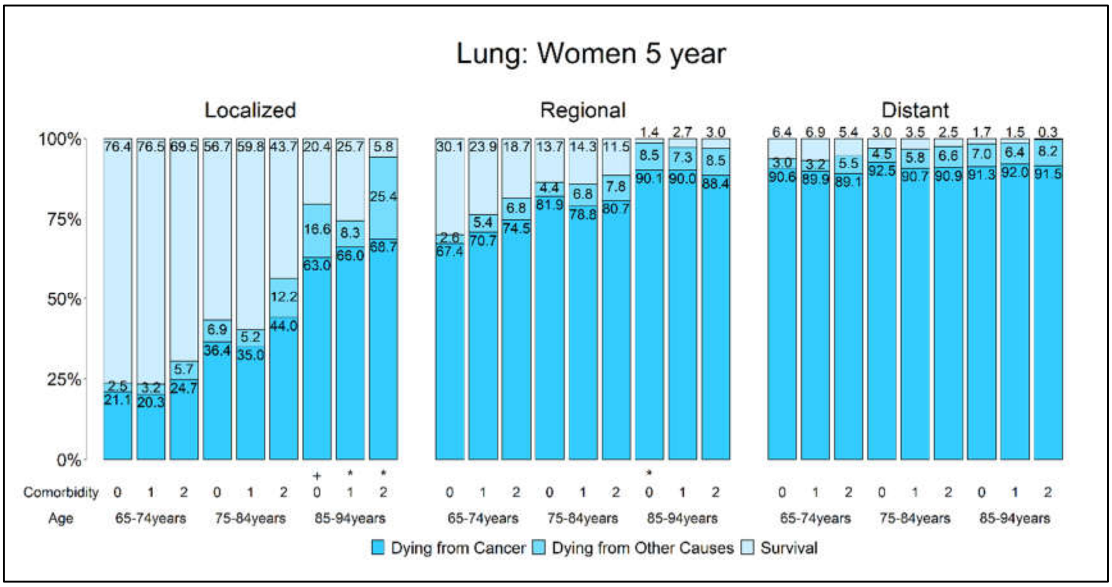
Lung: Women 5 year
Localized
Regional
Distant
100%
75%
50%
25%
0%
Comorbidity
Age
65-74years
75-84years
85-94years
Dying from Cancer
Dying from Other Causes
Survival

Figure S2E. Probabilities of dying from cancer, dying from other causes, and survival stratified by stage, comorbidity level, and age for **oral cancer**

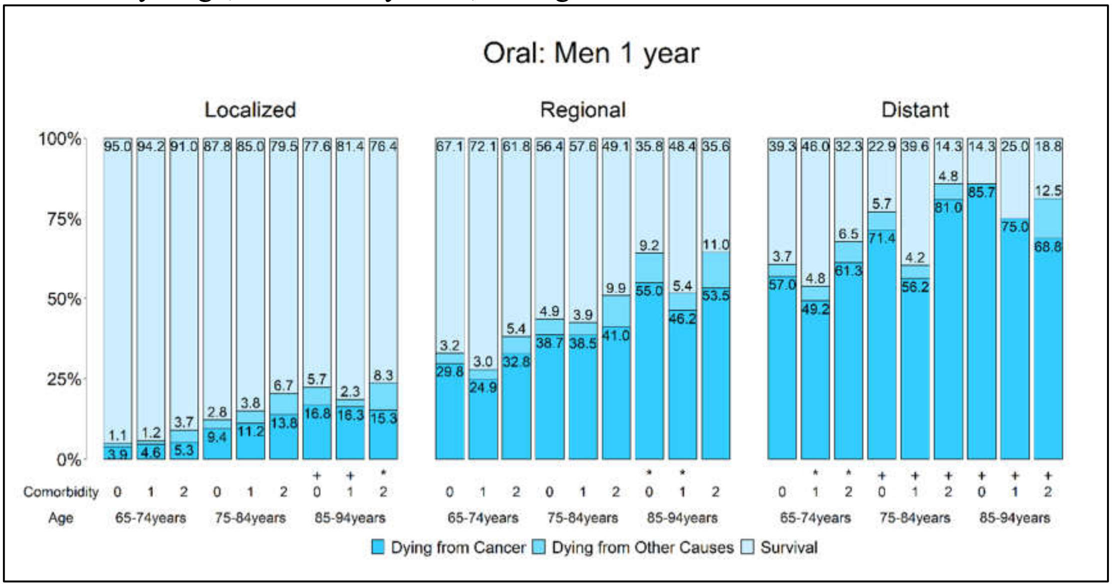

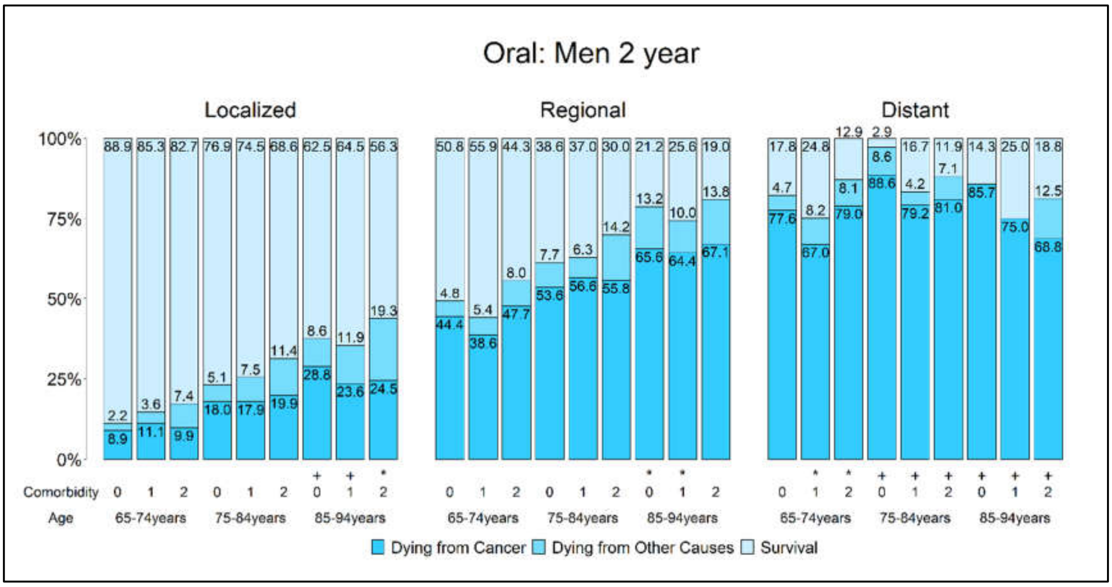

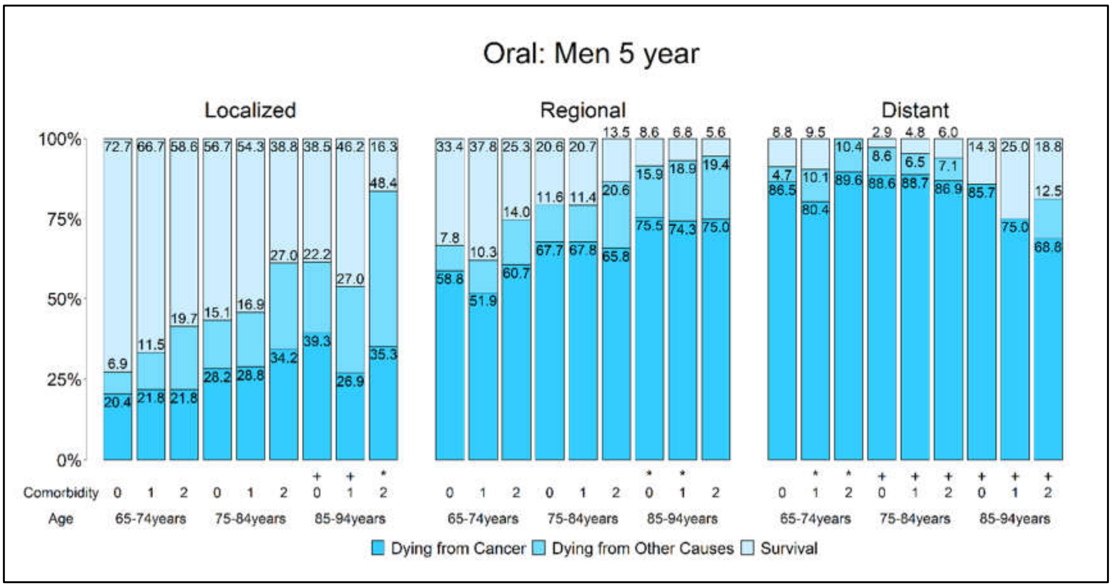

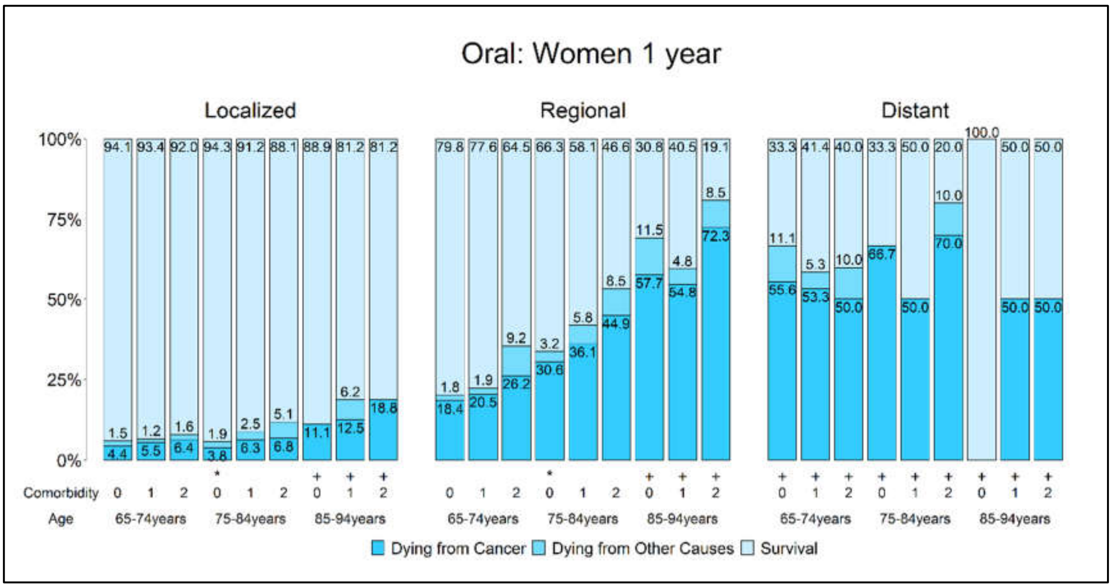
Oral: Women 1 year
Localized
Regional
Distant
100%
75%
50%
25%
0%
Comorbidity
Age
65-74years
75-84years
85-94years
Dying from Cancer
Dying from Other Causes
Survival

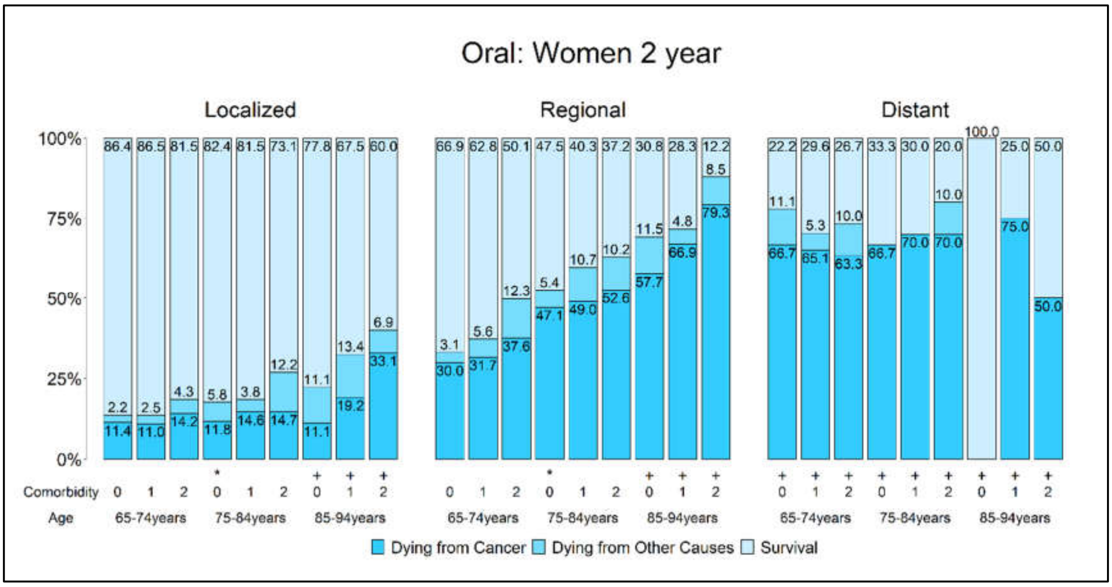
Oral: Women 2 year
Localized
Regional
Distant
100%
75%
50%
25%
0%
Comorbidity
Age
65-74years
75-84years
85-94years
Dying from Cancer
Dying from Other Causes
Survival

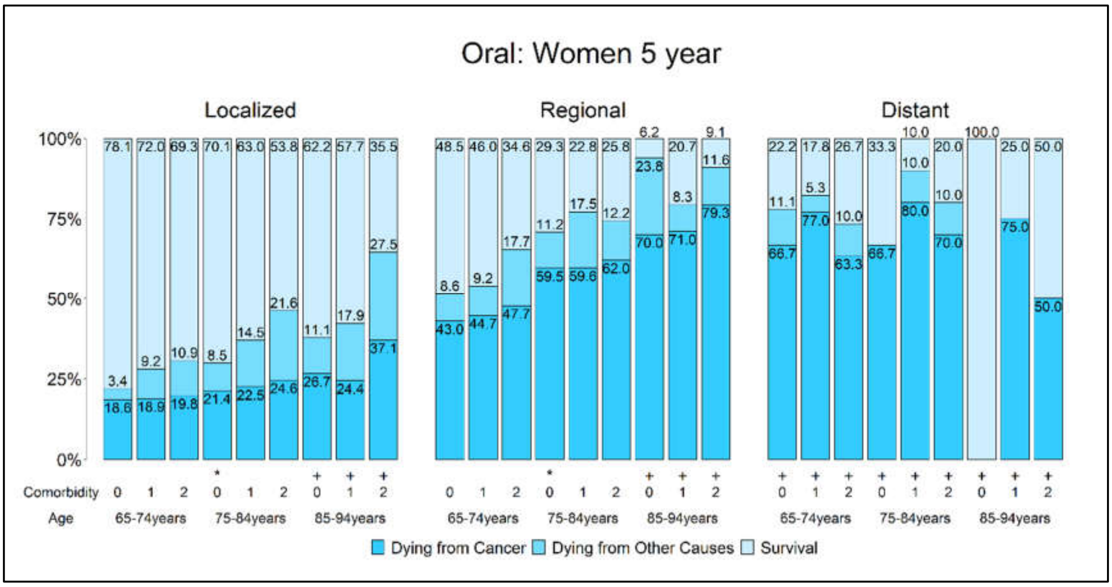
Oral: Women 5 year
Localized
Regional
Distant
100%
75%
50%
25%
0%
Comorbidity
Age
65-74years
75-84years
85-94years
Dying from Cancer
Dying from Other Causes
Survival

Figure S3A. One-year probabilities of dying from cancer, dying from other causes, and survival stratified by comorbidity level and subtype for distant **lung cancer** patients ages 30—94.

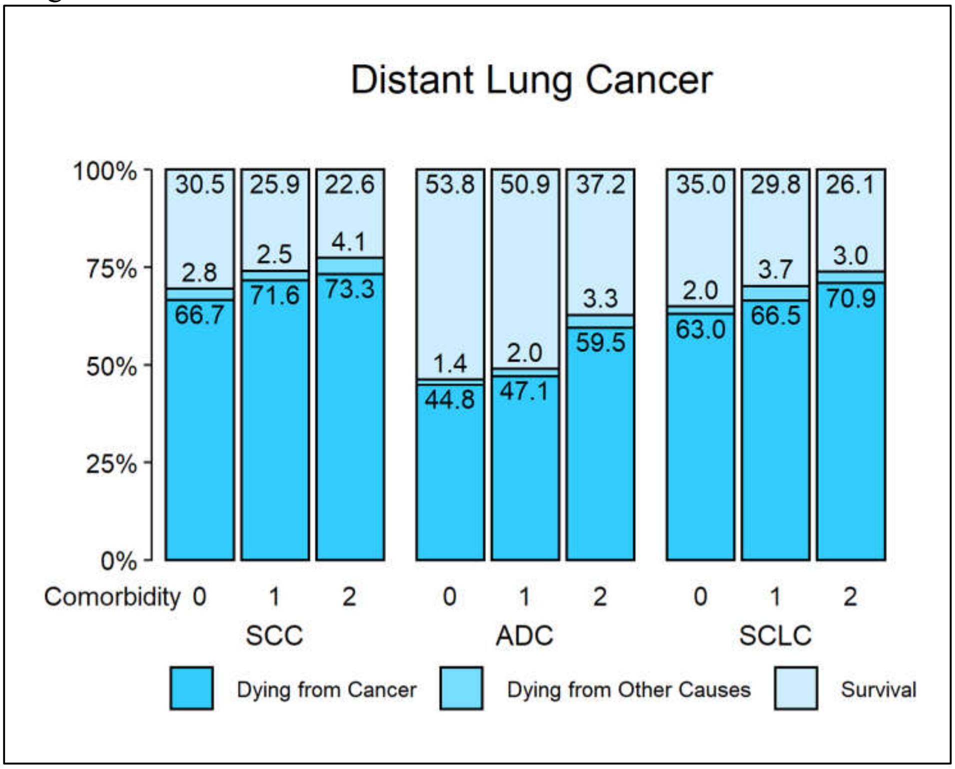

Figure S3B. Two-year probabilities of dying from cancer, dying from other causes, and survival stratified by comorbidity level and subtype for distant **lung cancer** patients ages 30—94.

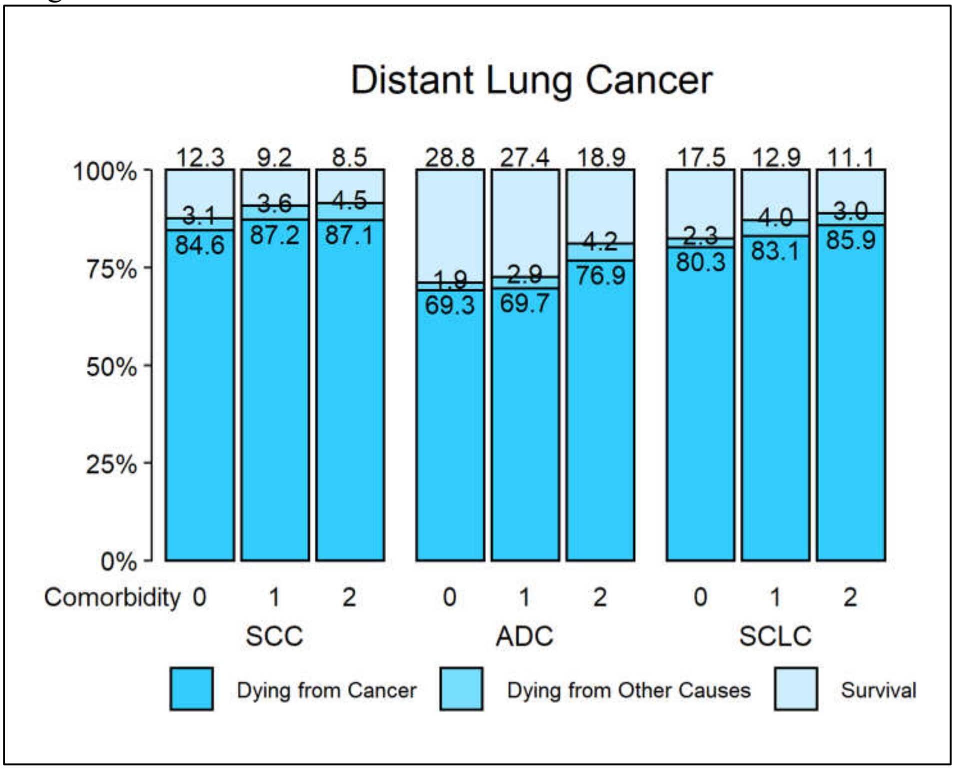

Figure S3C. Five-year probabilities of dying from cancer, dying from other causes, and survival stratified by comorbidity level and subtype for distant **lung cancer** patients ages 30—94.

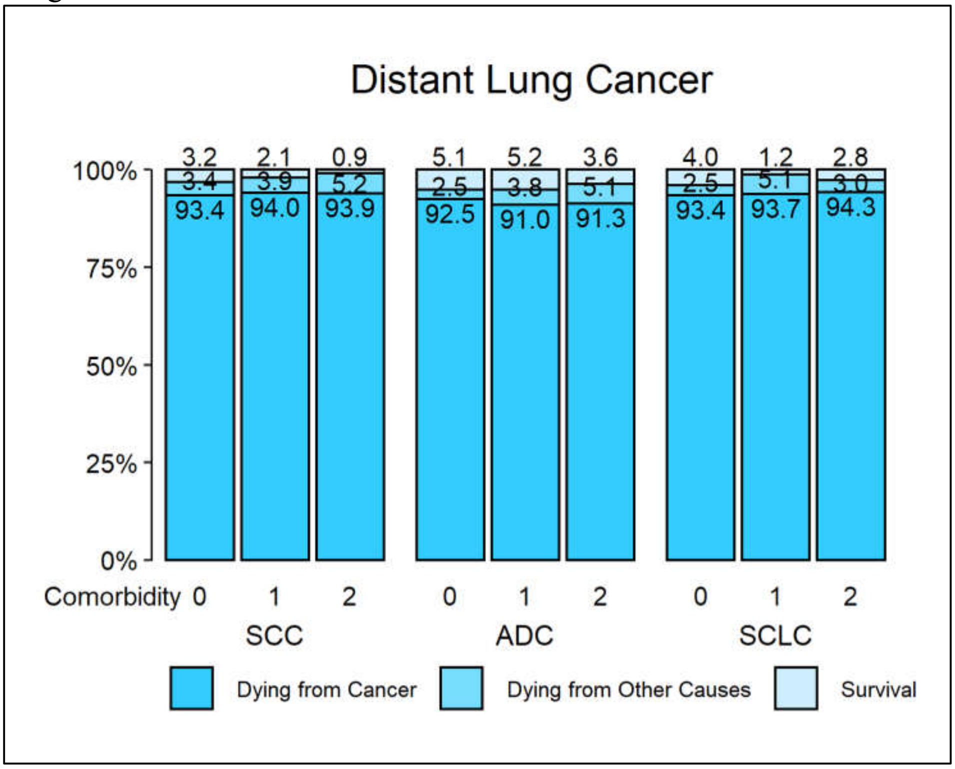

Figure S4A. One-year probabilities of dying from cancer, dying from other causes, and survival stratified by comorbidity level and year of diagnosis for distant **lung ADC** patients ages 30—94.

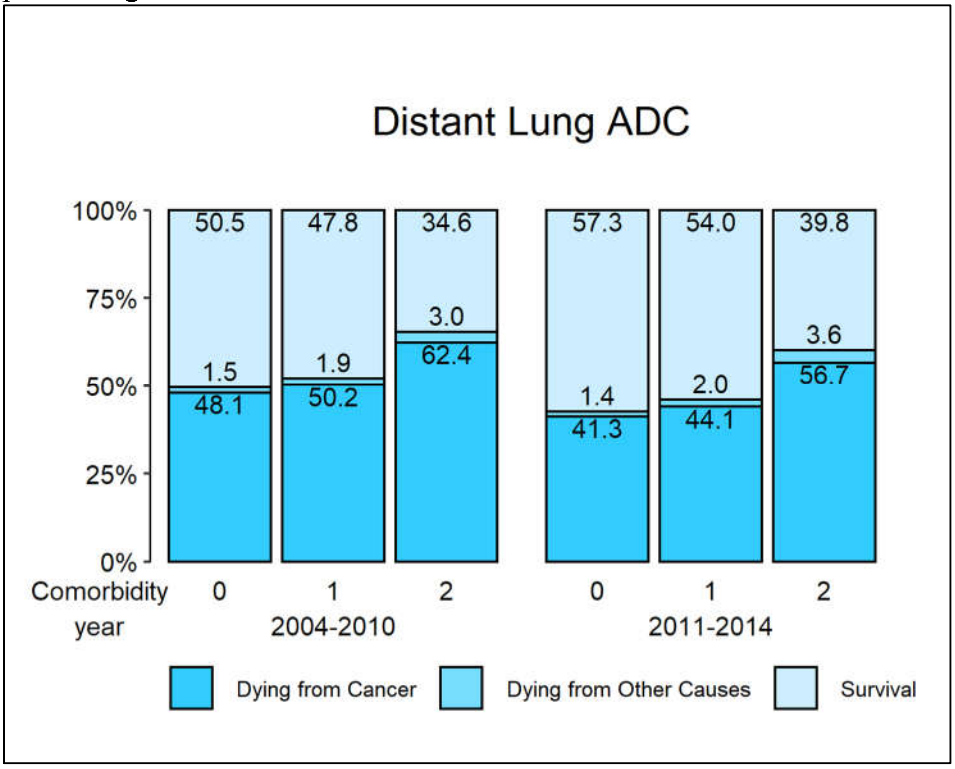

Figure S4B. Two-year probabilities of dying from cancer, dying from other causes, and survival stratified by comorbidity level and year of diagnosis for distant **lung ADC** patients ages 30—94.

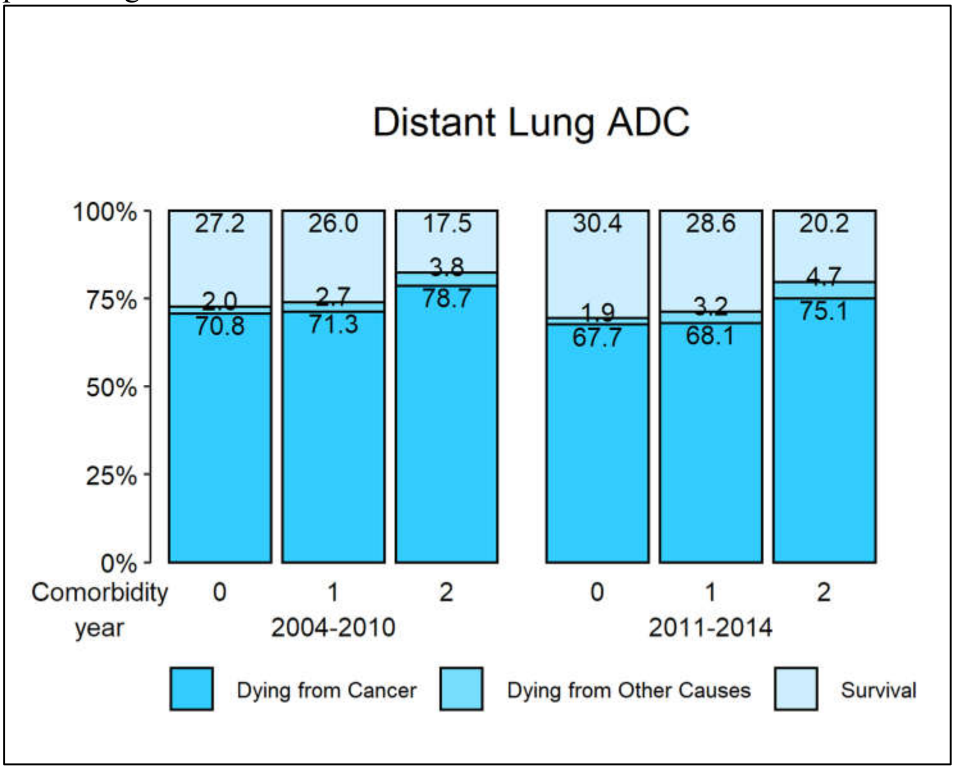

Figure S4C. Five-year probabilities of dying from cancer, dying from other causes, and survival are stratified by comorbidity level and year of diagnosis for distant **lung ADC** patients ages 30—94.

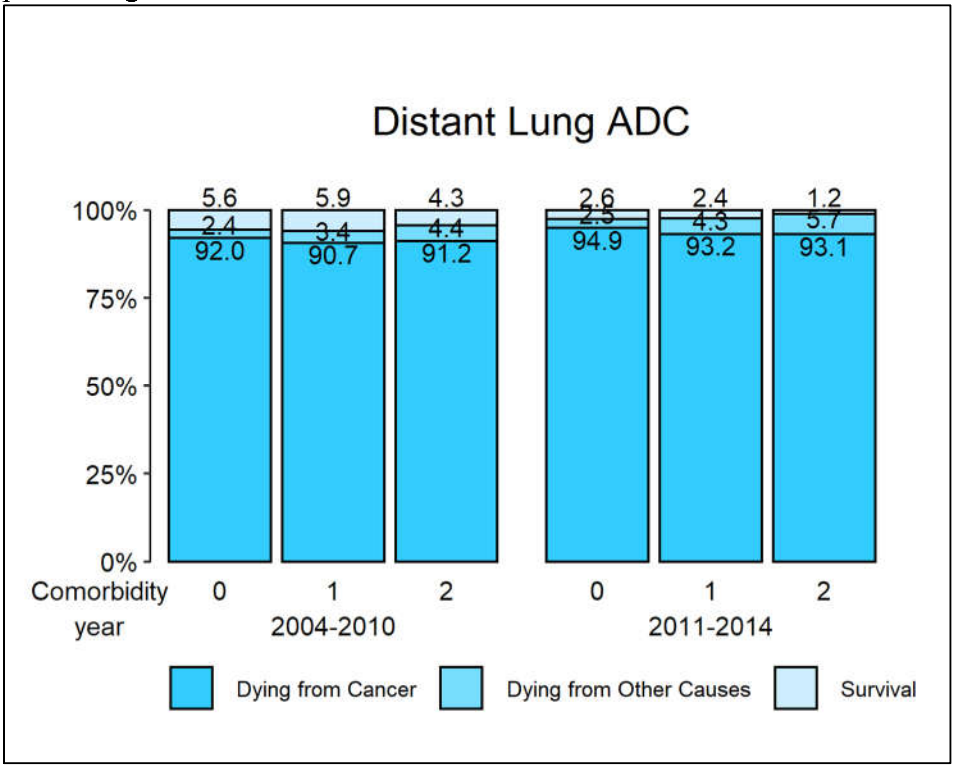

Figure S5. Probabilities of dying from cancer, dying from other causes and survival stratified by stage, comorbidity level, age, and certain specific sex-subtypes for **lung cancer**

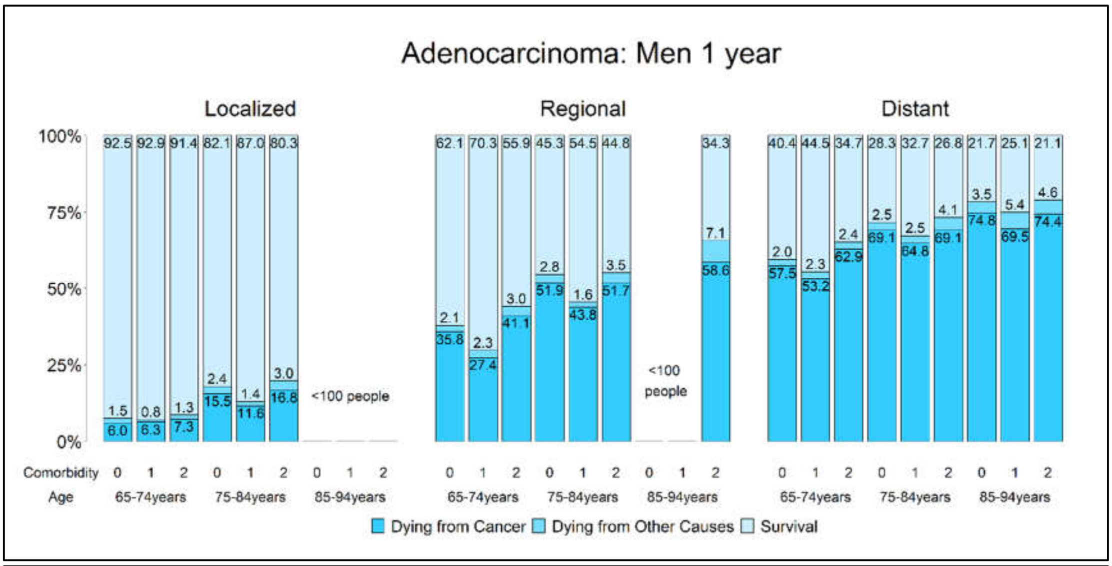

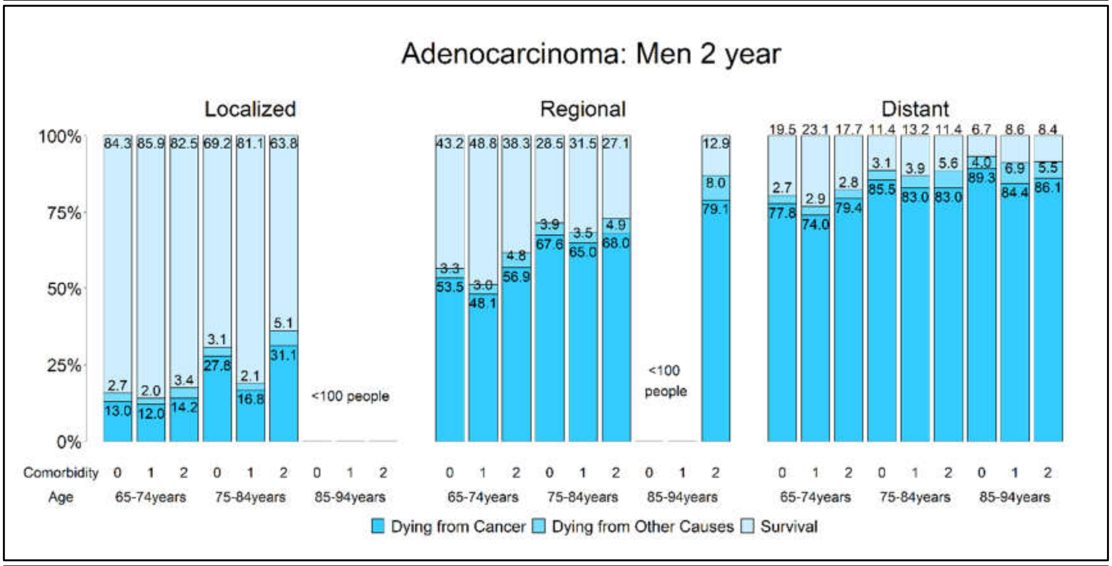

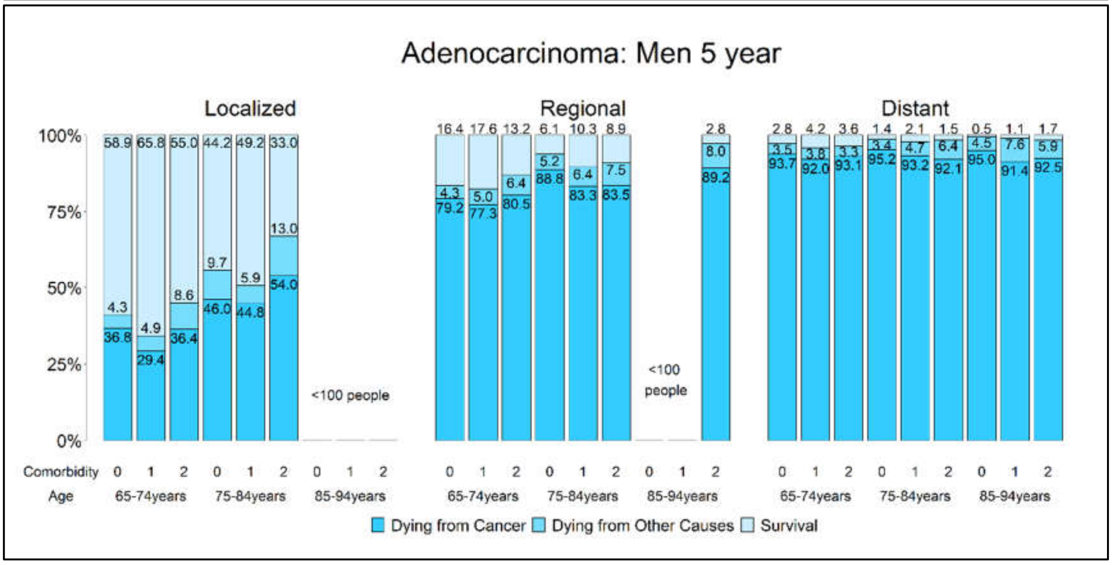

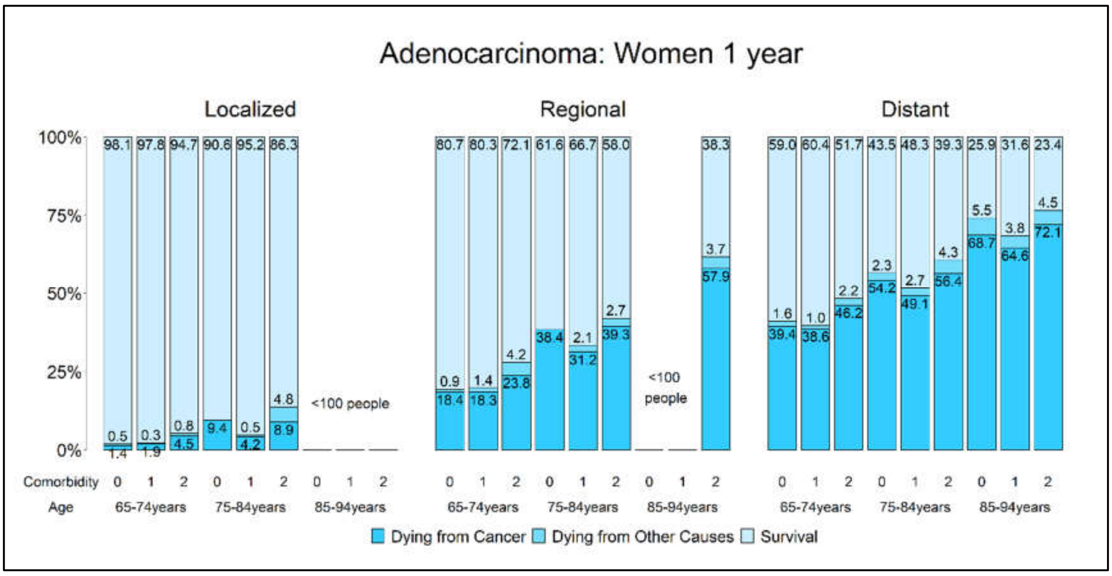
Adenocarcinoma: Women 1 year
Localized
Regional
Distant
<100 people
Comorbidity
Age
65-74years
75-84years
85-94years
Dying from Cancer Dying from Other Causes Survival

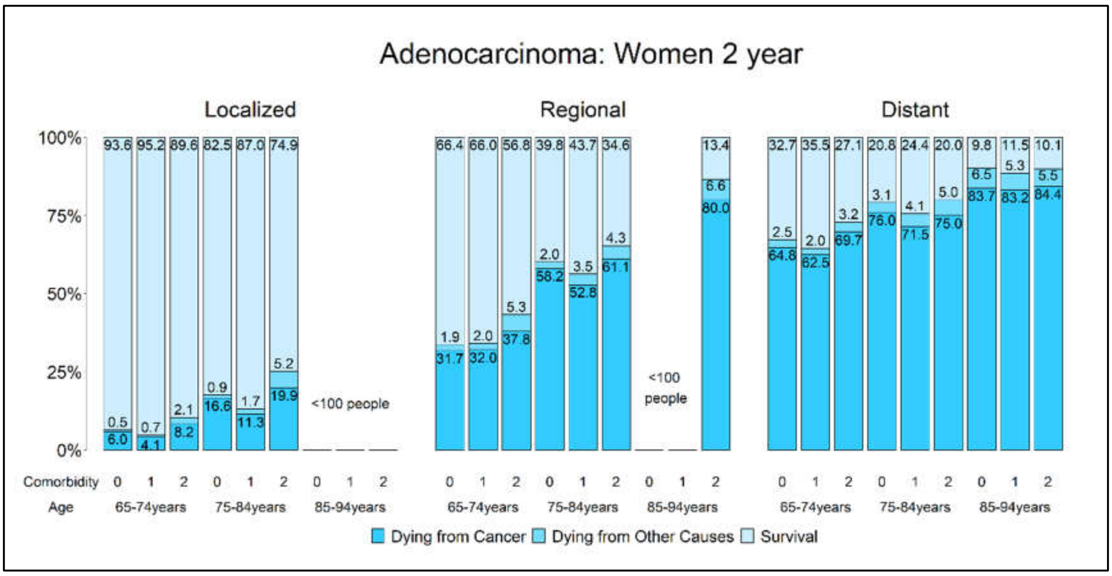
Adenocarcinoma: Women 2 year
Localized
Regional
Distant
<100 people
Comorbidity
Age
65-74years
75-84years
85-94years
Dying from Cancer Dying from Other Causes Survival

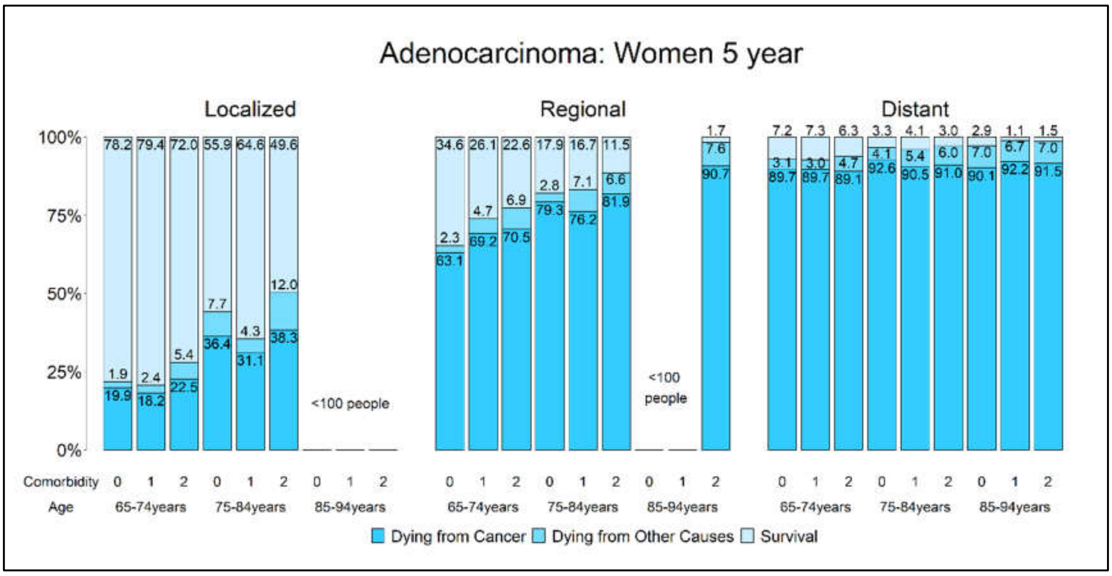
Adenocarcinoma: Women 5 year
Localized
Regional
Distant
<100 people
Comorbidity
Age
65-74years
75-84years
85-94years
Dying from Cancer Dying from Other Causes Survival

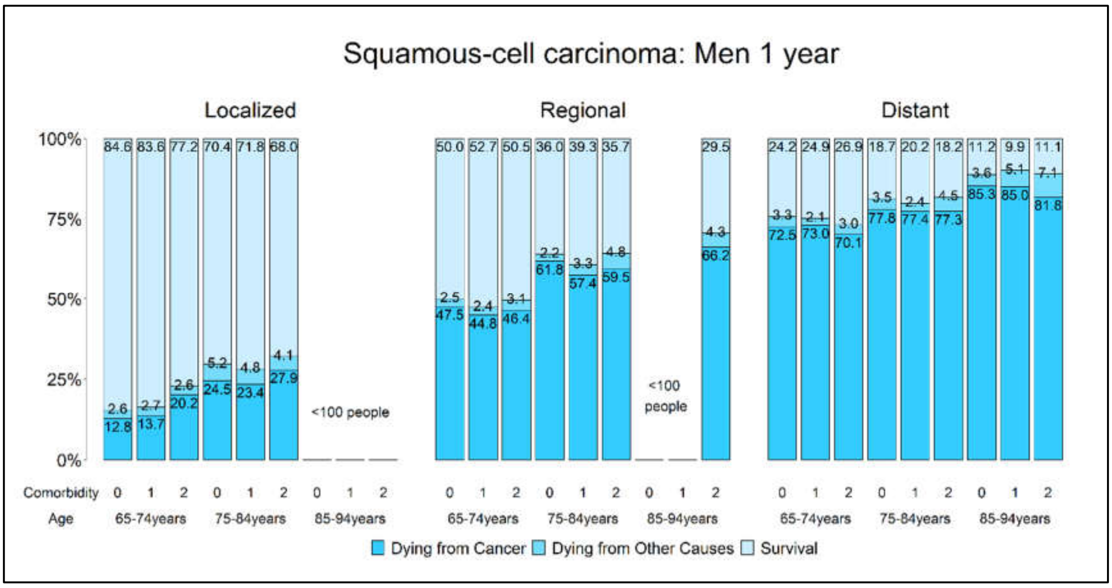
Squamous-cell carcinoma: Men 1 year
Localized
Regional
Distant
100%
75%
50%
25%
0%
84.6 83.6 77.2 70.4 71.8 68.0
2.6 2.7 2.6 5.2 4.8 4.1
12.8 13.7 20.2 24.5 23.4 27.9
<100 people
50.0 52.7 50.5 36.0 39.3 35.7
2.5 2.4 3.1 2.2 3.3 4.8
47.5 44.8 46.4 61.8 57.4 59.5
<100 people
29.5
4.3
66.2
24.2 24.9 26.9 18.7 20.2 18.2 11.2 9.9 11.1
3.3 2.1 3.0 3.5 2.4 4.5 3.6 5.1 7.1
72.5 73.0 70.1 77.8 77.4 77.3 85.3 85.0 81.8
Comorbidity 0 1 2 0 1 2 0 1 2
Age 65-74years 75-84years 85-94years
Dying from Cancer
Dying from Other Causes
Survival

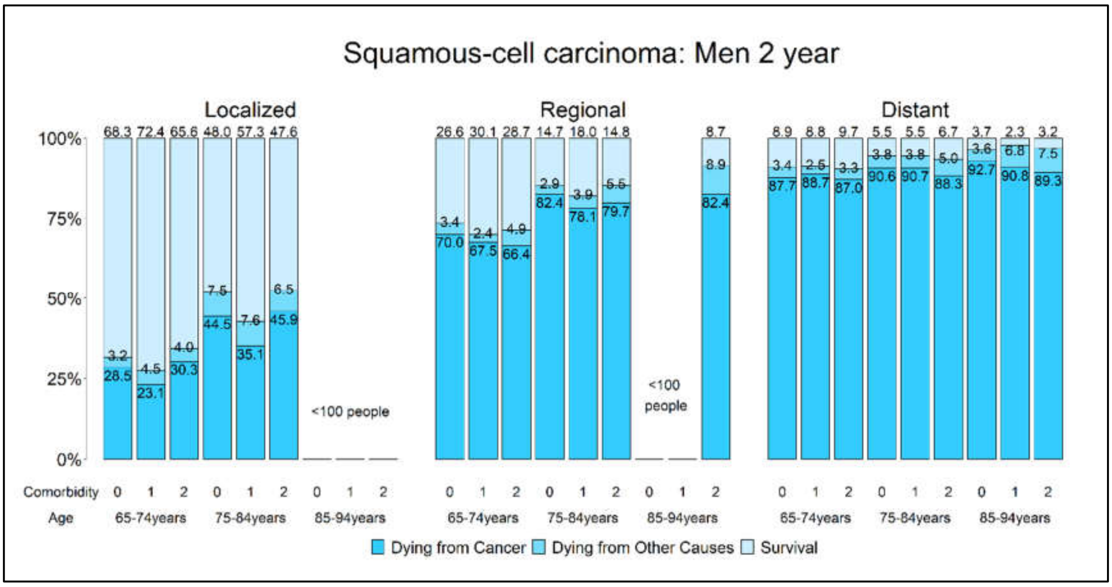
Squamous-cell carcinoma: Men 2 year
Localized
Regional
Distant
100%
75%
50%
25%
0%
68.3 72.4 65.6 48.0 57.3 47.6
3.2 4.5 4.0 7.5 7.6 6.5
28.5 23.1 30.3 44.5 35.1 45.9
<100 people
26.6 30.1 28.7 14.7 18.0 14.8
3.4 2.4 4.9 2.9 3.9 5.5
70.0 67.5 66.4 82.4 78.1 79.7
<100 people
8.7
8.9
82.4
8.9 8.8 9.7 5.5 5.5 6.7 3.7 2.3 3.2
3.4 2.5 3.3 3.8 3.8 5.0 3.6 6.8 7.5
87.7 88.7 87.0 90.6 90.7 88.3 92.7 90.8 89.3
Comorbidity 0 1 2 0 1 2 0 1 2
Age 65-74years 75-84years 85-94years
Dying from Cancer
Dying from Other Causes
Survival

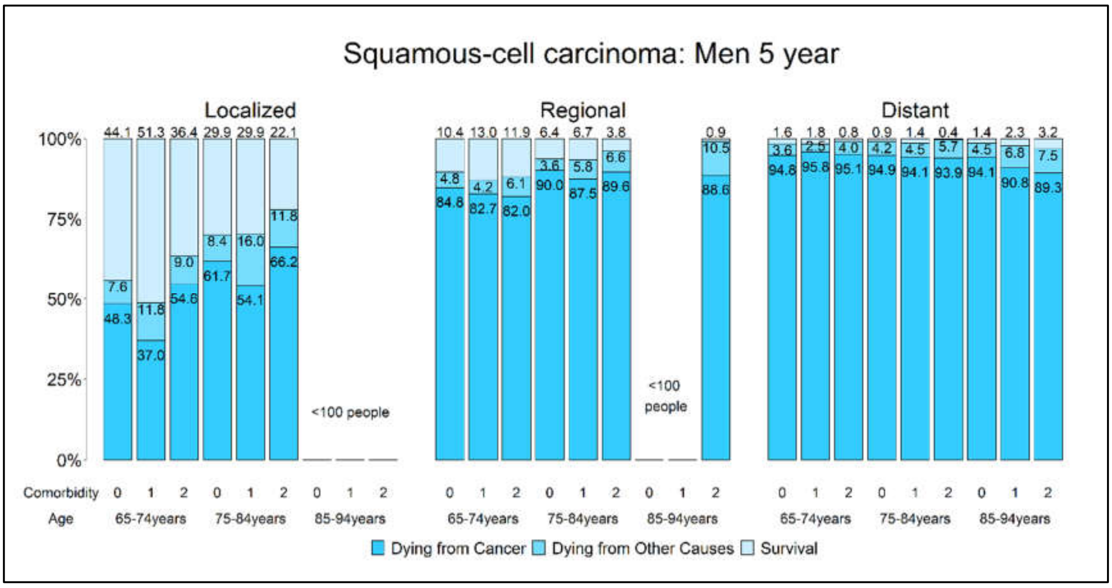
Squamous-cell carcinoma: Men 5 year
Localized
Regional
Distant
100%
75%
50%
25%
0%
44.1 51.3 36.4 29.9 29.9 22.1
7.6 11.8 9.0 8.4 16.0 11.8
48.3 37.0 54.6 61.7 54.1 66.2
<100 people
10.4 13.0 11.9 6.4 6.7 3.8
4.8 4.2 6.1 3.6 5.8 6.6
84.8 82.7 82.0 90.0 87.5 89.6
<100 people
0.9
10.5
88.6
1.6 1.8 0.8 0.9 1.4 0.4 1.4 2.3 3.2
3.6 2.5 4.0 4.2 4.5 5.7 4.5 6.8 7.5
94.8 95.8 95.1 94.9 94.1 93.9 94.1 90.8 89.3
Comorbidity 0 1 2 0 1 2 0 1 2
Age 65-74years 75-84years 85-94years
Dying from Cancer
Dying from Other Causes
Survival
